# Extracorporeal Shock Wave Therapy for Wound Management: Clinical Evidence, Energy Delivery Parameters and Mechanistic Insights — A Systematic Review

**DOI:** 10.1101/2025.07.28.25332291

**Authors:** Brent Musolf, Carmen Nussbaum-Krammer, Michael O’Neal, Tobias Wuerfel, Nicola Maffulli, Christoph Schmitz

## Abstract

**Background/Objectives:** Chronic wounds such as diabetic foot ulcers, venous leg ulcers and burns are a major health-care burden. Extracorporeal shock wave therapy (ESWT) is a non-invasive treatment that promotes angiogenesis and tissue regeneration. This review synthesizes the clinical evidence for ESWT in wound management, and asks how contemporary reviews reflect it.

**Methods:** PubMed and Ovid/Embase were searched from inception to 1 September 2026 according to PRISMA, regardless of wound type, study design or ESWT modality. Fifty-one clinical studies and 46 prior reviews were analyzed. Heterogeneity confined the primary synthesis to a qualitative one; an exploratory pooled analysis was performed for the one endpoint reported consistently enough to permit it.

**Results:** None of the 46 previously published reviews covered the original evidence in its full breadth: each cited a median of 19.4% of the clinical studies available at its reference date (range 0-90.9%), and systematic reviews covered no more than narrative ones. Most studies reported improved healing across all four modalities. In seven controlled trials reporting complete healing as events and denominators, the pooled risk ratio favored ESWT (1.51, 95% CI 1.22-1.87; I^2^ = 0%; 689 patients). Such data existed for three modalities only, resting on one trial for two, so modalities could not be compared.

**Conclusions:** ESWT is a useful adjunct for acute and chronic wounds. Because no previous review reflected the evidence base as a whole, conclusions drawn from it should be read with caution. Head-to-head trials with matched physical dose are needed before any modality can be preferred.

## 1. Introduction

Wound healing is a complex and tightly regulated biological process involving overlapping phases of inflammation, angiogenesis, cellular proliferation and tissue remodeling [1–3]. Under physiological conditions, these processes proceed in a coordinated manner that restores tissue integrity and function. However, systemic disease, infection, ischemia and other pathological factors can disrupt this sequence, resulting in wounds that fail to progress through the normal stages of healing [4]. Such chronic wounds, including diabetic foot ulcers (DFUs), venous leg ulcers (VLUs), pressure ulcers (PUs) and burn-related wounds, represent a major clinical challenge and are associated with substantial morbidity, impaired quality of life and high healthcare costs. As the global prevalence of diabetes, vascular disease and aging-related comorbidities continues to increase, the burden of chronic wounds is expected to rise further, underscoring the need for effective and accessible therapeutic strategies [5–7].

Among emerging therapeutic approaches, extracorporeal shock wave therapy (ESWT) has attracted increasing attention as a non-invasive modality capable of modulating biological processes involved in tissue repair [8–10]. Initially developed for lithotripsy, ESWT has subsequently been applied to a broad spectrum of musculoskeletal and soft-tissue conditions. In wound healing, extracorporeal shock waves (ESWs) are thought to exert biological effects through mechanotransduction pathways that stimulate angiogenesis, regulate inflammatory signaling, and promote cellular proliferation and tissue regeneration [11–13]. In experimental studies, shock wave exposure increased the expression of growth factors, enhanced microvascular perfusion and activated cellular pathways associated with tissue repair, suggesting that ESWT may influence multiple components of the wound healing cascade [12].

Several technological approaches have been developed to generate ESWs for therapeutic applications. These include electrohydraulic, electromagnetic, piezoelectric and ballistic (radial) systems [9,11,14]. Electrohydraulic devices generate ESWs through high-voltage electrical discharge in water, producing a rapidly expanding and collapsing vapor bubble whose resulting pressure wave can be focused or left unfocused, depending on the reflector geometry (Figure 1a,b).

**Figure 1.**
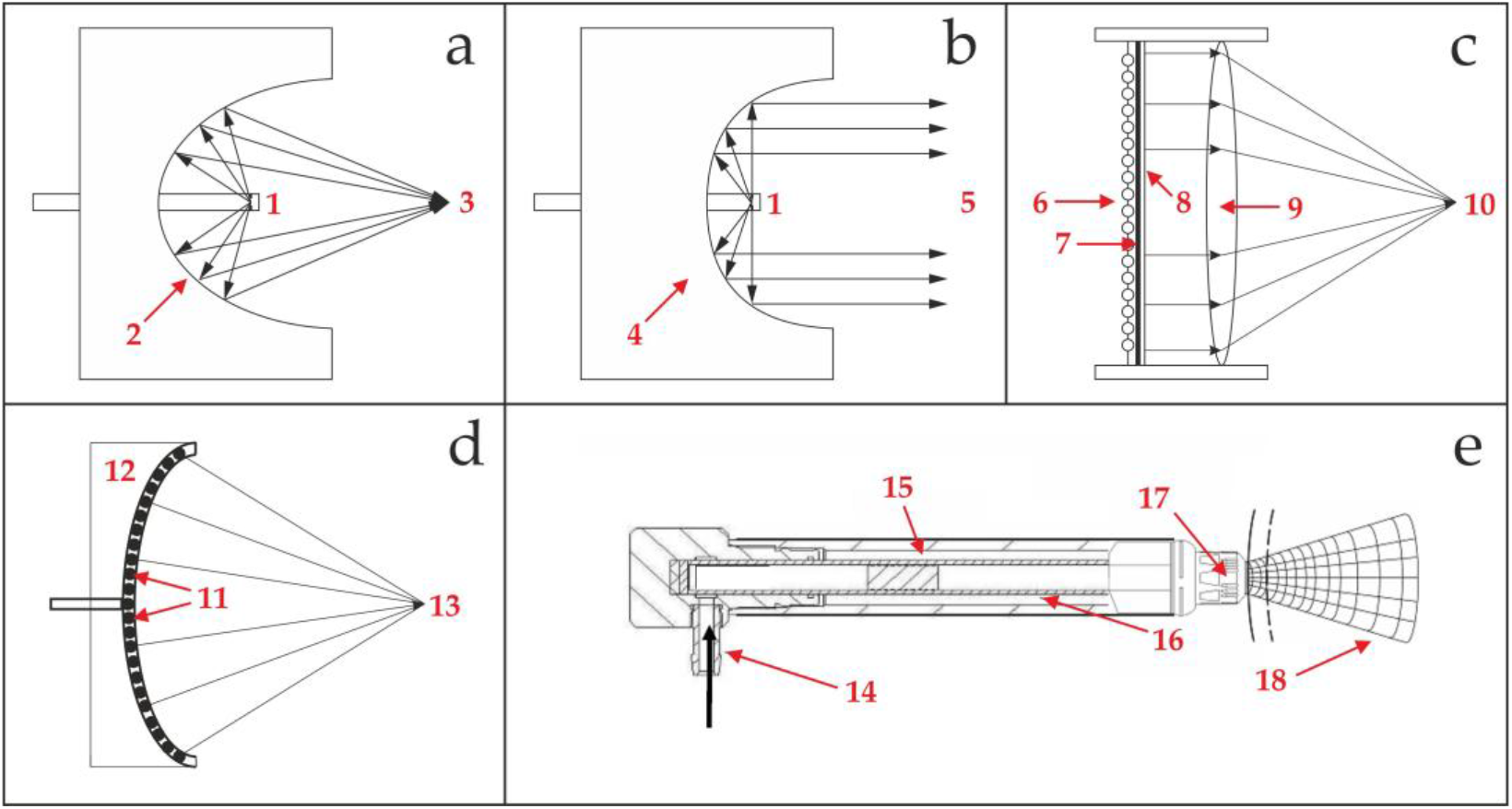
Schematic overview of different technical modalities for generating focused, unfocused and radial extracorporeal shock waves (ESWs) (modified from [9]). (**a**) Electrohydraulic modality to generate focused ESWs: a high-voltage discharge between two electrode tips (1) in water creates a vapor bubble filled with plasma, which rapidly expands and collapses, generating a shock wave. A reflector (2) focuses this shock wave at a second focal point (3). (**b**) Electrohydraulic modality to generate unfocused ESWs: with an alternative reflector (4), shock waves generated by high-voltage discharge between two electrode tips (1) in water remain unfocused (5). (**c**) Electromagnetic modality to generate focused ESWs: a magnetic field from a current in a coil (6) induces movement in a metal membrane (7), which propels another membrane (8) in fluid, producing a pressure wave. An acoustic lens (9) focuses this wave at a focal point (10). (**d**) Piezoelectric modality to generate focused ESWs: numerous piezocrystals (11) in a bowl-shaped device (12) deform in response to an electric discharge, creating pressure waves that focus at a point (13) due to the bowl’s design. (**e**) Ballistic modality to generate radial ESWs: compressed air (14) or a magnetic field propels a projectile (15) down a tube (16) to strike a metal applicator (17) on the skin, generating radial ESWs transmitted into tissue (18).

Electromagnetic systems use a magnetic field to induce rapid membrane displacement, producing pressure waves that are subsequently focused using an acoustic lens (Figure 1c). In piezoelectric devices, arrays of piezocrystals deform in response to electrical stimulation, producing convergent pressure waves that form a focused acoustic pulse (Figure 1d). In contrast, ballistic or radial systems accelerate a projectile against a metal applicator positioned on the skin surface, generating pressure waves that propagate radially into the surrounding tissue (Figure 1e).

The distinction between focused ESWT (fESWT) and radial ESWT (rESWT) is often discussed in both clinical and physical terms (e.g., [9,13,15–17]). In a strict physical sense, electromagnetic and piezoelectric devices initially generate pressure waves that can evolve into shock waves through nonlinear propagation during transmission through tissue [18]. At sufficiently high amplitudes, these waveforms display characteristic features of true shock waves, including rapid rise times and high peak pressures comparable to those produced by electrohydraulic sources. At lower amplitudes, however, the waveform characteristics may differ from true shock waves [18]. These physical considerations are relevant when interpreting the properties of specific devices, such as the Duolith SD1 system (Storz Medical, Tägerwilen, Switzerland), which produces focused pressure waves rather than true shock waves [19]. In clinical research, however, a broader operational definition of ESWs is commonly adopted. According to this definition, ESWs are characterized by peak pressures between approximately 10 and 100 MPa, rise times below 1 μs and a tensile component corresponding to approximately one-tenth of the peak positive pressure [20]. This definition encompasses focused, unfocused and radial modalities and reflects their shared therapeutic application in clinical practice.

Over the past two decades, a growing body of experimental and clinical research has explored the role of ESWT in wound management. Preclinical studies have demonstrated biological effects relevant to tissue repair, including stimulation of angiogenesis, modulation of inflammatory pathways and enhancement of fibroblast activity [12]. Clinical investigations, ranging from case reports and observational cohort studies to randomized controlled trials (RCTs), have examined the application of ESWT across diverse wound types, including DFUs, VLUs, PUs, postsurgical wounds and burn injuries. Collectively, these studies suggest that ESWT may accelerate wound healing, improve tissue perfusion and enhance epithelialization in selected patient populations.

Several narrative and systematic reviews have summarized aspects of this emerging literature. However, these reviews differ substantially in their methodological approaches, search strategies and inclusion criteria, leading to considerable variation in the number and types of studies considered. Consequently, the overall evidence base has remained fragmented, and comparisons between different ESWT technologies, treatment parameters and clinical outcomes have often been limited. In addition, relatively few reviews have examined the distribution of ESWT modalities across the clinical literature or explored whether differences in energy delivery parameters may influence therapeutic outcomes.

Given the increasing clinical interest in ESWT as a potential adjunctive therapy for wound management, a comprehensive and updated synthesis of the available clinical evidence is warranted. The present systematic review therefore aims to provide a structured evaluation of clinical studies investigating ESWT in wound healing. Specifically, the objectives of this review are to (i) identify as comprehensively as possible the clinical studies examining ESWT for wound management across different wound types and treatment settings, combining a database search with reference-list screening of all identified reviews; (ii) analyze the representation of different ESWT modalities within the clinical literature; (iii) evaluate reported therapeutic outcomes and methodological characteristics of the included studies; and (iv) explore potential relationships between ESWT treatment parameters and clinical outcomes.

## 2. Materials and Methods

This systematic review was conducted in accordance with the Preferred Reporting Items for Systematic Reviews and Meta-Analyses (PRISMA 2020) guidelines [21]. A systematic literature search was performed using the PubMed and Ovid/Embase databases to identify clinical studies investigating the use of ESWT in wound management. The search covered all records from database inception through September 1, 2026. Search terms were designed to capture studies addressing wound healing and shock wave therapy and included combinations of the terms “wound”, “ulcer”, “shock wave”, “shockwave” and “shockwaves.” Variations of these terms were applied in both databases to ensure comprehensive retrieval of relevant publications. The complete list of search terms used in the database queries is provided in Table 1.

**Table 1.** Search terms used in the systematic search on PubMed and Ovid/Embase (conducted on September 1, 2026), along with the number of records (N_R_).

| No. | PubMed |  | Ovid/Embase |  |
| --- | --- | --- | --- | --- |
|  | Search terms | N<sub>R</sub> | Search terms | N<sub>R</sub> |
| 1 | Wound healing shock wave | 218 | Wound AND healing AND shock AND wave | 412 |
| 2 | Wound healing shock waves | 169 | Wound AND healing AND shock AND waves | 80 |
| 3 | Wound healing shockwave | 211 | Wound AND healing AND shockwave | 164 |
| 4 | Wound healing shockwaves | 211 | Wound AND healing AND shockwaves | 45 |
| 5 | Ulcer shock wave | 60 | Ulcer AND healing AND shock AND wave | 243 |
| 6 | Ulcer shock waves | 35 | Ulcer AND healing AND shock AND waves | 39 |
| 7 | Ulcer shockwave | 88 | Ulcer AND healing AND shockwave | 75 |
| 8 | Ulcer shockwaves | 88 | Ulcer AND healing AND shockwaves | 17 |
| 9 | Ulcers shock wave | 60 | Ulcers AND healing AND shock AND wave | 161 |
| 10 | Ulcers shock waves | 35 | Ulcers AND healing AND shock AND waves | 33 |
| 11 | Ulcers shockwave | 88 | Ulcers AND healing AND shockwave | 70 |
| 12 | Ulcers shockwaves | 88 | Ulcers AND healing AND shockwaves | 20 |
| Total |  | 1351 |  | 1359 |

The initial search identified 1,351 records in PubMed and 1359 records in Ovid/Embase, yielding 2,710 records before duplicate removal. After removing 1,878 duplicates, the remaining records were screened based on titles and abstracts to identify potentially relevant studies.

Studies were considered eligible if they reported clinical investigations involving human subjects and evaluated the application of ESWT for wound management. A broad range of study designs was included to capture the full scope of available evidence, including reviews, RCTs, cohort studies with control groups, uncontrolled case series and case reports.

Records were excluded if they were editorials, commentaries, letters to the editor or studies investigating unrelated clinical conditions. Studies involving animal models or in vitro experiments were also excluded.

Following the initial screening, 832 records were assessed for relevance, of which 742 were excluded according to the predefined criteria. The remaining 90 publications were successfully retrieved through the E-Media library of Ludwig-Maximilians University of Munich (LMU Munich, Germany) or through alternative sources.

Additionally, the reference lists of all reviews identified during the literature search (n = 46) were screened to identify studies not captured in PubMed or Ovid/Embase. This process yielded 8 additional publications. One of these records (reference 41 in [22]) could not be verified because the provided link redirected to an unrelated publication and the journal website did not list the article; therefore, it was excluded from further analysis. The other 7 additional publications were also successfully retrieved through the E-Media library of LMU Munich or through alternative sources.

For articles published in languages other than English, translations were performed using the large language model Claude (Opus 5; 2026; Anthropic, San Francisco, CA, USA) to enable assessment of eligibility and extraction of relevant study data.

Each review was assigned a reference date, defined as the date up to which its literature search was reported to include studies. Where no such date was stated, the date of submission was used; where neither was stated, the date of publication minus six months was used. A clinical study was counted as available to a given review if it had been published on or before that review’s reference date. The inclusion and exclusion criteria applied by the individual reviews were deliberately not taken into account in this comparison, because the question addressed here is which part of the published evidence each review could have covered, not which part it set out to cover.

After full-text assessment, all 51 clinical studies met the inclusion criteria and were included in the final analysis. The study selection process is summarized in the PRISMA flow diagram (Figure 2).

**Figure 2.**
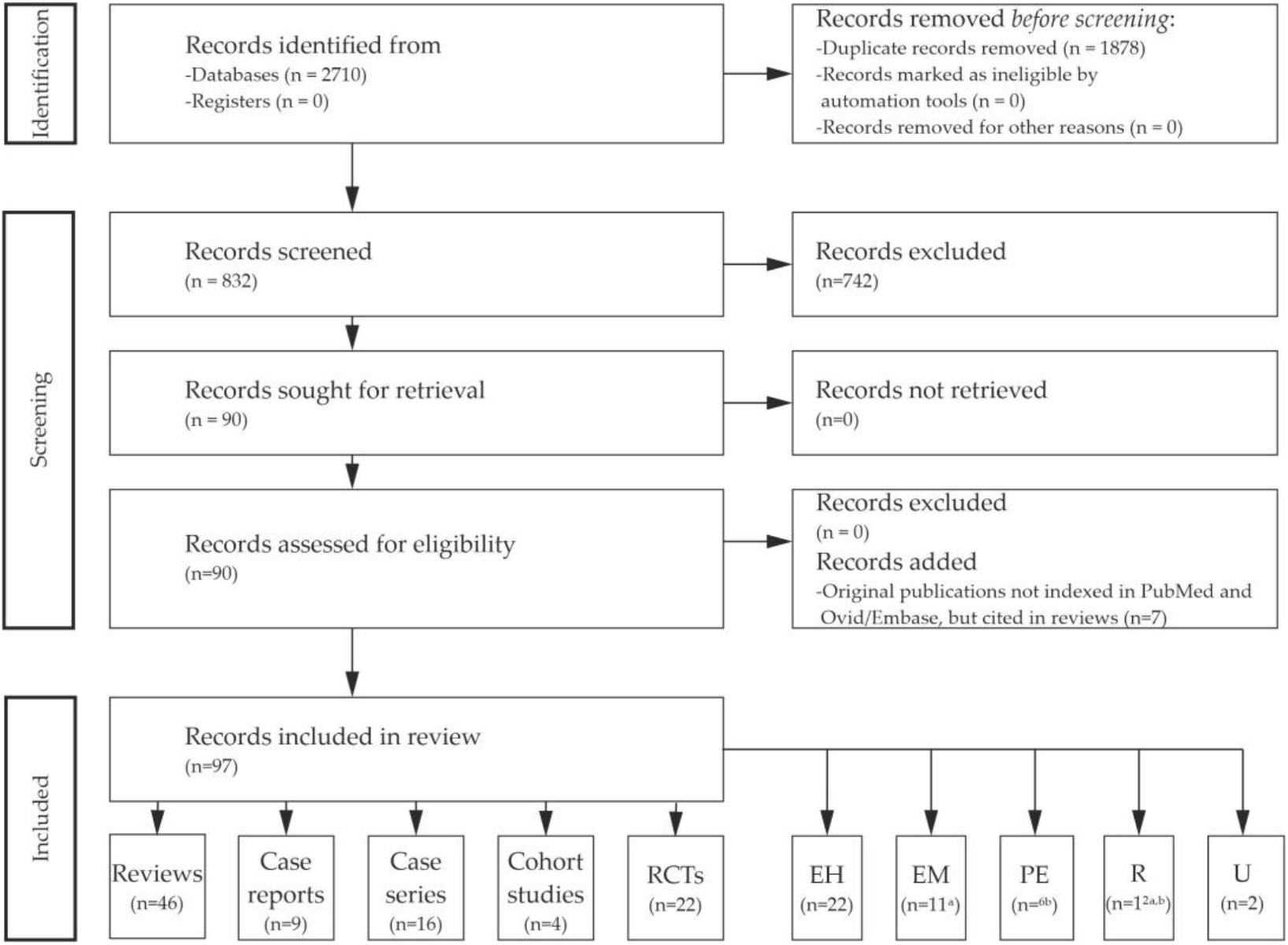
Systematic review flow chart of the literature search regarding extracorporeal shock wave therapy in wound management, performed according to the PRISMA guidelines [21] on September 1, 2026. Abbreviations: RCTs, randomized controlled trials; EH, electrohydraulic modality; EM, electromagnetic modality; PE, piezoelectric modality; R, radial modality; U, ESWT modality not specified. Notes: ^a^, one case report employed both electromagnetic fESWT and ballistic rESWT devices; ^b^, two RCTs employed both piezoelectric fESWT and ballistic rESWT devices

Among the 51 studies, 22 were reported by their authors as RCTs, 4 were cohort studies with non-randomized controls, 16 were uncontrolled case series and 9 were case reports. In three of the studies described as randomized, allocation followed the calendar date of referral, the order in which patients presented, or a method that was not stated [23–25]; these were treated in the present analysis as controlled but not randomized.

Of the 51 studies, 22 used electrohydraulic devices, 11 electromagnetic devices, 6 piezoelectric devices and 12 radial devices, while in 2 studies the technical modality was not disclosed. Two randomized trials used both a piezoelectric and a radial device, which is the reason why the sum of studies (53) reported here exceeds the number of studies (51). To enable systematic comparison across studies, 40 predefined variables were extracted from each publication, covering study design, patient characteristics, treatment parameters and reported outcomes. A detailed overview of all extracted variables is presented in Table 2.

**Table 2.** Variables (V) extracted from the 51 included records. Abbreviations: ESWs, extracorporeal shock waves; ESWT, extracorporeal shock wave therapy; fESWT, focused ESWT; uESWT, unfocused ESWT; rESWT, radial ESWT; PEDro, Physiotherapy Evidence Database (cf. [9]).

| V | Description |
| --- | --- |
| V1 | Wound type (e.g. ulcer, burn, mixed, etc.) |
| V2 | Mean size of wounds per group |
| V3 | Duration of wounds (acute, chronic, since when, etc.) |
| V4 | Study design (case report, case series, cohort study, randomized controlled trial) |
| V5 | Overall outcome |
| V6 | Types of intervention (ESWT group) |
| V7 | Types of intervention (control group, if applicable) |
| V8 | Number of patients in the ESWT group |
| V9 | Number of patients in the control group (if applicable) |
| V10 | ESWT device used |
| V11 | ESWT modality used |
| V12 | Energy flux density (EFD) used |
| V13 | Any description, whether the provided EFD was the positive or the total EFD |
| V14 | Number of ESWs per treatment session |
| V15 | Number of ESWs per square cm wound area, or any related measure |
| V16 | Frequency (in Hertz) at which the ESWs were applied |
| V17 | Number of treatment sessions |
| V18 | Interval between treatment sessions |
| V19 | Primary endpoint |
| V20 | Time post-baseline when the primary endpoint was assessed |
| V21 | Secondary endpoints (if any) |
| V22 | Time post-baseline when the secondary endpoints were assessed |
| V23 | Description of a prospective sample size calculation and/or prospective power analysis |
| V24 | Outcome of the prospective sample size calculation and/or prospective power analysis |
| V25 | Results obtained for the primary endpoint, separated for the ESWT group and the control group (if applicable), with results of the statistical analysis |
| V26 | Results obtained for the secondary endpoints, separated for the ESWT and the control group, with results of the statistical analysis (if applicable) |
| V27 | Description of a retrospective power analysis |
| V28 | Outcome of the retrospective power analysis |
| V29 | Description of any treatment-related adverse events |
| V30 | (PEDro assessment): eligibility criteria were specified (yes/no) |
| V31 | (PEDro assessment): subjects were randomly allocated to groups (yes/no) |
| V32 | (PEDro assessment): allocation was concealed (yes/no) |
| V33 | (PEDro assessment): the groups were similar at baseline regarding the most important prognostic indicators (yes/no/not applicable) |
| V34 | (PEDro assessment): there was blinding of all subjects (yes/no) |
| V35 | (PEDro assessment): there was blinding of all therapists who administered the therapy (yes/no) |
| V36 | (PEDro assessment): there was blinding of all assessors who measured at least one key outcome (yes/no) |
| V37 | (PEDro assessment): measures of at least one key outcome were obtained from more than 85% of the subjects initially allocated to groups (yes/no) |
| V38 | (PEDro assessment): all subjects for whom outcome measures were available received the treatment or control condition as allocated or, where this was not the case, data for at least one key outcome was analyzed by "intention to treat" (yes/no) |
| V39 | (PEDro assessment): the results of between-group statistical comparisons are reported for at least one key outcome (yes/no/not applicable) |
| V40 | (PEDro assessment): the study provides both point measures and measures of variability for at least one key outcome (yes/no) |

The overall result of each study was classified using the scheme summarized in Table 3.

**Table 3.** Categorization (C) of the overall outcome of the 51 included records (variable V5 in Table 2).

| C | Description |
| --- | --- |
| 1 | Study with control group; statistically significantly ($p < 0.05$) better treatment outcome with ESWT compared to sham treatment, no treatment or an alternative therapy. |
| 2a | Study with control group; statistically significant ($p < 0.05$) clinical improvement from baseline observed both in the ESWT group and in the group receiving an alternative treatment; no statistically significant difference in the outcome between the groups. |
| 2b | Study with control group; statistically significant ($p < 0.05$) clinical improvement from baseline observed both in the ESWT group and in the sham or no-treatment group; no statistically significant difference in the outcome between the groups. |
| 3 | Study with control group; no statistically significant ($p < 0.05$) clinical improvement from baseline in either the ESWT group or the sham, no-treatment or alternative treatment group. |
| 4 | Study with control group; statistically significantly better treatment outcome with sham treatment, no treatment or an alternative therapy compared to ESWT. |
| 5 | Study without control group. |

This classification distinguished between studies demonstrating statistically significant benefits of ESWT compared with control treatments, studies showing improvement without significant between-group differences, studies reporting no improvement and studies lacking control groups.

Methodological quality of RCTs was evaluated using criteria derived from the Physiotherapy Evidence Database (PEDro) scale, which assesses aspects of trial design such as randomization procedures, allocation concealment, baseline comparability, blinding of participants and outcome assessors, completeness of outcome reporting and statistical comparisons between groups (cf. [9]). Corresponding variables were extracted for each included study to provide an overview of methodological rigor across the available clinical evidence.

Because of substantial heterogeneity among the included studies in terms of wound types, ESWT technologies, treatment parameters, outcome definitions and follow-up intervals, a structured qualitative synthesis was chosen as the primary form of analysis, focusing on the characteristics of the included studies and their reported outcomes across wound types and ESWT modalities.

The extent to which this literature supports quantitative synthesis was itself assessed. Three questions were addressed: which endpoints were reported consistently enough to be pooled, whether the four ESWT modalities can be compared with one another, and whether treatment doses can be expressed on a common scale. For a dichotomous outcome, a record was considered poolable only if it reported the number of events together with the number of patients or wounds at risk in both an ESWT group and a control group; for a continuous outcome, only if it reported means with standard deviations. Values given as standard errors, or as medians with quartile ranges, were not converted, because the available conversions rest on distributional assumptions that cannot be verified from the published data. Where several publications reported the same participants, only one of them entered the quantitative analysis.

Complete wound healing was the only endpoint that met these requirements in more than a handful of records, and it was pooled in an exploratory analysis. Risk ratios with 95% confidence intervals were calculated for each trial and combined with a fixed-effect inverse-variance model; no event count was zero, so no continuity correction was applied.

Heterogeneity was quantified with Cochran Q and I-squared, and tau-squared was estimated by the DerSimonian-Laird method, with the corresponding random-effects estimate reported as a check on the fixed-effect result [26]. In the one three-arm trial [27], the two active arms were combined and compared with the sham arm. The robustness of the pooled estimate was examined by repeating the analysis after excluding the trial in which allocation followed the calendar date of referral [23], after restricting it to the sham-controlled trials [27,28], and after excluding the trials in which the comparator was hyperbaric oxygen therapy (HBOT) [23,30]. The calculations were performed with the standard formulae of the inverse-variance method rather than with a dedicated meta-analysis package.

Whether the modalities can be compared was assessed by counting, for each modality, the controlled trials that contributed healing counts and the uncontrolled records that contributed a healing proportion. An indirect comparison would have required either a meta-regression on modality, for which Cochrane guidance recommends at least ten studies, or a subgroup contrast in which each subgroup is independently estimable [26]. Whether dose can be expressed on a common scale was assessed by recording, for every study, the EFD and the quantity against which it was reported, the number of ESWs per session and per square centimeter of wound area, and whether the dose was scaled to the wound, fixed irrespective of wound size, or applied to an anatomical region.

## 3. Results

This systematic review assessed 46 reviews on ESWT for wound management published between 2010 and 2026 [22,31–75], as well as 51 clinical studies on ESWT published between 2005 and 2026 [23–25,27–30,76–119].

### 3.1. Assessment of Reviews of ESWT for Wound Management

Table 4 summarizes key characteristics and the main conclusions (in one sentence, each reflecting the conclusions of the authors of the respective review and not those of the present systematic review) for each review.

**Table 4.**
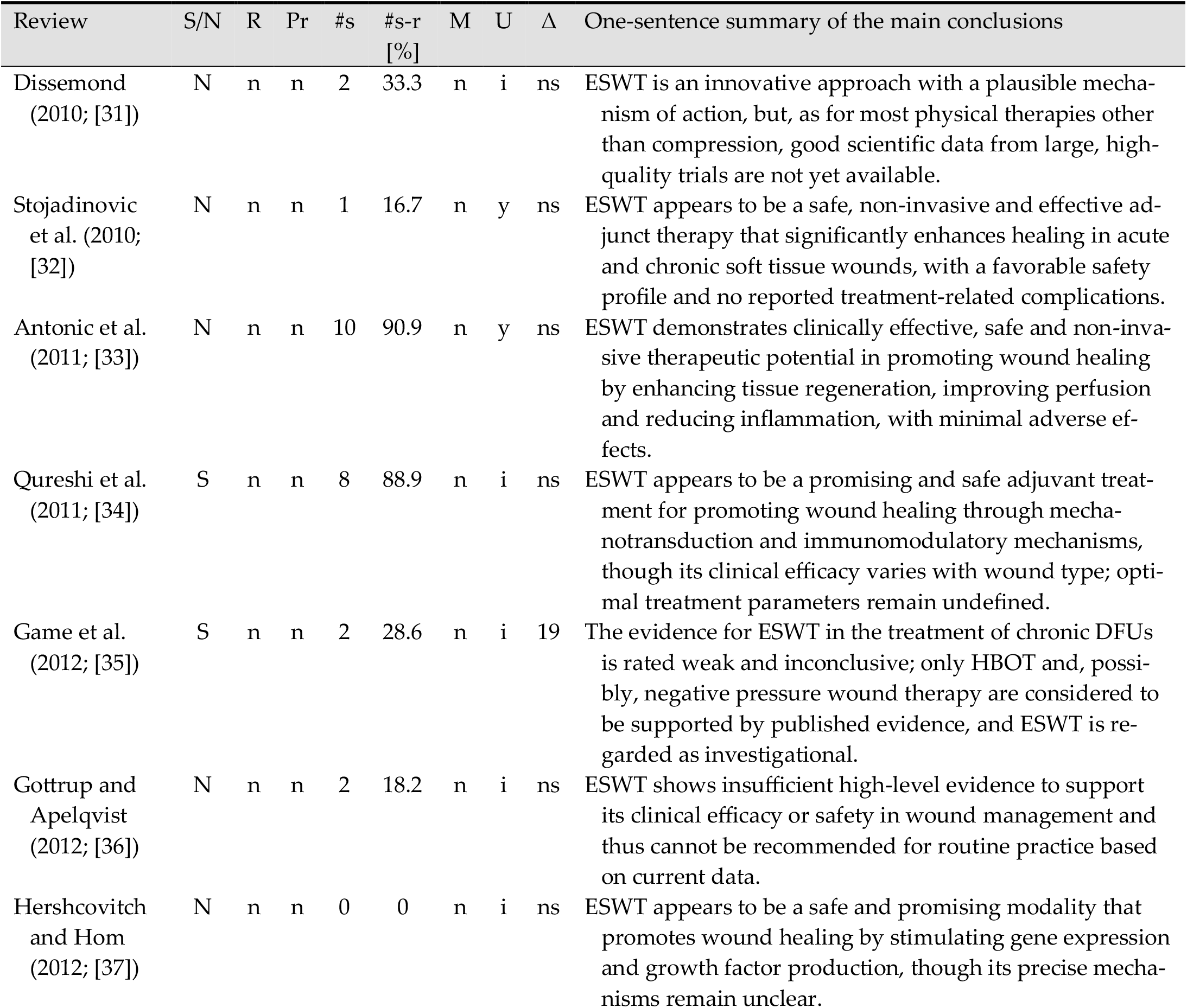
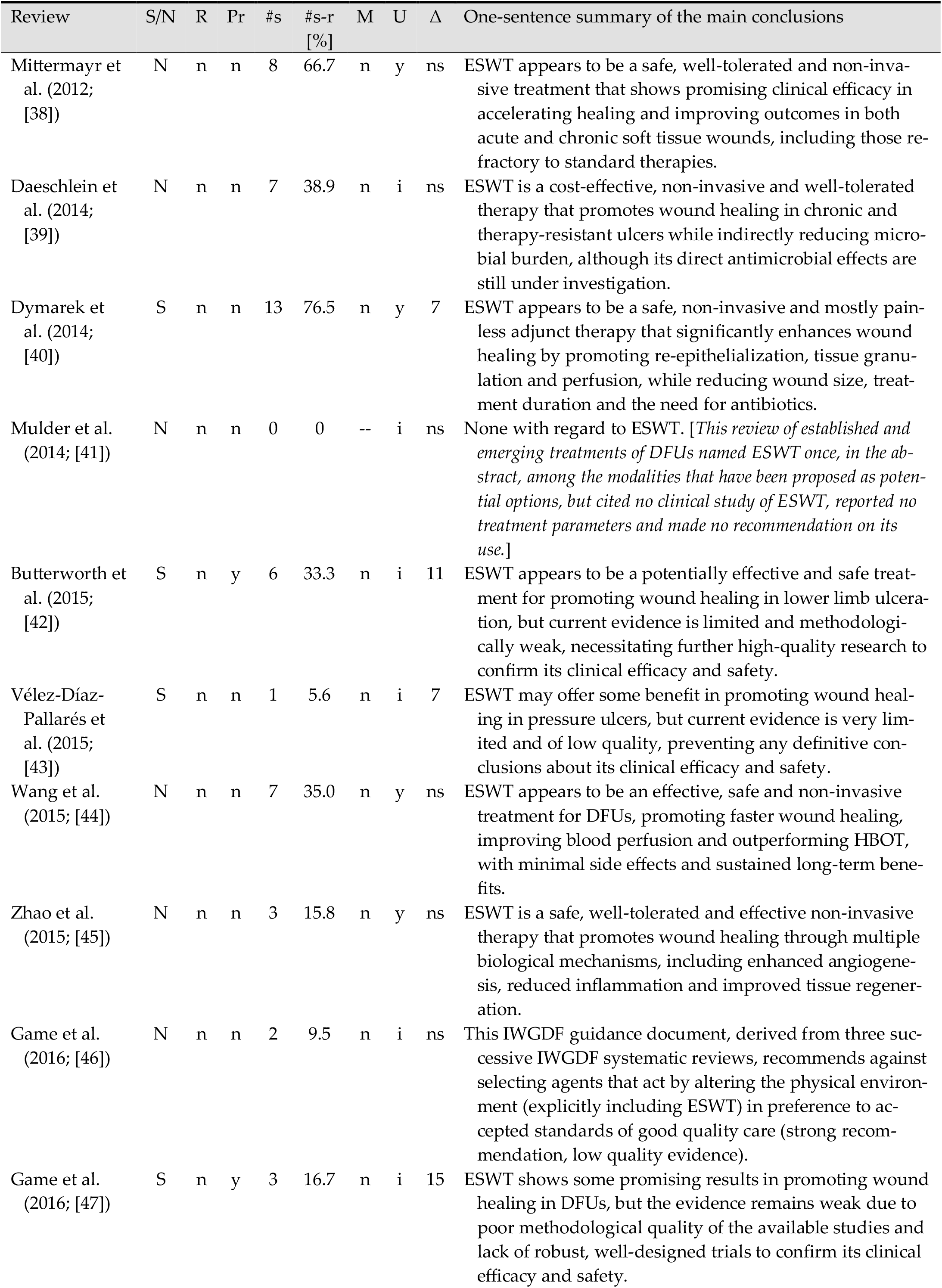
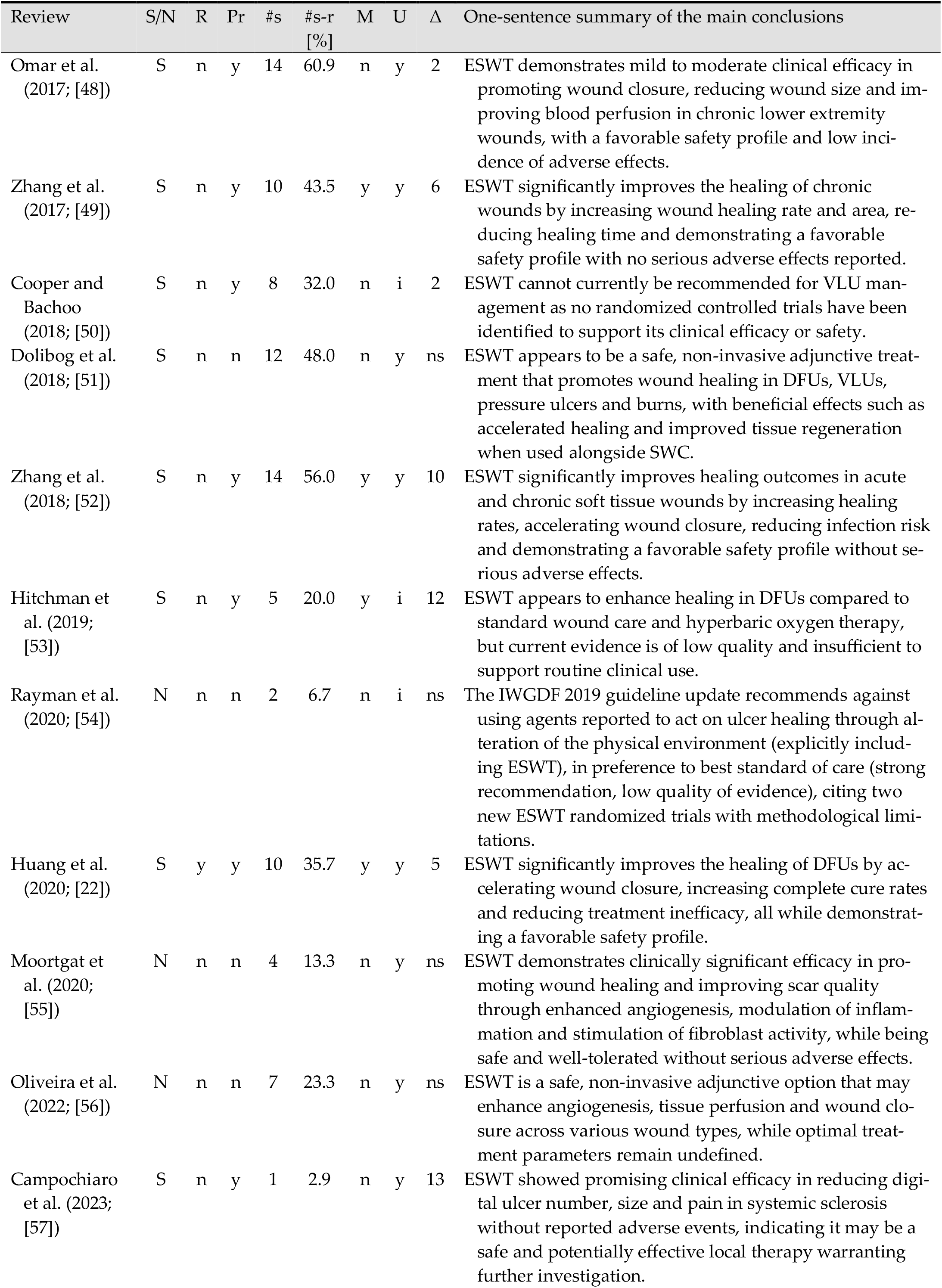
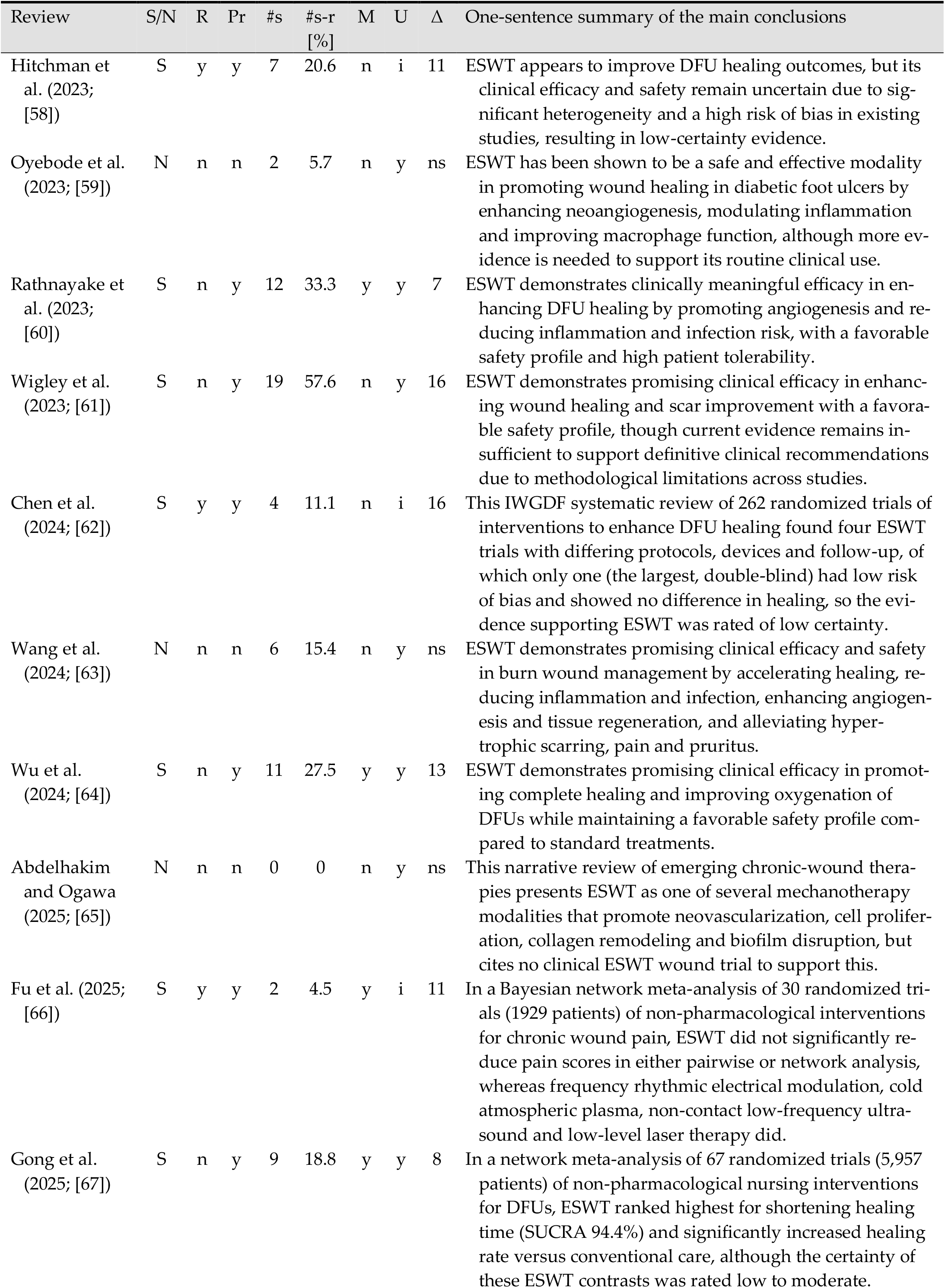
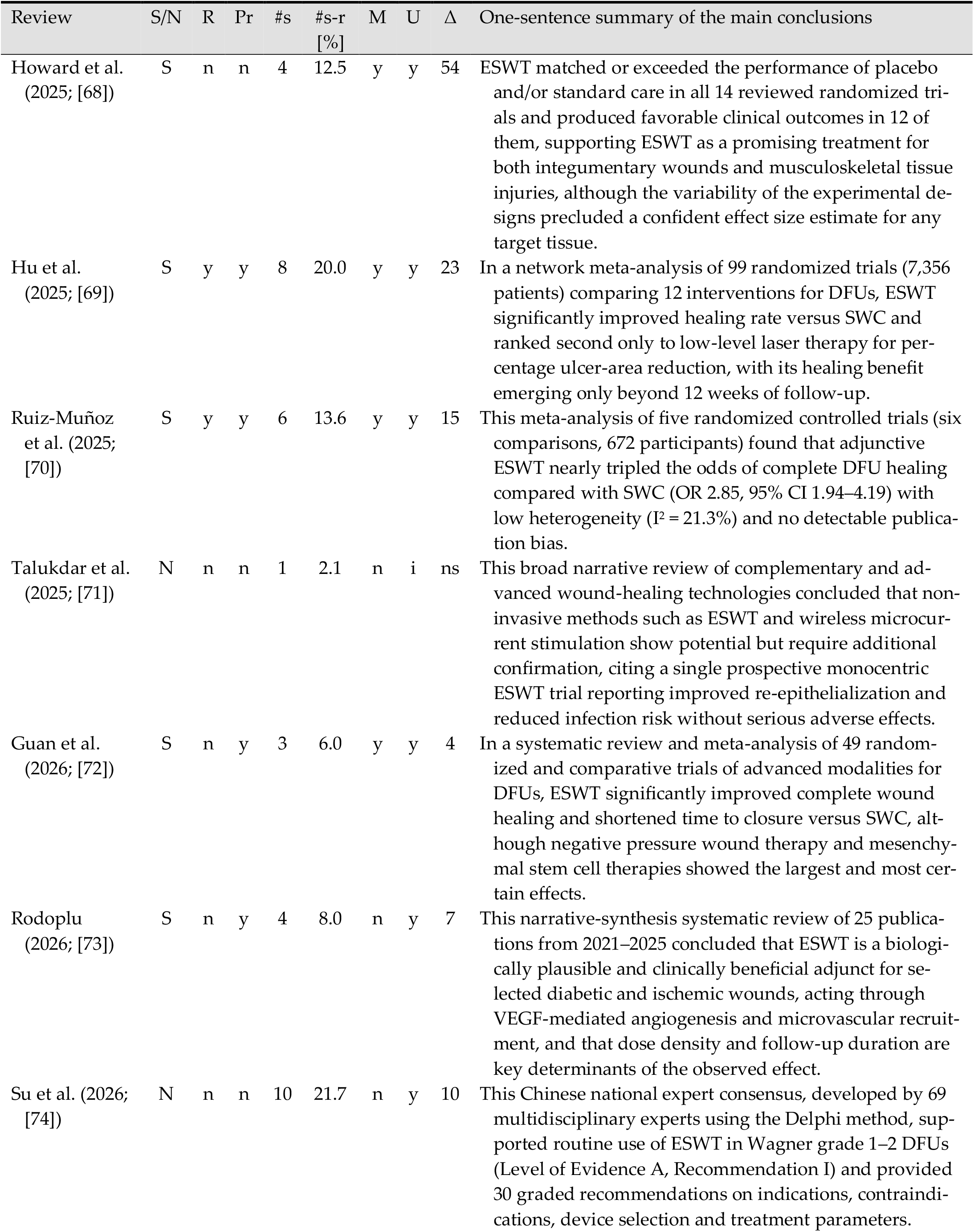
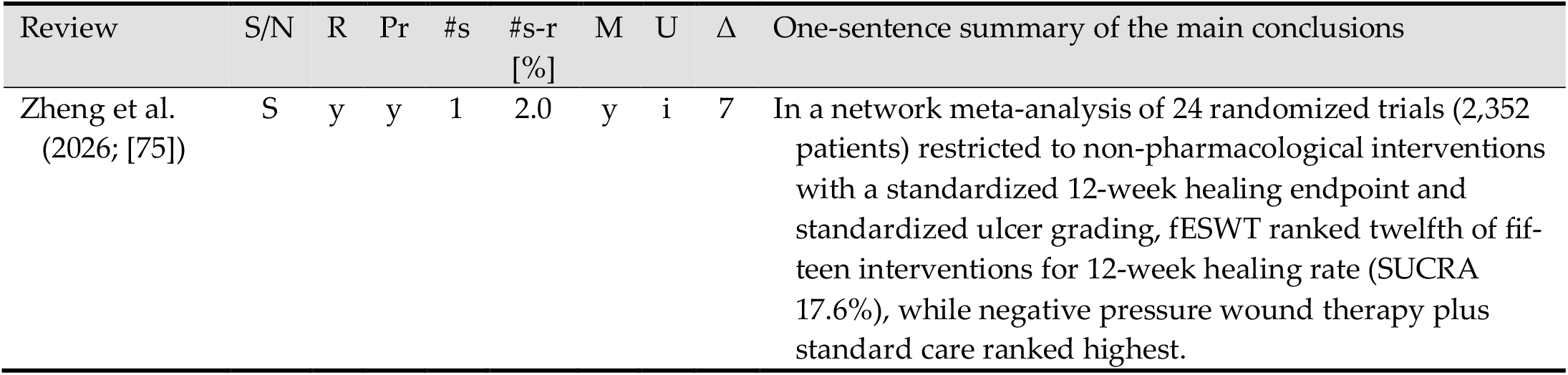
Key characteristics and one-sentence summaries of the main conclusions (each reflecting the conclusions of the authors of the respective review and not those of the present systematic review) of all reviews assessed in this systematic review, listed in chronological order according to the year of publication. Details are provided in the text. Abbreviations (in alphabetical order): CI, confidence interval; DFU(s), diabetic foot ulcer(s); ESWT, extracorporeal shock wave therapy; fESWT, focused ESWT; HBOT, hyperbaric oxygen therapy; i, inconclusive; I2, measure of statistical heterogeneity; IWGDF, International Working Group on the Diabetic Foot; M, meta-analysis of data on ESWT for wound management; N, narrative review; n, no; OR, odds ratio; Pr, review performed according to the PRISMA criteria [21]; R, registration of review with PROSPERO [120]; RCT, randomized controlled trial; S, systematic review; #s, number of clinical studies on ESWT for wound management considered in each review; #s-r, relative number of clinical studies on ESWT for wound management that had been published by the reference date of each review (c.f. Section 2); SUCRA, surface under the cumulative ranking curve; SWC, standard wound care; U, use of ESWT supported; VEGF, vascular endothelial growth factor; VLU(s), venous leg ulcer(s); y, yes; Δ, interval in months between the date to which the literature search of a review was current and the date of its publication (c.f. Table A1 in Appendix A), with “ns” indicating that no such date was reported.

Of the 46 reviews (Table 4), 27 (58.7%) were systematic reviews and 19 (41.3%) were narrative reviews. Seven of 27 systematic reviews (25.9%) [22,58,62,66,69,70,75] were registered with PROSPERO [120]. Twenty-one of 46 reviews (45.7%), corresponding to 21 of 27 systematic reviews (77.8%), reported that they were conducted according to the PRISMA criteria [21]. Supplementary File 1 presents standardized summaries of all reviews.

The number of clinical studies on ESWT for wound management included in each review ranged from 0 [37,41,65] to 19 [61], with a mean ± SD of 5.9 ± 4.5 and a median of 5.5 studies. More importantly, the proportion of available clinical studies included in each review, based on the number of studies published on or before the reference date of the review (c.f. Section 2), varied from 0% [37,41,65] to 90.9% [33], with a mean ± SD of 26.0% ± 23.4% and a median of 19.4%. Table A2 in Appendix A provides a matrix indicating which clinical studies were included in each review.

When stratified by review type, systematic reviews included 29.0% ± 23.1% of the available studies (median = 20.6%), whereas narrative reviews included 21.7% ± 23.6% (median = 15.8%). Thus, none of the 46 reviews provided a comprehensive synthesis of the clinical evidence on ESWT for wound management available at its reference date. For systematic reviews, this limited coverage may be attributable to restrictive inclusion criteria (e.g., inclusion limited to RCTs or studies involving specific patient populations such as individuals over 65 years of age). In narrative reviews, the basis on which the included studies were selected from those available was usually not stated.

A further question concerns not what the reviews included but how current their evidence was when they appeared. Twenty-six of the 46 reviews (56.5%) reported a date to which their literature search was current, whereas the remaining 20 reported neither a search date nor a search period. Among the 26 reviews that did, the interval between that date and publication had a median of 10.5 months (interquartile range 7 to 15 months, range 2 to 54 months, mean ± SD 12.0 ± 10.0 months), and in seven of them it was 15 months or longer. In 23 of these 26 reviews (88.5%), at least one further clinical study on ESWT for wound management had been published by the time the review appeared (median 2 studies, range 0 to 18), so that the evidence available to a reader had grown by a median of 8% (range 0% to 100%) while the review was in press. The longest interval was in the review by Howard et al. (2025; [68]), whose literature search was performed on June 4, 2021 and which appeared in December 2025, 54 months later; 18 of the 51 clinical studies assessed here had been published in the meantime. Table A1 in Appendix A lists these dates for all 46 reviews.

In 14 of 27 systematic reviews (51.9%), no meta-analysis was performed. Some authors explicitly discussed their reasons for not conducting a meta-analysis, whereas others implied such reasons or did not address the issue directly.

Qureshi et al. (2011; [34]) presented an early systematic review of ESWT applications in plastic and reconstructive surgery, and did not discuss the possibility of performing a meta-analysis. Game et al. (2012; [35]) included ESWT among several treatment modalities for DFUs and opted for a narrative synthesis, citing limited and heterogeneous data as barriers to quantitative pooling. Dymarek et al. (2014; [40]) examined ESWT within a broader review of physical therapies for wound management but did not address the potential use of meta-analytic methods. Butterworth et al. (2015; [42]) conducted a review focusing on methodological quality and critical appraisal of DFU studies using ESWT, implicitly suggesting that heterogeneity of outcomes and reporting limited the feasibility of meta-analysis. Vélez-Díaz-Pallarés et al. (2015; [43]) evaluated several adjunctive treatments for pressure ulcers, including ESWT, and did not state a reason for the absence of a meta-analysis. Game et al. (2016; [47]) presented an updated systematic review and explicitly cited the limited number of RCTs, low study quality and inconsistent outcome reporting as reasons for preferring qualitative synthesis over meta-analysis. Omar et al. (2017; [48]) presented study results narratively without directly explaining the absence of meta-analysis, although substantial variability in ESWT protocols and comparator treatments suggests clinical heterogeneity as a likely contributing factor. Cooper and Bachoo (2018; [50]) clearly stated in a Cochrane review that meta-analysis was not feasible because of substantial heterogeneity in study designs, interventions and outcome measures, including differing definitions of wound healing and inconsistent follow-up periods. Dolibog et al. (2018; [51]) reviewed the use of ESWT for soft tissue wounds and summarized clinical studies describing treatment parameters and outcomes across different wound types. The authors presented their findings descriptively and did not attempt a quantitative synthesis, likely reflecting the small number of studies and the heterogeneity of treatment protocols and reported outcomes. Campochiaro et al. (2023; [57]) reviewed therapies for systemic sclerosis and included ESWT, noting the lack of standardized outcome reporting and the limited number of studies, both of which likely discouraged quantitative synthesis. Hitchman et al. (2023; [58]) focused on ESWT for DFUs and reported their findings narratively without stating a rationale for omitting a meta-analysis; the included studies varied considerably. Wigley et al. (2023; [61]) assessed the role of ESWT in aesthetic and reconstructive wound care; although systematic in scope, their review did not address the possibility of meta-analysis and instead provided a descriptive synthesis of outcomes.

In contrast, 13 of 27 systematic reviews (48.1%) included a meta-analysis. Zhang et al. (2017; [49]) conducted a meta-analysis evaluating the efficacy of ESWT for DFUs by pooling data from four RCTs; however, the limited number of included studies and variability in outcome definitions constrained the robustness of the results. In a follow-up review, Zhang et al. (2018; [52]) expanded their earlier work by including 6 studies and examining outcomes such as healing rate and wound size reduction, although the overall number of trials remained limited and the authors acknowledged methodological limitations in the available data. Hitchman et al. (2019; [53]) conducted a focused meta-analysis comparing ESWT with SWC in vascular wound healing. Although several RCTs were included, the analysis was limited to a small number of outcomes, because of the limited data available across studies. Huang et al. (2020; [22]) performed a meta-analysis of eight RCTs (n = 339) examining ESWT for DFUs and reported pooled effects on wound surface area, re-epithelialization and complete healing. Despite a structured analytical approach, only a limited set of outcomes could be analyzed because of inconsistent reporting across trials. Rathnayake et al. (2023; [60]) included nine studies and primarily evaluated wound healing rate and healing time; however, relatively few studies met the inclusion criteria and the number of analyzable outcomes remained limited, restricting the generalizability of the conclusions. Wu et al. (2024; [64] included ten RCTs and examined outcomes such as complete healing, unchanged ulcers, transcutaneous oxygen pressure (TcPO_2_) levels and adverse events. Although this analysis incorporated a broader set of outcomes than earlier studies, the authors nevertheless noted limitations related to the small number of high-quality studies and variability in outcome definitions. Fu et al. (2025; [66]) and Zheng et al. (2026; [75]) performed a Bayesian and a frequentist network meta-analysis, respectively, in which ESWT was one node among a larger set of non-pharmacological interventions, and Gong et al. (2025; [67]) and Hu et al. (2025; [69]) employed the same approach for nursing interventions and for 12 treatments of DFUs. Ruiz-Muñoz et al. (2025; [70]) pooled six trials of ESWT for DFUs in a random-effects model with formal tests for small-study effects, and Guan et al. (2026; [72]) pooled three trials within a broader comparison of advanced therapies. Howard et al. (2025; [68]) pooled pain intensity, pressure pain threshold and time to re-epithelialization across randomized trials of ESWT in wounds and in musculoskeletal tissue injuries; each of the wound-related estimates rested on two or three trials, and the certainty of the evidence was rated moderate at best. In each of these analyses ESWT was represented by between one and nine trials, so the ranking statistics reported for it rest on the evidence base described above. Hu et al. [69] additionally stratified by follow-up duration and found the advantage of ESWT in healing rate only beyond 12 weeks.

Therefore, both types of systematic reviews (those with and those without meta-analyses) highlight substantial limitations that complicate the performance of a robust meta-analysis of ESWT for wound healing. Reviews without meta-analysis frequently cited or implied barriers such as clinical and methodological heterogeneity, inconsistent definitions of healing, variability in ESWT protocols, diverse comparator interventions and incomplete outcome reporting. Reviews that attempted meta-analyses generally included relatively few heterogeneous RCTs, often pooling only one or two outcomes, and acknowledged issues such as low trial quality and limited generalizability.

Twenty-seven of 46 reviews (58.7%) supported the use of ESWT for wound management, whereas 19 of 46 reviews (41.3%) were inconclusive regarding its effectiveness. In summary, across narrative reviews the prevailing interpretation was cautiously optimistic. ESWT was frequently described as a promising adjunctive therapy for wound healing, with several case series and small clinical studies suggesting improved healing outcomes, although authors consistently emphasized the need for more rigorous trials, particularly RCTs. Systematic reviews without meta-analysis likewise presented a generally positive but cautious perspective, acknowledging potential benefits while highlighting methodological limitations, heterogeneity and insufficient high-quality evidence as barriers to firm conclusions. Systematic reviews that included meta-analyses reported statistically significant effects in favor of ESWT for outcomes such as healing rate and tissue oxygenation; however, these analyses uniformly noted limitations related to small and heterogeneous data sets and the resulting low certainty of evidence. Overall, while the review literature suggests that ESWT may provide therapeutic benefit, the collective evidence remains inconclusive and insufficient to support routine clinical implementation without further high-quality research.

Two of the most recent reviews addressed the questions that motivated the present synthesis more directly than their predecessors. Su et al. (2026; [74]), in a national expert consensus developed by the Delphi method, distinguished between the modalities in its recommendations, reserving fESWT for deep tissue involvement and favoring rESWT for superficial defects, and specified energy, impulse number and treatment frequency. Rodoplu (2026; [73]) classified the included publications by protocol class, and related impulse density to healing time. Both therefore treated ESWT as a family of interventions rather than as a single one. Neither, however, compared the modalities at a dose matched on a physical scale, and both rested on the primary trials assessed here; in [73], the 25 included publications are not listed individually, so the underlying study set cannot be reconstructed.

### 3.2. Assessment of Clinical Studies of ESWT for Wound Management

Tables 5–9 summarize the key characteristics and main findings (in a single sentence) of each clinical study on fESWT, unfocused ESWT (uESWT) and rESWT for wound management identified in this systematic review, categorized by device type: electrohydraulic (Table 5), electromagnetic (Table 6), piezoelectric (Table 7), radial (Table 8) and undisclosed devices (Table 9). The manufacturers of the devices listed in Tables 5-8 are provided in Table 10. Note: some records reported the same participants; such records are listed separately in these tables for completeness, but each group of overlapping publications was counted once in the quantitative analyses; the six groups concerned are identified in Section 3.3.4.

**Table 5.** Key characteristics and one-sentence summaries of the main findings of all clinical studies identified in this systematic review that were conducted using electrohydraulic ESWT devices (listed in chronological order according to the year of publication). Supplementary File 2 provides standardized summaries of these studies, including the extraction of 40 predefined variables from each study (see Table 2 for details). Abbreviations (in alphabetical order): ASEPSIS, scoring system for postoperative wound infection; CS, case series (n>1); CO, cohort study with non-randomized controls; CR, case report (n=1); DFUs, diabetic foot ulcers; ESWs, extracorporeal shock waves; ESWT, extracorporeal shock wave therapy; fESWT, focused ESWT; HBOT, hyperbaric oxygen therapy; NC, number of patients in the control group; NE, number of patients in the ESWT group; O, overall outcome (as defined in Table 3); RCT, randomized controlled trial; SWC, standard wound care; T, type of study; uESWT, unfocused ESWT; VLUs, venous leg ulcers. Notes: ^a^, all patients received both treatments in crossover design; ^b^, 67 patients (72 ulcers), subdivided into diabetic ulcers (38 patients / 40 ulcers) and nondiabetic ulcers (29 patients / 32 ulcers).

| Study | Device | T | NE | NC | O | One-sentence summary of the main findings |
| --- | --- | --- | --- | --- | --- | --- |
| Meirer et al. (2005; [76]) | Evotron | CR | 1 | 0 | 5 | This case report described the treatment of deep partial thickness burns in one patient, resulting in near-complete wound re-epithelialization by day 15 and full healing without scarring at six months. |
| Schaden et al. (2007; [77]) | DermaGold | CS | 208 | 0 | 5 | In this prospective case series, 208 patients with acute and chronic soft tissue wounds – including postoperative dehiscence, posttraumatic necrosis, venous and arterial ulcers, pressure ulcers and burns – were treated, resulting in complete wound healing (100% epithelialization) in 156 patients (75%) without any reported adverse events. |
| Dumfarth et al. (2008; [78]) | DermaGold | RCT | 50 | 50 | 1 | In this RCT, acute surgical wounds from saphenous vein harvesting were treated in 100 patients, showing significantly improved healing in the 50 patients receiving ESWT, evidenced by lower ASEPSIS scores ($4.4 \pm 5.3$ vs. $11.6 \pm 8.3$, $p < 0.001$) and reduced need for antibiotics (4% vs. 22%, $p = 0.015$), compared to 50 control patients. |
| Saggini et al. (2008; [79]) | Evotron | CO | 30 | 10 | 1 | This study treated 32 chronic lower extremity ulcers, including 16 posttraumatic, 12 venous and 4 DFUs, in 30 patients with ESWT and compared outcomes to 10 chronic ulcers in a control group receiving SWC, finding that 16 treated ulcers (50%) healed completely with significant improvements in wound parameters and pain, while only 1 control ulcer healed and no significant improvements were observed in the control group. |
| Wang et al. (2009; [23]) | Orthowave | RCT | 34 | 36 | 1 | In this RCT, 34 patients with 36 chronic DFUs treated with ESWT showed significantly better healing outcomes (31% completely healed and 57% improved) compared to 36 patients with 36 ulcers in the control group receiving HBOT, where 22% completely healed and 50% improved. |
| Arnó et al. (2010; [80]) | dermaPACE | CS | 15 | 0 | 5 | Deep partial- and full-thickness burns in 15 patients were treated in a prospective observational study, with 12 out of 14 evaluable patients (86%) achieving spontaneous healing without surgery and no adverse events reported. |
| Larking et al. (2010; [81]) | Orthowave 180c (unfocused) | RCT | 8<sup>a</sup> | 8<sup>a</sup> | 1 | In this RCT, nine chronic decubitus ulcers in eight patients showed significantly greater healing following ESWT compared to placebo, with improvements observed 6–8 weeks after treatment initiation. |
| Ottomann et al. (2010; [82]) | DermaGold | RCT | 13 | 15 | 1 | In this RCT, 28 patients with acute traumatic wounds and burns requiring split-thickness skin grafts were treated; those who received ESWT (n = 13) showed significantly faster donor site healing ($13.9 \pm 2.0$ days) compared to the control group (n = 15) receiving SWC alone ($16.7 \pm 2.0$ days; $p < 0.001$). |
| Wang et al. (2011; [30]) | dermaPACE | RCT | 39 | 38 | 1 | In this RCT, 39 patients with 44 chronic DFUs treated with ESWT showed a significantly higher rate of complete healing (57%) compared to 38 patients with 40 ulcers treated with HBOT (25%, $p = 0.003$). |
| Wolff et al. (2011; [83]) | Orthowave | CS | 282 | 0 | 5 | Chronic soft tissue wounds, including post-surgical, post-traumatic, venous, arterial and decubitus ulcers as well as burns, were treated in 258 patients, of whom 191 (74%) achieved complete wound closure or significant improvement following ESWT in this one-armed prospective cohort study. |
| Fioramonti et al. (2012; [84]) | Evotron | CR | 1 | 1 | 5 | Chronic VLUs in one patient were treated on one leg with ESWT, resulting in complete healing after six weeks, while a control ulcer on the contralateral leg treated with SWC showed incomplete healing. |
| Ottomann et al. (2012; [85]) | DermaGold | RCT | 22 | 22 | 1 | In this RCT, 44 patients with superficial second-degree burns were treated; those receiving ESWT (n=22) achieved complete wound epithelialization significantly faster ($9.6 \pm 1.7$ days) than those receiving SWC alone (n=22; $12.5 \pm 2.2$ days; $p < 0.001$). |
| Saggini et al. (2013; [86]) | DermaGold / Evotron | RCT | 60 | 40 | 2a | This RCT compared two uESWT regimens in 124 patients with chronic ulcers of diabetic, pressure-related and traumatic etiology, without a non-ESWT comparator: 62 patients received uESWT at 0.10 mJ/mm<sup>2</sup> (DermaGold) and 62 at 0.04 mJ/mm<sup>2</sup> (Evotron). Wound area fell by 80% and pain by 79% in the higher-energy arm, compared with 67% and 48% in the lower-energy arm; 22 of the 62 patients in the lower-energy arm withdrew, 15 of them because of pain, so 62 and 40 patients respectively contributed outcome data. Because there was no non-ESWT comparator, this trial is informative about dose rather than about efficacy relative to SWC. |
| Wang et al. (2014; [87]) | dermaPACE | CS | 67<sup>b</sup> | 0 | 5 | Chronic foot ulcers of diabetic and nondiabetic origin were treated in 67 patients with 72 ulcers, resulting in complete healing in 55.6% and ≥50% improvement in 27.8% of cases at 1 year, with sustained healing in 57.4% at 5 years, demonstrating better outcomes in nondiabetic patients. |
| Aschermann et al. (2017; [88]) | CellSonic | CS | 60 | 0 | 5 | In this retrospective analysis of consecutive routine-care patients, 75 chronic therapy-refractory leg ulcers of various etiologies in 60 patients were treated, with complete healing in 41% of ulcers, a size reduction of more than 75% in 16%, a reduction of 20-75% in 35% and no change in 8%. |
| Snyder et al. (2018; [28]: analysis of pooled data) | dermaPACE | RCT | 172 | 164 | 1 | In this RCT, 336 patients with chronic DFUs were treated; significantly more wounds achieved complete closure at 20 weeks (35.5% vs. 24.4%) and 24 weeks (37.8% vs. 26.2%) in the treatment group compared to the sham control group. |
| Snyder et al. (2018; [28]: first RCT) | dermaPACE | RCT | 107 | 99 | 1 | In this RCT of 206 patients with chronic DFUs, significantly more ulcers achieved complete closure in the treatment group than in the sham group at 20 weeks (36.4% vs. 23.2%) and at 24 weeks (39.3% vs. 26.3%). |
| Snyder et al. (2018; [28]: second RCT) | dermaPACE | RCT | 65 | 65 | 2b | In this RCT of 130 patients with chronic DFUs, wound closure rates were higher in the treatment group than in the sham group at both 12 weeks (26.2% vs. 23.1%) and 24 weeks (35.4% vs. 26.2%), though these differences were not statistically significant. |
| Chou et al. (2019; [89]) | dermaPACE | CS | 4 | 0 | 5 | Chronic DFUs were treated in four patients, resulting in complete healing in two patients and partial improvement in the other two at 48 weeks following extended ESWT. |
| Galiano et al. (2019; [90]) | dermaPACE | RCT | 172 | 164 | 1 | This report of the secondary endpoints of the same two trials [28] found significantly greater wound area reduction (48.6% vs. 10.7%; $p = 0.015$) and perimeter reduction (46.4% vs. 25.0%; $p = 0.022$) in the ESWT group at 12 weeks. Complete closure in the pooled dataset was 22.7% vs. 18.3% at 12 weeks ($p = 0.320$), reaching significance at 20 weeks (35.5% vs. 24.4%; $p = 0.027$) and 24 weeks (37.8% vs. 26.2%; $p = 0.023$). |
| Holsapple et al. (2021; [91]) | DermaGold | CS | 9 | 0 | 5 | In 9 patients with chronic VLUs unresponsive to conventional treatment, ESWT led to improved healing in 7 patients, as evidenced by reduced wound area and increased angiogenesis |
| Jeong et al. (2023; [92]) | orthoPACE | CO | 25 | 18 | 1 | In this controlled cohort study, 25 patients with DFUs showed significantly improved microcirculation in the 32 ulcerated feet treated with ESWT compared to the 18 untreated contralateral feet, as evidenced by a statistically significant increase in transcutaneous partial oxygen pressure after two weeks of treatment. |
| Nemeth and Shah (2024; [93]) | dermaPACE | CS | 10 | 0 | 5 | Thirteen chronic wounds in 10 patients (primarily DFUs and postsurgical wounds) were treated with ESWT, resulting in complete healing in 12 cases within 12 weeks. |
| Marzella et al. (2026; [94]) | OrthoGold | CS | 64 | 0 | 5 | In this prospective, non-comparative case series, 64 patients with soft-tissue defects of the hand (mean baseline wound area $2.4 \pm 1.1$ cm<sup>2</sup>, treated a mean of 6.2 weeks after injury) received fESWT at 0.10 mJ/mm<sup>2</sup> and 350–1000 ESWs/cm<sup>2</sup> once weekly for a mean of $4.3 \pm 1.2$ sessions in addition to SWC, resulting in complete wound closure after a mean of $4.5 \pm 1.1$ weeks together with significant improvement from baseline in the Bates–Jensen Wound Assessment Tool score, pain and capillaroscopic microvascular density (all $p < 0.01$); no adverse events were reported. |

**Table 6.**
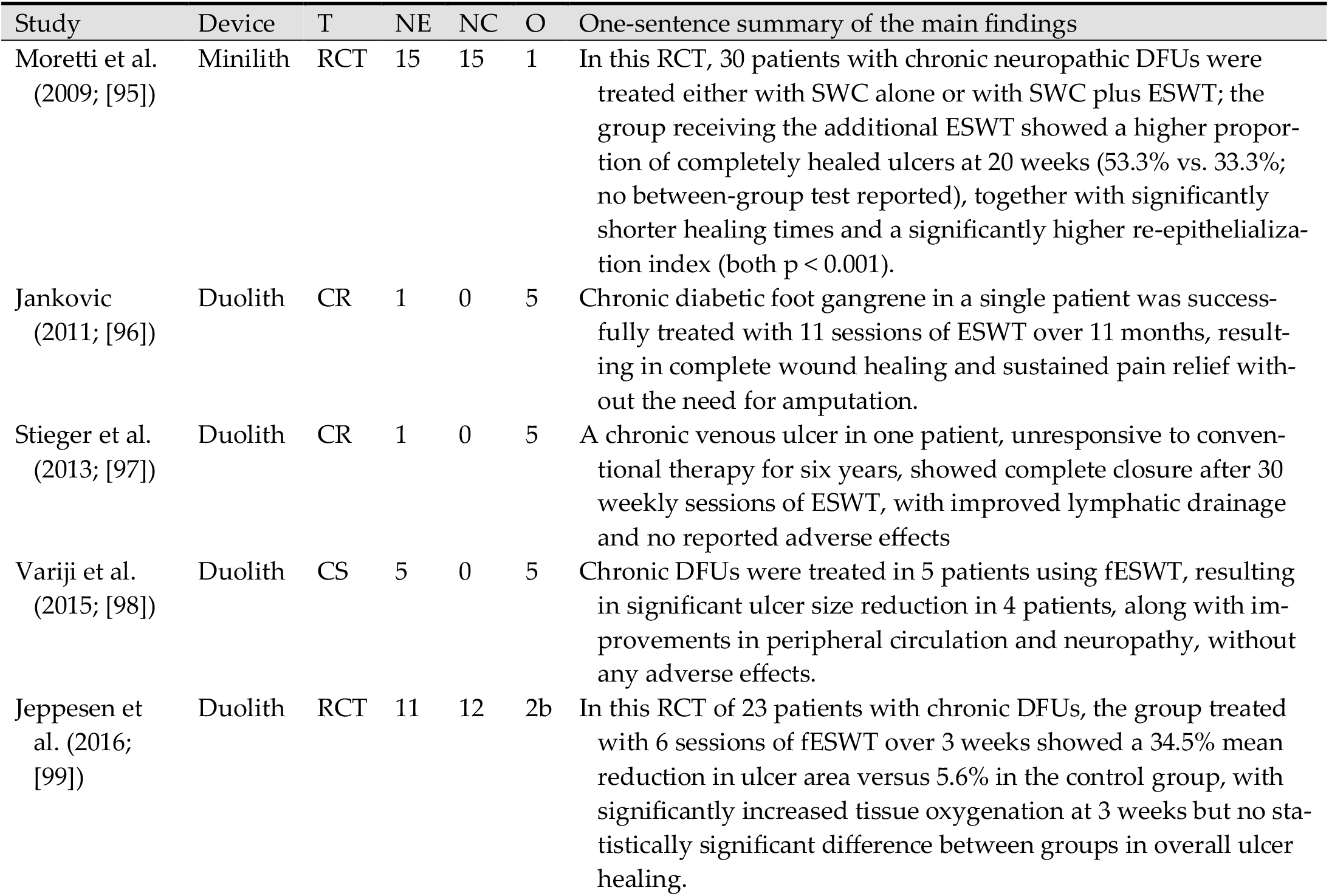
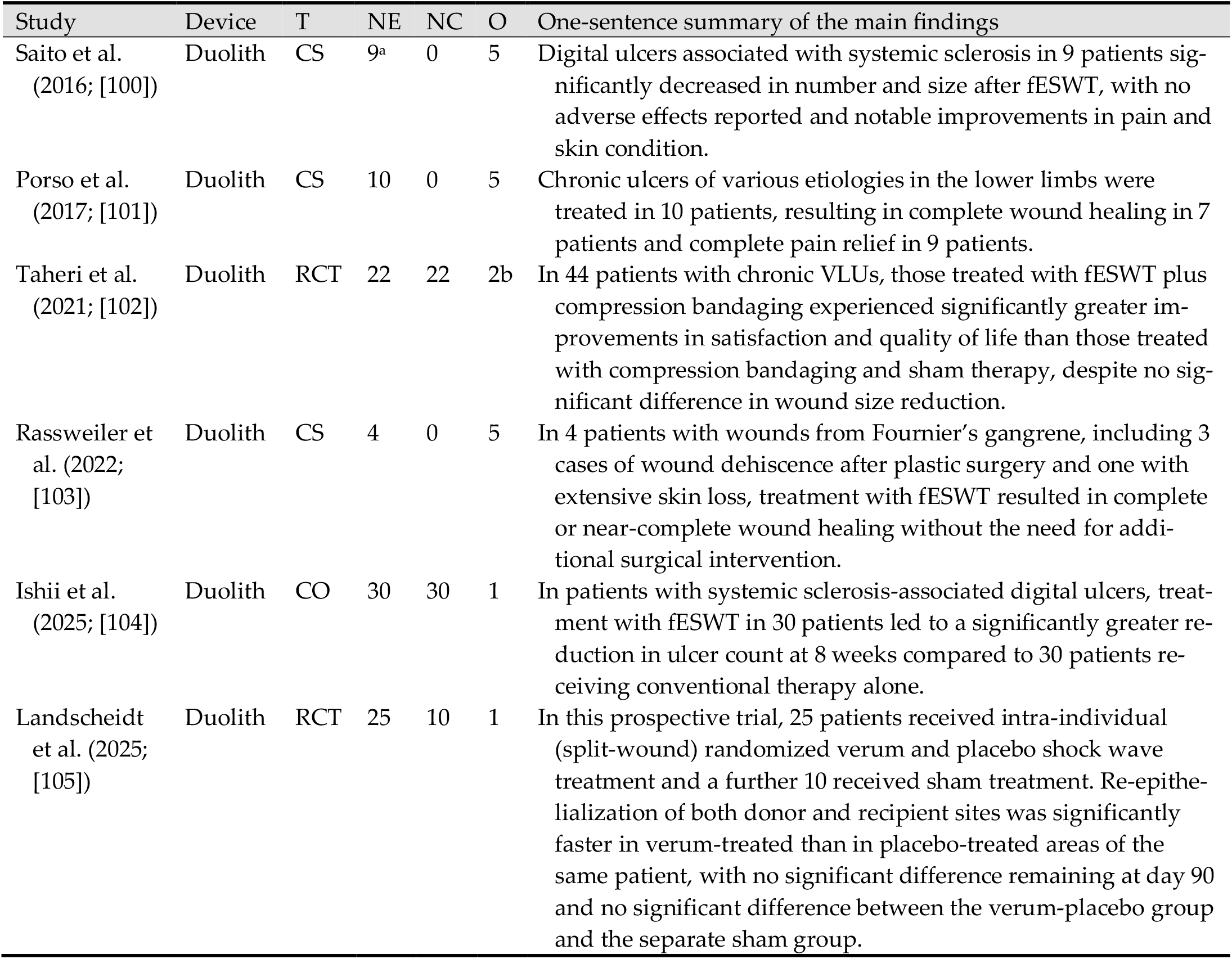
Key characteristics and one-sentence summaries of the main findings of all clinical studies identified in this systematic review that were conducted using electromagnetic ESWT devices (listed in chronological order according to the year of publication). Supplementary File 3 provides standardized summaries of these studies, including the extraction of 40 predefined variables from each study (see Table 2 for details). Abbreviations (in alphabetical order): CS, case series (n>1); CO, cohort study with non-randomized controls; CR, case report (n=1); DFUs, diabetic foot ulcers; ESWT, extracorporeal shock wave therapy; fESWT, focused ESWT; NC, number of patients in the control group; NE, number of patients in the ESWT group; O, overall outcome (as defined in Table 3); RCT, randomized controlled trial; SWC, standard wound care; T, type of study; VLUs, venous leg ulcers. Notes: ^a^, 9 patients with 49 digital ulcers.

**Table 7.**
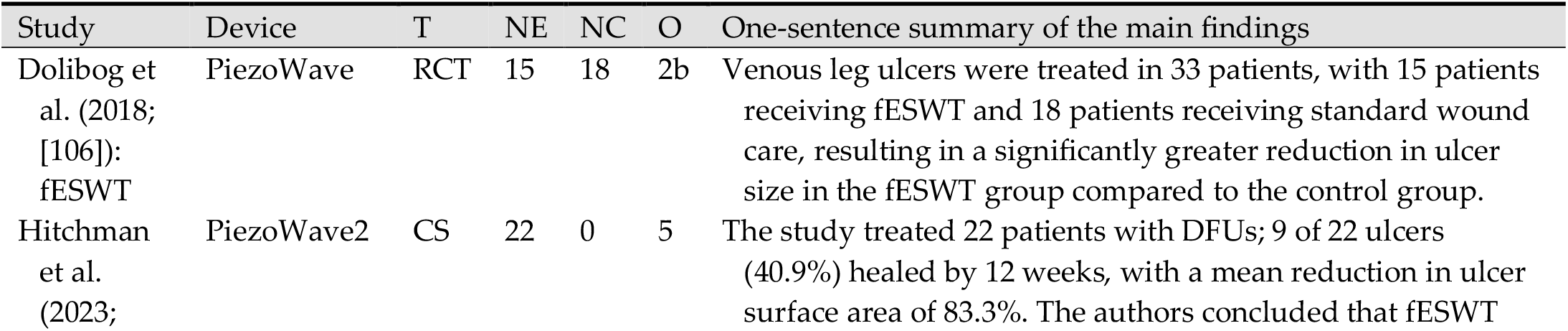
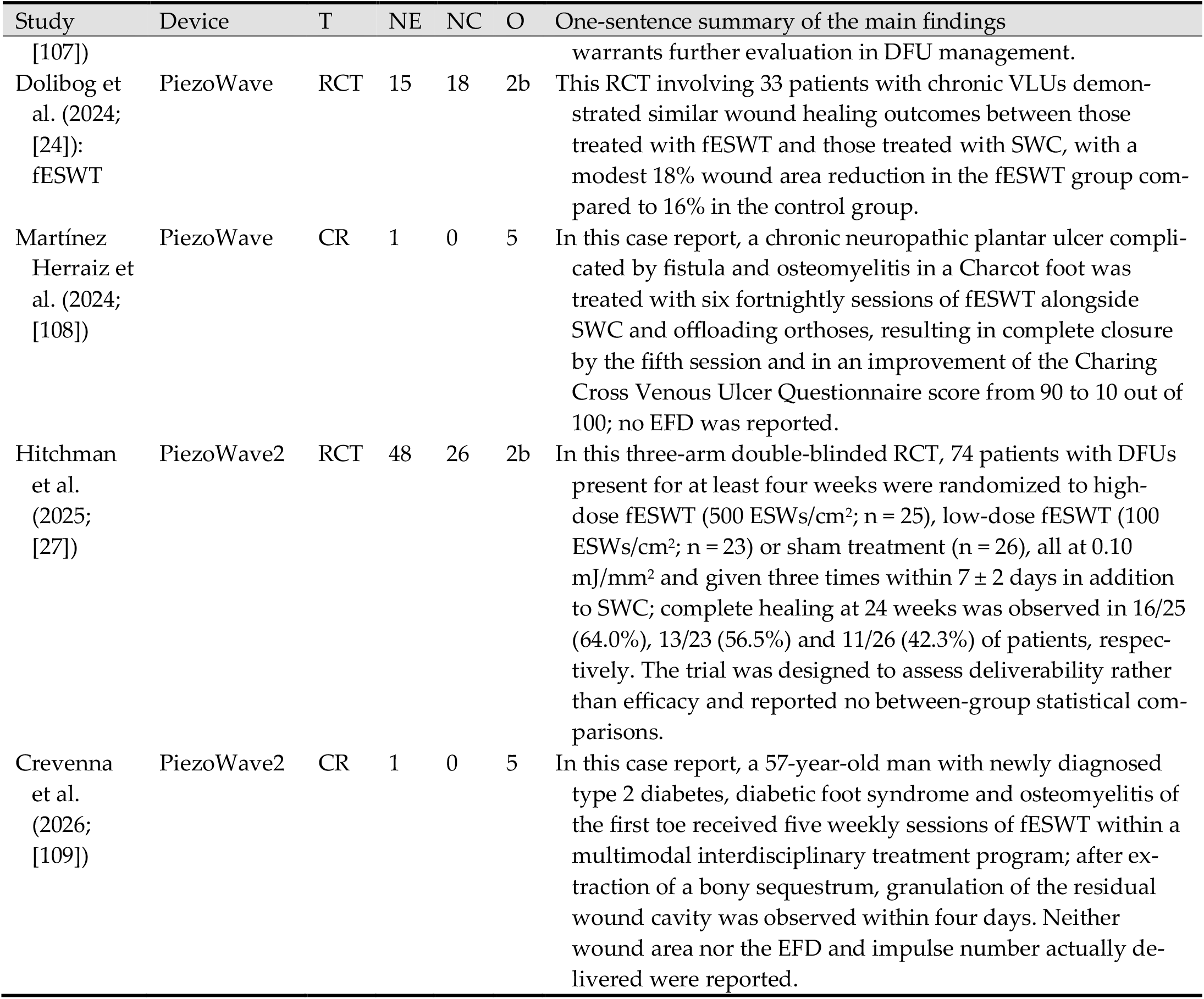
Key characteristics and one-sentence summaries of the main findings of all clinical studies identified in this systematic review that were conducted using piezoelectric ESWT devices (listed in chronological order according to the year of publication). Supplementary File 4 provides standardized summaries of these studies, including the extraction of 40 predefined variables from each study (see Table 2 for details). Abbreviations (in alphabetical order): CS, case series (n>1); CO, cohort study with non-randomized controls; CR, case report (n=1); DFU(s), diabetic foot ulcer(s); EFD, energy flux density; ESWs, extracorporeal shock waves; ESWT, extracorporeal shock wave therapy; fESWT, focused ESWT; NC, number of patients in the control group; NE, number of patients in the ESWT group; O, overall outcome (as defined in Table 3); RCT, randomized controlled trial; SWC, standard wound care; T, type of study; VLUs, venous leg ulcers. Notes: ^a^, the abstract of this paper reported 45.5% healing, which does not agree with the figures given in its Results section.

**Table 8.** Key characteristics and one-sentence summaries of the main findings of all clinical studies identified in this systematic review that were conducted using radial ESWT devices (listed in chronological order according to the year of publication). Supplementary File 5 provides standardized summaries of these studies, including the extraction of 40 predefined variables from each study (see Table 2 for details). Abbreviations (in alphabetical order): CS, case series (n>1); CO, cohort study with non-randomized controls; CR, case report (n=1); DFUs, diabetic foot ulcers; ESWT, extracorporeal shock wave therapy; NC, number of patients in the control group; NE, number of patients in the ESWT group; O, overall outcome (as defined in Table 3); rESWT, radial ESWT; RCT, randomized controlled trial; SWC, standard wound care; T, type of study; VLUs, venous leg ulcers. Notes: ^a^, radial ESWT (17 patients) vs ultrasound therapy (17 patients) and SWC only (17 patients); ^b^, this report describes neither randomization nor allocation concealment nor blinding, and reports that 46 of 87 consecutively enrolled patients received ESWT and the remaining 41 routine treatment; the study is therefore classified here as a controlled study without randomization.

| Study | Device | T | NE | NC | O | One-sentence summary of the main findings |
| --- | --- | --- | --- | --- | --- | --- |
| Nossair et al. (2013; [110]) | BTL-5000 | RCT | 20 | 20 | 1 | This RCT treated 40 patients with DFUs, finding that rESWT significantly reduced wound surface area and increased epithelialization compared to SWC, with a notable improvement in both outcomes in the treatment group. |
| Fekete et al. (2014; [111]) | Dolor-Clast | CR | 1 | 0 | 5 | This case report involved a single patient with a large venous ulcer who was treated with rESWT, resulting in significant wound size reduction and the preparation of the wound for surgical closure. |
| Dolibog et al. (2018; [106]): rESWT | Shock-Master | RCT | 17 | 18 | 1 | Venous leg ulcers were treated in 35 patients, with 17 patients receiving rESWT and 18 patients receiving standard wound care, resulting in a significantly greater reduction in ulcer size in the rESWT group compared to the control group. |
| Dolibog et al. (2018; [112]) | Shock-Master | RCT | 17/17<sup>a</sup> | 17 | 4 | This study treated 51 patients with VLUs and found that both rESWT (38.2% reduction) and ultrasound therapy (67.6% reduction) significantly outperformed SWC (15.8% reduction) in reducing ulcer area after 4 weeks of treatment. |
| Duan et al. (2020; [113]) | Dolor-Clast | CR | 1 | 0 | 5 | This case report demonstrated that rESWT combined with alginate dressing successfully healed the pressure ulcer of one elderly patient after 12 weeks of treatment, with the ulcer healing fully. |
| Kang et al. (2020; [114]) | Storz Medical radial device | CR | 1 | 0 | 5 | This case report treated a decubitus ulcer in a single patient with a spinal cord injury, achieving complete wound healing after three months of rESWT. |
| Fan et al. (2022; [25]) | Dolor-Clast | CO<sup>b</sup> | 46 | 41 | 1 | This RCT demonstrated that in 87 patients with recurrent DFUs, the group receiving rESWT showed significantly better wound healing, faster recovery times, fewer adverse reactions and improved life quality compared to the control group receiving routine treatment. |
| Dymarek et al. (2023; [115]) | Cellactor | RCT | 20 | 20 | 1 | Pressure ulcers were treated in 40 patients, with a significant improvement in wound size and clinical scores in the rESWT group compared to the placebo group, supporting the potential effectiveness of rESWT for pressure ulcer healing. |
| Dolibog et al. (2024; [24]): rESWT | Shock-Master | RCT | 16 | 18 | 2b | This randomized controlled trial involving 34 patients with chronic venous leg ulcers demonstrated a significantly greater reduction in wound area (31.6%) in the rESWT compared to a 16% reduction in the control group receiving SWC. |
| Dymarek et al. (2024; [116]) | Cellactor | CS | 19 | 0 | 5 | This study treated 31 chronic wounds (22 pressure ulcers, 7 venous leg ulcers and 2 DFUs) in 19 patients, demonstrating significant reductions in wound area and clinical improvements, as measured by the Wound Bed Score and Bates-Jansen Wound Assessment Tool. |
| Sopel et al. (2024; [117]) | Cellactor | CS | 10 | 0 | 5 | In a case series involving 10 patients with pressure ulcers, two sessions of rESWT led to significant improvements in wound size, clinical score, proliferation index and microvascular density, supporting its potential as an effective treatment for pressure ulcers. |
| Yang et al. (2025; [118]) | Dolor-Clast | RCT | 20 | 20 | 1 | This RCT demonstrated that rESWT significantly improved wound healing, pain relief and scar outcomes in 40 patients with deep dermal burns of the hand, with the rESWT group showing a higher median percentage reduction in wound area after 14 days (78%) than the sham-treated routine dressing group (33%). |

**Table 9.** Key characteristics and one-sentence summaries of the main findings of all clinical studies identified in this systematic review that were conducted using ESWT devices whose technical modality was not disclosed (listed in chronological order according to the year of publication). Supplementary File 6 provides standardized summaries of these studies, including the extraction of 40 predefined variables from each study (see Table 2 for details). Abbreviations (in alphabetical order): CS, case series (n>1); CO, cohort study with non-randomized controls; CR, case report (n=1); DFUs, diabetic foot ulcers; ESWT, extracorporeal shock wave therapy; NC, number of patients in the control group; NE, number of patients in the ESWT group; O, overall outcome (as defined in Table 3); RCT, randomized controlled trial; SWC, standard wound care; T, type of study; VLUs, venous leg ulcers.

| Study | Device | M | T | NE | NC | O | One-sentence summary of the main findings |
| --- | --- | --- | --- | --- | --- | --- | --- |
| Omar et al. (2014; [29]) | Not reported | U | RCT | 19 | 19 | 1 | Chronic DFUs in 38 patients (45 ulcers) were treated with either ESWT or SWC; the ESWT group showed significantly better outcomes, with 54% complete healing compared to 28.5% in the control group. |
| Vangaveti et al. (2023; [119]) | Not reported | U | RCT | 25 | 23 | 2b | This RCT involved 48 patients with DFUs, with 25 receiving ESWT plus standard wound care and 23 receiving only SWC; no significant difference in wound healing was found between the two groups after 6 weeks. |

**Table 10.** Manufacturers of the ESWT devices mentioned in Tables 5-8. Abbreviation: n, number of studies in which the respective device was used.

| Device | n | Manufacturer |
| --- | --- | --- |
| Duolith SD1 | 10 | Storz Medical, Tägerwil, Switzerland |
| dermaPACE | 7 | Sanuwave Health, Eden Prairie, MN, USA |
| DermaGold | 6 | Tissue Regeneration Technologies (TRT), Alpharetta, GA, USA |
| DolorClast | 4 | Electro Medical Systems (EMS), Nyon, Switzerland |
| Evotron | 4 | High Medical Technologies (HMT), Lengwil, Switzerland |
| Orthowave | 3 | MTS Medical, Konstanz, Germany |
| ShockMaster | 3 | Gymna Uniphy, Bilzen-Hoeselt, Belgium |
| Cellactor | 3 | Storz Medical, Tägerwil, Switzerland |
| PiezoWave | 3 | Richard Wolf, Knittlingen, Germany |
| BTL-5000 | 1 | BTL, Prague, Czech Republic |
| CellSonic | 1 | CellSonic, Las Vegas, NV, USA |
| Minilith SL1 | 1 | Storz Medical, Tägerwil, Switzerland |
| orthoPACE | 1 | Sanuwave Health, Eden Prairie, MN, USA |
| OrthoGold 100 | 1 | MTS Medical, Konstanz, Germany |
| PiezoWave2 | 3 | Richard Wolf, Knittlingen, Germany |
| Storz Medical radial device | 1 | Storz Medical, Tägerwil, Switzerland |
| Name of device not mentioned | 2 |  |

Tables 5–9 and Supplementary Files 2-6 show that the clinical literature on ESWT for wound management varies widely in wound type, treatment protocol, dosing, endpoints and methodological quality. All subsequent assessments refer to the 51 clinical studies listed in Tables 5–9; the variables V1 to V40 are summarized in Table 2.

#### 3.2.1. Wound Type (V1)

The wound types investigated across the studies encompassed a wide range of acute and chronic conditions, including DFUs, VLUs, PUs, posttraumatic wounds, postsurgical wounds and burns. Diabetic foot ulcers were by far the most frequently studied, reflecting both their prevalence and their clinical complexity, particularly in patients with neuropathy, ischemia or both (e.g., [30,95]). Other studies addressed conditions that are particularly difficult to treat, including wounds following surgical debridement for Fournier’s gangrene [103] and digital ulcers in systemic sclerosis (e.g., [100]). Acute wounds included burns (e.g., [80,118]), surgical wounds following saphenous vein harvesting for coronary artery bypass grafting [78] and skin graft donor sites (e.g., [82,105]) and acute soft-tissue defects of the hand following crush, sharp or burn injury [94].

#### 3.2.2. Wound Size (V2)

Wound size varied widely across studies, ranging from small superficial ulcers to large chronic wounds exceeding 20 cm^2^. Many studies applied a minimum wound size of 1 cm^2^, with several enrolling ulcers with mean baseline areas of approximately 9-12 cm^2^ to allow evaluation of measurable treatment effects of ESWT (e.g., [28,112]). A few studies specifically emphasized large or refractory wounds, particularly in cases involving DFUs or VLUs with prolonged healing times (e.g., [83,91]).

#### 3.2.3. Duration of Wounds (V3)

Most studies targeted chronic wounds, typically defined as those persisting for more than 4 to 6 weeks, with some wounds present for several months or even years. Inclusion criteria often required a minimum wound duration of one month or longer to ensure chronicity (e.g., [99,104]). Several studies specifically highlighted long-standing, non-healing wounds, such as DFUs persisting for more than six months or recurrent VLUs, as potentially responsive to ESWT (e.g., [77,87]).

#### 3.2.4. Study Design (V4)

The studies included a broad spectrum of designs, ranging from single-patient case reports and small case series to large RCTs. While RCTs represent the highest level of evidence and have become more common in recent publications (e.g., [28,118]), numerous case series and cohort studies provided valuable real-world clinical data, particularly for rare or complex wounds (e.g., [104,117]). Case reports were especially common in early or exploratory applications of ESWT (e.g., [76,96]).

#### 3.2.5. Overall Outcome (V5)

Most studies reported beneficial effects on wound healing, including accelerated epithelialization, reduced wound size, improved tissue perfusion and enhanced granulation tissue formation. Complete or near-complete healing was achieved in a substantial proportion of cases in both observational studies and RCTs (e.g., [77,95]). Some studies also reported secondary benefits such as pain reduction or decreased infection rates (e.g., [104]). The magnitude of the reported effect varied with ESWT modality, wound type and study design. Throughout this review, ESWT was administered in addition to SWC.

#### 3.2.6. Types of Intervention (ESWT Group) (V6)

In the ESWT groups, patients received fESWT, uESWT or rESWT in addition to SWC, with protocols varying in energy level, impulse count and treatment frequency. The number of treatment sessions ranged from a single application, used for acute surgical and burn wounds [78,82,85] and in single-session mechanistic studies [115,116], to 30 weekly sessions in a case of refractory chronic ulceration [97]. Treatment parameters were adjusted according to wound type, chronicity and individual patient response (e.g., [28,104]).

#### 3.2.7. Types of Intervention (Control Group, if Applicable) (V7)

In studies with control groups, patients typically received SWC, which included interventions such as regular debridement, moist wound dressings, infection control and offloading where appropriate. Some studies incorporated additional therapies into the control protocols, such as HBOT (e.g., [23]), intermittent pneumatic compression (IPC) (e.g., [24]) or sham ESWT to maintain blinding in RCTs (e.g., [28,105]). All control regimens included debridement and moisture-retentive dressings; offloading and compression were reported inconsistently.

#### 3.2.8. Number of Patients in the ESWT Group (V8)

The number of patients in ESWT groups varied substantially across studies (mean ± SD: 36.7 ± 56.2; median: 20; range: 1–282). Case reports and small case series included as few as 1 to 5 patients (e.g., [84,98]), whereas larger RCTs and cohort studies enrolled 10 to more than 280 patients [83]. The largest ESWT group reported in an RCT consisted of 172 patients, treated with ESWT across two multicenter trials [28].

#### 3.2.9. Number of Patients in the Control Group (V9)

In studies including control groups, the number of control patients generally mirrored that of the ESWT group, although slight imbalances were common (mean ± SD: 35.0 ± 39.9; median: 20; range: 1–164). Smaller trials and cohort studies included 10 to 50 control patients (e.g., 10 controls in [79] and 41 controls in [25]), while larger RCTs enrolled substantially more participants. The largest control group consisted of 164 patients in a sham control arm closely matched to the 172 ESWT patients in the same study [28].

#### 3.2.10. ESWT Device Used (V10)

Frequently used devices included Duolith SD1 (electromagnetic; 10 studies), dermaPACE (electrohydraulic; 7 studies), DermaGold (electrohydraulic; 6 studies), DolorClast (radial; 4 studies), Evotron (electrohydraulic; 4 studies), Orthowave (electrohydraulic; 3 studies), ShockMaster (radial; 3 studies), Cellactor SC1 (radial; 3 studies), PiezoWave and PiezoWave2 (piezoelectric; 3 studies each) and OrthoGold 100 (electrohydraulic; 1 study). Manufacturers of these and of the remaining devices are listed in Table 10.

#### 3.2.11. Wound Type and ESWT Modality Used (V1 and V11)

Different wound types were treated with different ESWT modalities, often reflecting the depth, chronicity and complexity of the wounds. Electrohydraulic ESWT was most commonly applied to DFUs, burns and complex posttraumatic wounds (e.g., [30,77]), reflecting its early clinical adoption. Electromagnetic ESWT was frequently used for VLUs, PUs and systemic sclerosis–related ulcers, often in outpatient or rehabilitation settings (e.g., [103,104]). Piezoelectric ESWT was also used for DFUs and VLUs, with recent RCTs focusing on comparisons with other physical therapies (e.g., [24]). Radial ESWT was typically applied to pressure ulcers, superficial chronic ulcers and burns (e.g., [25,115]).

#### 3.2.12. Energy Flux Density (EFD) (V12, V13)

Reported EFD values ranged from 0.01 mJ/mm^2^ [118] to 0.25 mJ/mm^2^ [25,97,98,100,103–105,109], with most protocols between 0.1 and 0.2 mJ/mm^2^. Five studies did not report an EFD [93,102,108,113,114]; in one of these the dose was specified only as a device intensity level [108]. In several rESWT studies the quoted mJ/mm^2^ value was a nominal manufacturer figure rather than a measured energy flux density (c.f. Section 3.3.5).

None of the studies specified whether the reported EFD referred to positive energy flux density (EFD_+_) or total energy flux density (EFD_total_). This introduces considerable ambiguity because these two measures differ substantially in magnitude and clinical interpretation [9].

#### 3.2.13. Number of ESWs per Treatment Session and/or per cm^2^ Wound Area (V14, V15)

Where an absolute number of ESWs per treatment session was reported, it ranged from 300 to 7000 [86,100]. Because many studies did not specify the number of ESWs per treatment session, corresponding mean and median values cannot be provided.

Many studies scaled the number of ESWs to wound area, with 100 ESWs/cm^2^ wound area, representing a commonly applied dose (e.g., [81,106]). In some studies, doses as high as 1000 ESWs/cm^2^ wound area were applied [111]. One RCT compared two impulse densities of the same device directly, randomizing patients with DFUs to 500 or 100 ESWs/cm^2^ at an identical EFD alongside a sham arm [27].

#### 3.2.14. Frequency (in Hertz) at Which the ESWs Were Applied (V16)

The frequency of ESW application varied across studies but generally ranged from 3 to 5 Hz, except in studies using rESWT (8 Hz in [118], 10 Hz in [114] and 9–18 Hz in [25]). Some studies did not report the exact frequency of ESW application (e.g., [76,94,107]).

#### 3.2.15. Number of Treatment Sessions (V17)

The number of treatment sessions per patient also varied widely, from 1 to 30 [82,97]. Treatments were administered over periods ranging from a few days to several weeks. When the number of ESWs per session and the number of sessions per treatment course are considered together, total treatment exposure varied by more than two orders of magnitude, both in cumulative ESWs and in total energy delivered. Only a minority of studies combined ESWs per unit wound area and number of sessions into an explicit dosing rationale (e.g., [101]).

#### 3.2.16. Interval Between Treatment Sessions (V18)

The interval between ESWT treatment sessions varied considerably across studies, reflecting differences in wound type, chronicity and treatment protocol. Most studies administered 3 to 12 sessions, usually once or twice weekly (e.g., 4-8 sessions in [28] and 6 sessions in [112]). The shortest interval was used in an RCT that delivered three sessions within 7 ± 2 days [27], and the longest in a case report using fortnightly sessions [108]. The total number of sessions was frequently adapted to wound response, wound size or patient tolerance.

#### 3.2.17. Primary Endpoint (V19)

Across studies, primary endpoints most commonly focused on wound healing outcomes, particularly reduction in wound size, rate of complete wound closure or time to healing. Many RCTs defined percentage reduction in wound area over a defined time period (e.g., 4 or 12 weeks) as the primary efficacy measure (e.g., [98,105]). Some studies used complete epithelialization or number of healed wounds as binary endpoints (e.g., [95]), while others incorporated composite outcome measures including granulation tissue formation, pain reduction and infection control (e.g., [79]).

#### 3.2.18. Time Post-Baseline When the Primary Endpoint Was Assessed (V20)

The timing of primary endpoint assessment varied with study design, typically between 4 and 12 weeks post-baseline. Many studies assessed the primary endpoint at 4 weeks (e.g., [102,106,112]). A smaller number extended follow-up to 24 weeks [28,90] or, in one case series, to 1 and 5 years [87].

#### 3.2.19. Secondary Endpoints (V21)

Secondary endpoints varied across studies, but frequently included pain reduction, quality of life (QoL), granulation tissue formation, wound perfusion and adverse events. Several studies assessed pain using visual analog scales (VAS) or similar tools (e.g., [99,103]), while QoL was often evaluated using instruments such as EQ-5D or disease-specific questionnaires (e.g., [100,107]). Other commonly reported secondary outcomes included infection rates, time to first signs of granulation tissue formation and rates of wound recurrence. Some studies also assessed biological indicators of healing, such as thermography or perfusion imaging (e.g., [30,87]).

#### 3.2.20. Time Post-Baseline When the Secondary Endpoints Were Assessed (V22)

The timing of secondary endpoint assessments generally mirrored that of the primary endpoints, with most evaluations conducted 4 to 12 weeks post-baseline. Pain levels, granulation tissue formation and early tissue responses were often measured weekly or biweekly during treatment, whereas QoL measures and recurrence outcomes were typically assessed at final follow-up visits, such as 8, 12 or 24 weeks (e.g., [100,107]). In some studies, secondary endpoints such as infection status or perfusion changes were monitored at intermediate time points (e.g., 2 or 6 weeks) to capture early physiological responses to treatment (e.g., [100,105]).

#### 3.2.21. Prospective Sample Size Calculation and/or Prospective Power Analysis (V23, V24)

Most studies did not report a prospective sample size calculation or power analysis, particularly among case reports, case series and non-randomized cohort studies. A prospective sample-size calculation was reported by a minority of studies, almost all of them controlled trials, for example [28,30,104,119]. In contrast, many smaller RCTs and exploratory trials did not report such planning, reflecting either pilot-study status or limitations in recruitment capacity.

#### 3.2.22. Results Obtained for the Primary Endpoint (V25)

Results for the primary endpoint (most commonly wound size reduction or complete healing) were generally favorable for ESWT compared with control treatments. Many studies reported statistically significant improvements in wound area reduction or healing rates in the ESWT group (e.g., [98,106]), with several trials documenting greater than 50% reduction in wound size over 4–12 weeks. In larger RCTs, ESWT significantly outperformed sham treatment with respect to wound closure rates [28]. However, a few studies did not demonstrate statistically significant differences, often a consequence of small sample sizes or insufficient statistical power (e.g., [99]). Complete healing was reported as a proportion in 22 of the 51 records, and these proportions differed markedly with the study design: the 14 uncontrolled records reached a median of 89%, with complete healing in each of the six case reports, whereas the eight controlled trials reached a median of 52% in their ESWT groups, against a median of 26% in their control groups (Figure 3).

**Figure 3.**
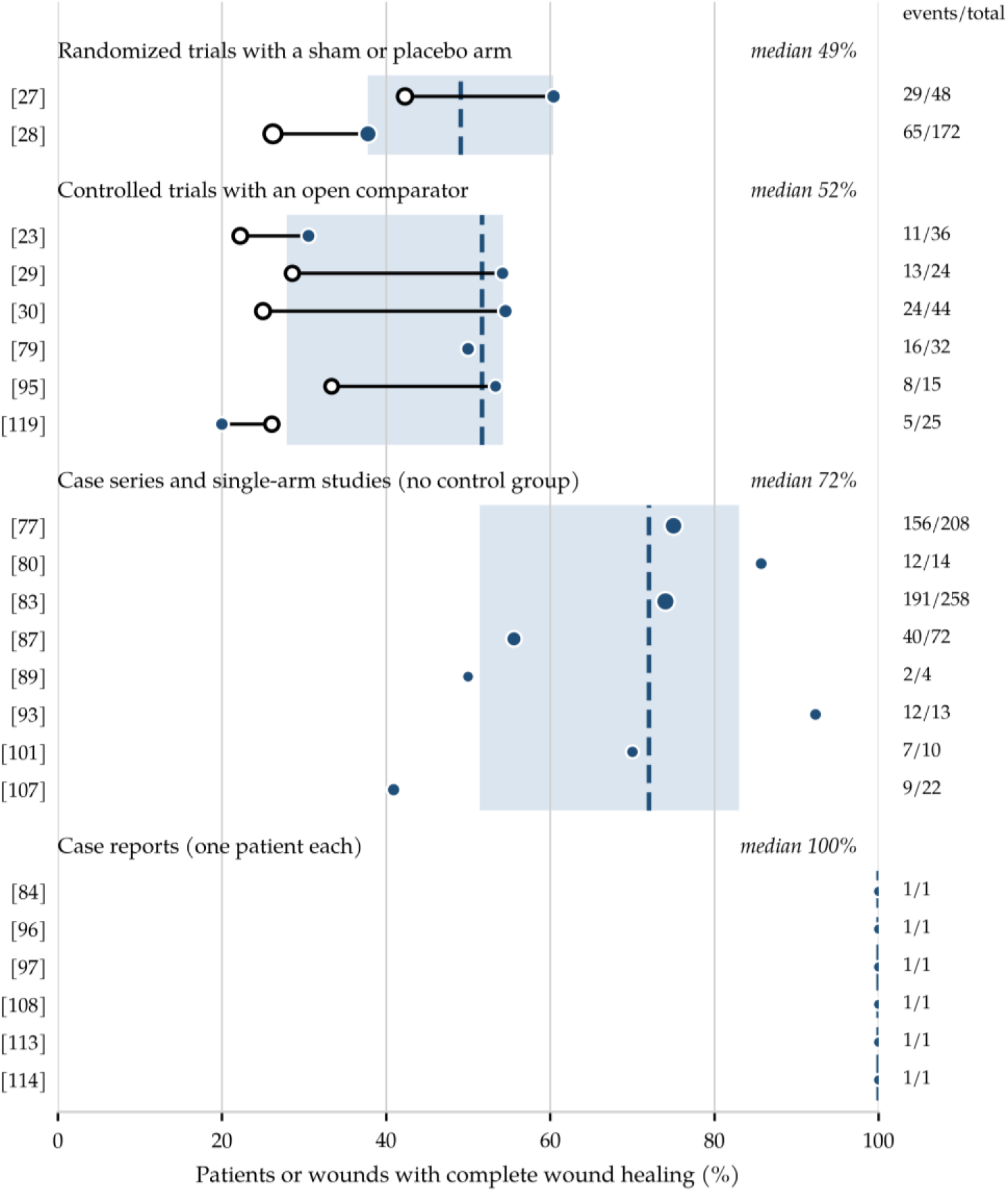
Proportion of patients or wounds with complete wound healing in the ESWT group, by study design. Each row is one record, identified by its reference number; filled circles show the ESWT group and open circles the control group of the same record, joined by a line. Marker size increases with the number of patients or wounds on which the proportion is based (events/total, given at the right). The dashed line and the shaded band give the median and the interquartile range of the ESWT groups within each stratum. Only the 22 of the 51 records that reported complete healing as a proportion are shown; the remaining records reported wound area, perfusion, pain, molecular or histological endpoints instead.

#### 3.2.23. Results Obtained for the Secondary Endpoint (V26)

Results for secondary endpoints generally supported the potential benefits of ESWT, with many studies reporting reduced pain levels, improved quality of life and enhanced granulation tissue formation. Pain scores decreased significantly in several trials using VAS or comparable instruments (e.g., [100,102]), and improvements in quality-of-life measures were observed in both generic and disease-specific scales (e.g., [100,107]). Other secondary findings included increased tissue perfusion (e.g., [30]) and faster onset of granulation tissue formation (e.g., [79]); adverse events are reported in Section 3.2.25.

#### 3.2.24. Retrospective Power Analysis (V27, V28)

Retrospective (post hoc) power analyses were rarely reported across the studies. Most trials that did not achieve statistical significance did not include a formal post hoc power calculation to evaluate the possibility of Type II error. One exception was a study that explicitly acknowledged that the trial may have been underpowered to detect a statistically significant difference, implying retrospective consideration of statistical power [99].

#### 3.2.25. Adverse Events (V29)

Adverse events associated with ESWT were rare and generally mild and transient across nearly all studies. The most frequently reported events included local discomfort, erythema, bruising or mild pain at the treatment site, which typically resolved without medical intervention (e.g [28,30]). Serious adverse events occurred in these populations, as is expected in patients with chronic wounds and multiple comorbidities, but none was attributed to ESWT. In the two largest sham-controlled trials, such events were less frequent in the ESWT group than in the sham group (32.0% versus 43.3%; p = 0.042) [28], and in an RCT comparing two ESWT doses with sham treatment, adverse events were likewise more frequent in the sham arm [27]. Several studies explicitly noted the favorable safety profile of ESWT, even in patient populations with impaired wound healing capacity, such as individuals with diabetes or systemic sclerosis (e.g., [77,104]).

#### 3.2.26. PEDro Assessment (V30–V40)

PEDro scores (maximum score 10) across the 26 records with a control group ranged from 4 to 9 (mean ± SD: 6.6 ± 1.5; median: 6.5), indicating moderate methodological quality. The dataset included several well-conducted RCTs alongside studies with limitations related to sample size or blinding. The highest score, 9, was reached by three studies, each of which met every criterion except blinding of the treating therapist, including randomization with concealed allocation, blinding of participants and outcome assessors, and high retention rates [28,82,85]. The companion report of the secondary endpoints of the same two trials scored 8 [90]. Overall, the most frequently unmet criteria were blinding of the treating therapist (unmet in all 26 records), concealed allocation (18 of 26) and blinding of the participants (16 of 26), whereas outcome measures and statistical analyses were generally reported consistently.

#### 3.2.27. Distribution of Overall Outcomes across ESWT Modalities

The distribution of overall outcomes according to the classification scheme outlined in Table 3 showed the same range of outcome categories across ESWT modalities, although their relative frequencies differed. Among the 22 studies using electrohydraulic devices (Table 5), 10 (45.5%) were controlled studies reporting significantly better outcomes with ESWT than with the comparator (category 1), one (4.5%) compared two ESWT regimens that both improved from baseline without a demonstrated difference between them (category 2a) and 11 (50.0%) were uncontrolled (nine case series and two case reports; category 5). Within the largest of these studies, the pooled analysis and the first of the two constituent trials met category 1, whereas the second trial met category 2b. Of the 11 studies using electromagnetic devices (Table 6), three (27.3%) reported category-1 outcomes and two (18.2%) reported category-2b outcomes, while six studies (54.5%) were uncontrolled (four case series and two case reports; category 5). The six studies using piezoelectric devices (Table 7) comprised three category-2b studies (50.0%) and three uncontrolled studies (50.0%; one case series and two case reports, category 5); none of them showed a statistically significant advantage over its comparator. One of the category-2b studies was an RCT that compared two ESWT doses with sham treatment but performed no statistical testing. Therefore, its classification rested on the pattern of the reported results rather than on a formal comparison [27]. Among the records obtained with radial devices (Table 8), which comprised 10 studies using a radial device only and the two trials that also used a piezoelectric device, five (41.7%) reported category-1 outcomes, one (8.3%) a category-2b outcome, one (8.3%) a category-4 outcome and five (41.7%) were uncontrolled (three case reports and two case series; category 5). Of the two studies in which the modality was not disclosed (Table 9), one reported a category-1 outcome [29] and the other a category-2b outcome [119].

No study was classified as category 3. One study met the definition of category 4: in a three-arm trial in patients with VLUs, ultrasound therapy produced a significantly greater reduction in ulcer area than rESWT (67.6% versus 38.2%; p < 0.05), although both were significantly superior to SWC alone [112]. In a second trial from the same group, IPC produced the largest median area reduction, ahead of both ESWT modalities [24]. In one further trial the proportion of ulcers healed at 6 weeks was numerically lower with ESWT than with SWC alone (5/25 versus 6/23; p = 0.73), while the mean reduction in wound surface area favored ESWT (36.1 versus 5.1 mm^2^; p = 0.33) [119]. Most controlled studies therefore reported outcomes favoring ESWT over SWC or sham treatment, but the categorization should not be read as evidence that no study produced a result favoring a comparator. The near-absence of negative categories across a literature in which 25 of 51 records are uncontrolled should also be interpreted with the possibility of publication bias in mind.

### 3.3. Feasibility of Quantitative Synthesis

The heterogeneity described above precludes a conventional meta-analysis of the evidence base as a whole. The three questions defined in Section 2 are addressed in turn, and identify more precisely where the evidence is thin.

#### 3.3.1. Endpoints Available for Pooling

Of the 51 records, seven reported complete wound healing as events and denominators in both an ESWT group and a control group [23,27–30,95,119]. For continuous outcomes, the position was less favorable. Means with standard deviations for the percentage reduction in wound area were available in nine records, whereas four reported standard errors and five reported medians with quartile ranges only. Several records reported no wound-closure or wound-area outcome at all, most often because the study was designed around perfusion, molecular or histological endpoints.

#### 3.3.2. Exploratory Pooled Analysis of Complete Wound Healing

In these seven studies [23,27–30,95,119], complete healing occurred in 155 of 364 patients or wounds treated with ESWT and in 89 of 325 controls (Table 12).

The pooled risk ratio was 1.51 (95% CI 1.22-1.87). Cochran Q was 3.60 on six degrees of freedom, so I^2^ was 0% and tau-squared was zero, and the random-effects estimate was identical. This value is close to that reported by the most recent meta-analysis restricted to DFUs (risk ratio 1.57, 95% CI 1.26-1.95) [64], with which it shares four of seven trials.

The estimate was stable across the restrictions that the data permit. Excluding the trial in which allocation followed the calendar date of referral [23] gave 1.52 (1.22-1.90); restricting the analysis to the two sham-controlled trials [27,28] gave 1.44 (1.10-1.88); and excluding the two trials in which the comparator was HBOT [23,30] gave 1.44 (1.14-1.83). The absence of statistical heterogeneity should not be read as agreement between the trials. They differed in comparator, wound type, assessment timepoint, unit of analysis and device, and the confidence intervals of the individual trials are wide enough to overlap in spite of those differences. The analysis is therefore best understood as a check that the qualitative synthesis is consistent with the pooled literature, and not as a substitute for it.

#### 3.3.3. Availability of Controlled Data by Modality

The seven trials in Table 12 were not distributed evenly across the four modalities (Table 13).

**Table 11.**
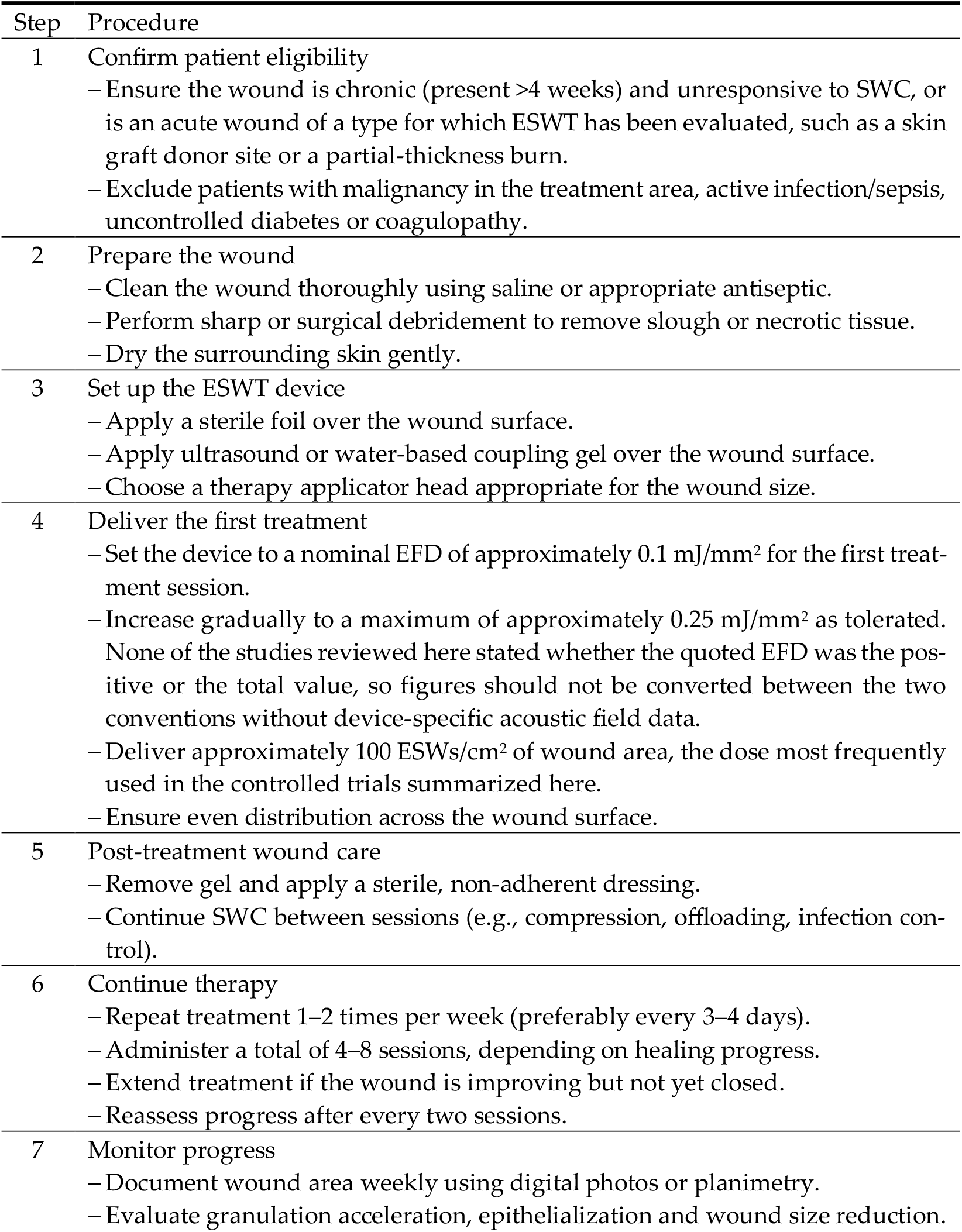
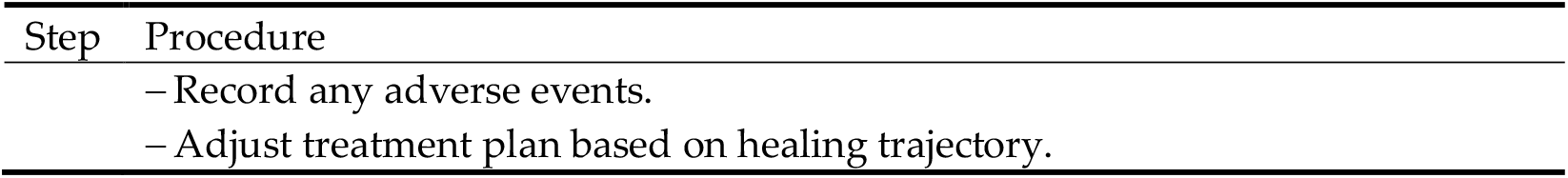
Outline of a generalized, evidence-informed ESWT treatment protocol for wound management.

**Table 12.** Controlled studies reporting complete wound healing as events and denominators in both study groups, with an exploratory pooled estimate. In the three-arm trial [27] the two active arms were combined (Section 2). Risk ratios above 1 favor ESWT. Abbreviations: EH, electrohydraulic; EM, electromagnetic; HBOT, hyperbaric oxygen therapy; RR, risk ratio; SWC, standard wound care.

| Study | Modality | Comparator | ESWT | Control | Assessment | RR (95% CI) |
| --- | --- | --- | --- | --- | --- | --- |
| Wang et al. 2009 [23] | EH | HBOT | 11/36 | 8/36 | End of follow-up | 1.38 (0.63-3.01) |
| Moretti et al. 2009 [95] | EM | SWC | 8/15 | 5/15 | 20 weeks | 1.60 (0.68-3.77) |
| Wang et al. 2011 [30] | EH | HBOT | 24/44 | 10/40 | After one course | 2.18 (1.20-3.98) |
| Omar et al. 2014 [29] | Not stated | SWC | 13/24 | 6/21 | 20 weeks | 1.90 (0.88-4.09) |
| Snyder et al. 2018 [28] | EH | Sham + SWC | 65/172 | 43/164 | 24 weeks | 1.44 (1.05-1.99) |
| Vangaveti et al. 2023 [119] | Not stated | SWC | 5/25 | 6/23 | 6 weeks | 0.77 (0.27-2.18) |
| Hitchman et al. 2025 [27] | PE | Sham + SWC | 29/48 | 11/26 | 24 weeks | 1.43 (0.86-2.36) |
| Pooled (fixed effect) |  |  | 155/364 | 89/325 |  | 1.51 (1.22-1.87) |

**Table 13.** Distribution of the evidence across ESWT modalities. The pooled estimates are shown for completeness only: the electrohydraulic estimate rests on three trials, two of which used HBOT as comparator, and the piezoelectric estimate on a single trial that was not powered for efficacy. Abbreviations: RR, risk ratio.

| Modality | Controlled trials with healing counts | Patients | Pooled RR (95% CI) | Uncontrolled records contributing a healing proportion |
| --- | --- | --- | --- | --- |
| Electrohydraulic | 3 | 492 | 1.55 (1.19-2.03) | Seven series, approximately 645 wounds, plus one series of 64 patients without an event count |
| Electromagnetic | 1 | 30 | Not estimable ($k = 1$) | Four series, 28 patients |
| Piezoelectric | 1 | 74 | 1.43 (0.86-2.36) | Two records, 23 ulcers |
| Radial | 0 | - | Not estimable | Two series, neither reporting a closure outcome |
| Not stated | 2 | 93 | Not applicable | None |

Controlled data on complete healing were available for electrohydraulic devices in three trials, for piezoelectric devices in one and for electromagnetic devices in one, and were not available for radial devices in chronic wounds; the only controlled rESWT trial in acute wounds, in deep dermal burns of the hand, reported the median percentage reduction in wound area after 14 days rather than complete healing [118]. Two of the seven trials [29,119] did not name the device used and could not be assigned to any modality.

Neither of the requirements for an indirect comparison between modalities defined in Section 2 was met: the five trials that could be assigned to a modality fell short of the ten studies recommended for a meta-regression [26], and only one modality was independently estimable. The uncontrolled literature was distributed even more unevenly: approximately 645 wounds in seven electrohydraulic case series, together with a further series of 64 patients reporting complete closure for the whole cohort without giving an event count [94]; 28 patients in four electromagnetic series; 23 ulcers in two piezoelectric records; and no radial series reporting a closure outcome at all. A proportion pooled across the modalities would consequently be an electrohydraulic estimate. This distribution, rather than any observed difference in effect, is what currently prevents a comparison between technologies.

#### 3.3.4. Records Reporting Overlapping Participants

Six groups of publications among the 51 records reported overlapping or identical participants. Snyder et al. [28] and Galiano et al. [90] reported the primary and the secondary endpoints of the same two phase III trials and thus the same 172 patients treated with ESWT and 164 sham-treated controls; only [28] entered Table 12. The two reports of Ottomann et al. [82,85] were two wound strata of one clinical program. Schaden et al. [77] and Wolff et al. [83] were uncontrolled series from the same center with overlapping recruitment windows. The three trials of Dolibog et al. [24,106,112] shared a single ethics approval, the same two devices and the same treatment parameters, with overlapping recruitment, so participant overlap between them cannot be excluded. Dymarek et al. [115,116] and Sopel et al. [117] belonged to one research program and shared device and dose. Wang et al. [87] and Chou et al. [89] were series from the same center. These relationships do not diminish the value of the individual reports, but they should be taken into account whenever the number of studies or the number of patients is used as a measure of the strength of the evidence.

#### 3.3.5. Comparability of Treatment Dose across Modalities

A quantitative comparison of dose across modalities is not possible with the data as reported. Radial devices are ballistic sources that generate a pressure wave rather than a true shock wave, and EFD is not the quantity that governs their output; the parameter reported by their manufacturers is the air pressure driving the projectile, in bar. Several radial studies nevertheless quoted an EFD alongside a pressure, and the implied conversions are not mutually consistent: 2.5 bar was equated with 0.15 mJ/mm^2^ in one trial [115] and 1.0 bar with 0.01 mJ/mm^2^ in another [118], a sixfold difference in the ratio between the two quantities, in studies that used the same protocol of 300 followed by 100 ESWs/cm^2^. A third study quoted a single EFD against a pressure range of 2-4 bar [25]. These figures are therefore best understood as device-specific nominal values rather than as measured energy flux densities, and any analysis that would regress clinical outcome on EFD across modalities would in part be regressing on these nominal values.

A second obstacle is that three incompatible dosing conventions coexist in the literature. Most studies scaled the number of ESWs to the wound area, with 100 ESWs/cm^2^ as the modal protocol; others delivered a fixed number of impulses irrespective of wound size, such as the 500 impulses used with one electrohydraulic device or the 2000 impulses used in several electromagnetic protocols; and a third group dosed an anatomical region rather than a wound, as in the 7000 impulses applied across both hands and forearms in patients with systemic sclerosis [100]. Cumulative exposure across a treatment course was consequently not comparable between studies even within a single modality, and the observation that the four modalities occupy different regions of this parameter space confounds any indirect comparison between them.

## 4. Discussion

The present systematic review synthesizes the currently available clinical evidence on ESWT in wound management by evaluating 51 clinical studies [23–25,27–30,76–119] and placing these findings in the context of 46 previously published reviews [22,31–75]. Most of the included studies reported the improvements in wound healing parameters summarized in Section 3.2.5, and reported them across DFUs, VLUs, PUs, burns and postsurgical wounds alike. The consistency in the direction of effect across wound types, rather than its magnitude, is the feature of this evidence base that most requires explanation. An exploratory pooled analysis of the seven controlled trials that reported complete healing as events and denominators gave a risk ratio of 1.51 (95% CI 1.22-1.87) in favor of ESWT (Section 3.3.2), a value consistent with published meta-analyses restricted to DFUs [64,70] as well as with published network analyses in which ESWT appeared as one node among several interventions [67,69,72]. Those network analyses did not agree on where ESWT ranks: it was placed first for healing time in one [67] and second for area reduction in another [69], but 12 of 15 for the 12-week healing rate in a third [75]. The three networks were drawn on different trial sets and defined the ESWT node differently, with [75] restricting it to focused devices, which is itself an illustration of the difficulty this review describes.

A finding that applies to the review literature as a whole is that none of the 46 previously published reviews reflected the available original evidence in its full breadth. Coverage of the clinical studies that had been published by a review’s own reference date ranged from 0% to 90.9%, with a median of 19.4%, and was no higher in systematic reviews (median 20.6%) than in narrative ones (median 15.8%). Table A2 makes the pattern concrete: a trial by Wang et al. (2011; [30]), one of the most frequently cited in this field, appeared in 18 of the 42 reviews (42.9%) published after it became available, and three reviews cited no clinical study of ESWT for wound management [37,41,65]. Restrictive inclusion criteria account for part of this in the systematic reviews, but not for the pattern in the narrative literature, where an explicit justification for the selection was usually not reported. The consequence for a reader is that no single review, meta-analyses included, reflected the evidence base as a whole, and that the conclusions of these reviews, favorable and inconclusive alike, rested on subsets of the evidence that differed from one another. Assembling the full set of primary studies is therefore a first requirement for further work on this topic.

A second and independent limitation of the review literature is its currency. Just over half of the reviews (26 of 46) stated a date to which their literature search was current, and among those the median interval between that date and publication was 10.5 months, with seven reviews at 15 months or more. Because the primary literature in this field has continued to grow, this delay has consequences beyond the publication process: the median review was already missing 8% of the then-available evidence on the day it became citable. In a review by Howard et al. (2025; [68]) the interval was 54 months and 18 further clinical studies had appeared in the meantime; the synthesis covered 32 studies at a time when 50 were available. The 20 reviews that reported no search date are harder to judge, because a reader cannot tell how current such a review is, and the year of publication is not a reliable guide to it. Two conclusions follow for the use and the reporting of reviews in this field. First, the date to which the search was current, rather than the year of publication, is the date that should be quoted whenever a review is cited as evidence, and it should be stated in the abstract. Second, the interval between search and publication belongs in the appraisal of a review alongside its inclusion criteria and its risk-of-bias assessment, because the currency of the search limits how much a review adds to the primary literature, independently of how carefully it was conducted.

The present analysis also highlights an important feature of the existing literature: despite clear technological differences between ESWT systems, clinical studies have generally treated ESWT as a unified therapeutic concept. Across narrative reviews, systematic reviews without meta-analysis and those including meta-analysis, different ESWT modalities (including electrohydraulic, electromagnetic, piezoelectric and radial devices) have typically been grouped together when evaluating clinical outcomes [31,33,42,44,49,50,52,64]. Although these technologies differ substantially in their methods of generating acoustic energy (cf. Figure 1), the available clinical evidence does not demonstrate a therapeutic advantage of one modality over another. This is an absence of evidence rather than evidence of equivalence: as shown in Section 3.3.3, controlled data on complete healing exist for only three of the four modalities, in a ratio of three electrohydraulic trials to one electromagnetic and one piezoelectric trial, and two of the seven trials did not report which device was used. The assumption of functional equivalence has nevertheless become widespread, although it has been tested directly in only two trials [24,106]. This observation reflects both the pragmatic way in which ESWT has been incorporated into clinical practice and the current lack of comparative studies specifically designed to evaluate modality-dependent treatment effects. Only two reviews departed from this assumption: a national expert consensus recommended radial ESWT for superficial defects and focused ESWT for deep tissue involvement [74], and a further review classified the published protocols by dose density [73]. Neither recommendation rests on a direct comparison at matched physical dose, so both remain inferences from device physics rather than conclusions from trial evidence.

The analysis of the review literature also reveals a recurrent pattern of overlapping clinical indications and patient populations in ESWT studies. Many narrative reviews described the application of ESWT across a wide spectrum of acute and chronic wounds, including DFUs, VLUs, burns and postsurgical wounds, without systematically stratifying outcomes according to wound etiology or patient characteristics [31,36,38,44]. Similarly, several systematic reviews combined results from heterogeneous patient populations and wound types to reach overall conclusions regarding ESWT efficacy [35,48,50]. Even in meta-analyses that focused primarily on DFUs, substantial variability in ulcer characteristics, comorbidities and treatment protocols was often present among the included trials [22,60,64]. While this broad applicability reflects the versatility of ESWT as a therapeutic modality, it also introduces heterogeneity that complicates the interpretation of clinical outcomes and limits the development of indication-specific treatment recommendations.

A consistent theme across both the review literature and the primary clinical studies is the proposed biological basis of ESWT-induced tissue repair. Narrative reviews frequently described common biological pathways through which shock waves may influence wound healing, including stimulation of angiogenesis, increased expression of vascular endothelial growth factor (VEGF), activation of fibroblasts and enhancement of local microcirculation [31,33,38,44,56]. Angiogenesis occupies a central position in this account, as it does in cutaneous wound repair generally: restoration of the vascular supply to the regenerating tissue is a prerequisite for closure and is the target of several other regenerative strategies as well [121]. These mechanisms are supported by both experimental and clinical observations. For example, increased expression of angiogenesis-related markers such as endothelial nitric oxide synthase (eNOS), VEGF and proliferating cell nuclear antigen (PCNA) has been reported following ESWT treatment [23]. Improvements in tissue oxygenation and perfusion have also been documented, including higher TcPO_2_ levels after treatment [99]. Additional studies have demonstrated increased numbers of CD31positive vessels and improved vascularization in treated tissues [91,117].

Mechanotransduction appears to represent a central mechanism underlying these biological effects. Shock waves generate mechanical stresses within tissue that are converted into biochemical signals capable of activating cellular repair pathways. For example, increased extracellular signal-regulated kinase activity in macrophages has been observed following ESWT exposure, suggesting activation of intracellular signaling cascades involved in tissue regeneration [91]. In addition, activation of yes-associated protein 1 (YAP1) signaling has been associated with enhanced proliferation of keratinocytes and fibroblasts and increased extracellular matrix remodeling in response to rESWT [117]. These mechanotransductive responses are accompanied by modulation of inflammatory processes. Chronic wounds are often characterized by persistent inflammatory activation, and several studies have reported reductions in inflammatory markers such as C-reactive protein following ESWT treatment [25,119]. In addition, decreases in macrophage density within wound biopsies have been described, suggesting resolution of chronic inflammatory activity [91].

ESWT has also been associated with stimulation of fibroblast activity and differentiation of myofibroblasts, processes that are essential for wound contraction and collagen deposition. Increased expression of α-smooth muscle actin-positive myofibroblasts has been observed after treatment with rESWT, indicating enhanced remodeling capacity within the healing tissue [117]. Increased granulation tissue formation, collagen synthesis and epithelialization following treatment have been described both histologically and clinically. Re-epithelialization has been reported as an outcome in clinical studies using electrohydraulic [77], electromagnetic [105], piezoelectric [107] and radial [110] ESWT devices, suggesting that the biological effects of acoustic stimulation are broadly shared across ESWT technologies and likely arise from common mechanotransductive processes within the treated tissue.

One physical mechanism proposed to initiate this signaling is acoustic cavitation. This consists of the formation, oscillation and collapse of microbubbles within tissue fluids exposed to high-pressure acoustic waves, and the rapid pressure fluctuations this produces in the surrounding microenvironment [11,122]. These dynamic physical forces generate localized shear stress and micro-mechanical strain that can stimulate cellular signaling pathways and promote the release of pro-regenerative growth factors. Although cavitation has been proposed as a mechanistic explanation in several clinical studies of ESWT for wound management [79,86,95,98], its occurrence within human wound tissue during treatment has not yet been demonstrated experimentally.

The clinical effects reported in the included studies are compatible with this mechanism, although no study measured mechanotransductive signaling and clinical outcome in the same patients. Additional biological responses reported in clinical and experimental studies, such as upregulation of angiogenic mediators (e.g., VEGF and eNOS), modulation of inflammatory pathways [123] and increased fibroblast activity, are consistent with the concept that mechanically induced signaling cascades contribute to the improvements in wound healing that have been observed. How energy delivery parameters, cavitation and wound-specific outcomes relate to one another is not yet known, and comparative and biomarker-driven studies are needed to establish it.

The clinical literature also illustrates considerable variability in ESWT treatment protocols, reflecting both the evolving nature of the field and the incomplete understanding of how specific energy delivery parameters influence the biological mechanisms described above. Across the analyzed studies, parameters such as EFD, number of impulses per session, treatment frequency and interval between treatment sessions varied substantially. Some studies applied relatively low energy levels and repeated treatments over several weeks, whereas others used higher energies with fewer sessions. Several reviews identified this variability in treatment parameters as a major source of heterogeneity in the evidence base [42,50]. Although energy delivery is a potentially important determinant of treatment response, the existing literature provides no clear evidence that higher EFD consistently produces superior clinical outcomes [22,52,64]. Instead, the available studies suggest that therapeutic effectiveness depends on the interaction between energy delivery, tissue characteristics and wound chronicity rather than on the absolute energy level alone. A further difficulty is that dose has not been reported on a common scale (c.f. Section 3.3.5). Focused devices are specified by EFD, ballistic devices by the air pressure driving the projectile. For radial devices, the difficulty is compounded by the fact that the acoustic output is not determined by the pressure setting alone. In a standardized in vitro comparison of two such devices, raising the pulse repetition rate from 1 Hz to about 20 Hz left the output of the DolorClast (Electro Medical Systems) almost unchanged, with losses of up to 12% in peak positive pressure and up to 18.8% in the positive pulse intensity integral, whereas the same increase reduced these quantities by up to 68.4% and up to 90.2% in the MasterPuls 200 Ultra (Storz Medical); operating the latter at 5 bar and 21 Hz yielded a positive pulse intensity integral comparable to operating it at 1 bar and 1 Hz [124]. The difference was attributed to the air-flow management of the two devices, that is, to a property of the device rather than of the handpiece or applicator fitted to it [124].

The consequence is stronger than a reporting deficiency. The air pressure driving the projectile is an input setting and carries no information about the acoustic energy that leaves the applicator, and for an uncharacterized device that energy may fall by an order of magnitude across the repetition rates used in practice. The energy delivered by a radial device therefore has to be treated as unknown unless the acoustic output of that device has been measured, independently of the manufacturer’s specifications, across the repetition rates at which it was operated. Four of the 12 radial records analyzed here used a DolorClast device, at 1 to 4 bar and 4 to 18 Hz where these were reported [25,113,118] and without a stated repetition rate in one case [111]; for this device, the published measurements cover the settings used, and, while the absolute output still depends on the handpiece and applicator fitted, it does not collapse with the repetition rate. For the remaining eight records, the delivered energy is unknown: three used a ShockMaster (Gymna Uniphy) [24,106,112], three a Cellactor (Storz Medical) [115–117], one a BTL-5000 (BTL) [110] and one a radial device identified only by its manufacturer [114], and for none of these has the dependence of the acoustic output on the pulse repetition rate been reported. The mJ/mm^2^ values quoted in these eight records are therefore manufacturer labels rather than measured energies, and they cannot be placed alongside the energy flux densities of focused devices. Comparisons of dose across modalities, and any attempt at meta-regression on EFD that pools focused and radial devices, are therefore not currently interpretable.

A first step towards resolving this has now been taken: an RCT randomized patients with DFUs to two different impulse densities of the same piezoelectric device (500 versus 100 ESWs/cm^2^ at an identical EFD of 0.1 mJ/mm^2^) and to sham treatment [27]. At 6 weeks, complete healing occurred in 9/25 (36.0%) of patients in the high-dose arm, 4/23 (17.4%) in the low-dose arm and 3/26 (11.5%) in the sham arm, corresponding to risk ratios against sham of 3.12 (95% CI 0.95-10.21) and 1.51 (0.38-6.04), respectively; by 24 weeks the three proportions had converged to 64.0%, 56.5% and 42.3%. The trial was not powered for efficacy and performed no statistical testing, so these figures are hypothesis-generating; they are nevertheless the first direct comparison of two ESWT doses in a randomized human trial, and their ordering is consistent with a dose-dependent effect on early healing. Preclinical work points in a more complex direction: dose-gradient experiments in animals have described a non-monotonic relationship in which re-epithelialization improved at low impulse numbers and was impaired at higher ones [125], and the doses at which impairment was observed fell within the range used clinically. A dose-finding study designed for that purpose remains to be performed.

Collectively, these observations suggest that, although ESWT treatment protocols varied considerably across studies, the parameters most frequently reported clustered in a narrow band. On this basis, the protocols most frequently reported in controlled trials and well-designed cohort studies can be summarized as a generalized treatment framework (Table 11).

Study design also shapes how the reported outcomes should be read. Twenty-five of the 51 records were uncontrolled (16 case series and 9 case reports), and where complete healing was reported as a proportion these reported the highest values in the review (median 89%, and 100% in each of the case reports; Figure 3), whereas the controlled trials reported a median of 52% in their ESWT groups, against control groups that reached a median of 26%. This pattern is consistent with the known susceptibility of wound assessment to performance and detection bias [126], although the studies differed in population, device and endpoint, so the comparison is descriptive rather than causal. It nonetheless underlines the importance of sham control and blinded assessment for an endpoint with a visual component such as wound closure.

An additional limitation of the current evidence base is the near absence of direct comparative trials between ESWT modalities. Only two studies directly compared fESWT and rESWT in patients with VLUs [24,106]. In the first [106], 50 patients were allocated to rESWT (n = 17), piezoelectric fESWT (n = 15) or SWC (n = 18) at matched nominal dose (0.17 versus 0.173 mJ/mm^2^, 100 ESWs/cm^2^, 5 Hz, six sessions). The mean relative reduction in ulcer area was 46.3%, 53.7% and 31.7%, respectively, at 4 weeks, and 67.7%, 63.5% and 38.9% at 8 weeks. The two ESWT modalities did not differ from one another at either timepoint (post-hoc p = 1.000 in both instances): the authors concluded that there was no statistically significant difference between rESWT and fESWT in this indication. Relative to SWC, rESWT reached significance at 8 weeks for ulcer area (p = 0.023) whereas fESWT did not (p = 0.084); the complementary analysis of the modeled time to half closure reported in the same study showed the reverse pattern, and this internal inconsistency should be taken into account when interpreting the trial. In the second study [24], the median relative reduction in ulcer area at 4 weeks was 52.9% with IPC, 31.6% with rESWT, 18.0% with fESWT and 16.0% with SWC (omnibus p = 0.001); the only between-group difference the authors described as statistically significant was that between compression therapy and SWC, and the difference between the two ESWT modalities was not formally tested. These two studies do not establish a difference in efficacy between fESWT and rESWT, and the direction of the numerical difference between the modalities was not consistent between them. Both trials were moreover conducted by the same group at the same center under a single ethics approval, using the same two devices at identical parameters and with overlapping recruitment periods, and a third trial from the same protocol has also been published [112]; they are therefore best regarded as one program of work rather than as independent replications.

The safety profile summarized in Section 3.2.25 is consistent across wound types, modalities and patient groups, including elderly patients and patients with diabetes. Treatment-related events were mild and transient, most often local discomfort, erythema or minor bruising at the treatment site (e.g., [23,28]). Because ESWT is non-invasive and its energy level can be titrated to patient tolerance, it can be delivered in both outpatient and hospital-based wound care settings.

In the broader context of wound care, ESWT was used as an adjunct rather than as a standalone therapy in every included study. Within this multimodal framework, ESWT adds a biological stimulus to the healing cascade. However, its relative position among other physical or biophysical therapies, including ultrasound therapy, electrical stimulation or negative pressure wound therapy, remains insufficiently defined due to the lack of direct comparative trials [40].

Current clinical guidelines only partially reflect the growing body of evidence on ESWT in wound management. The clinical practice recommendations of the International Diabetes Federation [127] do not address ESWT as a therapeutic option, and the American Diabetes Association Standards of Care [128] mention it only in passing, among biophysical modalities whose supporting studies are characterized as retrospective observational studies or poor-quality randomized trials, while the International Working Group on the Diabetic Foot guidelines discuss it only briefly and recommend against the use of physical therapies in general [129]. In contrast, the Wound Healing Society guidelines endorse ESWT for diabetic foot ulcers and assign a Level I recommendation, carried forward unchanged in their most recent update, which states that ESWT increases the incidence of healing and reduces the time to heal [130]; that recommendation rests on two of the reviews and one of the clinical trials assessed here [22,28,53]. The four documents were issued between 2017 and 2026, and drew on different subsets of the trials summarized here, which accounts for part of the discrepancy. As additional high-quality trials become available, the role of ESWT in these guidelines should be reassessed.

Several limitations of the current evidence base should be acknowledged. The clinical studies analyzed here varied widely with respect to study design, patient populations, wound characteristics, treatment protocols and outcome measures. Many were small single-center trials with relatively short follow-up. Direct comparisons between ESWT modalities or dosing strategies were rare, and molecular mechanisms were more often inferred than measured in patients. Co-interventions were also reported inconsistently: offloading and compression were specified in only part of the control regimens (c.f. Section 3.2.7), and mechanical offloading is itself a determinant of healing and of ulcer remission in the diabetic foot [131], so differences in standard care between the study groups cannot be excluded as a contributor to the reported effects. Several groups of publications reported overlapping participants (c.f. Section 3.3.4), so the number of studies overstates the number of independent patient cohorts. Three studies described as randomized allocated participants by referral date, by order of presentation, or by an unstated method [23–25], and were treated here as controlled but not randomized. Taken together, these features limit the ability to define optimal treatment protocols or to identify modality-specific advantages.

The present review has limitations of its own. Methodological quality was appraised with criteria derived from the PEDro scale [9] rather than with an instrument designed for risk-of-bias assessment in systematic reviews, such as RoB 2 [132] or ROBINS-I [133], and the 25 uncontrolled records could not be appraised with any of these instruments. The search covered two databases, so studies indexed only elsewhere may have been missed, and seven of the 51 records were identified by screening the reference lists of reviews rather than by the database search itself.

## 5. Conclusions

The present systematic review provides a comprehensive synthesis of the clinical literature examining ESWT in wound management. Across a diverse set of clinical studies involving multiple wound types, the majority of studies reported improvements in wound healing parameters, including reductions in wound size, accelerated re-epithelialization, enhanced tissue perfusion and increased rates of complete wound closure. Where complete healing was reported as events and denominators in both study groups, pooling the seven eligible controlled trials gave a risk ratio of 1.51 (95% CI 1.22-1.87) in favor of ESWT. These findings were observed across studies employing different ESWT technologies, including electrohydraulic, electromagnetic, piezoelectric and radial devices.

Although the four modalities differ in how they generate the acoustic pulse, the available clinical evidence does not establish the superiority of any one technology. This should be read as an absence of comparative evidence rather than as a demonstration of equivalence: controlled data on complete healing exist for three of the four modalities but rest on a single trial for two of them, and only two trials, both from a single center, compared modalities directly. Treatment parameters such as EFD, impulse number and treatment frequency may matter more than device type, but these parameters are not currently reported on a scale that permits comparison between focused and ballistic sources.

At the same time, the evidence base has remained heterogeneous in the respects set out in Section 4. Many studies involved relatively small sample sizes, methodological quality varied, and differences in outcome measures and reporting practices complicated direct comparison. These limitations underscore the need for additional well-designed RCTs with standardized treatment protocols and clearly defined clinical endpoints.

The biological responses reported alongside clinical improvement, namely angiogenesis, fibroblast activation, granulation tissue formation and re-epithelialization, point to an effect on the wound healing process itself rather than on symptoms alone. Responses of this kind have been described with electrohydraulic, electromagnetic, piezoelectric and radial devices alike, which is compatible with mechanotransduction as a mechanism shared across the four technologies. In the studies included here, molecular or histological responses and clinical endpoints were measured in different patients, at different timepoints, or not within the same study; tissue-level measurements in patients were reported by only a small number of uncontrolled series; and no study examined whether treatment effects persist after therapy ends. Establishing whether ESWT modifies the pathophysiology of impaired healing, and not only its clinical course, will require trials that pair biomarker and histological endpoints with wound closure in the same cohort.

Four priorities follow from this synthesis. First, an adequately powered dose-finding study in humans. The eightfold range of EFD and the more than hundredfold range of impulse density currently in use have been compared systematically only once, in a trial that randomized patients to two impulse densities and to sham treatment but was not powered to detect a difference between them [27]; its results are compatible with a dosedependent effect on early healing and warrant confirmation. Second, head-to-head trials of the modalities with dose matched on a physical rather than a nominal basis. Third, agreement on a dose metric that is meaningful for ballistic devices. Fourth, consistent reporting of complete wound closure with denominators at the conventional timepoints of 12 and 24 weeks, analyzed at patient level, together with the device model and settings used.

In summary, the available clinical literature indicates that ESWT is a useful adjunctive approach for the treatment of acute and chronic wounds. While the direction of effect is consistent across studies, further high-quality research will be essential to refine treatment protocols, to establish whether the modalities differ, and to define the role of ESWT within multidisciplinary wound care.

## Supplementary Materials

The following supporting information can be downloaded at https://doi.org/10.5281/zenodo.22998149:

– standardized summaries of all 46 reviews assessed in this systematic review.
– standardized summaries and 40 key variables (cf. Table 2 in the main text) from clinical studies on electrohydraulic ESWT for wound management.
– standardized summaries and 40 key variables (cf. Table 2 in the main text) from clinical studies on electromagnetic ESWT for wound management.
– standardized summaries and 40 key variables (cf. Table 2 in the main text) from clinical studies on piezoelectric ESWT for wound management.
– standardized summaries and 40 key variables (cf. Table 2 in the main text) from clinical studies on radial ESWT for wound management.
– standardized summaries and 40 key variables (cf. Table 2 in the main text) from clinical studies on ESWT with undisclosed ESWT modality in wound management.

## Author Contributions

Conceptualization, B.M., C.N.-K., M.O., T.W., N.M. and C.S.; data curation, C.N.-K. and C.S.; formal analysis, B.M. and C.S.; methodology, B.M., C.N.-K. and C.S.; project administration, C.S.; software, C.S.; visualization, C.S.; writing—original draft preparation, C.S.; writing—review and editing, B.M., C.N.-K., M.O., T.W. and N.M. All authors have read and agreed to the published version of the manuscript.

## Institutional Review Board Statement

Not applicable.

## Informed Consent Statement

Not applicable.

## Data Availability Statement

The data underlying this article are available in the article and in its online supplementary material.

## Declaration of generative AI and AI-assisted technologies in the writing process

During the preparation of this manuscript, the authors used the large language model Claude (Opus 5; 2026; Anthropic, San Francisco, CA, USA) for the purpose of translating articles published in languages other than English, and to improve the language and readability of the text. All AI-assisted revisions were subsequently reviewed and edited by the authors, who take full responsibility for the content of the manuscript.

## Abbreviations

The following abbreviations are used in this manuscript:

DFUs: Diabetic foot ulcers
VLUs: Venous leg ulcers
Pus: Pressure ulcers
ESWT: Extracorporeal shock wave therapy
ESWs: Extracorporeal shock waves
fESWT: Focused extracorporeal shock wave therapy
rESWT: Radial extracorporeal shock wave therapy
RCTs: Randomized controlled trials
PRISMA: Preferred Reporting Items for Systematic Reviews and Meta-Analyses
uESWT: Unfocused extracorporeal shock wave therapy
PEDro: Physiotherapy Evidence Database
EFD: Energy flux density
HBOT: Hyperbaric oxygen therapy
CI: Confidence interval
I²: Measure of statistical heterogeneity
IWGDF: International Working Group on the Diabetic Foot
OR: Odds ratio
PROSPERO: International Prospective Register of Systematic Reviews
SUCRA: Surface under the cumulative ranking curve
SWC: Standard wound care
VEGF: Vascular endothelial growth factor
SD: Standard deviation
TcPO_2_: Transcutaneous oxygen pressure
ASEPSIS: Scoring system for postoperative wound infection (Additional treatment, Serous discharge, Erythema, Purulent exudate, Separation of deep tissues, Isolation of bacteria, Stay as inpatient prolonged over 14 days)
CS: Case series
IPC: Intermittent pneumatic compression
QoL: Quality of life
VAS: Visual analog scale
EQ-5D: EuroQol five-dimension questionnaire
RR: Risk ratio
eNOS: Endothelial nitric oxide synthase
PCNA: Proliferating cell nuclear antigen
CD31: Cluster of differentiation 31 (platelet endothelial cell adhesion molecule 1)
YAP1: Yes-associated protein 1
RoB 2: Cochrane risk-of-bias tool for randomized trials, version 2
ROBINS-I: Risk Of Bias In Non-randomized Studies – of Interventions
ClSt: Clinical study

## Appendix A

**Table A1.**
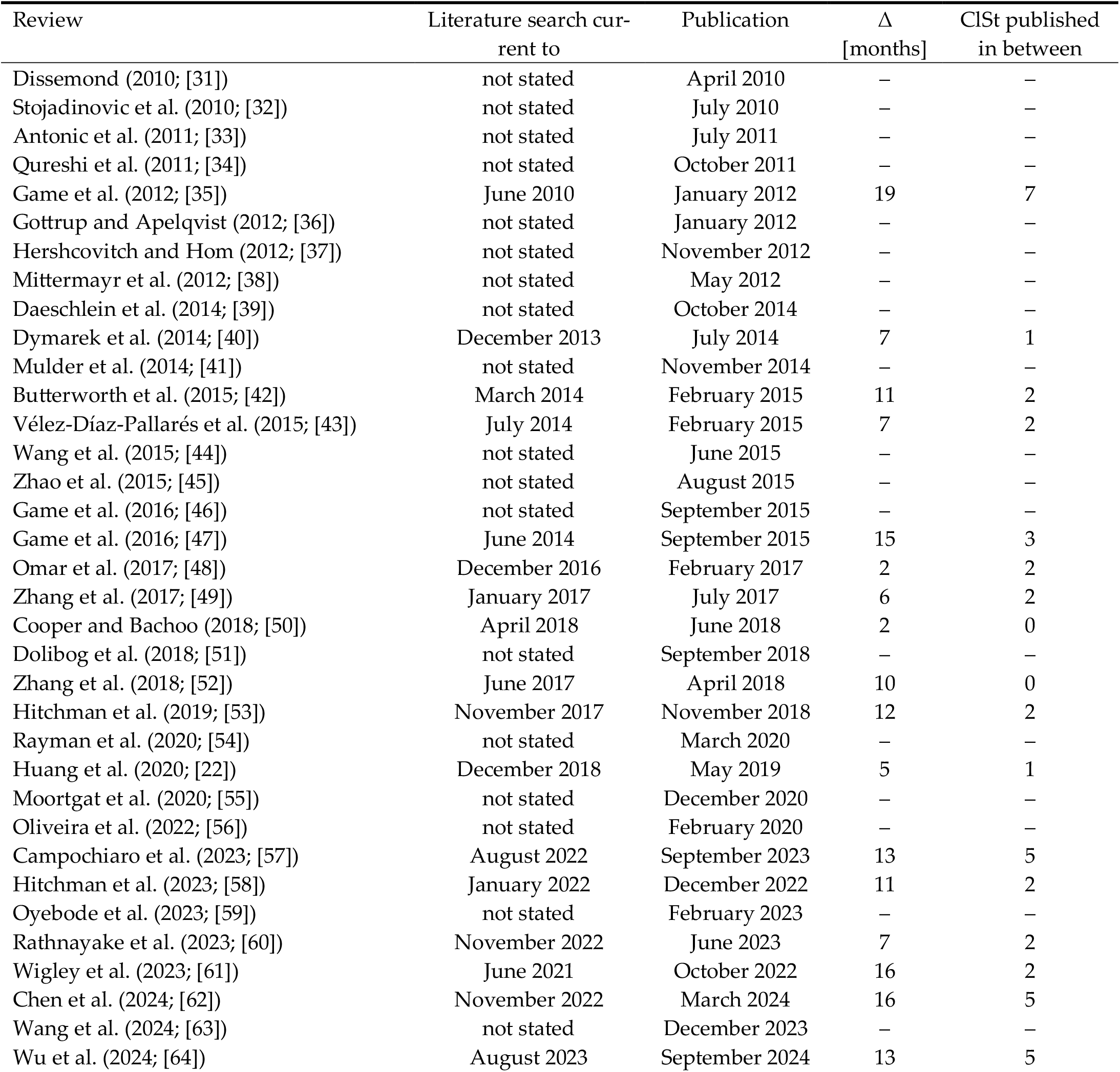
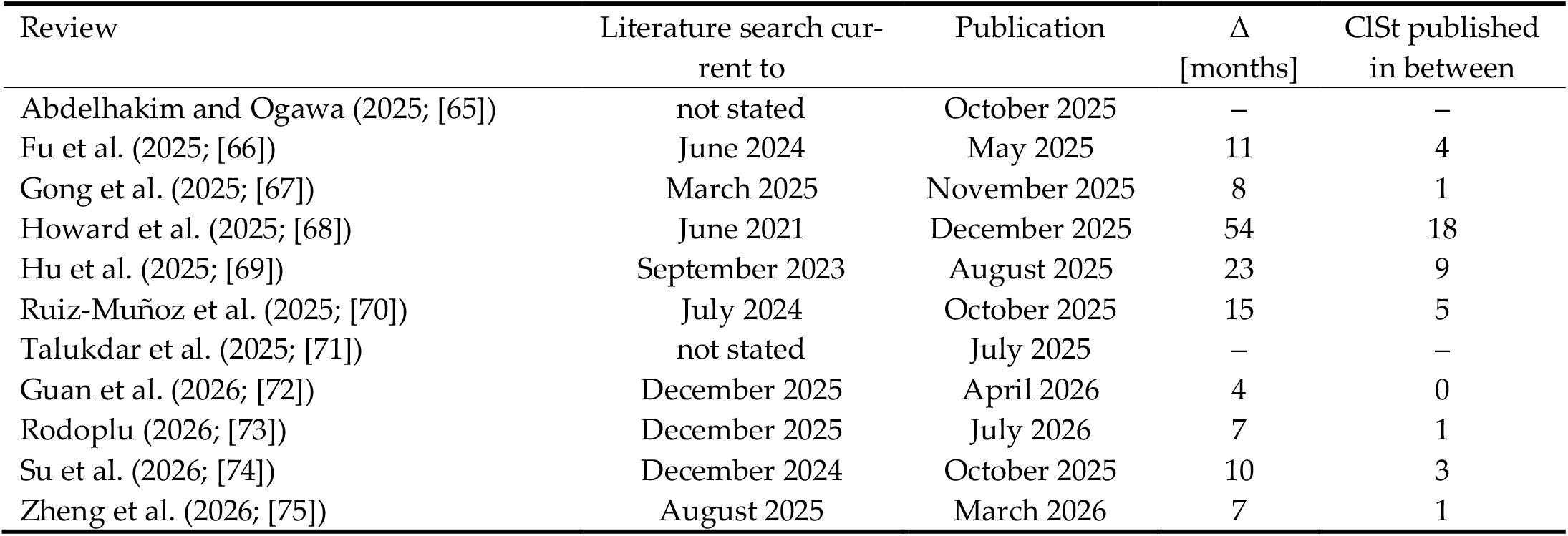
Currency of the literature at the time of publication for all reviews assessed in this systematic review. “Literature search current to” is the date up to which the review reported that its search included studies; where only the period covered by the search was given, the end of that period is shown. “Publication” is the date on which the article first became available, which for articles published online ahead of their issue may precede the year given in the citation. Δ is the interval between the two dates in whole months. “ClSt published in between” is the number of clinical studies among [23–25,27–30,76–119] that appeared after the search date and on or before publication, that is, evidence that already existed when the review was published but could not have been covered by it. Abbreviations: ClSt, clinical study; Δ, interval between the literature search and publication. Note: Twenty of the 46 reviews (43.5%) reported no date to which their literature search was current. For these, neither Δ nor the number of clinical studies published in between could be determined; the reference date used elsewhere in this review (Section 2) was derived from the date of submission or from the date of publication minus six months and is not a search date.

| Review | Literature search current to | Publication | $\Delta$<br>[months] | CISt published in between |
| --- | --- | --- | --- | --- |
| Dissemond (2010; [31]) | not stated | April 2010 | – | – |
| Stojadinovic et al. (2010; [32]) | not stated | July 2010 | – | – |
| Antonic et al. (2011; [33]) | not stated | July 2011 | – | – |
| Qureshi et al. (2011; [34]) | not stated | October 2011 | – | – |
| Game et al. (2012; [35]) | June 2010 | January 2012 | 19 | 7 |
| Gottrup and Apelqvist (2012; [36]) | not stated | January 2012 | – | – |
| Hershcovitch and Hom (2012; [37]) | not stated | November 2012 | – | – |
| Mittermayr et al. (2012; [38]) | not stated | May 2012 | – | – |
| Daeschlein et al. (2014; [39]) | not stated | October 2014 | – | – |
| Dymarek et al. (2014; [40]) | December 2013 | July 2014 | 7 | 1 |
| Mulder et al. (2014; [41]) | not stated | November 2014 | – | – |
| Butterworth et al. (2015; [42]) | March 2014 | February 2015 | 11 | 2 |
| Vélez-Díaz-Pallarés et al. (2015; [43]) | July 2014 | February 2015 | 7 | 2 |
| Wang et al. (2015; [44]) | not stated | June 2015 | – | – |
| Zhao et al. (2015; [45]) | not stated | August 2015 | – | – |
| Game et al. (2016; [46]) | not stated | September 2015 | – | – |
| Game et al. (2016; [47]) | June 2014 | September 2015 | 15 | 3 |
| Omar et al. (2017; [48]) | December 2016 | February 2017 | 2 | 2 |
| Zhang et al. (2017; [49]) | January 2017 | July 2017 | 6 | 2 |
| Cooper and Bachoo (2018; [50]) | April 2018 | June 2018 | 2 | 0 |
| Dolibog et al. (2018; [51]) | not stated | September 2018 | – | – |
| Zhang et al. (2018; [52]) | June 2017 | April 2018 | 10 | 0 |
| Hitchman et al. (2019; [53]) | November 2017 | November 2018 | 12 | 2 |
| Rayman et al. (2020; [54]) | not stated | March 2020 | – | – |
| Huang et al. (2020; [22]) | December 2018 | May 2019 | 5 | 1 |
| Moortgat et al. (2020; [55]) | not stated | December 2020 | – | – |
| Oliveira et al. (2022; [56]) | not stated | February 2020 | – | – |
| Campochiaro et al. (2023; [57]) | August 2022 | September 2023 | 13 | 5 |
| Hitchman et al. (2023; [58]) | January 2022 | December 2022 | 11 | 2 |
| Oyebode et al. (2023; [59]) | not stated | February 2023 | – | – |
| Rathnayake et al. (2023; [60]) | November 2022 | June 2023 | 7 | 2 |
| Wigley et al. (2023; [61]) | June 2021 | October 2022 | 16 | 2 |
| Chen et al. (2024; [62]) | November 2022 | March 2024 | 16 | 5 |
| Wang et al. (2024; [63]) | not stated | December 2023 | – | – |
| Wu et al. (2024; [64]) | August 2023 | September 2024 | 13 | 5 |

| Review | Literature search current to | Publication | $\Delta$<br>[months] | ClSt published<br>in between |
| --- | --- | --- | --- | --- |
| Abdelhakim and Ogawa (2025; [65]) | not stated | October 2025 | – | – |
| Fu et al. (2025; [66]) | June 2024 | May 2025 | 11 | 4 |
| Gong et al. (2025; [67]) | March 2025 | November 2025 | 8 | 1 |
| Howard et al. (2025; [68]) | June 2021 | December 2025 | 54 | 18 |
| Hu et al. (2025; [69]) | September 2023 | August 2025 | 23 | 9 |
| Ruiz-Muñoz et al. (2025; [70]) | July 2024 | October 2025 | 15 | 5 |
| Talukdar et al. (2025; [71]) | not stated | July 2025 | – | – |
| Guan et al. (2026; [72]) | December 2025 | April 2026 | 4 | 0 |
| Rodoplu (2026; [73]) | December 2025 | July 2026 | 7 | 1 |
| Su et al. (2026; [74]) | December 2024 | October 2025 | 10 | 3 |
| Zheng et al. (2026; [75]) | August 2025 | March 2026 | 7 | 1 |

**Table A2.**
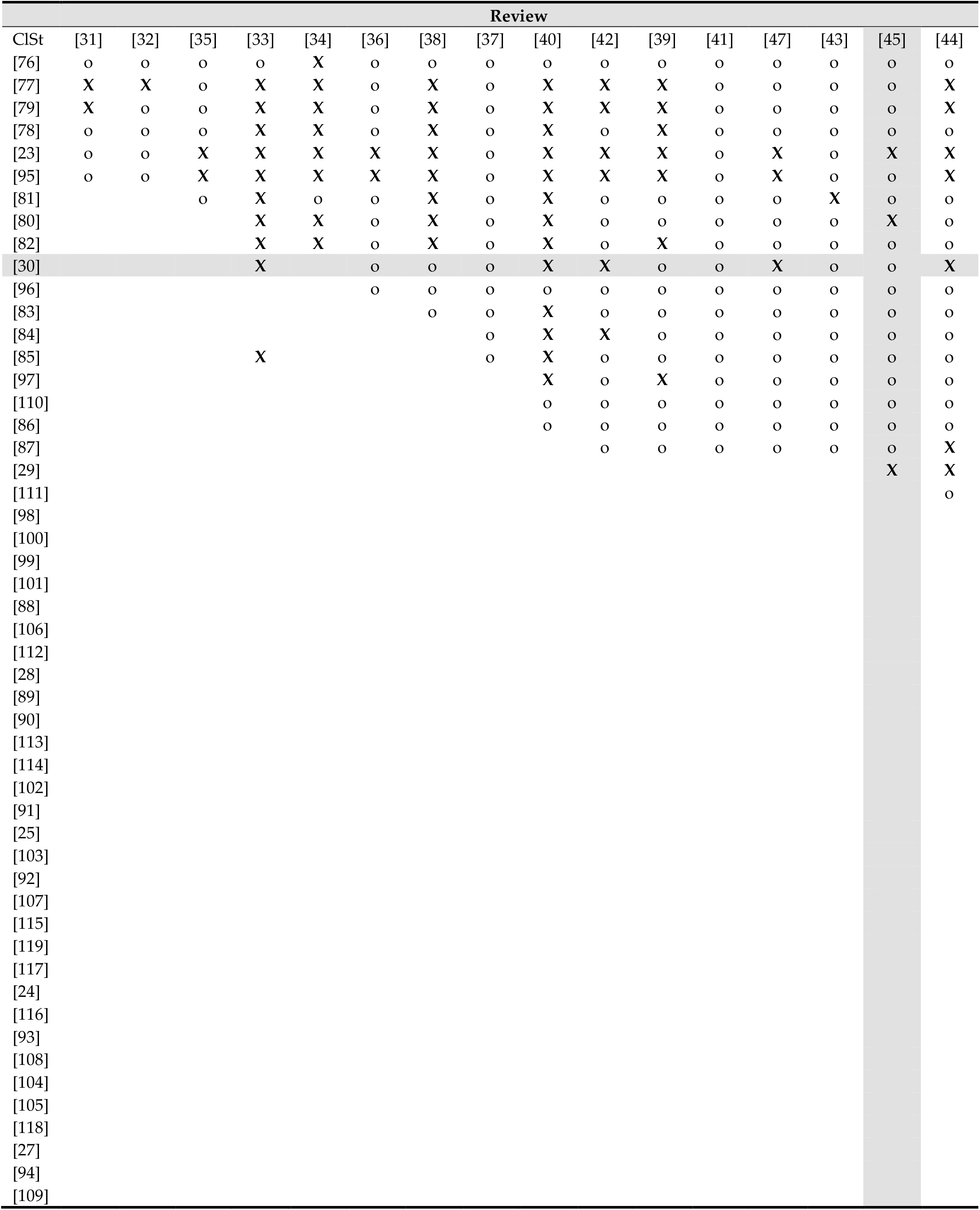
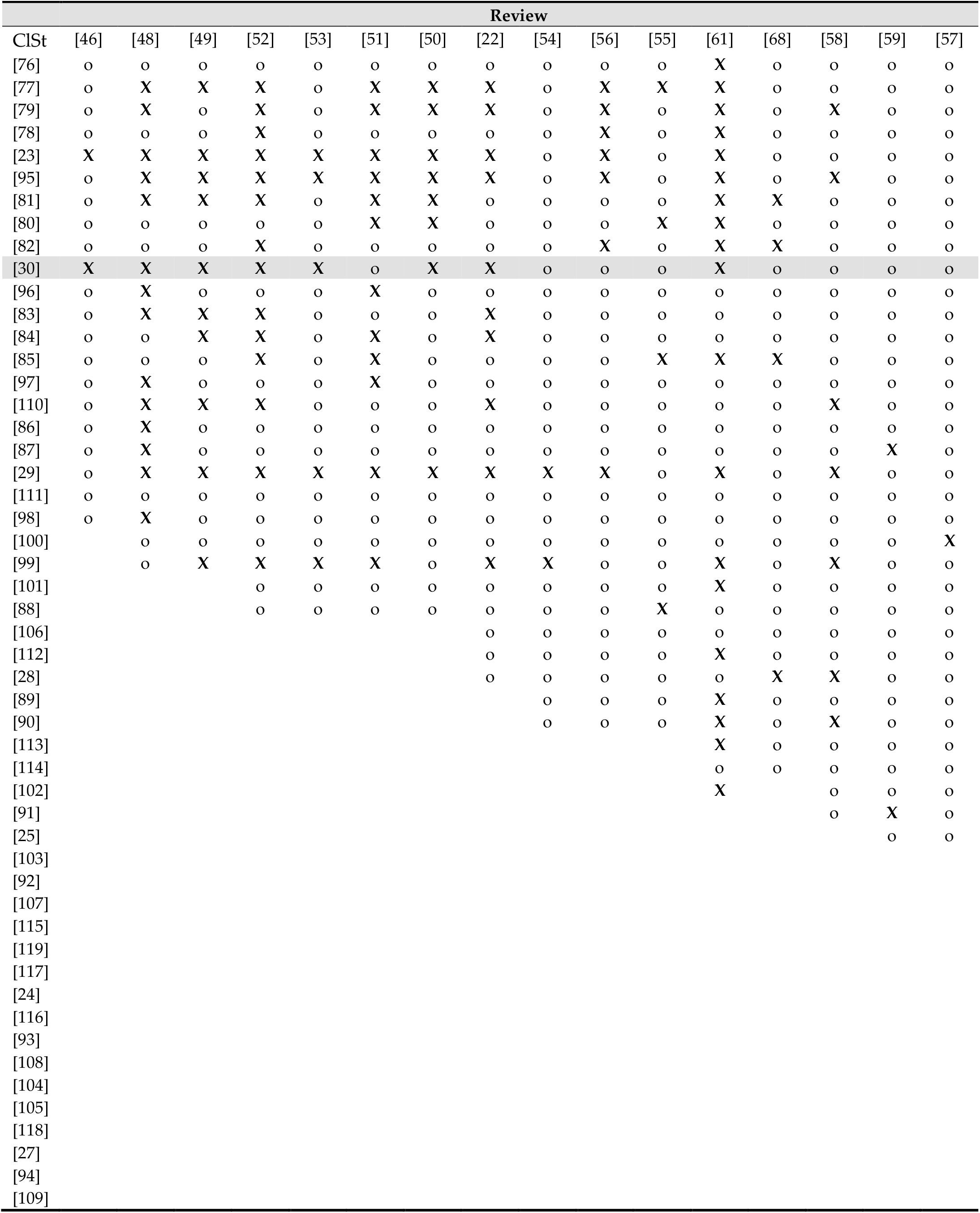
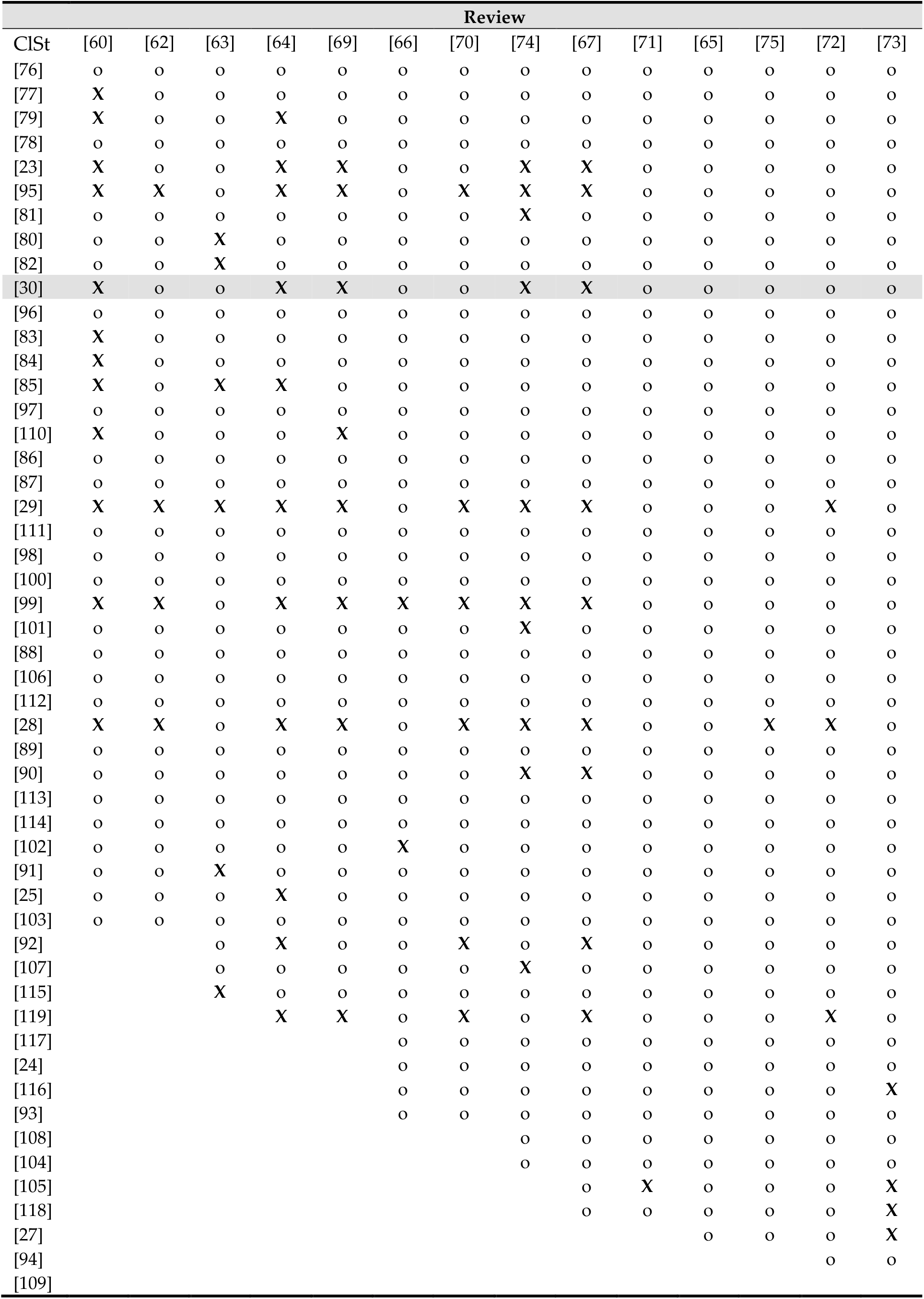
Matrix indicating which clinical studies [23–25,27–30,76–119] were included in each review [22,31–75] The matrix is presented in three panels. Abbreviations: ClSt, clinical study; lowercase “o”, the respective clinical study could have been cited in the corresponding review; uppercase, bold “X”, the clinical study was in fact cited in the respective review. **Explanation of Table A2 by means of an example:** the clinical study by Wang et al. (2011; [30]) was cited in the reviews by Antonic et al. (2011; [33]), Dymarek et al. (2014; [40]), Butterworth et al. (2015; [42]), Wang et al. (2015; [44]), Game et al. (2016; [46,47]), Omar et al. (2017; [48]), Zhang et al. (2017; [49]), Cooper and Bachoo (2018; [50]), Zhang et al. (2018; [52]), Hitchman et al. (2019; [53]), Huang et al. (2020; [22]), Rathnayake et al. (2023; [60]), Wigley et al. (2023; [61]), Wu et al. (2024; [64]), Gong et al. (2025; [67]), Hu et al. (2025; [69]) and Su et al. (2026; [74]) (indicated by gray fields in row [30]). Accordingly, this clinical study by Wang et al. (2011; [30]) was cited in 18 of the 42 reviews (42.9%) published after its release. The review by Zhao et al. (2015; [45]) was among those that did not cite the clinical study by Wang et al. (2011; [30]). In fact, it cited only 3 of the 19 clinical studies (15.8%) that had been published prior to its release: Wang et al. (2009; [23]), Arnó et al. (2010; [80]) and Omar et al. (2014; [29]) (indicated by gray fields in column [45]). Complete reference details are available in the main text. Table A1 lists, for every review, the date to which its literature search was current, its date of publication and the interval between the two. Note: Antonic et al. (2011; [33]) cited two clinical studies that appeared after their reference date, one of them as ‘in press’; these are shown as X below the availability boundary of the column.

| ClSt | Review |  |  |  |  |  |  |  |  |  |  |  |  |  |  |  |
| --- | --- | --- | --- | --- | --- | --- | --- | --- | --- | --- | --- | --- | --- | --- | --- | --- |
|  | [31] | [32] | [35] | [33] | [34] | [36] | [38] | [37] | [40] | [42] | [39] | [41] | [47] | [43] | [45] | [44] |
| [76] | o | o | o | o | X | o | o | o | o | o | o | o | o | o | o | o |
| [77] | X | X | o | X | X | o | X | o | X | X | X | o | o | o | o | X |
| [79] | X | o | o | X | X | o | X | o | X | X | X | o | o | o | o | X |
| [78] | o | o | o | X | X | o | X | o | X | o | X | o | o | o | o | o |
| [23] | o | o | X | X | X | X | X | o | X | X | X | o | X | o | X | X |
| [95] | o | o | X | X | X | X | X | o | X | X | X | o | X | o | o | X |
| [81] |  |  | o | X | o | o | X | o | X | o | o | o | o | X | o | o |
| [80] |  |  |  | X | X | o | X | o | X | o | o | o | o | o | X | o |
| [82] |  |  |  | X | X | o | X | o | X | o | X | o | o | o | o | o |
| [30] |  |  |  | X |  | o | o | o | X | X | o | o | X | o | o | X |
| [96] |  |  |  |  |  | o | o | o | o | o | o | o | o | o | o | o |
| [83] |  |  |  |  |  |  | o | o | X | o | o | o | o | o | o | o |
| [84] |  |  |  |  |  |  |  | o | X | X | o | o | o | o | o | o |
| [85] |  |  |  | X |  |  |  | o | X | o | o | o | o | o | o | o |
| [97] |  |  |  |  |  |  |  |  | X | o | X | o | o | o | o | o |
| [110] |  |  |  |  |  |  |  |  | o | o | o | o | o | o | o | o |
| [86] |  |  |  |  |  |  |  |  | o | o | o | o | o | o | o | o |
| [87] |  |  |  |  |  |  |  |  |  | o | o | o | o | o | o | X |
| [29] |  |  |  |  |  |  |  |  |  |  |  |  |  |  | X | X |
| [111] |  |  |  |  |  |  |  |  |  |  |  |  |  |  |  | o |
| [98] |  |  |  |  |  |  |  |  |  |  |  |  |  |  |  |  |
| [100] |  |  |  |  |  |  |  |  |  |  |  |  |  |  |  |  |
| [99] |  |  |  |  |  |  |  |  |  |  |  |  |  |  |  |  |
| [101] |  |  |  |  |  |  |  |  |  |  |  |  |  |  |  |  |
| [88] |  |  |  |  |  |  |  |  |  |  |  |  |  |  |  |  |
| [106] |  |  |  |  |  |  |  |  |  |  |  |  |  |  |  |  |
| [112] |  |  |  |  |  |  |  |  |  |  |  |  |  |  |  |  |
| [28] |  |  |  |  |  |  |  |  |  |  |  |  |  |  |  |  |
| [89] |  |  |  |  |  |  |  |  |  |  |  |  |  |  |  |  |
| [90] |  |  |  |  |  |  |  |  |  |  |  |  |  |  |  |  |
| [113] |  |  |  |  |  |  |  |  |  |  |  |  |  |  |  |  |
| [114] |  |  |  |  |  |  |  |  |  |  |  |  |  |  |  |  |
| [102] |  |  |  |  |  |  |  |  |  |  |  |  |  |  |  |  |
| [91] |  |  |  |  |  |  |  |  |  |  |  |  |  |  |  |  |
| [25] |  |  |  |  |  |  |  |  |  |  |  |  |  |  |  |  |
| [103] |  |  |  |  |  |  |  |  |  |  |  |  |  |  |  |  |
| [92] |  |  |  |  |  |  |  |  |  |  |  |  |  |  |  |  |
| [107] |  |  |  |  |  |  |  |  |  |  |  |  |  |  |  |  |
| [115] |  |  |  |  |  |  |  |  |  |  |  |  |  |  |  |  |
| [119] |  |  |  |  |  |  |  |  |  |  |  |  |  |  |  |  |
| [117] |  |  |  |  |  |  |  |  |  |  |  |  |  |  |  |  |
| [24] |  |  |  |  |  |  |  |  |  |  |  |  |  |  |  |  |
| [116] |  |  |  |  |  |  |  |  |  |  |  |  |  |  |  |  |
| [93] |  |  |  |  |  |  |  |  |  |  |  |  |  |  |  |  |
| [108] |  |  |  |  |  |  |  |  |  |  |  |  |  |  |  |  |
| [104] |  |  |  |  |  |  |  |  |  |  |  |  |  |  |  |  |
| [105] |  |  |  |  |  |  |  |  |  |  |  |  |  |  |  |  |
| [118] |  |  |  |  |  |  |  |  |  |  |  |  |  |  |  |  |
| [27] |  |  |  |  |  |  |  |  |  |  |  |  |  |  |  |  |
| [94] |  |  |  |  |  |  |  |  |  |  |  |  |  |  |  |  |
| [109] |  |  |  |  |  |  |  |  |  |  |  |  |  |  |  |  |
|  | [46] | [48] | [49] | [52] | [53] | [51] | [50] | [22] | [54] | [56] | [55] | [61] | [68] | [58] | [59] | [57] |
| [76] | o | o | o | o | o | o | o | o | o | o | o | X | o | o | o | o |
| [77] | o | X | X | X | o | X | X | X | o | X | X | X | o | o | o | o |
| [79] | o | X | o | X | o | X | X | X | o | X | o | X | o | X | o | o |
| [78] | o | o | o | X | o | o | o | o | o | X | o | X | o | o | o | o |
| [23] | X | X | X | X | X | X | X | X | o | X | o | X | o | o | o | o |
| [95] | o | X | X | X | X | X | X | X | o | X | o | X | o | X | o | o |
| [81] | o | X | X | X | o | X | X | o | o | o | o | X | X | o | o | o |
| [80] | o | o | o | o | o | X | X | o | o | o | X | X | o | o | o | o |
| [82] | o | o | o | X | o | o | o | o | o | X | o | X | X | o | o | o |
| [30] | X | X | X | X | X | o | X | X | o | o | o | X | o | o | o | o |
| [96] | o | X | o | o | o | X | o | o | o | o | o | o | o | o | o | o |
| [83] | o | X | X | X | o | o | o | X | o | o | o | o | o | o | o | o |
| [84] | o | o | X | X | o | X | o | X | o | o | o | o | o | o | o | o |
| [85] | o | o | o | X | o | X | o | o | o | o | X | X | X | o | o | o |
| [97] | o | X | o | o | o | X | o | o | o | o | o | o | o | o | o | o |
| [110] | o | X | X | X | o | o | o | X | o | o | o | o | o | X | o | o |
| [86] | o | X | o | o | o | o | o | o | o | o | o | o | o | o | o | o |
| [87] | o | X | o | o | o | o | o | o | o | o | o | o | o | o | X | o |
| [29] | o | X | X | X | X | X | X | X | X | X | o | X | o | X | o | o |
| [111] | o | o | o | o | o | o | o | o | o | o | o | o | o | o | o | o |
| [98] | o | X | o | o | o | o | o | o | o | o | o | o | o | o | o | o |
| [100] |  | o | o | o | o | o | o | o | o | o | o | o | o | o | o | X |
| [99] |  | o | X | X | X | X | o | X | X | o | o | X | o | X | o | o |
| [101] |  |  |  | o | o | o | o | o | o | o | o | X | o | o | o | o |
| [88] |  |  |  | o | o | o | o | o | o | o | X | o | o | o | o | o |
| [106] |  |  |  |  |  |  |  | o | o | o | o | o | o | o | o | o |
| [112] |  |  |  |  |  |  |  | o | o | o | o | X | o | o | o | o |
| [28] |  |  |  |  |  |  |  | o | o | o | o | o | X | X | o | o |
| [89] |  |  |  |  |  |  |  |  | o | o | o | X | o | o | o | o |
| [90] |  |  |  |  |  |  |  |  | o | o | o | X | o | X | o | o |
| [113] |  |  |  |  |  |  |  |  |  |  |  | X | o | o | o | o |
| [114] |  |  |  |  |  |  |  |  |  |  |  | o | o | o | o | o |
| [102] |  |  |  |  |  |  |  |  |  |  |  | X |  | o | o | o |
| [91] |  |  |  |  |  |  |  |  |  |  |  |  |  | o | X | o |
| [25] |  |  |  |  |  |  |  |  |  |  |  |  |  |  | o | o |
| [103] |  |  |  |  |  |  |  |  |  |  |  |  |  |  |  |  |
| [92] |  |  |  |  |  |  |  |  |  |  |  |  |  |  |  |  |
| [107] |  |  |  |  |  |  |  |  |  |  |  |  |  |  |  |  |
| [115] |  |  |  |  |  |  |  |  |  |  |  |  |  |  |  |  |
| [119] |  |  |  |  |  |  |  |  |  |  |  |  |  |  |  |  |
| [117] |  |  |  |  |  |  |  |  |  |  |  |  |  |  |  |  |
| [24] |  |  |  |  |  |  |  |  |  |  |  |  |  |  |  |  |
| [116] |  |  |  |  |  |  |  |  |  |  |  |  |  |  |  |  |
| [93] |  |  |  |  |  |  |  |  |  |  |  |  |  |  |  |  |
| [108] |  |  |  |  |  |  |  |  |  |  |  |  |  |  |  |  |
| [104] |  |  |  |  |  |  |  |  |  |  |  |  |  |  |  |  |
| [105] |  |  |  |  |  |  |  |  |  |  |  |  |  |  |  |  |
| [118] |  |  |  |  |  |  |  |  |  |  |  |  |  |  |  |  |
| [27] |  |  |  |  |  |  |  |  |  |  |  |  |  |  |  |  |
| [94] |  |  |  |  |  |  |  |  |  |  |  |  |  |  |  |  |
| [109] |  |  |  |  |  |  |  |  |  |  |  |  |  |  |  |  |

| ClSt | Review |  |  |  |  |  |  |  |  |  |  |  |  |  |
| --- | --- | --- | --- | --- | --- | --- | --- | --- | --- | --- | --- | --- | --- | --- |
|  | [60] | [62] | [63] | [64] | [69] | [66] | [70] | [74] | [67] | [71] | [65] | [75] | [72] | [73] |
| [76] | o | o | o | o | o | o | o | o | o | o | o | o | o | o |
| [77] | X | o | o | o | o | o | o | o | o | o | o | o | o | o |
| [79] | X | o | o | X | o | o | o | o | o | o | o | o | o | o |
| [78] | o | o | o | o | o | o | o | o | o | o | o | o | o | o |
| [23] | X | o | o | X | X | o | o | X | X | o | o | o | o | o |
| [95] | X | X | o | X | X | o | X | X | X | o | o | o | o | o |
| [81] | o | o | o | o | o | o | o | X | o | o | o | o | o | o |
| [80] | o | o | X | o | o | o | o | o | o | o | o | o | o | o |
| [82] | o | o | X | o | o | o | o | o | o | o | o | o | o | o |
| [30] | X | o | o | X | X | o | o | X | X | o | o | o | o | o |
| [96] | o | o | o | o | o | o | o | o | o | o | o | o | o | o |
| [83] | X | o | o | o | o | o | o | o | o | o | o | o | o | o |
| [84] | X | o | o | o | o | o | o | o | o | o | o | o | o | o |
| [85] | X | o | X | X | o | o | o | o | o | o | o | o | o | o |
| [97] | o | o | o | o | o | o | o | o | o | o | o | o | o | o |
| [110] | X | o | o | o | X | o | o | o | o | o | o | o | o | o |
| [86] | o | o | o | o | o | o | o | o | o | o | o | o | o | o |
| [87] | o | o | o | o | o | o | o | o | o | o | o | o | o | o |
| [29] | X | X | X | X | X | o | X | X | X | o | o | o | X | o |
| [111] | o | o | o | o | o | o | o | o | o | o | o | o | o | o |
| [98] | o | o | o | o | o | o | o | o | o | o | o | o | o | o |
| [100] | o | o | o | o | o | o | o | o | o | o | o | o | o | o |
| [99] | X | X | o | X | X | X | X | X | X | o | o | o | o | o |
| [101] | o | o | o | o | o | o | o | X | o | o | o | o | o | o |
| [88] | o | o | o | o | o | o | o | o | o | o | o | o | o | o |
| [106] | o | o | o | o | o | o | o | o | o | o | o | o | o | o |
| [112] | o | o | o | o | o | o | o | o | o | o | o | o | o | o |
| [28] | X | X | o | X | X | o | X | X | X | o | o | X | X | o |
| [89] | o | o | o | o | o | o | o | o | o | o | o | o | o | o |
| [90] | o | o | o | o | o | o | o | X | X | o | o | o | o | o |
| [113] | o | o | o | o | o | o | o | o | o | o | o | o | o | o |
| [114] | o | o | o | o | o | o | o | o | o | o | o | o | o | o |
| [102] | o | o | o | o | o | X | o | o | o | o | o | o | o | o |
| [91] | o | o | X | o | o | o | o | o | o | o | o | o | o | o |
| [25] | o | o | o | X | o | o | o | o | o | o | o | o | o | o |
| [103] | o | o | o | o | o | o | o | o | o | o | o | o | o | o |
| [92] |  |  | o | X | o | o | X | o | X | o | o | o | o | o |
| [107] |  |  | o | o | o | o | o | X | o | o | o | o | o | o |
| [115] |  |  | X | o | o | o | o | o | o | o | o | o | o | o |
| [119] |  |  |  | X | X | o | X | o | X | o | o | o | X | o |
| [117] |  |  |  |  |  | o | o | o | o | o | o | o | o | o |
| [24] |  |  |  |  |  | o | o | o | o | o | o | o | o | o |
| [116] |  |  |  |  |  | o | o | o | o | o | o | o | o | X |
| [93] |  |  |  |  |  | o | o | o | o | o | o | o | o | o |
| [108] |  |  |  |  |  |  |  | o | o | o | o | o | o | o |
| [104] |  |  |  |  |  |  |  | o | o | o | o | o | o | o |
| [105] |  |  |  |  |  |  |  |  | o | X | o | o | o | X |
| [118] |  |  |  |  |  |  |  |  | o | o | o | o | o | X |
| [27] |  |  |  |  |  |  |  |  |  |  | o | o | o | X |
| [94] |  |  |  |  |  |  |  |  |  |  |  |  | o | o |
| [109] |  |  |  |  |  |  |  |  |  |  |  |  |  |  |

