## Supplementary File 1 for "Extracorporeal Shock Wave Therapy for Wound Management: Clinical Evidence, Energy Delivery Parameters and Mechanistic Insights — A Systematic Review"

**Extracorporeal Shock Wave Therapy in Wound Management: A Comprehensive Systematic Review of Clinical Evidence, Modalities and Mechanisms**

by Brent Musolf, Carmen Nussbaum-Krammer, Michael O’Neal, Nicola Maffulli and Christoph Schmitz

**Supplementary File 1**

**Standardized summaries of all 46 reviews assessed in this systematic review**

**(numbers in brackets refer to the reference numbers in the main text)**

**Note on the standardized summaries:** Each summary condenses one review as it was published, together with the interpretation that its own authors placed on the literature they had assessed. The summaries therefore reproduce the position of the respective authors and the state of the field at the time of that publication, and not the assessment of the present systematic review. They follow a uniform structure (Motivation, Hypothesis, Methods, Meta-Analysis, Results and Conclusions), because the original abstracts differ widely in structure, in length and in the information they report, which makes direct comparison between publications difficult; the standardized form is intended to remove this obstacle.

**Abbreviations (in alphabetical order):** ABI, ankle-brachial index; AMT, amniotic membrane therapy; ASEPSIS, scoring system for postoperative wound infection; BBN, Bayesian belief network; CABG, coronary artery bypass grafting; CAP, cold atmospheric plasma; CASPe, Spanish Critical Appraisal Skills Programme; CDO, continuous diffusion of oxygen; CI, confidence interval; CINeMA, Confidence in Network Meta-Analysis; CONSORT, Consolidated Standards of Reporting Trials; COX-2, cyclooxygenase-2; CWI, Combat Wound Initiative; DFU(s), diabetic foot ulcer(s); DU(s), digital ulcer(s); EFD, energy flux density; EGF, epidermal growth factor; eNOS, endothelial nitric oxide synthase; EQ-5D, EuroQol five-dimension questionnaire; ERK, extracellular signal-regulated kinase; ESWT, extracorporeal shock wave therapy; ESWs, extracorporeal shock waves; EWMA, European Wound Management Association; EXER, exercise therapy; fESWT, focused ESWT; FREMS, frequency rhythmic electrical modulation system; GRADE, Grading of Recommendations, Assessment, Development and Evaluation; HbA1c, glycated hemoglobin; HBOT, hyperbaric oxygen therapy; HIF, hypoxia-inducible factor; I2, measure of statistical heterogeneity; IGF-1, insulin-like growth factor 1; IL, interleukin; iNOS, inducible nitric oxide synthase; IPC, intermittent pneumatic compression; IWGDF, International Working Group on the Diabetic Foot; LFU, low-frequency ultrasound; LLLT, low-level laser therapy; LT, light therapy; MD, mean difference; MeSH, Medical Subject Headings; NCLFU, non-contact low-frequency ultrasound; NF-κB, nuclear factor kappa B; NPWT, negative pressure wound therapy; OR, odds ratio; PAR, percentage area reduction; PCNA, proliferating cell nuclear antigen; PDGF, platelet-derived growth factor; PEDro, Physiotherapy Evidence Database; PICO, population, intervention, comparison and outcome; PRISMA, Preferred Reporting Items for Systematic Reviews and Meta-Analyses; ProCT, procalcitonin; PROSPERO, International Prospective Register of Systematic Reviews; PRP, platelet-rich plasma; PU(s), pressure ulcer(s); RANTES, regulated on activation, normal T cell expressed and secreted; RCT(s), randomized controlled trial(s); rESWT, radial ESWT; RoB, risk of bias; ROBINS-I, Risk Of Bias In Non-randomised Studies of Interventions; RR, risk ratio; SC, stem cells; SD, standard deviation; SIGN, Scottish Intercollegiate Guidelines Network; SMD, standardized mean difference; SSc, systemic sclerosis; SUCRA, surface under the cumulative ranking curve; SWC, standard wound care; TcPO₂, transcutaneous partial oxygen pressure; TEAEs, treatment-emergent adverse events; TENS, transcutaneous electrical nerve stimulation; TGF-β, transforming growth factor beta; TLR3, toll-like receptor 3; TNF-α, tumor necrosis factor alpha; TOT, topical oxygen therapy; uESWT, unfocused ESWT; VAC, vacuum-assisted closure; VAS, visual analogue scale; VEGF, vascular endothelial growth factor; VLUs, venous leg ulcers; VRT, virtual reality technology; WMDs, weighted mean differences; α-SMA, alpha smooth muscle actin.

**[31] Dissemond, J. Physikalische Therapien des chronischen Ulcus cruris. *Hautarzt* 2010, *61*(5), 387–396. https://doi.org/10.1007/s00105-009-1882-2.**

**Motivation:** Chronic wounds such as ulcus cruris represent a growing therapeutic challenge, particularly among elderly patients in industrialized nations. Standard wound care is often insufficient for complete healing, prompting the need to evaluate adjunctive physical therapies.

**Hypothesis:** While no singular hypothesis was tested, this review operated under the assumption that various physical modalities, including ESWT, can significantly enhance wound healing outcomes in patients with chronic leg ulcers.

**Methods:** This narrative review examined a broad spectrum of physical therapies applied in chronic wound management, with ESWT as one of several modalities analyzed. The review did not follow a systematic review protocol; no specific search strategy, list of databases or controlled key search terms were provided. Literature included was selected based on clinical relevance and availability at the time of publication. No quantitative synthesis was performed; instead, a qualitative summary of clinical trials, Cochrane reviews and expert consensus was presented. The scope encompassed multiple physical interventions, such as compression therapy, vacuum therapy, therapeutic ultrasound, ESWT, electrical and electromagnetic stimulation, photodynamic therapy, hydrotherapy and water-filtered infrared-A radiation.

**Meta-Analysis:** No meta-analysis was performed.

**Results:** Compression therapy was the only modality with robust, consistent evidence supporting its effectiveness and is widely regarded as standard care in VLUs. Vacuum-assisted closure has become a practical mainstay despite limited high-quality data. ESWT was presented as a promising innovation, with mechanistic studies demonstrating enhanced angiogenesis, increased local blood flow and upregulation of regenerative markers like VEGF and eNOS. In a clinical study of 30 patients with treatment-resistant wounds, ESWT achieved complete healing in 50% within six weeks, with the remainder showing marked improvement. A larger multicenter prospective study involving 208 patients reported complete epithelialization in 75% of cases after three ESWT sessions. Other therapies, including ultrasound, electrostimulation and photodynamic therapy, showed mixed results or suffered from methodological limitations and small sample sizes.

**Conclusions:** The review concluded that while many physical therapies offer potential benefits in chronic wound care, most (aside from compression therapy) lack sufficient scientific validation through large, high-quality trials. Among the evaluated modalities, ESWT was presented as an innovative approach for which, as for most physical therapies other than compression, good scientific data are not yet available. Its ability to stimulate neoangiogenesis and modulate inflammation highlights its therapeutic relevance, especially in complex or refractory wounds. Nevertheless, ESWT should currently be considered an adjunctive therapy, integrated into a comprehensive wound management strategy that addresses underlying pathologies such as venous insufficiency or arterial disease. Continued clinical research is essential to firmly establish its role and standardize treatment protocols.

**[32] Stojadinovic, A.; Elster, E.; Potter, B.K.; Davis, T.A.; Tadaki, D.K.; Brown, T.S.; Ahlers, S.; Attinger, C.E.; Andersen, R.C.; Burris, D.; Centeno, J.; Champion, H.; Crumbley, D.R.; Denobile, J.; Duga, M.; Dunne, J.R.; Eberhardt, J.; Ennis, W.J.; Forsberg, J.A.; Hawksworth, J.; Helling, T.S.; Lazarus, G.S.; Milner, S.M.; Mullick, F.G.; Owner, C.R.; Pasquina, P.F.; Patel, C.R.; Peoples, G.E.; Nissan, A.; Ring, M.; Sandberg, G.D.; Schaden, W.; Schultz, G.S.; Scofield, T.; Shawen, S.B.; Sheppard, F.R.; Stannard, J.P.; Weina, P.J.; Zenilman, J.M. Combat Wound Initiative program. *Mil. Med.* 2010, *175*(7 Suppl.), 18–24. https://doi.org/10.7205/milmed-d-10-00156.**

**Motivation:** Combat-related injuries frequently result in complex, contaminated and nonhealing wounds that are poorly responsive to conventional surgical and wound care strategies. Given the limitations of current treatments, there is a critical need for innovative modalities that accelerate healing, minimize complications and support the functional recovery of military personnel.

**Hypothesis:** Although a specific hypothesis was not tested in this review, the authors advanced the premise that ESWT offers therapeutic benefits in the management of acute and chronic wounds through anti-inflammatory and pro-angiogenic mechanisms.

**Methods:** This publication presented a narrative review embedded within the Combat Wound Initiative (CWI) framework, focusing on the translational development and clinical application of ESWT for wound management, particularly in military trauma care. The review synthesized findings from preclinical animal models and human trials, as well as ongoing studies integrating molecular biomarker profiling and advanced bioinformatics. No systematic search strategy, database listing or predefined key search terms were provided. The analysis was qualitative and included bench-to-bedside translational evidence from institutional research programs. Clinical outcomes were interpreted using Bayesian belief network (BBN) modeling to predict wound healing responses.

**Meta-Analysis:** No meta-analysis was performed.

**Results:** Preclinical studies demonstrated that a single ESWT session significantly reduced inflammatory cell infiltration and suppressed gene expression of key cytokines and matrix metalloproteinases in a murine burn model. Additional experiments revealed enhanced angiogenesis and skin graft integration following ESWT, accompanied by upregulated expression of pro-angiogenic growth factors. Clinically, a phase II trial involving 208 patients with acute and chronic wounds treated with unfocused ESWT showed a 75% complete healing rate, with no treatment-related toxicity or complications. Wound size (≤10 cm^2^) and shorter duration (≤1 month) were independent predictors of complete epithelialization. A phase III RCT is underway to assess ESWT’s efficacy versus standard care in acute traumatic wounds, with endpoints including healing time, quality of life and biomarker changes. Biomarker analyses identified specific cytokines in serum and wound effluent (e.g., ProCT, IL-13, RANTES) associated with wound dehiscence or healing, supporting their use in predictive modeling.

**Conclusions:** The integrated clinical and translational efforts of the CWI underscore ESWT’s significant potential as a safe, noninvasive and biologically active adjunctive therapy for acute and chronic soft tissue wounds. ESWT has shown both anti-inflammatory and pro-angiogenic effects that translate into improved wound healing outcomes. While further RCTs are needed to confirm its superiority over standard treatments, ESWT currently represents one of the most promising emerging technologies in the field of combat and complex wound management. Its clinical utility is enhanced by favorable safety, ease of application and compatibility with precision medicine approaches that incorporate molecular diagnostics and decision support algorithms.

**[33] Antonic, V.; Mittermayr, R.; Schaden, W.; Stojadinovic, A. Evidence supporting extracorporeal shock wave therapy for acute and chronic soft tissue wounds. *Wounds* 2011, *23*(7), 204–215.**

**Motivation:** Chronic and complex soft tissue wounds remain a significant therapeutic challenge, particularly in patients with comorbidities or trauma-related injuries. Standard treatments are often time-consuming, costly and inconsistent in efficacy, highlighting the need for new, safe and cost-effective modalities that promote rapid and durable healing.

**Hypothesis:** While no explicit hypothesis was tested, this review assumed that ESWT exerts beneficial effects on acute and chronic soft tissue wounds by promoting angiogenesis, modulating inflammation and enhancing tissue regeneration.

**Methods:** This narrative review compiled and analyzed preclinical and clinical studies on ESWT in the treatment of acute and chronic soft tissue wounds, including burns, surgical wounds, VLUs and DFUs. The review did not describe a systematic search strategy, listed specific databases searched or defined key search terms. The synthesis was qualitative, with emphasis on outcomes related to wound healing time, angiogenic and inflammatory markers, as well as treatment safety and tolerability. The reviewed studies included both RCTs and mechanistic investigations in animal models and humans.

**Meta-Analysis:** No meta-analysis was performed.

**Results:** Preclinical studies demonstrated that ESWT suppresses early pro-inflammatory cytokine and chemokine expression, reduces leukocyte infiltration and enhances angiogenic signaling such as VEGF expression and nitric oxide production. These effects translated into improved tissue perfusion, accelerated revascularization of grafts and better wound integration. Clinically, a phase II feasibility study involving 208 patients with various acute and chronic wounds showed a 75% complete healing rate without treatment-related adverse events. ESWT significantly accelerated epithelialization of skin graft donor sites and burn wounds, with reductions in healing time and infection rates compared to standard care. In chronic wound populations, ESWT improved wound closure, reduced exudate and pain, and enhanced granulation tissue formation. In DFU patients, ESWT outperformed HBOT, demonstrating higher healing rates, faster epithelialization and greater increases in markers of angiogenesis and cell proliferation. Notably, a RCT showed 57% complete healing with ESWT versus 25% with HBOT, along with superior improvements in oxygenation and wound perfusion.

**Conclusions:** ESWT has emerged as a clinically effective, non-invasive and safe adjunctive therapy for the treatment of both acute and chronic soft tissue wounds. Its therapeutic benefits are linked to anti-inflammatory and pro-angiogenic mechanisms that promote tissue regeneration without surgical morbidity or systemic side effects. The reviewed evidence supports ESWT as a viable and cost-effective option, particularly in outpatient settings, with broad applicability across wound types. While the exact mechanisms remain under investigation, current data justify continued clinical integration and further research to standardize treatment protocols. In the field of wound management, ESWT holds substantial promise as a tool to improve outcomes and reduce the burden of chronic nonhealing wounds.

**[34] Qureshi, A.A.; Ross, K.M.; Ogawa, R.; Orgill, D.P. Shock wave therapy in wound healing. *Plast. Reconstr. Surg.* 2011, *128*(6), 721e–727e. https://doi.org/10.1097/PRS.0b013e318230c7d1.**

**Motivation:** Despite advances in wound management, acute and chronic wounds continue to impose a substantial clinical and economic burden due to delayed healing, infection risk and scarring. The potential of ESWT to accelerate healing and improve outcomes has garnered increasing attention, but its clinical application remains limited by gaps in understanding and inconsistent study design.

**Hypothesis:** While not testing a formal hypothesis, this review proposed that ESWT may serve as a safe and effective adjuvant modality in wound healing by promoting angiogenesis, reducing inflammation and modulating cellular behavior through mechanotransduction.

**Methods:** This review employed an evidence-based narrative approach, analyzing peer-reviewed preclinical and clinical studies from the preceding ten years on the application of ESWT for wound management. The authors searched Medline and Cochrane databases using MeSH terms and relevant keywords related to wounds, shock wave therapy and tissue regeneration. The inclusion criteria allowed for RCTs and non-randomized clinical studies, animal models and in vitro studies. Clinical evidence was graded according to the Oxford Centre for Evidence-Based Medicine levels; RCTs were assessed using CONSORT criteria. The focus was exclusively on ESWT for wound healing, not on general physical therapy modalities.

**Meta-Analysis:** No meta-analysis was performed.

**Results:** Preclinical studies demonstrated that ESWT stimulates expression of pro-healing molecules such as VEGF, nitric oxide synthase and proliferating cell nuclear antigen, and reduces pro-inflammatory cytokines and leukocyte infiltration. In diabetic animal models, ESWT improved perfusion and accelerated healing, although some studies reported delayed healing with multiple treatments. Clinical trials showed variable but promising outcomes: in a study of 208 patients with complex wounds, 75% achieved complete epithelialization after ESWT. Other clinical studies reported reduced healing time for graft donor sites, improved outcomes in DFUs and lower infection rates in surgical wounds. Comparative studies indicated that ESWT may outperform HBOT in DFUs. Applications in burns and skin flaps also revealed benefits, including improved perfusion and reduced necrosis, although most studies lacked long-term follow-up and uniform methodology. Across the reviewed literature, serious adverse effects were rare and ESWT was well tolerated.

**Conclusions:** ESWT emerges as a promising, non-invasive and safe adjunctive therapy for both acute and chronic wound care. It exerts beneficial effects via mechanotransductive and immunomodulatory mechanisms, supporting angiogenesis, reducing inflammation and enhancing cellular repair activity. Despite encouraging results, inconsistencies in treatment protocols, such as focus type, energy level, number of extracorporeal shock waves and treatment intervals, underscore the need for standardized protocols and well-powered RCTs. Nonetheless, ESWT holds significant potential to improve healing outcomes, particularly in complex or refractory wounds.

**[35] Game, F.L.; Hinchliffe, R.J.; Apelqvist, J.; Armstrong, D.G.; Bakker, K.; Hartemann, A.; Löndahl, M.; Price, P.E.; Jeffcoate, W.J. A systematic review of interventions to enhance the healing of chronic ulcers of the foot in diabetes. *Diabetes Metab. Res. Rev.* 2012, *28*(Suppl. 1), 119–141. https://doi.org/10.1002/dmrr.2246.**

**Motivation:** Chronic DFUs remain a major cause of morbidity, amputation and healthcare costs worldwide. Despite numerous emerging therapies, uncertainty persists regarding the most effective interventions for enhancing wound healing in these patients.

**Hypothesis:** While no hypothesis was tested, this systematic review aimed to evaluate the clinical efficacy and methodological quality of various therapies, including ESWT, to determine their effectiveness in promoting healing in chronic DFUs.

**Methods:** This study was a systematic review conducted by the International Working Group on the Diabetic Foot (IWGDF). It included RCTs, cohort studies, case-control studies and other controlled designs published between December 2006 and June 2010. The review focused exclusively on interventions for chronic DFUs, not on wound care in general. Studies were sourced from Medline and Embase, and were assessed for methodological quality using SIGN (Scottish Intercollegiate Guidelines Network) criteria. A total of 1322 publications were screened, with 43 studies ultimately included. Data were synthesized in a narrative format.

**Meta-Analysis:** No meta-analysis was performed.

**Results:** The evidence supporting most adjunctive wound therapies was generally poor or limited. Among physical modalities, ESWT was included under the category of electrical, electromagnetic, laser, shock wave and ultrasound therapies. One small RCT found no difference in healing at 20 weeks, although the time to healing in the small number of patients who healed was reported to be significantly shorter, but the overall quality of evidence was low and the findings could not be generalized. Compared with other modalities like HBOT and NPWT, which showed more consistent and higher-quality evidence of benefit, ESWT lacked sufficient supportive data. This review noted considerable methodological shortcomings in the ESWT studies, such as small sample sizes, lack of blinding, incomplete reporting of outcomes and inadequate statistical analyses. The review also emphasized that despite promising biological mechanisms, ESWT’s clinical impact remains unproven due to insufficient rigorous evidence.

**Conclusions:** This systematic review concluded that evidence for ESWT in the treatment of chronic DFUs remains weak and inconclusive. While preliminary data suggest that ESWT may have some therapeutic potential, , the lack of high-quality, adequately powered RCTs prevents strong clinical recommendations. In contrast, HBOT and, possibly, NPWT were the only interventions for which the authors found published evidence that could justify their use. The authors recommend that future research should focus on well-designed, high-quality RCTs to assess ESWT’s role in diabetic wound healing. Until then, ESWT should be considered investigational and its use restricted to clinical study settings.

**[36] Gottrup, F.; Apelqvist, J. Present and new techniques and devices in the treatment of DFU: A critical review of evidence. *Diabetes Metab. Res. Rev.* 2012, *28*(Suppl. 1), 64–71. https://doi.org/10.1002/dmrr.2242.**

**Motivation:** Despite extensive clinical use of advanced wound therapies for DFUs, their effectiveness remains uncertain due to a lack of high-quality evidence. This review aimed to critically evaluate the clinical efficacy of established and emerging technologies, including ESWT, for the treatment of DFUs.

**Hypothesis:** While no formal hypothesis was tested, this review was grounded in the premise that only interventions supported by high-level evidence (RCTs or meta-analyses) should inform changes in clinical practice for DFU management.

**Methods:** This narrative critical review was based on a synthesis of recent systematic reviews, Cochrane analyses and RCTs published in the field of DFU treatment. The review included techniques such as debridement, topical negative pressure, HBOT, growth factors, bioengineered skin and physical therapies including ESWT. The authors adopted Cochrane-level evidence standards (Level I RCTs and meta-analyses) and drew on the work of the International Working Group of the Diabetic Foot and the Patient Outcome Group. No search strategy or list of databases was explicitly described. The review used a qualitative synthesis of evidence and included comparisons across interventions based on study design, sample size, outcome measures and risk of bias.

**Meta-Analysis:** No meta-analysis was performed.

**Results:** Among all interventions assessed, ESWT was classified under physical modalities (electrical, electromagnetic, laser, ultrasound and ESWT). Although preclinical studies and pilot clinical data suggested biological plausibility for ESWT in enhancing angiogenesis and modulating inflammation, clinical trials yielded insufficient evidence of efficacy. Two studies showed no significant differences in healing outcomes, though a shorter healing time in a subset of patients was noted. Another low-quality study suggested that ESWT may outperform HBOT, but methodological limitations, including small sample sizes and lack of blinding, prevented reliable conclusions. Compared to HBOT, for which the authors found increasing evidence on the highest level of an effect in DFU patients, ESWT lacked robust clinical validation; the evidence for topical negative pressure was judged insufficient as well. Overall, this review emphasized that across the wound care field, many treatments are supported by studies with methodological weaknesses, limiting their generalizability and impact on clinical guidelines.

**Conclusions:** The authors concluded that there is currently insufficient high-level evidence to support the use of ESWT in routine DFU management. Although ESWT demonstrates theoretical and experimental promise, its clinical relevance remains unproven, especially in comparison to more established modalities. The review underscored the urgent need for better-designed clinical trials, with adequate sample sizes, clearly defined endpoints and robust methodology. Until such data are available, ESWT should be considered experimental and its use confined to research settings. Broader concerns about evidence quality in DFU treatment suggest that outcome measures and research standards in wound care require re-evaluation to generate clinically meaningful evidence.

**[37] Hershcovitch, M.D.; Hom, D.B. Update in wound healing in facial plastic surgery. *Arch. Facial Plast. Surg.* 2012, *14*(6), 387–393. https://doi.org/10.1001/2013.jamafacial.33.**

**Motivation:** Facial plastic surgeons increasingly face the challenge of managing complex wounds, particularly in irradiated or poorly vascularized tissues. Advances in adjunctive therapies such as ESWT offer novel opportunities to improve healing outcomes, yet clinical adoption is hampered by limited mechanistic clarity and variable evidence quality.

**Hypothesis:** While no explicit hypothesis was tested, the authors reviewed emerging evidence under the assumption that ESWT and other biologically active technologies stimulate cellular pathways involved in wound healing, offering practical benefits in clinical settings such as facial reconstruction.

**Methods:** This narrative review evaluated contemporary adjunctive therapies in wound healing relevant to facial plastic surgery. The review spanned multiple biologic and physical modalities, including growth factors, platelet-derived products, bioengineered skin, VAC, HBOT, electrostimulation and ESWT. The focus was on integrating findings from recent preclinical studies, clinical case series and a limited number of small trials. No formal search strategy, database list or inclusion/exclusion criteria were reported. The synthesis was qualitative and descriptive, based on peer-reviewed publications and selected illustrative case examples.

**Meta-Analysis:** No meta-analysis was performed.

**Results:** ESWT was highlighted as a noninvasive modality delivering high-energy acoustic extracorporeal shock waves to tissue, initially developed for lithotripsy. Its mechanism in wound healing remains incompletely understood but is hypothesized to involve cytoskeletal mechanotransduction, stimulation of gene expression and cellular proliferation. Preclinical and early-phase clinical studies suggested ESWT induces favorable changes in growth factor production and promotes healing in both acute and chronic wounds. Notably, ESWT was found to be safe, with no reported adverse events such as infection, bleeding or tissue damage. In parallel, the review described other adjunctive therapies (e.g., PRP, VAC, bioengineered grafts), but consistently noted that their benefits are only demonstrable when layered onto robust foundational wound care practices.

**Conclusions:** This review concluded that ESWT is a promising adjunct in wound healing, potentially affecting the expression of genes and the production of growth factors known to promote wound healing. However, its role in clinical practice remains investigational, particularly in facial plastic surgery. Given its favorable safety profile and mechanistic rationale, further RCTs are warranted to define optimal protocols, indications and patient populations. Importantly, the authors emphasized that advanced technologies such as ESWT should not replace, but rather complement, standard wound care principles including infection control, tissue oxygenation and debridement.

**[38] Mittermayr, R.; Antonic, V.; Hartinger, J.; Kaufmann, H.; Redl, H.; Téot, L.; Stojadinovic, A.; Schaden, W. Extracorporeal shock wave therapy (ESWT) for wound healing: Technology, mechanisms, and clinical efficacy. *Wound Repair Regen.* 2012, *20*(4), 456–465. https://doi.org/10.1111/j.1524-475X.2012.00796.x.**

**Motivation:** Chronic and nonhealing wounds present an increasing burden on global healthcare systems due to prolonged treatment needs, reduced patient quality of life and high costs. ESWT has emerged as a potential noninvasive, cost-effective approach to improve tissue repair and reduce treatment-associated burdens.

**Hypothesis:** Although this review did not formally test a hypothesis, it was based on the premise that ESWT stimulates cellular and molecular processes, including angiogenesis and anti-inflammatory responses, that can accelerate wound healing.

**Methods:** This narrative review synthesized findings from both preclinical and clinical studies exploring ESWT for soft tissue wound healing, with a focus on its mechanisms of action, physical parameters and clinical efficacy. The review did not follow a systematic search protocol; rather, it compiled and critically interpreted relevant literature and institutional clinical data. The authors described treatment parameters (e.g., energy flux density, number of extracorporeal shock waves, frequency) and addressed the challenges in standardizing protocols across studies. The review also reported on outcomes from an ongoing cohort of approximately 600 patients treated with unfocused ESWT.

**Meta-Analysis:** No meta-analysis was performed.

**Results:** Mechanistically, ESWT was shown to enhance tissue perfusion, upregulate angiogenic factors such as VEGF and nitric oxide, and stimulate endothelial progenitor cell recruitment, cell proliferation and extracellular matrix metabolism. It also suppresses proinflammatory pathways (e.g., NF-κB) and demonstrates antibacterial effects against wound pathogens. Clinical studies included patients with DFUs, VLUs, pressure sores, burns and skin graft donor sites. In a pivotal prospective study involving 208 patients, 75% achieved full wound epithelialization, with smaller and shorter-duration wounds responding better. Comparative trials showed ESWT to be more effective than HBOT in improving perfusion, reducing wound size and enhancing histologic markers of healing. Additional studies confirmed safety and feasibility, with no adverse events reported. One placebo-controlled crossover trial noted that ESWT-stimulated wounds improved after an initial increase in wound size, suggesting early debridement-like effects.

**Conclusions:** ESWT is a safe, noninvasive and promising therapy for the treatment of acute and chronic soft tissue wounds. The therapy exerts biologically active, tissue-regenerative effects through a combination of enhanced angiogenesis, modulation of inflammation and stem cell recruitment. Clinical evidence, although encouraging, is still limited by small sample sizes and variability in treatment protocols. Future large-scale, randomized controlled trials are required to standardize dosing parameters, validate efficacy and define optimal indications. Until then, ESWT represents an innovative adjunct to standard wound care with the potential to significantly enhance healing in complex wounds.

**[39] Daeschlein, G.; Lutze, S.; Arnold, A.; von Podewils, S.; Jünger, M. Stellenwert moderner physikalischer Behandlungsverfahren bei infizierten und kolonisierten Wunden in der Dermatologie. *Hautarzt* 2014, *65*(11), 949–959. https://doi.org/10.1007/s00105-014-3526-4.**

**Motivation:** The increasing prevalence of chronic wounds and tumor-related skin lesions necessitates the development of effective, well-tolerated and outpatient-compatible therapies. ESWT has recently gained attention as a promising adjunctive treatment for therapy-resistant ulcers in dermatology.

**Hypothesis:** While this review did not propose a formal hypothesis, it was based on the clinical assumption that ESWT, through its physical stimulation of wound tissue, accelerates wound healing and exerts at least indirect antimicrobial effects in chronic, colonized or infected wounds.

**Methods:** This narrative review focused on modern physical treatment modalities in dermatology, with specific attention to ESWT as part of a multimodal wound management approach. The review discussed ESWT alongside other physical treatments such as cold plasma, infrared-A therapy, electrostimulation and low-level laser therapy. While not systematic, the analysis drew from institutional clinical experiences and relevant literature. No detailed search strategy or list of databases was provided. The focus was on ESWT for chronic ulcer wounds, especially those resistant to standard treatments.

**Meta-Analysis:** No meta-analysis was performed.

**Results:** ESWT was found to be a noninvasive, cost-effective and outpatient-suitable therapy for chronic therapy-refractory ulcers. The authors described successful treatment outcomes in patients with VLUs using 100 ESWs per cm^2^ at 5 Hz, administered biweekly over a minimum of five sessions per patient. Approximately 70% of patients experienced wound area reduction; 50% achieved full wound closure. Mechanistically, ESWT was reported to promote angiogenesis, microcirculation, granulation tissue formation and pain reduction, while potentially reducing fibrin deposition and the need for antibiotics. Although the primary antimicrobial effect is indirect, via promotion of wound healing, direct bactericidal activity is currently under investigation. ESWT was well tolerated, caused minimal discomfort and could be applied rapidly and easily in outpatient settings. No severe side effects were noted. Contraindications included acute infections with deep tissue involvement (e.g., phlegmonous infections) or the presence of highly virulent pathogens in immunocompromised patients.

**Conclusions:** ESWT represents a safe, well-tolerated and effective adjunctive therapy for chronic, therapy-refractory wounds in dermatological settings. Its noninvasiveness, short application time and outpatient feasibility make it particularly suitable for long-term wound care. While its primary therapeutic value lies in stimulation of tissue regeneration, indirect antimicrobial activity and pain relief, potential direct bactericidal effects require further study. Given its clinical utility, ESWT may play a key role in future complex wound care regimens. However, larger RCTs are needed to standardize treatment parameters and confirm long-term benefits across diverse wound etiologies.

**[40] Dymarek, R.; Halski, T.; Ptaszkowski, K.; Słupska, L.; Rosińczuk, J.; Taradaj, J. Extracorporeal shock wave therapy as an adjunct wound treatment: A systematic review of the literature. *Ostomy Wound Manage.* 2014, *60*(7), 26–39.**

**Motivation:** Chronic and complex soft tissue wounds, including VLUs, DFUs, PUs and burns, are a persistent therapeutic challenge. Given the limitations of standard care, ESWT has emerged as a promising adjunctive method to accelerate healing and improve patient outcomes.

**Hypothesis:** This systematic review aimed to test the hypothesis that ESWT is a safe and effective adjunctive treatment for chronic and acute soft tissue wounds, promoting faster healing through biological mechanisms such as angiogenesis, anti-inflammatory modulation and tissue regeneration.

**Methods:**
This was a systematic review of the literature, including studies published between 2000 and 2013 that evaluated ESWT in human subjects with soft tissue wounds. Five databases (MEDLINE, PubMed, Scopus, EBSCOhost and PEDro) were searched using defined keywords. A total of 13 clinical studies (n = 919 patients) were included, comprising seven RCTs, one clinical controlled trial, three prospective clinical trials and two case reports. The included studies were assessed using Cochrane Collaboration criteria, focusing on study design, blinding, randomization, intervention protocols and outcomes such as wound closure, perfusion, granulation tissue formation and adverse events. The analysis distinguished between focused, unfocused and radial ESWT using electrohydraulic, electromagnetic or ballistic ESWT devices.

**Meta-Analysis:** No meta-analysis was performed.

**Results:** Across the reviewed studies, ESWT was shown to significantly improve wound healing outcomes, including higher rates of complete closure, faster epithelialization, increased granulation tissue, improved perfusion and reduced need for antibiotics compared to controls (standard wound care, sham ESWT or HBOT). For example, complete wound healing rates for chronic DFUs ranged from 31% to 57% in ESWT groups versus 22% to 33% in controls. A study on surgical wounds following coronary artery bypass grafting (CABG) showed significantly lower ASEPSIS scores and reduced need for antibiotics in the ESWT group. In patients with split-thickness skin graft donor sites and burns, ESWT reduced healing times by several days. The EFD ranged from 0.03 to 0.25 mJ/mm^2^, with most studies using 100–500 ESWs/cm^2^ at 4–5 Hz, once or twice weekly for 3–6 sessions. ESWT was generally well tolerated and safe, with few minor adverse events reported (e.g., transient erythema or localized pain). Histological and immunohistochemical analyses demonstrated upregulation of VEGF, eNOS and PCNA, as well as suppression of pro-inflammatory cytokines and apoptosis markers.

**Conclusions:** This systematic review supports ESWT as a safe, noninvasive and effective adjunctive therapy for both acute and chronic soft tissue wounds. ESWT promotes wound healing through multiple biological mechanisms, including angiogenesis, improved perfusion, enhanced cell proliferation and anti-inflammatory effects. While substantial clinical evidence exists, further large-scale, sham-controlled and multicenter RCTs are warranted to refine treatment parameters, assess long-term outcomes and develop evidence-based clinical guidelines for routine application.

**[41] Mulder, G.; Tenenhaus, M.; D’Souza, G.F. Reduction of diabetic foot ulcer healing times through use of advanced treatment modalities. *Int. J. Low. Extrem. Wounds* 2014, *13*(4), 335–346. https://doi.org/10.1177/1534734614557925.**

**Motivation:** DFUs are a growing global health concern due to their high prevalence, delayed healing, risk of amputation and associated socioeconomic burden. Although numerous standard and advanced therapies exist, wound healing remains suboptimal, driving interest in adjunctive modalities such as ESWT.

**Hypothesis:** No hypothesis was tested. The review was based on the premise that advanced and adjunctive modalities can shorten healing times in DFUs when they are matched to the wound and to the factors delaying its healing.

**Methods:** This narrative review surveyed peer-reviewed literature on emerging wound care treatments, covering therapies such as dressings, off-loading, hyperbaric oxygen therapy, growth factors, bioengineered skin constructs, electrical stimulation, pulsed electromagnetic fields, phototherapy and negative pressure wound therapy. The review included evidence from clinical studies and meta-analyses, drawing comparisons between standard wound care and adjunctive options. No formal search strategy or systematic selection process was applied.

**Meta-Analysis:** No meta-analysis was performed.

**Results:** Off-loading in total-contact casts or removable walkers was described as among the most studied and simplest treatments for DFUs, and dressings and antimicrobial products as the established elements of local care. Among bioengineered constructs, a 12-week randomized trial of a dermal construct reported closure rates of 30% with the active product and 18% with the control, and a trial of a composite graft in 208 patients reported complete wound healing in 63 patients (56%) compared with 36 patients (38%) in the control group at 12 weeks. Application of recombinant human platelet-derived growth factor to full-thickness lower-extremity diabetic ulcers was reported to increase complete healing significantly compared with placebo, with the note that this product had received a boxed warning from the US Food and Drug Administration. Under future treatments the review covered electrical stimulation, for which a trial reported a wound area reduction of 31% with stimulation versus 4% without, pulsed electromagnetic fields, recombinant erythropoietin, phototherapy and negative pressure wound therapy. ESWT was named once in this article, in the abstract, among the treatments that have been further proposed as potential options; no clinical study of ESWT was cited, no treatment parameters were given and no mechanism of action was described.

**Conclusions:** The authors concluded that although national and international guidelines on the treatment of the diabetic foot ulcer exist, there is no definitive or universal consensus on the choice of specific treatment modalities. Optimizing comorbidities and the disease state, hemodynamics, local and peripheral skin and wound care and metabolic challenges, while reducing biological and bacterial burden and minimizing trauma, was presented as the primary approach, followed by the choice of the most appropriate treatment material or product. The review made no recommendation on ESWT.

**[42] Butterworth, P.A.; Walsh, T.P.; Pennisi, Y.D.; Chesne, A.D.; Schmitz, C.; Nancarrow, S.A. The effectiveness of extracorporeal shock wave therapy for the treatment of lower limb ulceration: A systematic review. *J. Foot Ankle Res.* 2015, *8*, 3. https://doi.org/10.1186/s13047-014-0059-0.**

**Motivation:** Lower limb ulceration, particularly in diabetic populations, remains a major source of morbidity and healthcare burden worldwide. ESWT has recently emerged as a promising adjunctive modality, yet the quality and consistency of clinical evidence supporting its use in ulcer treatment remain unclear.

**Hypothesis:** This systematic review tested the hypothesis that ESWT improves healing outcomes in lower limb ulceration of neurovascular origin, especially when used alongside standard care.

**Methods:** This study was a systematic review conducted in accordance with PRISMA guidelines. Five electronic databases (MEDLINE, CINAHL, Web of Knowledge, Scopus and Ovid AMED) were searched for articles published through December 2013. All human studies assessing ESWT for neurovascular lower limb ulcers (e.g., diabetic, venous) were eligible, excluding pressure, burn or surgical wounds. The methodological quality of included studies was evaluated using the Downs and Black Quality Index Tool; Cohen’s d was calculated to estimate effect sizes where data permitted. The review used a narrative synthesis due to clinical and methodological heterogeneity.

**Meta-Analysis:** No meta-analysis was performed.

**Results:** Five studies met the inclusion criteria, comprising three RCTs, one quasi-experimental trial and one case-series, with overall methodological quality ranging from 38% to 63%. Four studies involved DFUs, while one included a broader ulcer mix (e.g., venous and arterial). ESWT protocols varied in frequency (3 to 10 sessions), energy levels (EFD 0.03–0.27 mJ/mm^2^) and total number of ESWs (100–500/cm^2^). In the only RCT comparing ESWT plus SWC with SWC alone, ESWT-treated patients had significantly higher complete healing rates (53.3%) and shorter healing time (60.8 vs. 82.2 days; p < 0.001) compared to controls. Other RCTs showed similar findings, with ESWT outperforming HBOT in healing rates (57% vs. 25%) and reducing non-responding ulcers (11% vs. 60%; p < 0.001). One study reported complete healing in 16 of 32 ulcers after six ESWT sessions. Safety data across studies indicated no serious adverse events, with ESWT generally well tolerated. However, the lack of blinding, inconsistent ulcer classification, poor external validity and variation in ESWT dosing reduced the certainty of conclusions.

**Conclusions:** Preliminary evidence suggests that ESWT is a safe and potentially effective adjunct to standard care for lower limb ulcers, particularly of diabetic origin. ESWT appears to enhance healing rates and reduce time to closure, outperforming conventional therapies in some studies. However, due to low to moderate study quality, heterogeneity in protocols and limited sample sizes, current evidence remains insufficient for firm clinical recommendations. Future high-quality RCTs with standardized protocols and validated outcome measures are required to establish ESWT’s role in ulcer management and to develop evidence-based treatment guidelines.

**[43] Vélez-Díaz-Pallarés, M.; Lozano-Montoya, I.; Abraha, I.; Cherubini, A.; Soiza, R.L.; O’Mahony, D.; Montero-Errasquín, B.; Cruz-Jentoft, A.J. Nonpharmacologic interventions to heal pressure ulcers in older patients: An overview of systematic reviews (The SENATOR-ONTOP Series). *J. Am. Med. Dir. Assoc.* 2015, *16*(6), 448–469. https://doi.org/10.1016/j.jamda.2015.01.083.**

**Motivation:** Pressure ulcers are highly prevalent in older adults, often leading to prolonged hospitalization, increased mortality and decreased quality of life. While pharmacologic treatments are commonly used, the effectiveness of nonpharmacologic interventions, such as ESWT, for PU healing in elderly populations remains uncertain and under-investigated.

**Hypothesis:** Although no formal hypothesis was tested, this review aimed to assess whether any nonpharmacologic intervention, including ESWT, improves PU healing outcomes in older patients, with a specific focus on complete ulcer healing as the critical outcome.

**Methods:** This study was an overview of systematic reviews within the EU-funded SENATOR-ONTOP project. Multiple databases (PubMed, Cochrane Database of Systematic Reviews, EMBASE and CINAHL) were searched from inception to October 2013, with an updated search of the Cochrane Database in July 2014. Reviews were included if they examined at least one comparative primary study involving nonpharmacologic PU interventions in older adults (≥65 years). Among the various adjunctive therapies examined, ESWT was represented by a single RCT. Risk of bias was assessed using Cochrane Collaboration criteria; the GRADE framework was used to evaluate the quality of evidence. Data synthesis was conducted narratively.

Meta-Analysis: Studies were pooled where possible; a fixed-effects model was used, or a random-effects model when I2 exceeded 50%. Pooling was possible for ultrasound and electrotherapy but not for ESWT, which was represented by a single RCT.

**Results:** Of 110 systematic reviews analyzed, 45 studies were included, encompassing interventions such as support surfaces, nutritional supplementation, electrotherapy and ESWT. Only one RCT involving ESWT in elderly patients with chronic PUs was identified. This crossover, placebo-controlled study (n=8) found no statistically significant difference in complete healing or healing time between ESWT and sham treatment. The trial was small and although the risk of bias was considered low, the GRADE evidence quality for ESWT was rated as very low due to imprecision, limited sample size and lack of replication. Across all evaluated nonpharmacologic interventions, electrotherapy was the only modality supported by low-quality evidence for improved healing rates. No other intervention, including ESWT, demonstrated consistent or generalizable efficacy for PU healing in older patients.

**Conclusions:** Current evidence is insufficient to support the routine use of ESWT or other nonpharmacologic therapies in the treatment of PUs in older adults. Although ESWT is biologically plausible and shows promise in younger populations and other ulcer types, the clinical utility in geriatrics remains unproven. Well-designed, adequately powered RCTs focusing specifically on older adults with PUs are needed to evaluate ESWT’s effectiveness, establish standardized treatment protocols and guide clinical practice. Until such evidence emerges, ESWT should be considered experimental in the geriatric pressure ulcer population.

**[44] Wang, C.J.; Cheng, J.H.; Kuo, Y.R.; Schaden, W.; Mittermayr, R. Extracorporeal shockwave therapy in diabetic foot ulcers. *Int. J. Surg.* 2015, *24*(Pt B), 207–209. https://doi.org/10.1016/j.ijsu.2015.06.024.**

**Motivation:** DFUs represent a severe and increasingly prevalent complication of diabetes mellitus, often resulting in prolonged treatment, infection, limb amputation and substantial healthcare costs. Despite numerous standard and adjunctive therapies, healing outcomes remain inconsistent and often unsatisfactory, creating a critical need for innovative, noninvasive treatment options.

**Hypothesis:** Although no formal hypothesis was tested, this review examined the premise that ESWT improves healing outcomes in chronic DFUs and may offer superior efficacy compared to other adjunctive methods such as HBOT.

**Methods:** This was a narrative review summarizing results from previously published clinical trials and cohort studies evaluating ESWT in DFUs and non-DFUs. No systematic search strategy or keyword documentation was performed. The review synthesized comparative and longitudinal data regarding ESWT’s efficacy, optimal dosage, safety profile and long-term outcomes, including head-to-head comparisons with HBOT.

**Meta-Analysis:** No meta-analysis was performed.

**Results:** Multiple studies reviewed showed favorable results for ESWT. A 2009 RCT found 53.3% complete healing in the ESWT group versus 33.3% in controls, with significantly shorter healing times (60.8 vs. 82.2 days; p < 0.001). Another study involving 38 DFU patients reported complete healing in 54% of ESWT-treated ulcers after 20 weeks, compared to 28.5% in controls. In a direct comparison with HBOT, ESWT yielded higher complete healing rates (57% vs. 25%; p = 0.003), better perfusion and enhanced histological markers of angiogenesis and cell proliferation. A 5-year follow-up cohort of 67 patients with chronic ulcers (both diabetic and non-diabetic) revealed sustained healing in over 55% of cases, though perfusion declined over time. Notably, ESWT remained well tolerated, with no reported serious adverse events in any of the referenced studies.

**Conclusions:** ESWT demonstrates consistent short- and long-term benefits for healing chronic DFUs, outperforming conventional therapies and HBOT in clinical outcomes such as wound closure, tissue perfusion and histological regeneration markers. Its noninvasive nature, safety and cost-effectiveness further support its utility as a viable adjunctive treatment. Nonetheless, broader implementation is limited by a lack of standardized treatment protocols and variability in study design. The authors advocated for larger, well-controlled RCTs to confirm optimal dosing, frequency and timing of ESWT in the diabetic wound care algorithm. Until such guidelines are established, ESWT should be considered a promising alternative, especially in patients with therapy-refractory DFUs.

**[45] Zhao, J.; Xue, Y.; Yu, J.; Shi, K.; Xian, C.; Zhou, X. Advances in the research of mechanism of enhancement of wound healing with extracorporeal shock wave therapy. *Zhonghua Shao Shang Za Zhi* 2015, *31*(4), 315–317.**

**Motivation:** Wound healing, particularly in chronic and complex wounds, remains a major clinical challenge. ESWT has emerged as a non-invasive, safe and effective adjunct in wound management, yet the underlying mechanisms of its therapeutic effects are not fully elucidated.

**Hypothesis:** This review aimed to investigate whether ESWT promotes wound healing through specific cellular and molecular mechanisms that enhance tissue regeneration, angiogenesis and modulation of inflammation.

**Methods:** This was a narrative review focused exclusively on the mechanisms of ESWT for wound management. The review did not report a systematic search strategy, nor did it list specific databases searched, keywords used or selection criteria. Instead, it synthesized data from a broad range of previously published basic science and clinical studies on ESWT, encompassing both in vitro and in vivo research. The synthesis was qualitative in nature.

**Meta-Analysis:** No meta-analysis was performed.

**Results:** This review identified multiple biological effects of ESWT relevant to wound healing. Mechanistically, ESWT increases the expression of angiogenic factors such as vascular endothelial growth factor, endothelial nitric oxide synthase and transforming growth factor beta1, and promotes endothelial cell proliferation, migration and capillary formation. It also stimulates the activation of signaling pathways including Ras/ERK and HIF-1α, enhances stem cell recruitment and induces nitric oxide production. Additionally, ESWT modulates inflammatory responses by downregulating pro-inflammatory cytokines (e.g., IL-1β, TNF-α), thereby promoting a regenerative wound microenvironment. Antibacterial and microcirculatory benefits were also described.

**Conclusions:** This review supported the biological plausibility and multifaceted regenerative potential of ESWT for wound management. Although clinical evidence is still evolving, the cellular and molecular mechanisms outlined provide a strong rationale for its integration into wound management protocols. ESWT may be especially valuable in difficult-to-treat wounds due to its ability to simultaneously enhance angiogenesis, regulate inflammation and improve tissue perfusion.

**[46]	Game, F.L.; Apelqvist, J.; Attinger, C.; Hartemann, A.; Hinchliffe, R.J.; Löndahl, M.; Price, P.E.; Jeffcoate, W.J.; on behalf of the International Working Group on the Diabetic Foot (IWGDF). IWGDF guidance on use of interventions to enhance the healing of chronic ulcers of the foot in diabetes. *Diabetes Metab. Res. Rev.* 2016, 32(Suppl. 1), 75–83. https://doi.org/10.1002/dmrr.2700.**

**Motivation:** Chronic ulcers of the foot in diabetes are associated with protracted healing, infection and amputation, and promoted adjunctive interventions continue to grow in number. The authors stated that evidence was needed to substantiate particular interventions, and that clinicians should not adopt newer, more expensive treatments unless these improved healing more than existing methods.

**Hypothesis:** No formal hypothesis was tested. For each intervention category the review asked whether an adjunctive treatment, including agents acting through alteration of the physical environment, such as ESWT, improved DFU healing compared with accepted standards of good quality care.

**Methods:** This narrative guidance document was derived from three previous IWGDF systematic reviews, published in 2008, 2012 and 2015. No search strategy, databases, search terms, date limits, PROSPERO registration or PRISMA flow diagram was reported. The Grading of Recommendations Assessment, Development and Evaluation (GRADE) system was used. Evidence quality was rated high, moderate or low from risk of bias, effect sizes and expert opinion, inconsistency, indirectness and imprecision being unassessable for much of the older data; each recommendation was rated strong or weak by that quality, the balance of benefits and harms, patient values and costs. Ten intervention categories matched those of the systematic reviews, ESWT grouped with electricity, magnetism and ultrasound as agents altering the physical environment.

**Meta-Analysis:** No meta-analysis was performed.

**Results:** Evidence on debridement was limited and at high risk of bias. A large single-blind RCT at low risk of bias comparing three dressings showed no difference in wound healing or new infection. Of two methodologically good RCTs of systemic HBOT, the larger showed treated patients more likely to heal within 12 months; a retrospective cohort study in 83 centers across 31 US states found no effect on amputation or healing. NPWT was associated with benefit in post-operative wounds in three RCTs subject to bias. Electrical stimulation, ultrasound, normothermic therapy, magnetism and laser therapy showed no convincing evidence of benefit. Two ESWT studies reported apparent superiority of ESWT over HBOT, but were limited by per protocol analysis and other methodological problems.

**Conclusions:** Nine recommendations were issued. Recommendation 8 stated that agents reported to have an impact on wound healing through alteration of the physical environment, including electricity, magnetism, ultrasound and shock waves, should not be selected in preference to accepted standards of good quality care (strong recommendation; low quality of evidence), and no evidence existed justifying adoption of any reported physical therapy in routine practice. Weak recommendations were made for systemic HBOT and NPWT in post-operative wounds. Three unresolved issues were identified: a low evidence base, trial design difficulties, and few data on effectiveness and cost-effectiveness.

**[47] Game, F.L.; Apelqvist, J.; Attinger, C.; Hartemann, A.; Hinchliffe, R.J.; Löndahl, M.; Price, P.E.; Jeffcoate, W.J.; International Working Group on the Diabetic Foot. Effectiveness of interventions to enhance healing of chronic ulcers of the foot in diabetes: A systematic review. *Diabetes Metab. Res. Rev.* 2016, *32*(Suppl. 1), 154–168. https://doi.org/10.1002/dmrr.2707.**

**Motivation:** DFUs remain a major global therapeutic and financial challenge. Despite numerous emerging therapies, the effectiveness of many remains uncertain, particularly for chronic wounds that are slow to heal. Therefore, an updated systematic review was commissioned by the International Working Group on the Diabetic Foot (IWGDF) to assess the evidence supporting interventions such as ESWT.

**Hypothesis:** This systematic review was designed to test whether any specific intervention, including ESWT, improves healing rates of chronic DFUs based on high-quality evidence.

**Methods:** This systematic review followed the methodology of previous IWGDF assessments. It included controlled clinical studies (RCTs and comparative cohort studies) evaluating interventions for healing of DFUs in adults with type 1 or type 2 diabetes. A comprehensive search of Medline and EMBASE (June 2010–June 2014) was conducted using structured terms, followed by independent assessment of study quality using SIGN criteria. A total of 2161 studies were screened, with 33 meeting inclusion criteria. Due to heterogeneity in interventions, designs and outcomes, data synthesis was narrative.

**Meta-Analysis:** No meta-analysis was performed.

**Results:** Two earlier trials and one newly identified study evaluated ESWT for DFUs. One small RCT found no significant difference in complete healing between ESWT and sham therapy at 20 weeks. Two other studies, both by the same authors, compared ESWT with HBOT and reported higher healing rates with ESWT, although the methodology was weak and endpoints included composite measures such as partial healing. The new study replicated previous findings but did not clarify whether it was a reanalysis or a new cohort. Collectively, the ESWT studies demonstrated limited sample sizes, inconsistent endpoints and methodological flaws, including open-label design, lack of blinding, per-protocol analysis and selective reporting. As a result, the quality of evidence for ESWT was rated as low and no clear conclusions could be drawn about its efficacy in DFU treatment. In contrast, some other modalities (e.g., negative pressure wound therapy for postoperative wounds) were supported by stronger evidence, but most interventions lacked sufficient data to support widespread adoption.

**Conclusions:** Despite early enthusiasm, ESWT currently lacks robust, high-quality evidence to support its routine use in the treatment of chronic DFUs. While small trials suggested it may be superior to HBOT in selected cases, these findings were undermined by methodological limitations and small sample sizes. This review underscored the ongoing need for rigorous, well-powered, blinded RCTs with clearly defined endpoints to evaluate physical adjuncts like ESWT in chronic wound management.

**[48] Omar, M.T.; Gwada, R.F.; Shaheen, A.A.; Saggini, R. Extracorporeal shockwave therapy for the treatment of chronic wound of lower extremity: Current perspective and systematic review. *Int. Wound J.* 2017, *14*(6), 898–908. https://doi.org/10.1111/iwj.12723.**

**Motivation:** Chronic wounds of the lower extremities, including DFUs and VLUs, remain a major clinical and economic burden. The lack of uniform success with standard therapies has driven interest in adjunctive treatments such as ESWT, which has shown promise but remains variably supported by clinical evidence.

**Hypothesis:** This systematic review sought to determine whether ESWT is an effective and safe adjunctive therapy to SWC for enhancing the healing of chronic wounds of the lower extremities.

**Methods:** This was a systematic review of clinical trials evaluating the efficacy of ESWT in chronic lower extremity wounds, published between 2000 and 2016. A total of 11 studies (7 RCTs, 1 controlled clinical trial and 3 case series) involving 925 patients were identified through 10 databases (including PubMed, MEDLINE, EMBASE, PEDro and Cochrane CENTRAL). Inclusion criteria covered randomized and quasi-experimental designs in adults with various chronic lower extremity ulcers. Two reviewers independently extracted data and assessed methodological quality using the PEDro scale; studies were classified by Sackett’s levels of evidence. A narrative synthesis was applied.

**Meta-Analysis:** No meta-analysis was performed.

**Results:** The majority of studies showed significant improvements in healing rates, wound size reduction and time to closure with ESWT compared to controls. Reported complete healing rates in ESWT-treated groups ranged from 31% to 83%, compared to 10% to 49% in control groups. Time to full epithelialization was consistently shorter in ESWT groups (e.g., 64.5 days vs. 81.2 days; p < 0.05). In comparative trials, ESWT outperformed HBOT in healing rate, perfusion improvement and fewer adverse events. Most studies used unfocused ESWT at doses between 0.03–0.11 mJ/mm^2^. Only three studies evaluated blood perfusion and all reported significant improvements following ESWT. No serious adverse events were reported, though one trial noted minor infection managed with antibiotics. Based on PEDro scoring, three studies were of good quality, five fair and three poor; common limitations included lack of blinding and absence of intention-to-treat analysis.

**Conclusions:** This review found mild to moderate evidence supporting the use of ESWT as a safe and effective adjunct to standard wound care in chronic lower extremity wounds. ESWT improves healing outcomes, reduces wound surface area and enhances blood perfusion with minimal risk of adverse effects. However, variability in treatment protocols, methodological weaknesses and short follow-up durations in many studies prevented definitive conclusions. High-quality, standardized and adequately powered RCTs are needed to confirm these findings and determine optimal ESWT parameters and cost-effectiveness in clinical practice.

**[49] Zhang, L.; Weng, C.; Zhao, Z.; Fu, X. Extracorporeal shock wave therapy for chronic wounds: A systematic review and meta-analysis of randomized controlled trials. *Wound Repair Regen.* 2017, *25*(4), 697–706. https://doi.org/10.1111/wrr.12566.**

**Motivation:** Chronic wounds, particularly DFUs and PUs, pose significant therapeutic and socioeconomic challenges due to their poor healing potential and high recurrence rates. Conventional therapies often fail to achieve satisfactory outcomes, prompting investigation into adjunctive treatments such as ESWT.

**Hypothesis:** This systematic review and meta-analysis was designed to test the hypothesis that ESWT, when used alongside SWC, significantly improves healing outcomes for chronic wounds compared to SWC alone.

**Methods:** This was a systematic review and meta-analysis of RCTs published between January 2000 and January 2017. Seven databases, including PubMed, EMBASE, Cochrane Central, PEDro and Medline, were searched using predefined terms related to ESWT and chronic wound healing. Seven RCTs involving 301 patients met the inclusion criteria. Trials were assessed using the Jadad scale; data were extracted independently by two reviewers. Meta-analyses were performed using Review Manager 5.3, calculating odds ratios (ORs), standard mean differences (SMDs) and 95% confidence intervals (CIs). Heterogeneity was evaluated with the I^2^ statistic and both fixed and random effects models were applied based on heterogeneity levels.

Meta-Analysis: Meta-analyses were performed with Review Manager 5.3.5, using a fixed-effects model when studies were homogeneous (p > 0.05, I^2^ < 50%) and a random-effects model otherwise.

**Results:** Meta-analysis revealed that ESWT significantly improved wound healing outcomes. The wound healing rate was 1.86 times higher in the ESWT group than in controls (OR = 2.86, 95% CI: 1.63–5.03; p < 0.001), with minimal heterogeneity (I^2^ = 0%). The percentage of wound healing area was increased by 30.5% in ESWT-treated patients (SMD = 30.5; 95% CI: 23.8–37.1; p < 0.001); healing time was shortened by 19 days (SMD = –19.1 days; 95% CI: –23.7 to –14.5; p < 0.001). Subgroup analyses showed consistent superiority of ESWT over both standard care and hyperbaric oxygen therapy. No serious adverse effects or complications were reported in any included study. Jadad scores ranged from 3 to 5, with most studies rated as high quality.

**Conclusions:** This systematic review provided robust evidence that ESWT is a safe and effective adjunctive therapy that accelerates healing of chronic wounds, improves wound closure rates and reduces healing time when compared with SWC alone. The positive effects of ESWT appear consistent across various wound etiologies and energy settings. However, further high-quality RCTs are needed to optimize treatment protocols, determine long-term efficacy and assess cost-effectiveness. Despite current limitations, ESWT represents a promising option in the multidisciplinary management of chronic wounds.

**[50] Cooper, B.; Bachoo, P. Extracorporeal shock wave therapy for the healing and management of venous leg ulcers. *Cochrane Database Syst. Rev.* 2018, *6*(6), CD011842.** [**https://doi.org/10.1002/14651858.CD011842.pub2**](https://doi.org/10.1002/14651858.CD011842.pub2)**.**

**Motivation:** VLUs are the most common form of chronic lower limb wounds, contributing significantly to morbidity, impaired quality of life and healthcare costs. ESWT has been proposed as a novel treatment modality for promoting healing in these wounds through stimulation of angiogenesis and reduction of inflammation, but its efficacy remains unclear.

**Hypothesis:** This Cochrane systematic review was conducted to assess the hypothesis that ESWT improves healing and clinical outcomes in patients with VLUs when compared to SWC or other interventions.

**Methods:** This was a systematic review following Cochrane methodology. A comprehensive search was conducted in April 2018 across multiple databases including the Cochrane Wounds Specialised Register, CENTRAL, MEDLINE, Embase and CINAHL Plus, along with ClinicalTrials.gov and the WHO ICTRP. Inclusion criteria were RCTs comparing focused or unfocused ESWT to placebo, no treatment, SWC or alternative therapies in adults with VLUs. Primary outcomes included time to complete healing, proportion of ulcers healed and adverse effects. Secondary outcomes included ulcer size reduction, quality of life, pain, exudate, recurrence and cost. No language or publication status restrictions were applied.

**Meta-analysis:** No meta-analysis was performed.

**Results:** Despite extensive literature screening of 235 records, no RCTs met the inclusion criteria for this review. Five candidate studies were excluded upon full-text review because the patient cohorts did not include VLUs, despite investigating ESWT in chronic wounds. Consequently, no data were available to assess the efficacy, safety or cost-effectiveness of ESWT in the management of VLUs.

**Conclusions:** This review found no RCTs evaluating ESWT specifically for VLUs, highlighting a significant evidence gap. While observational studies and non-RCTs suggest that ESWT may be a safe and potentially beneficial therapy for soft tissue wounds, including some cases of VLUs, such findings lack the methodological rigor required for clinical guidance. Future research should prioritize well-designed RCTs that incorporate current best-practice comparators such as multilayer compression therapy, include patient-centered outcome measures like quality of life and assess cost-effectiveness. Until such data become available, no recommendations can be made regarding the use of ESWT for VLUs in clinical practice.

**[51] Dolibog, P.; Franek, A.; Brzezińska-Wcisło, L.; Dolibog, P.; Wróbel, B.; Arasiewicz, H.; Chmielewska, D. Shockwave therapy in selected soft tissue diseases: A literature review. *J. Wound Care* 2018, *27*(9), 573–583. https://doi.org/10.12968/jowc.2018.27.9.573.**

**Motivation:** Soft tissue wounds such as DFUs, VLUs, PUs and burns are frequent and often challenging to heal with conventional methods alone. ESWT has emerged as a potential adjunctive modality, but treatment parameters and outcomes vary widely across studies, necessitating a consolidated review of existing evidence.

**Hypothesis:** Although no formal hypothesis was tested, the authors sought to evaluate whether ESWT, when used in combination with SWC, contributes meaningfully to the healing of soft tissue wounds of various etiologies.

**Methods:** This review systematically searched PubMed, Embase and Web of Science using multiple keyword combinations related to ESWT and wound healing. A total of 14 studies, including RCTs, controlled clinical trials, prospective case studies and case reports, were included. The review analyzed parameters such as energy density, number of ESWs, frequency, treatment regimen, type of wound and methods of generating ESWs. Data were synthesized qualitatively.

**Meta-analysis:** No meta-analysis was performed.

**Results:** This review encompassed 191 soft tissue wounds across diverse etiologies. Focused ESWT was most commonly applied for DFUs, with reported healing rates ranging from 25% to 53.3%, typically using energy densities of 0.03–0.11 mJ/mm^2^ and 100 ESWs/cm^2^, over 3 to 10 sessions. For VLUs, both fESWT and uESWT were used, with healing rates of 36–100% depending on protocol, duration and study design. PUs were generally treated with uESWT, with healing rates ranging from 55.5% to 71.4%, while burn wounds treated with uESWT demonstrated 80–100% healing, often after just one or two sessions. The use of rESWT was reported infrequently and typically in combination with fESWT. In one notable case report, rESWT and fESWT led to complete healing of diabetic gangrene. Across the studies, reported adverse events included pain during treatment, infection, micro-traumatic effects with petechiae, hematoma or seroma formation, and a local inflammatory reaction. However, details on standard wound care, cost-effectiveness and long-term outcomes were often lacking.

**Conclusions:** The findings support the beneficial role of ESWT as an adjunct to SWC for various chronic wounds, especially DFUs, VLUs, PUs and burns. ESWT appears to enhance tissue perfusion, promote angiogenesis and reduce healing time. However, inconsistencies in protocols, lack of parameter standardization and sparse data on standalone effectiveness preclude definitive conclusions. While ESWT is safe and well-tolerated, future research should focus on larger, controlled trials, standardize treatment parameters and evaluate long-term outcomes and cost-effectiveness. Until such evidence is available, ESWT should be considered a supportive tool within a broader wound management strategy.

**[52] Zhang, L.; Fu, X.B.; Chen, S.; Zhao, Z.B.; Schmitz, C.; Weng, C.S. Efficacy and safety of extracorporeal shock wave therapy for acute and chronic soft tissue wounds: A systematic review and meta-analysis. *Int. Wound J.* 2018, *15*(4), 590–599. https://doi.org/10.1111/iwj.12902.**

**Motivation:** Acute and chronic soft tissue wounds, including DFUs, VLUs, PUs and burns, represent a growing public health challenge due to their delayed healing, pain and risk of infection. Standard wound care often falls short, prompting interest in adjunctive modalities such as ESWT, which has shown regenerative potential in musculoskeletal and ischemic conditions.

**Hypothesis:** This systematic review and meta-analysis tested the hypothesis that ESWT enhances wound healing outcomes, compared to SWC alone, in patients with acute and chronic soft tissue wounds.

**Methods:** The review included RCTs published before June 2017, identified through PubMed, Medline, Embase, Cochrane CENTRAL, Cochrane Library, PEDro and HealthSTAR. Inclusion required randomized allocation, comparison of ESWT versus SWC (± HBOT), wound healing as a monitored outcome, ≥80% participant retention and English language publication. The Cochrane risk-of-bias tool was used for quality assessment. Outcomes analyzed via RevMan 5.3.5 included healing rate, wound area reduction, healing time and infection incidence. Fixed or random-effects models were applied based on heterogeneity.

**Meta-analysis:** A meta-analysis was performed.

**Results:** Ten RCTs involving 473 patients were included. ESWT significantly improved wound healing outcomes versus SWC. The odds of complete healing were increased by 2.7-fold (OR = 3.7, 95% CI: 2.3–6.0, p < 0.001); the percentage reduction in wound area improved by 30.5% (SMD = 30.5; 95% CI: 23.8–37.1; p < 0.001). ESWT shortened healing time by 3 days for acute wounds and 19 days for chronic wounds (p < 0.001), and reduced the risk of wound infection by 53% (OR = 0.5, 95% CI: 0.2–0.9; p = 0.03). Heterogeneity was low for all but healing time. No serious adverse effects were reported; minor effects included local skin redness and mild discomfort. Subgroup analysis was performed on acute versus chronic wounds; the included trials applied fESWT in 5 studies, uESWT in 4 studies and rESWs in 1 study, with energy densities ranging from 0.03–0.23 mJ/mm^2^.

**Conclusions:** This systematic review and meta-analysis provided strong evidence that ESWT is a safe and effective adjunct to SWC for both acute and chronic soft tissue wounds. ESWT enhances healing rates, reduces wound size and infection risk, and accelerates recovery. While current findings are compelling, standardization of ESWT protocols and further well-powered, high-quality RCTs are needed to define optimal treatment regimens and assess long-term benefits and cost-effectiveness. Clinicians may consider ESWT particularly in cases refractory to conventional approaches.

**[53] Hitchman, L.H.; Totty, J.P.; Raza, A.; Cai, P.; Smith, G.E.; Carradice, D.; Wallace, T.; Harwood, A.E.; Chetter, I.C. Extracorporeal shockwave therapy for diabetic foot ulcers: A systematic review and meta-analysis. *Ann. Vasc. Surg.* 2019, *56*, 330–339. https://doi.org/10.1016/j.avsg.2018.10.013.**

**Motivation:** DFUs affect approximately 10% of diabetic patients and are associated with poor healing outcomes, risk of infection and limb loss. Despite SWC and advanced therapies like HBOT, healing rates remain suboptimal. ESWT has been suggested as a novel adjunctive treatment to improve healing outcomes in DFUs.

**Hypothesis:** This systematic review and meta-analysis tested the hypothesis that ESWT improves DFU healing compared to SWC or HBOT, evaluating outcomes such as healing rate, time to healing, perfusion and safety.

**Methods:** A comprehensive search was conducted through PubMed, Ovid MEDLINE, Embase, Web of Science, CINAHL Plus, Cochrane Central Register of Controlled Trials and ClinicalTrials.gov up to November 2017. Eligible studies were all clinical controlled trials involving patients older than 18 years with DFUs of at least 3 weeks’ duration; one of the five included studies was a controlled cohort study. The primary outcome was ulcer healing. Secondary outcomes included blood flow perfusion, infection rate, amputation rate and quality of life. Data extraction and bias assessment were performed independently by two reviewers using RevMan 5.3 and the Cochrane Risk of Bias Tool.

**Meta-analysis:** A meta-analysis was performed.

**Results:** Five studies involving 255 patients were included. Three trials compared ESWT with SWC; two compared ESWT with HBOT. ESWT was superior to SWC in terms of complete wound healing (odds ratio (OR) = 2.7; 95% CI: 1.0–6.9; I^2^ = 0%) and significantly shortened healing time (average 64.5 vs. 81.2 days; p < 0.05). ESWT also outperformed HBOT in healing outcomes (OR = 2.5; 95% CI: 1.1–5.61 I^2^ = 28%). Some studies reported increased blood perfusion, while one reported reduced bacterial load post-ESWT. No studies reported on amputation rates or quality of life; adverse effects were not observed. However, risk of bias was high or unclear in all studies due to poor blinding, allocation concealment and inconsistent reporting.

**Conclusions:** Preliminary evidence suggests that ESWT may improve healing in DFUs, performing better than both SWC and HBOT in small clinical trials. Nevertheless, the evidence is not robust enough to support routine clinical use. Key limitations include small sample sizes, methodological weaknesses, lack of blinding and heterogeneity in treatment protocols. Larger, high-quality RCTs with standardized ESWT parameters and comprehensive reporting are urgently needed to confirm efficacy and guide clinical implementation. Until then, ESWT remains an investigational therapy for DFUs.

**[54]	Rayman, G.; Vas, P.; Dhatariya, K.; Driver, V.; Hartemann, A.; Londahl, M.; Piaggesi, A.; Apelqvist, J.; Attinger, C.; Game, F.; on behalf of the International Working Group on the Diabetic Foot (IWGDF). Guidelines on use of interventions to enhance healing of chronic foot ulcers in diabetes (IWGDF 2019 update). *Diabetes Metab. Res. Rev.* 2020, 36(S1), e3283. https://doi.org/10.1002/dmrr.3283.**

**Motivation:** DFUs remain associated with protracted healing, infection, amputation and death, and consume rising health care resources, so interventions promoted to enhance healing need evidence of effectiveness. Four previous IWGDF systematic reviews had identified poor study design as the main obstacle to assessing these therapies; well-designed studies had since followed the 2016 IWGDF/EWMA 21-point checklist.

**Hypothesis:** No formal hypothesis was tested. The guideline used Population, Intervention, Comparator, Outcome (PICO) questions, one asking whether products altering wound biology through mechanical and physical means, including lasers, ESWT, ultrasound, magnetism and electric current, promote healing added to standard care.

**Methods:** This narrative guideline accompanied, but was published separately from, its supporting systematic review and followed GRADE methodology. An independent multidisciplinary working group installed by the IWGDF Editorial Board devised the clinical questions and defined critically important outcomes from Jeffcoate and colleagues' outcome set. The literature was assessed with the SIGN guideline, the Cochrane review system and the IWGDF/EWMA 21-point scoring system. Evidence per outcome was graded high, moderate or low from risk of bias, effect sizes, inconsistency and publication bias, and each recommendation was strong or weak, for or against the intervention. No search strategy, databases, search terms, date limits, PROSPERO registration or PRISMA flow diagram was reported. Thirteen recommendations resulted.

**Meta-Analysis:** No meta-analysis was performed.

**Results:** The review identified 97 published clinical trials between 2015 and 2019, versus 33 between 2011 and 2015. Two interventions were newly supported by single large RCTs: a sucrose-octasulfate impregnated dressing (adjusted odds ratio 2.6; 95% CI, 1.4-4.7, healing at week 20) and an autologous combined leukocyte, platelet and fibrin patch (complete healing 34% versus 22% of controls). Previous reviews had found nine studies of mechanical and physical therapies, including ESWT. The review added studies of ultrasound (one), ESWT (two), low-level laser therapy (three), a class IV laser (one), photodynamic therapy (two), infrared radiation (one) and pneumatic compression (one), all at high risk of bias or without evidence of benefit. One RCT of therapeutic magnetic resonance therapy was at low risk of bias but showed no benefit.

**Conclusions:** Recommendation 12 stated that agents reported to have an effect on ulcer healing through alteration of the physical environment, including electricity, magnetism, ultrasound and shock waves, should not be used in preference to best standard of care (strong recommendation; low quality of evidence), unchanged from the 2016 guidance. Poor study design left little evidence to recommend mechanical and physical therapies in hard-to-heal DFUs. New recommendations were added for sucrose-octasulfate dressings and placental-derived products. The recommendations were derived mostly from studies in specialist multidisciplinary foot clinics in high-income countries, their applicability elsewhere has remained unknown.

**[22]	Huang, Q.; Yan, P.; Xiong, H.; Shuai, T.; Liu, J.; Zhu, L.; Lu, J.; Shi, X.; Yang, K.; Liu, J. Extracorporeal shock wave therapy for treating foot ulcers in adults with type 1 and type 2 diabetes: A systematic review and meta-analysis of randomized controlled trials. *Can. J. Diabetes* 2020, *44*(2), 196–204.e3. https://doi.org/10.1016/j.jcjd.2019.05.006.**

**Motivation:** DFUs are among the most severe complications of diabetes, with high rates of morbidity, prolonged healing time and risk of amputation. Current treatments often fail to achieve complete healing, underscoring the need for effective adjunctive therapies. ESWT has emerged as a promising option, but comprehensive evidence for its efficacy in DFU treatment is limited.

**Hypothesis:** This systematic review and meta-analysis tested the hypothesis that ESWT improves wound healing outcomes, including wound surface area reduction, re-epithelialization and complete healing, in adults with DFUs compared to SWC or HBOT.

**Methods:** Following PRISMA guidelines and registered in PROSPERO (CRD42018118096), the authors searched PubMed, Embase, Web of Science, Cochrane Library, China Biology Medicine and relevant reference lists up to December 2018. Included studies were RCTs involving adult patients with DFUs treated with ESWT plus SWC versus SWC or SWC plus HBOT. Data were synthesized using StataSE 14.0, with outcomes expressed as risk ratios (RRs) or weighted mean differences (WMDs). Heterogeneity was assessed with I^2^ and chi-square tests; subgroup analyses were conducted where appropriate.

**Meta-analysis:** A meta-analysis was performed.

**Results:** Eight RCTs involving 339 patients were included. ESWT significantly increased the complete healing rate by 2.22-fold at the end of treatment (RR = 2.2; 95% CI: 1.5–3.4; p < 0.001) and reduced the proportion of unchanged ulcers by 4.8-fold (RR = 0.2; 95% CI: 0.1–0.4; p < 0.001). At follow-up, ESWT led to a mean reduction in wound surface area by 1.54 cm^2^ and an increase in re-epithelialization by 26.3%. Subgroup analyses demonstrated ESWT to be superior to both SWC (RR for complete healing = 2.4) and HBOT (RR = 1.8). ESWT also shortened healing time by 19 days in studies reporting this outcome. Adverse effects were minimal and transient, including local erythema, mild pain and hematoma, with no serious events reported.

**Conclusions:** This meta-analysis confirmed that ESWT is a safe and effective adjunctive therapy for DFUs, capable of accelerating wound closure, improving re-epithelialization and reducing treatment failure. It outperformed both SWC and HBOT in selected endpoints, offering an evidence-based option for treating chronic and refractory diabetic wounds. However, further large-scale RCTs are necessary to refine treatment protocols, explore early-phase efficacy and assess long-term outcomes and cost-effectiveness in diverse patient populations.

**[55] Moortgat, P.; Anthonissen, M.; Van Daele, U.; Meirte, J.; Vanhullebusch, T.; Maertens, K. Shock wave therapy for wound healing and scar treatment. In *Textbook on Scar Management: State of the Art Management and Emerging Technologies*; Téot, L.; Mustoe, T.A.; Middelkoop, E.; Gauglitz, G.G., Eds.; Springer: Cham, Switzerland, 2020; Chapter 55.**

**Motivation:** Chronic wounds and pathological scars remain major challenges in dermatology and reconstructive medicine. Conventional approaches offer limited success in promoting healing and restoring normal tissue architecture. ESWT has gained increasing interest for its regenerative effects in wound healing and scar modulation.

**Hypothesis:** This review tested the hypothesis that ESWT promotes tissue regeneration, modulates inflammation and improves structural and functional outcomes in soft tissue wounds and scars through mechanotransduction pathways.

**Methods:** This was a narrative review synthesizing preclinical and clinical evidence on ESWT's mechanisms, dose-response relationships and therapeutic effects in wound healing and scar treatment. The authors reviewed findings from cellular models, animal studies and clinical trials, with emphasis on angiogenesis, fibroblast activity, macrophage polarization, cytokine expression and scar remodeling. Key parameters such as EFD, number of ESWs per treatment session and frequency of treatment sessions were discussed in relation to therapeutic outcomes. No systematic search strategy was performed.

**Meta-analysis:** No meta-analysis was performed.

**Results:** Mechanistically, ESWT induces angiogenesis via upregulation of nitric oxide and vascular endothelial growth factor, and suppresses apoptosis in endothelial cells. It modulates macrophage polarization (enhancing M2 anti-inflammatory phenotype) and regulates inflammation through TLR3 and interleukin pathways. In fibroblasts and keratinocytes, ESWT activates gene expression involved in proliferation, cytoskeletal reorganization and extracellular matrix metabolism. In clinical trials, ESWT shortened epithelialization time in burn wounds, improved perfusion and accelerated closure of therapy-resistant ulcers. A referenced meta-analysis reported that ESWT increased healing rates 2.7-fold, reduced wound area by 30.5%, shortened healing time by 19 days for chronic wounds and reduced infection risk by 53%, compared to conventional wound therapy. In scar management, ESWT was associated with improvements in height, pliability, pigmentation, vascularity, range of motion and pain reduction. Histological studies revealed downregulation of pro-fibrotic markers (e.g., TGF-β1, alpha-SMA) and increased remodeling with thinner, better-aligned collagen fibers.

**Conclusions:** ESWT is a promising noninvasive therapy that enhances wound healing and facilitates scar remodeling through mechanotransduction, immunomodulation and angiogenesis. The therapy is safe, well-tolerated and adaptable to outpatient use, with favorable effects on both acute and chronic wounds, burn scars and hypertrophic scars. However, dose–response relationships remain underexplored and optimal treatment parameters (EFD, number of ESWs per treatment session and frequency of treatment sessions) must be defined for each indication. Future controlled studies should stratify results by tissue type, wound etiology and treatment settings to fully establish ESWT's clinical potential.

**[56] Oliveira, A.; Simões, S.; Ascenso, A.; Reis, C.P. Therapeutic advances in wound healing. *J. Dermatolog. Treat.* 2022, *33*(1), 2–22. https://doi.org/10.1080/09546634.2020.1730296.**

**Motivation:** Chronic and acute wounds, including DFUs, VLUs and burns, pose a growing clinical and economic burden due to delayed healing, pain, complications and the need for long-term care. With conventional therapies often failing to achieve satisfactory outcomes, ESWT has emerged as a non-invasive, adjunctive modality under active investigation.

**Hypothesis:** Although no formal hypothesis was tested, the authors explored the premise that ESWT accelerates healing of acute and chronic wounds through mechanotransduction and immunomodulation, improving clinical outcomes in otherwise treatment-resistant cases.

**Methods:** This narrative review was part of a broader article on emerging technologies in wound healing and included a section summarizing preclinical and clinical studies on ESWT. The review highlighted ESWT’s proposed mechanisms of action, technical parameters (e.g., focused vs. unfocused application, EFD, number of ESWs per treatment session) and its observed clinical effects in various wound types. The review did not describe a systematic search strategy, database list or search terms. Results were presented descriptively.

**Meta-analysis:** No meta-analysis was performed.

**Results:** Preclinical studies in diabetic rat and mouse models demonstrated that ESWT improves local perfusion, reduces inflammatory responses and enhances expression of angiogenesis-related genes. One animal study reported that multiple treatments with uESWT exacerbated the delayed wound healing after initially increasing the size of the wound. Clinical studies varied in protocol but commonly employed 100–1000 ESWs at approximately 0.1 mJ/mm^2^, administered weekly or biweekly. A multicenter trial of 208 patients with complicated wounds showed 75% complete wound epithelialization after a mean of three ESWT treatments. In one IIb study of 30 diabetic patients with foot ulcers, 53% of those treated with debridement followed by uESWT had complete wound closure after 20 weeks compared with SWC controls; in another study of 72 patients with chronic DFUs treated with ESWT, only 31% healed completely. For skin graft donor sites, ESWT significantly reduced epithelialization time. ESWT was well tolerated across studies, with no major adverse events reported.

**Conclusions:** ESWT showed consistent promise as a safe, non-invasive adjunctive therapy in the management of both acute and chronic wounds, particularly DFUs, VLUs and burns. It may improve angiogenesis, tissue perfusion and inflammation control through mechanotransduction. However, optimal treatment parameters remain undefined; larger RCTs are needed to confirm efficacy, explore long-term outcomes and establish clinical guidelines. Until then, ESWT should be considered a complementary intervention within comprehensive wound care protocols, particularly for non-healing or refractory wounds.

**[57] Campochiaro, C.; Suliman, Y.A.; Hughes, M.; Schoones, J.W.; Giuggioli, D.; Moinzadeh, P.; Baron, M.; Chung, L.; Ross, L.; Maltez, N.; Allanore, Y.; Denton, C.P.; Distler, O.; Frech, T.; Furst, D.E.; Khanna, D.; Krieg, T.; Kuwana, M.; Matucci-Cerinic, M.; Pope, J.; Alunno, A. Non-surgical local treatments of digital ulcers in systemic sclerosis: A systematic literature review. *Semin. Arthritis Rheum.* 2023, *63*, 152267. https://doi.org/10.1016/j.semarthrit.2023.152267.**

**Motivation:** Digital ulcers (DUs) are a common and burdensome manifestation in systemic sclerosis (SSc), contributing significantly to patient morbidity. While systemic pharmacological therapy remains the standard of care, there is a growing interest in local, non-surgical interventions such as ESWT to improve ulcer healing and minimize systemic side effects.

**Hypothesis:** This review was conducted to evaluate whether local, non-surgical therapies, including ESWT, offer safe and effective alternatives or adjuncts to systemic therapies for treating DUs in SSc patients.

**Methods:** This systematic review followed PRISMA guidelines and included original clinical studies (RCTs and observational studies) published up to August 29, 2022, focusing on local, non-surgical treatments for DUs in SSc patients. Databases searched included PubMed, Embase, MEDLINE, Cochrane Library, Web of Science, Emcare and Academic Search Premier. The PICO framework guided inclusion criteria; studies had to report DU outcomes in adults with SSc. The Cochrane RoB tool and ROBINS-I were used to assess bias. Owing to heterogeneity, findings were synthesized narratively.

**Meta-analysis:** No meta-analysis was performed.

**Results:** Of the 14 included studies, only one evaluated ESWT, a phase 2, single-arm prospective pilot study involving nine patients. ESWT demonstrated potential antalgic, anti-inflammatory and tissue regenerative effects. After four weeks, the number of DUs significantly decreased from 49 to 20 (p < 0.05), with continued reductions in DU dimensions (from 10.9 mm to 2.5 mm) and VAS pain observed at 20 weeks. No treatment-related complications were reported. Most patients in the ESWT study also received systemic vasodilator therapy. Other local treatments examined in the systematic review included botulinum toxin A (5 studies), hydrocolloid membranes, vitamin E gel, tadalafil cream, low-level laser therapy and dimethyl sulfoxide, each showing varying degrees of efficacy and safety. ESWT and vitamin E were among the most promising with high healing rates and favorable tolerability.

**Conclusions:** Although limited by small sample size and lack of a control group, ESWT showed encouraging results in reducing DU burden and pain in SSc patients. It represents a potentially safe and effective adjunctive local therapy, particularly for patients refractory to or intolerant of systemic treatments. However, the evidence base has remained weak. Larger, controlled trials are needed to validate these findings. ESWT may be considered as part of a multidisciplinary strategy for DU management, but standardized protocols and outcome definitions must be developed to enable robust comparisons and clinical recommendations.

**[58] Hitchman, L.; Totty, J.; Smith, G.E.; Carradice, D.; Twiddy, M.; Iglesias, C.; Russell, D.; Chetter, I.C. Extracorporeal shockwave therapy compared with standard care for diabetic foot ulcer healing: An updated systematic review. *Int. Wound J.* 2023, *20*(6), 2303–2320.** [**https://doi.org/10.1111/iwj.14035**](https://doi.org/10.1111/iwj.14035)**.**

**Motivation:** DFUs are a major complication of diabetes, associated with prolonged healing, risk of infection, amputation and substantial healthcare costs. ESWT has emerged as a non-invasive adjunctive therapy, yet its clinical effectiveness and optimal dosing remain uncertain.

**Hypothesis:** While no formal hypothesis was tested, this systematic review evaluated whether ESWT, in addition to SWC, improves healing outcomes for DFUs compared to standard care alone, and whether a dose-response relationship influences time to healing.

**Methods:** This was a systematic review registered on PROSPERO (CRD42022312509) and conducted according to PRISMA 2020 and Cochrane guidelines. A comprehensive search was conducted across eight databases (e.g., MEDLINE, PubMed, Embase and Cochrane CENTRAL) and gray literature sources up to January 28, 2022. Inclusion criteria were RCTs comparing ESWT plus SWC versus SWC or sham ESWT in adults with DFUs. Primary outcome was time to ulcer healing; secondary outcomes included healing rates, ulcer size reduction, recurrence, adverse events and cost-effectiveness. Risk of bias was assessed using the Cochrane Risk of Bias 2 tool and GRADE was used to evaluate certainty of evidence. Due to heterogeneity, narrative synthesis was performed.

**Meta-analysis:** No meta-analysis was performed.

**Results:** Six RCTs involving 471 patients were included. ESWT protocols varied widely in the number of ESWs applied (100–500/cm^2^), energy levels (0.03–0.23 mJ/mm^2^), frequency (1–3 sessions/week) and treatment duration (1–10 weeks). Four trials reported time to healing, but only two reported it in days and found it significantly shorter with ESWT (60.8 vs. 82.2 days; 64.5 vs. 81.2 days). Healing rates at 20–24 weeks were higher in ESWT-treated groups (e.g., 54% vs. 28.5%, 39% vs. 26%) across trials. ESWT also led to greater ulcer size reduction (e.g., 83% vs. 36%, 34.5% vs. 5.6%). Ulcer recurrence and adverse event rates were similar or slightly lower in ESWT arms, with no serious ESWT-related complications. However, no trials assessed quality of life or cost-effectiveness; all were rated at high or unclear risk of bias, leading to low certainty of evidence for all outcomes.

**Conclusions:** ESWT appears to enhance healing outcomes in DFUs when combined with standard care, but current evidence is limited by methodological heterogeneity, inconsistent reporting and high risk of bias. The review found no definitive evidence on dose-response relationships, despite biological plausibility from preclinical models. Standardization of trial design, ESWT dosing, outcome measures and reporting is urgently needed. Until then, the integration of ESWT into routine DFU care should proceed cautiously, preferably within research settings.

**[59] Oyebode, O.A.; Jere, S.W.; Houreld, N.N. Current therapeutic modalities for the management of chronic diabetic wounds of the foot. *J. Diabetes Res.* 2023, *2023*, 1359537. https://doi.org/10.1155/2023/1359537.**

**Motivation:** Chronic DFUs represent one of the most debilitating complications of diabetes, with high risk of infection, limb amputation and increased mortality. Effective, affordable and accessible therapeutic interventions are needed to improve healing outcomes and reduce healthcare burdens.

**Hypothesis:** While no formal hypothesis was tested, this review was grounded in the assumption that adjunctive therapies, including ESWT, significantly improve chronic DFU healing by targeting cellular and molecular dysfunctions inherent in diabetic wounds.

**Methods:** This was a narrative review that synthesized current knowledge on therapeutic approaches for managing chronic DFUs, with a dedicated section evaluating the evidence and rationale for ESWT. The review also covered wound debridement and dressing, transcutaneous electrical nerve stimulation (TENS), nanomedicine, HBOT and topical oxygen therapy, and low-level laser therapy. The authors did not perform a systematic literature search. Instead, representative clinical and preclinical studies were discussed, with mechanistic insights and clinical relevance highlighted.

**Meta-analysis:** No meta-analysis was performed.

**Results:** ESWT has shown promising short- and long-term outcomes in the treatment of chronic wounds. Clinical studies cited included one in which 67 patients received ESWT (500 ESWs at 0.11 mJ/mm^2^, 4 Hz, twice weekly for 3 weeks), with results indicating improved wound healing. Another study demonstrated that ESWT enhanced macrophage ERK activity, suggesting a mechanism for improved inflammatory resolution. While adverse events such as bruising and localized discomfort have been reported, ESWT is generally considered safe and well tolerated. Compared to other adjunctive therapies, ESWT offers the advantage of being noninvasive and mechanistically targeted toward key impairments in diabetic wound healing, increasing neoangiogenesis and proliferation and reducing inflammatory effects. However, the review acknowledged that insufficient high-quality evidence exists to support routine use in all patients, and optimal treatment parameters have remained undefined.

**Conclusions:** ESWT is an emerging therapeutic modality with demonstrated capacity to accelerate healing in chronic DFUs by promoting neoangiogenesis, modulating macrophage function and reducing inflammation. Despite encouraging findings, its routine clinical implementation is limited by a lack of standardized protocols and insufficient large-scale randomized controlled trials. Future research should aim to define patient selection criteria, optimal dosing regimens and comparative effectiveness versus other therapies. Nonetheless, ESWT stands out as a valuable noninvasive adjunct in the multimodal management of diabetic foot wounds.

**[60] Rathnayake, A.; Saboo, A.; Vangaveti, V.; Malabu, U. Electromechanical therapy in diabetic foot ulcers patients: A systematic review and meta-analysis. *J. Diabetes Metab. Disord.* 2023, *22*(2), 967–984. https://doi.org/10.1007/s40200-023-01240-2.**

**Motivation:** DFUs are a frequent and severe complication of diabetes mellitus, often leading to infection, limb amputation and high mortality. While standard therapies are essential, they are frequently insufficient due to the complex pathophysiology of DFUs. As such, adjunctive electromechanical therapies, including ESWT, have emerged as promising interventions warranting thorough evaluation.

**Hypothesis:** While no formal hypothesis was tested, this systematic review and meta-analysis was designed to assess whether electromechanical therapies, including ESWT, are effective adjuncts to standard care for promoting healing in DFUs, and to compare outcomes between experimental and control groups.

**Methods:** A systematic review and meta-analysis was conducted in accordance with PRISMA guidelines and the Cochrane Handbook for Systematic Reviews of Interventions. Databases searched included PubMed, Medline, Embase, Cochrane Library and Google Scholar for literature from 1990 to 2022. The search targeted studies involving electromechanical therapies (e.g., ESWT, ultrasound, phototherapy, electrical stimulation) in DFUs. Inclusion criteria followed the PICO framework and included RCTs and controlled trials in English. Risk of bias was assessed using Cochrane tools. Meta-analyses were conducted using Review Manager 5.4; forest plots were created to assess overall treatment effects.

**Meta-analysis:** A meta-analysis was performed.

**Results:** After 8200 duplicate articles had been removed and a further 3651 papers excluded on title and abstract, 39 studies involving 1779 patients met the inclusion criteria. Among these, 10 studies evaluated ESWT, either alone or in combination with SWC. The meta-analysis of 15 studies revealed that electromechanical therapies significantly improved healing outcomes in DFUs compared to controls (p < 0.001). Specifically for ESWT, included studies reported accelerated healing, reduced ulcer size, increased expression of growth factors (e.g., VEGF, TGF-β, IGF-1), improved angiogenesis and shortened inflammation. One trial reported a full healing rate of 74% with ESWT in previously nonresponsive chronic wounds. While heterogeneity among trials was high (I^2^ = 98%), the overall effect favored treatment. Additionally, safety outcomes indicated that ESWT and similar therapies were well tolerated, with only minor, transient side effects reported (e.g., erythema, discomfort). Serious adverse events were rare.

**Conclusions:** This review confirmed that electromechanical therapies, including ESWT, are safe and effective adjuncts to standard DFU care. ESWT, in particular, showed robust biological and clinical potential, improving vascularization, cellular proliferation and wound closure rates. However, methodological heterogeneity and lack of standardized treatment parameters limit generalizability. Future trials should prioritize standardized protocols, cost-effectiveness evaluation and exploration of ESWT’s dose–response relationship. Despite these limitations, ESWT is a viable, noninvasive therapeutic option for accelerating DFU healing and should be considered in refractory cases or as part of a multimodal treatment strategy.

**[61] Wigley, C.H.; Janssen, T.J.; Mosahebi, A. Shock wave therapy in plastic surgery: A review of the current indications. *Aesthet. Surg. J.* 2023, *43*(3), 370–386. https://doi.org/10.1093/asj/sjac262.**

**Motivation:** ESWT has demonstrated regenerative, angiogenic and anti-inflammatory effects across various clinical settings. In plastic surgery, ESWT has gained increasing interest as a non-invasive adjunct for the treatment of chronic wounds, including DFUs, yet its therapeutic role and clinical value remain incompletely defined.

**Hypothesis:** This review tested the hypothesis that ESWT improves tissue regeneration and wound healing outcomes, including in DFUs, through mechanotransduction, and that its efficacy and safety warrant detailed evaluation within plastic surgery contexts.

**Methods:** A systematic literature review was conducted in line with PRISMA guidelines, covering studies up to June 1, 2021, using PubMed, Embase and the Cochrane Library. Inclusion criteria encompassed clinical studies of any design investigating ESWT applications in plastic surgery, with search terms targeting “shockwave” and “wound” alongside related keywords. After screening 364 records, 46 clinical studies with a total of 1496 patients were included. The review applied narrative synthesis, categorizing ESWT applications into cellulite, body contouring, scar management, burns and chronic wounds (including DFUs).

**Meta-analysis:** No meta-analysis was performed.

**Results:** Nine included studies specifically addressed DFUs, and a further four addressed other chronic wounds. Across RCTs and case series, ESWT demonstrated significantly higher healing rates and shorter healing times compared to SWC or HBOT. Reported outcomes included (i) a 53.3% complete healing rate vs. 33.3% in controls over 20 weeks, (ii) significantly reduced healing time (64.5 ± 8.1 days vs. 81.2 ± 4.4 days) and increased healing rates (54% vs. 28.5%), (iii) greater wound area and perimeter reduction compared to sham-treated controls, and (iv) favorable perfusion and angiogenic marker changes on histology. Furthermore, a large case series (n = 208) showed complete re-epithelialization in 75% of patients with therapy-resistant ulcers. Despite the variability in ESWT protocols (e.g., 100–500 ESWs/cm^2^; 0.03–0.23 mJ/mm^2^), all studies reported improved healing metrics. No serious adverse events were reported; ESWT was generally well tolerated. Limitations included inconsistent methodology, lack of protocol standardization and occasional industry funding.

**Conclusions:** The reviewed evidence suggested that ESWT is a safe, non-invasive and effective adjunctive therapy for chronic wounds, including DFUs, when combined with standard care. Its mechanism of action involves angiogenesis, reduced inflammation and enhanced tissue regeneration through mechanotransduction. However, the lack of standardized dosing protocols and methodological consistency limits definitive clinical recommendations. Further high-quality RCTs are needed to optimize ESWT parameters and confirm its comparative effectiveness, especially in relation to HBOT and other advanced wound therapies. In plastic surgery, ESWT holds promise as a cost-effective, outpatient-friendly treatment for complex wounds and scars.

**[62]	Chen, P.; Vilorio, N.C.; Dhatariya, K.; Jeffcoate, W.; Lobmann, R.; McIntosh, C.; Piaggesi, A.; Steinberg, J.; Vas, P.; Viswanathan, V.; Wu, S.; Game, F. Effectiveness of interventions to enhance healing of chronic foot ulcers in diabetes: A systematic review. *Diabetes Metab. Res. Rev.* 2024, 40(3), e3786. https://doi.org/10.1002/dmrr.3786.**

**Motivation:** DFUs burden health care systems and individuals substantially, so interventions to enhance healing must be backed by high-quality evidence and cost-effectiveness. Previous IWGDF systematic reviews had made four-yearly search updates including non-randomized designs; the IWGDF/EWMA 21-point checklist introduced in 2016 set a new standard for re-evaluating older articles.

**Hypothesis:** No formal hypothesis was tested. The review used PICO questions covering ten intervention categories; for physical alteration of the wound bed, which included ESWT, the question was whether the intervention enhanced healing compared with standard care.

**Methods:** A PROSPERO-registered protocol (CRD42022309184) and the PRISMA guidelines were followed. MEDLINE (PubMed), Scopus and Web of Science were searched without restrictions in January and November 2022, covering publications up to and including November 2022; reference lists but not the gray literature were checked. Titles, abstracts and full texts were screened independently in duplicate. Included RCTs had more than 80% of participants with DFUs or DFU outcomes reported separately and at least five participants per arm, the comparator being SWC. Methodological quality was scored with the 21-point criteria of Jeffcoate and colleagues and the Cochrane Risk of Bias 2 tool, certainty of evidence with GRADE against ten outcomes critical to decision-making.

**Meta-Analysis:** No meta-analysis was performed.

**Results:** The search identified 22,250 articles; 532 full texts were assessed and 262 studies included in ten intervention categories. Twenty-nine studies investigated physical therapies, four ESWT; three reported complete wound healing: the only trial blinding both participants and outcome assessors (206 participants) found no difference at 12 weeks, a single-blind study (38 participants) none at 4, 8 or 20 weeks, and an unblinded study (30 participants) reported improved complete healing at 20 weeks after three ESWT applications. Protocols were considerably heterogeneous, all three studies at risk of bias, two at high risk. Two reported faster healing with ESWT, one unblinded, the other a per-protocol analysis. Of three studies reporting ulcer area reduction, the 38-participant study found a significant reduction at 8 and 20 weeks, the other two none at 7 and 20 weeks.

**Conclusions:** The evidence statement for ESWT recorded (i) differing protocols, devices and follow-up times, (ii) that the one study blinding both participants and outcome measures showed no difference in healing, and (iii) that, all three studies being at risk of bias, evidence suggesting improved healing over standard of care was of low certainty. Certainty was low or very low for most interventions across the review, moderate for two and low for four others. Further high-quality RCTs were needed, few studies being at low risk of bias.

**[63] Wang, Y.; Hua, Z.; Tang, L.; Song, Q.; Cui, Q.; Sun, S.; Yuan, Y.; Zhang, L. Therapeutic implications of extracorporeal shock waves in burn wound healing. *J. Tissue Viability* 2024, *33*(1), 96–103. https://doi.org/10.1016/j.jtv.2023.12.003.**

**Motivation:** Burn wounds, particularly those resulting in hypertrophic scarring and chronic inflammation, pose significant clinical challenges and long-term disability. Conventional treatments often fail to prevent complications, prompting exploration of adjunctive physical therapies such as ESWT to improve outcomes in burn wound management.

**Hypothesis:** While no formal hypothesis was tested, this review was based on the premise that ESWT enhances burn wound healing and mitigate sequelae, including infection, hypertrophic scarring, pain and pruritus, by modulating inflammation, promoting angiogenesis and accelerating tissue regeneration.

**Methods:** This narrative review summarized preclinical and clinical studies of ESWT in burn wound healing, including its history, physical principles, mechanisms of action, application parameters and therapeutic implications. No formal systematic search strategy was applied. The article reviewed both fESWT and rESWT, and classified therapeutic effects by mechanistic domains such as inflammation control, angiogenesis, scar remodeling and symptom relief.

**Meta-analysis:** No meta-analysis was performed.

**Results:** ESWT has demonstrated multiple beneficial effects in burn wound healing. Preclinical studies showed that ESWT suppresses early proinflammatory responses by reducing macrophage infiltration, inhibiting NF-κB activation and downregulating proinflammatory cytokines (e.g., TNF-α, iNOS, COX-2). It also exhibits direct and indirect antimicrobial activity, increasing bacterial membrane permeability and enhancing antibiotic delivery. In murine and rat burn models, ESWT enhanced re-epithelialization, neovascularization and scar remodeling, with improved collagen alignment and reduced fibroblast proliferation. Clinical trials reported that ESWT improves hypertrophic scar appearance, reduces pain and pruritus and accelerates healing of acute burn wounds and skin graft donor sites. Treatment protocols varied widely but generally involved low-to-medium energy settings (0.05–0.3 mJ/mm^2^), weekly sessions and 1000–2000 ESWs per session. Adverse events were rare and mild. However, therapeutic outcomes varied, with some studies failing to show statistically significant benefits due to heterogeneity in study design, patient population and treatment dosing.

**Conclusions:** ESWT offers a safe, non-invasive and promising adjunctive therapy for both acute burn wounds and post-burn complications such as hypertrophic scarring, pain and itch. It acts via anti-inflammatory, angiogenic, anti-fibrotic and antimicrobial mechanisms, supporting faster healing and improved tissue remodeling. While current evidence is encouraging, definitive clinical protocols are lacking and high-quality RCTs are needed to standardize dosing parameters, treatment duration and patient selection. ESWT represents a valuable tool in modern burn rehabilitation, with potential to significantly improve outcomes and quality of life for burn survivors.

**[64] Wu, F.; Qi, Z.; Pan, B.; Tao, R. Extracorporeal shock wave therapy (ESWT) favors healing of diabetic foot ulcers: A systematic review and meta-analysis. *Diabetes Res. Clin. Pract.* 2024, *217*, 111843. https://doi.org/10.1016/j.diabres.2024.111843.**

**Motivation:** DFUs are a severe complication of diabetes mellitus, leading to prolonged hospitalization,

amputation and increased mortality. Despite current standard therapies, healing outcomes remain suboptimal, motivating investigation into adjunctive treatments such as ESWT.

**Hypothesis:** This systematic review and meta-analysis evaluated the hypothesis that ESWT improves ulcer healing and microcirculatory function in DFU patients, with an acceptable safety profile when compared to SWC or HBOT.

**Methods:** A systematic review and meta-analysis was conducted following Cochrane Handbook and PRISMA guidelines. Databases searched included PubMed, EMBASE, Cochrane CENTRAL and Web of Science for RCTs published before August 8, 2023. Studies had to report at least one of the following: complete ulcer healing, ≥50% improvement, unchanged ulcers, transcutaneous oxygen pressure (TcPO_2_) changes or treatment-emergent adverse events (TEAEs). Meta-analyses were performed using STATA 14.0; heterogeneity, publication bias and sensitivity analyses were assessed.

**Meta-analysis:** A meta-analysis was performed.

**Results:** Ten RCTs involving 754 patients were included. ESWT significantly increased the rate of complete healed ulcers (RR = 1.6; 95% CI: 1.3–2.0; p < 0.001) and decreased the rate of unchanged ulcers (RR = 0.25; 95% CI: 0.1–0.4; p < 0.001) compared to controls (SWC or HBOT). Subgroup analyses confirmed ESWT's superiority over both SWC and HBOT in these outcomes. ESWT also significantly improved TcPO_2_ levels (MD = 1.7 mmHg; 95% CI: 1.2–2.2; p < 0.001), suggesting enhanced local perfusion. However, no significant difference was found in the proportion of patients achieving ≥50% ulcer reduction (RR = 1.2; p = 0.444). The rate of TEAEs was comparable between ESWT and controls (RR = 0.6; p = 0.327), confirming its favorable safety profile. Included studies varied in ulcer size, HbA1c, intervention protocols and follow-up times, but showed no publication bias. All results were robust in sensitivity analyses.

**Conclusions:** This systematic review and meta-analysis confirmed that ESWT is a safe and effective adjunctive therapy for the treatment of DFUs. It enhances complete healing and perfusion while maintaining a low adverse event profile. Although ESWT did not significantly increase moderate ulcer improvement rates, the findings support its inclusion in DFU treatment protocols. Further large-scale, high-quality RCTs are needed to optimize treatment parameters and better understand the underlying mechanisms, including the role of glycemic control and ESWT dose-response.

**[65]	Abdelhakim, M.; Ogawa, R. Emerging Therapies in Chronic Wound Healing: Advances in Stem Cell Therapy, Growth Factor Modulation, Mechanical Strategies and Adjuvant Interventions. *Dermatol. Ther.* 2025, 15(12), 3533–3545. https://doi.org/10.1007/s13555-025-01564-2.**

**Motivation:** Chronic wounds such as DFUs, VLUs and pressure injuries affect millions of people worldwide at high economic cost. The authors stated that standard therapies often fail to close them, because inflammation persists, angiogenesis is impaired and cells respond poorly, and that no single treatment suits every wound type.

**Hypothesis:** No hypothesis was formulated or tested. The review assumed that regenerative and mechanotherapy-based approaches (stem cells, growth factor modulation and mechanical treatments such as ESWT) correct the biological deficits of the wound, a shift toward personalized regenerative care.

**Methods:** This was a narrative review. The authors searched PubMed/MEDLINE and Embase for work on regenerative cell therapy, growth factor modulation, mechanical and physical strategies, adjuvant therapies and platelet-derived products, preferring randomized and controlled studies, good observational cohorts, meta-analyses and consensus guidelines. They reported no search dates, search terms, eligibility criteria, record numbers, PRISMA flow diagram, protocol registration or risk-of-bias assessment. The scope covered stem cell therapy, growth factor modulation (PDGF, VEGF and EGF), platelet-derived extracellular vesicles, mechanobiological treatments (NPWT, ESWT, ultrasound and compression therapy), photobiomodulation, HBOT, topical oxygen therapy and three-dimensional bioprinting.

**Meta-Analysis:** No meta-analysis was performed.

**Results:** ESWT was presented as one of four mechanotherapy-oriented treatments, with NPWT, ultrasound and compression therapy. Its high-energy acoustic waves were described to exert mechanical forces that make cells produce bioactive molecules and proliferate, fibroblasts in particular. According to the authors, ESWs raise VEGF and other growth factors and so promote angiogenesis and vascularization; they remodel collagen fibers, which improves structural integrity and helps prevent excessive scarring; and they reduce inflammation and pain by activating anti-inflammatory cytokines and disrupting biofilm. A summary table added stem cell recruitment to these mechanisms, citing an experimental study of ESWT on incisional wounds in diabetic rats. No clinical trial of ESWT in wound management was cited, and no treatment parameters (EFD, ESW or session number), wound types or healing rates were reported.

**Conclusions:** The authors concluded that stem cell therapy, growth factor modulation and mechanical treatments each offer distinct benefits and, used together, could transform outcomes in refractory wounds. They identified three obstacles. Wounds and patients differ widely in age, comorbidity and wound etiology. Regeneration reaches its limit where tissue destruction is pronounced, making reconstructive strategies essential. And protocols for stem cell isolation, growth factor delivery and mechanical therapy are not standardized, which the authors stated hinders comparison across studies. Regulatory hurdles and high costs were described as further brakes on translation.

**[66]	Fu, C.; Li, N.; Li, M.; Li, H.; Cheng, L.; Wang, L. A systematic review and Bayesian network meta-analysis of the effectiveness of non-pharmacological interventions in treating chronic wound pain in patients. *Chin. J. Burns Wounds* 2025, 41(5), 491–500. https://doi.org/10.3760/cma.j.cn501225-20241213-00486.**

**Motivation:** Pain is a core problem of chronic wounds, contributing to anxiety and depression and lowering quality of life. Opioids and local anesthetics have adverse effects and carry a risk of dependence, while research on non-pharmacological adjuncts had addressed single modalities, leaving their relative effectiveness undetermined.

**Hypothesis:** No singular hypothesis was tested. The review assumed that non-pharmacological modalities added to SWC differ in relieving chronic wound pain, the most effective identifiable from direct and indirect evidence.

**Methods:** The review was registered in PROSPERO (CRD42024536186) and followed the PRISMA extension for network meta-analyses. Eight databases (PubMed, Embase, the Cochrane Library, Web of Science and four Chinese ones) were searched from inception to 30 June 2024. Eligible were RCTs in adults with chronic wounds comparing a non-pharmacological intervention with SWC, placebo or another and reporting post-treatment pain (visual analogue or numerical rating scale, EQ-5D, Lattinen pain scale). Two reviewers screened, extracted data and applied Cochrane RoB 2. Of 8975 records, 30 RCTs with 1929 patients (1016 treated, 913 control) in ten intervention categories, one of them ESWT, were included.

**Meta-Analysis:** The pairwise analysis used ADDIS 1.16.8 (SMD, random effects), the Bayesian network meta-analysis Stata 17.0 with ADDIS 1.16.8, node splitting preceding the consistency model. Three RCTs formed the ESWT node, all of ESWT plus SWC versus SWC (72 treated, 73 control). Pairwise, ESWT gave an SMD of -1.7 (95% CI -4.1 to 0.6; I^2^ 96.8%); in the network, -1.2 (95% CI -2.5 to 0.4). The frequency rhythmic electrical modulation system (FREMS) was superior to ESWT (1.94, 95% CI 0.2 to 4.0). Ranking used probability plots rather than SUCRA, rank 14 being the most effective; probabilities were given for FREMS (rank 14, 8%), cold atmospheric plasma (CAP; rank 12, 2%) and two further modalities, but none for ESWT. The comparison-adjusted funnel plot showed no significant asymmetry.

**Results:** In the pairwise analysis, pain fell significantly with low-level laser therapy (LLLT; SMD -0.5, 95% CI -0.8 to -0.2), FREMS (-4.1, -6.0 to -2.2), virtual reality technology (VRT; -1.0), non-contact low-frequency ultrasound (NCLFU; -0.6), CAP (-1.9, -3.2 to -0.6) and intermittent pneumatic compression (IPC; -0.6), but not with ESWT, topical oxygen therapy, NPWT or exercise training. In the network, FREMS (-3.1, -4.4 to -2.0), CAP (-1.8, -3.2 to -0.3), NCLFU (-1.2, -2.4 to -0.1) and LLLT (-1.1, -2.2 to -01) were superior to SWC, VRT and IPC were not, and FREMS surpassed ESWT and all others. Risk of bias across the 30 trials was uncertain.

**Conclusions:** The review concluded that LLLT, FREMS, NCLFU and CAP relieve chronic wound pain effectively, FREMS most, then CAP, NCLFU and LLLT; neither analysis showed that ESWT reduces pain. The limitations named were the exclusion of dressings, possible residual bias, the single outcome, the absence of subgroup analyses by wound type, and differences in device cost and regional resources.

**[67]	Gong, J.; Sun, X.; Fu, H.; Zhang, Y.; Gao, T. Comparative efficacy and safety of non-pharmacological nursing interventions for diabetic foot ulcers: a systematic review and network meta-analysis. *BMC Nurs.* 2025, 24(1), 1445. https://doi.org/10.1186/s12912-025-04065-x.**

**Motivation:** DFUs were presented as a severe complication of diabetes, with 589 million adults projected to have diabetes in 2025. Antibiotic resistance and adverse drug reactions limit drug treatment, and earlier syntheses could not rank several non-pharmacological nursing interventions at once.

**Hypothesis:** No singular hypothesis was tested. The review assumed that non-pharmacological nursing interventions, ESWT among them, differ in efficacy and safety across DFU outcomes, and that a network synthesis would order them.

**Methods:** The review followed PRISMA and its network meta-analysis extension. Five databases (including PubMed, Embase and the Cochrane Central Register of Controlled Trials) were searched from inception to 31 March 2025. Eligible were RCTs in DFU patients comparing a non-pharmacological adjunct with SWC or another and reporting healing, wound area, recurrence, amputation or adverse events; four non-English studies were excluded. Two reviewers applied Cochrane RoB 2, certainty being appraised with CINeMA. Of 3033 records, 67 RCTs with 5957 patients in ten intervention nodes were included, 9 on ESWT.

**Meta-Analysis:** The methods described a frequentist random-effects network meta-analysis in Stata 17.0 (network, mvmeta), the abstract calling the modeling Bayesian. Effects were odds ratios (ORs) or standardized mean differences (SMDs), ranked by the surface under the cumulative ranking curve (SUCRA). Versus conventional care, ESWT increased the healing rate (OR 3.6, 95% CI 1.8 to 7.1) and shortened healing time (SMD -2.0, 95% CI -3.3 to -0.7), ranking first (SUCRA 94.4%). It did not differ for change in wound area (-0.6, -1.8 to 0.6) or amputation (OR 0.2, 95% CI 0.01 to 5.1), despite the highest amputation SUCRA (68.4%). In the results tables the only significant amputation contrast was HBOT (OR 0.4, 95% CI 0.2 to 0.7), the estimate the abstract attributed to ESWT.

**Results:** Light therapy (LT) gave the highest healing rate (OR 7.6, 95% CI 2.9 to 19.9), followed by ultrasound therapy, exercise therapy (EXER), HBOT, NPWT and continuous diffusion of oxygen (CDO), all significantly improving healing. Ultrasound therapy gave the largest reduction in wound area (SMD -3.2, 95% CI -4.8 to -1.5; SUCRA 98.4%), EXER reduced recurrence (OR 0.1, 95% CI 0.02 to 0.3) and CDO caused the fewest adverse events. Six of 67 trials were at high risk of bias, and Egger tests indicated publication bias for healing and amputation rate. Global and local inconsistency tests gave p above 0.05.

**Conclusions:** The review concluded that the network yields an outcome-specific hierarchy: LT for complete healing, ultrasound therapy for wound area reduction, ESWT for shorter healing time and lower major amputation risk, EXER for recurrence prevention and CDO for safety. ESWT was presented as suitable where rapid closure and limb preservation are priorities; CINeMA certainty for its contrasts being low to moderate, the findings were called hypothesis-strengthening rather than definitive.

**[68]	Howard, T.J.; Amir, L.; Regulski, M.J.; Mullins, J.D. Use of extracorporeal shock wave therapy for wounds and musculoskeletal treatment: a systematic review. *J. Wound Care* 2025, 34(Sup. 12a), S5–S16. https://doi.org/10.12968/jowc.2024.0113.**

**Motivation:** ESWT converts acoustic into mechanical energy and promotes tissue regeneration through mechanotransduction, and two devices held US Food and Drug Administration clearance for hard-to-heal wounds. Previous reviews reached differing conclusions, and heterogeneity in EFD and other parameters limited earlier meta-analyses.

**Hypothesis:** No singular hypothesis was tested. The purpose was to evaluate the results and error estimates of recent comparative studies of fESWT and uESWT and its efficacy for wounds of the skin and musculoskeletal injuries.

**Methods:** Google Scholar and MEDLINE via PubMed were searched on 4 June 2021 (term extracorporeal shock wave therapy, clinical trial filters, 2010 to 2021); the article appeared in December 2025. Eligible were RCTs from 2010 onward in wound or musculoskeletal injury patients with a placebo or SWC arm; rESWT and non-English reports were excluded. From 304 records, 14 RCTs were reviewed: six on acute wounds (five on burns), two on hard-to-heal wounds (a decubitus ulcer trial with 9 patients, a DFU trial with 336) and six on musculoskeletal injuries. Trials were stratified by EFD, assessed with RoB 2 and graded with GRADE.

**Meta-Analysis:** Outcomes reported by more than one trial per group were pooled in R 4.3.2; three of the four concerned acute wounds. For pain intensity (three trials; 67 treated, 66 placebo) the random-effects mean difference was -2.3 (95% CI -4.4 to -0.3; I^2^ 97%). For pressure pain threshold (two trials; 85 patients) the random-effects standardized mean difference was 1.1 (95% CI 0.5 to 1.8; I^2^ 50%). For time to re-epithelialization (two trials; 72 patients) the fixed-effects mean difference was -2.9 days (95% CI -3. 8 to -2.0; I^2^ 0%). The two hard-to-heal trials were not pooled. For musculoskeletal pain intensity the mean difference was -1.7 (95% CI -3.2 to -0.2; I^2^ 77%). All four estimates favored ESWT; GRADE certainty was moderate for three acute wound outcomes and very low for musculoskeletal pain. Publication bias could not be assessed from so few studies, and no ranking statistics were produced.

**Results:** ESWT matched or exceeded its comparator in all 14 trials, and 12, including all 8 wound trials, reported significantly faster healing or greater pain reduction. Burn trials showed less pain and pruritus and better scar measures; two recorded re-epithelialization after 13.9 versus 16.7 and 9.6 versus 12.5 days, favoring ESWT. In the decubitus ulcer trial mean ulcer size changed by -0.7 cm^2^ with ESWT and 0.3 cm^2^ with placebo, and in the DFU trial 37.8% of ulcers closed by 24 weeks versus 26.2% of controls. ESWT was significantly better in four of six musculoskeletal trials.

**Conclusions:** The review concluded that ESWT supports wound healing and is promising for musculoskeletal injuries, the outcome favoring it in 12 of 14 trials. Design variability precluded a confident estimate of effect size, and the association between EFD and efficacy could not be tested, six trials spanning more than one energy band. Standardized, tissue-specific protocols and placebo comparators were recommended.

**[69]	Hu, X.; Meng, H.; Liang, J.; An, H.; Zhou, J.; Gao, Y.; You, C.; Zhang, Z.; Gong, X.; Liu, Y. Comparison of the efficacy of 12 interventions in the treatment of diabetic foot ulcers: a network meta-analysis. *PeerJ* 2025, 13, e19809. https://doi.org/10.7717/peerj.19809.**

**Motivation:** In 2021, diabetes affected 10.5% of people aged 20 to 79 years, and 19% to 34% of patients with diabetes develop a DFU. SWC alone is often not enough, most earlier syntheses were pairwise, and the two existing network meta-analyses had omitted common treatments.

**Hypothesis:** No singular hypothesis was stated. The review assumed that the 12 interventions differ across the DFU healing endpoints, and that indirect comparison through a common control would rank their effects.

**Methods:** The review followed PRISMA and was prospectively registered in PROSPERO (CRD42023461811). Two authors searched PubMed, Web of Science, the Cochrane Library and Embase from inception to September 2023. Eligible were RCTs in DFU patients comparing one of 12 interventions with SWC or placebo and reporting healing rate, healing time, percentage area reduction (PAR) or amputation rate. The 12 included ESWT, stem cells (SC), amniotic membrane therapy (AMT), low-level laser therapy (LLLT), platelet-rich plasma (PRP), NPWT, low-frequency ultrasound (LFU), HBOT and topical oxygen therapy (TOT). Of 1981 records, 99 RCTs with 7356 participants were included, the ESWT node 8 RCTs, six versus SWC and two versus HBOT.

**Meta-Analysis:** A frequentist random-effects network meta-analysis across the 12 nodes used Stata 16.0, with ORs and MDs ranked by the surface under the cumulative ranking curve (SUCRA). Global inconsistency gave p = 0.227, with no small-study effects in the funnel plot. In the healing rate network (95 RCTs, 6807 patients), ESWT was one of ten interventions significantly better than SWC, its estimate reported only in a figure. In the PAR network (15 RCTs, 880 patients), ESWT improved PAR over SWC (MD 27.5, 95% CI 11.0 to 44.0) and TOT, ranking second (SUCRA 84.0%) behind LLLT (93.9%). ESWT did not significantly shorten healing time and was absent from the amputation network. It showed no benefit within 12 weeks but improved healing thereafter (OR 2.7, 95% CI 1.1 to 6.7).

**Results:** For healing rate, only LFU (OR 2.2, 95% CI 1.0 to 4.9) and electric stimulation (OR 1.9, 95% CI 0.9 to 4.1) were no better than SWC. SC ranked highest (SUCRA 89.7%; OR 5.7, 95% CI 2.6 to 12.3), followed by AMT (OR 5.1, 95% CI 3.1 to 8.4). AMT (MD -26.9 days, 95% CI -44.3 to -9.6), PRP and NPWT significantly shortened healing time. For PAR, LLLT (MD 34.3, 95% CI 17.4 to 51.2) and ESWT were superior to SWC. SC (OR 0.1, 95% CI 0.03 to 0.6) and HBOT (OR 0.4, 95% CI 0.2 to 0.8) significantly reduced the amputation rate.

**Conclusions:** The review concluded that SC and AMT were the most effective interventions for the DFU healing rate, that AMT also shortened healing time, that LLLT was most effective for reducing ulcer area and that SC reduced the amputation rate. HBOT and ESWT were described as time-dependent, their effects accumulating beyond 12 weeks, which places ESWT where follow-up is long. The principal limitation was the lack of ulcer type classification.

**[70]	Ruiz-Muñoz, M.; Rueda-Zapata, L.; Martinez-Barrios, F.-J.; Nováková, T.; Lopezosa-Reca, E.; Gonzalez-Sanchez, M.; Fernandez-Torres, R.; Galan-Mercant, A. Efficacy of extracorporeal shockwave therapy in the management of chronic diabetic foot ulcer: a systematic review and meta-analysis. *Med. Sci.* 2025, 13(4), 219. https://doi.org/10.3390/medsci13040219.**

**Motivation:** DFUs are a frequent complication of diabetes, which affects approximately 463 million adults. The figures reported were a lifetime risk of ulceration of 30-34%, amputation in up to 40% of patients and death within five years in 70% of those amputated. The most recent meta-analysis covered trials only up to 2014, so an update was required.

**Hypothesis:** No hypothesis was formulated. The stated objective was to evaluate ESWT as an addition to SWC in chronic DFUs, and to determine whether more ulcers close completely than with SWC alone.

**Methods:** The review followed PRISMA 2020 and was registered in PROSPERO in advance (CRD420250527214). Eligible were RCTs in adults with chronic DFUs comparing ESWT with SWC; the primary outcome was the healing rate. WoS, EMBASE, MEDLINE Complete, CINAHL Complete, Academic Search Ultimate, AMED, Scopus and PubMed were searched in July 2024 with Boolean terms for DFUs and ESWT. Two reviewers screened independently; risk of bias was assessed with the robvis tool and quality with the Spanish Critical Appraisal Skills Programme (CASPe), cut-off 8 of 11 points.

**Meta-Analysis:** A random-effects model on the log odds ratio was back-transformed to an OR. The results section gave a pooled OR for complete healing with ESWT versus SWC of 2.9 (95% CI 1.9-4.2; z = 5.3, p < 0.001), the abstract a different figure (OR 2.8, 95% CI 2.0-3.8). Heterogeneity was low: Q(5) = 6.4, p = 0.274; I^2^ = 21.3%. Neither the Kendall rank correlation test (tau = -0.333, p = 0.200) nor the Egger test (intercept 0.488, p = 0.385) indicated funnel-plot asymmetry; trim-and-fill imputed two studies, adjusted OR 2.5 (95% CI 1.7-3.5).

**Results:** Of 206 records, 21 full texts were assessed and 8 articles included. Six data sets entered the quantitative synthesis: Moretti et al. (2009; [95]), Omar et al. (2014; [29]), Jeppesen et al. (2016; [99]), the two trials of Snyder et al. (2018; [28]) and Vangaveti et al. (2023; [119]), together 672 patients, all scoring 10 or 11 of 11 points on CASPe. The log odds ratio was positive in every trial and significant in three. Protocols varied in device, EFD (0.03-0.23 mJ/mm^2^), pulse number and treatment period (3-20 weeks).

**Conclusions:** The authors concluded that adjunctive ESWT produced significantly higher rates of complete healing in chronic DFUs than SWC alone, patients being almost three times as likely to reach closure, and recommended it as a valuable addition to standard care. The finding was regarded as robust, heterogeneity being low and the sensitivity and trim-and-fill estimates stable. Trials varied in patient characteristics and protocols, dose, frequency and duration being unstandardized, and the International Working Group on the Diabetic Foot does not recommend ESWT for routine use.

**[71]	Talukdar, S.; Medhi, J.; Barbhuiya, P.A.; Chakrabarti, S. Complementary approaches and advanced technologies in ameliorating rate of wound healing. *Int. J. Pharm.* 2025, 682, 125910. https://doi.org/10.1016/j.ijpharm.2025.125910.**

**Motivation:** Wound healing is a multi-phase process of hemostasis, inflammation, proliferation and matrix remodeling. Diabetes and extensive burns disrupt it, producing chronic wounds that do not close and are prone to infection and abnormal scarring, while traditional treatments often fall short and high cost limits advanced wound care technologies. The review summarized complementary approaches and advanced technologies for accelerating healing in acute and chronic wounds.

**Hypothesis:** No hypothesis was formally tested. The review assumed that complementary and advanced approaches acting at cellular and molecular level, among them ESWT, can accelerate repair in wounds that respond poorly to conventional treatment.

**Methods:** This was a narrative review. The authors reported no protocol, registration, database list, search terms, date limits, eligibility criteria, selection procedure or risk-of-bias assessment, and did not follow PRISMA. Material was organized by theme: conventional versus complementary approaches; cell and gene therapy and the delivery of growth factors, bioactive substances and biomaterials; advanced technologies (injectable hydrogels, artificial-intelligence-enabled sensor dressings, bioengineered skin grafts, three-dimensional bioprinting, siRNA and microRNA therapeutics); preclinical wound models; and advanced therapies in clinical trials. Preclinical and clinical studies were tabulated with intervention, enrollment, duration, dose, outcome and registration number.

**Meta-Analysis:** No meta-analysis was performed.

**Results:** Only one clinical ESWT wound study was cited (Landscheidt et al., 2025; [105]), in the clinical trials section and again in the conclusion: a prospective single-center trial in acute and chronic wounds under the Ärztekammer Berlin (Germany), with 25 control and 10 intervention participants, topical application and three months' duration; re-epithelialization was faster and the risk of wound infection lower, with no serious side effects. No device type, EFD, number of EFDs or session schedule was given. ESWT was grouped with noncontact normothermic wound therapy and ON101 cream as non-invasive and topical therapies with encouraging healing and infection outcomes. Peripheral blood mononuclear and umbilical cord mesenchymal stem cells improved perfusion and ulcer healing, whereas NPWT was no better than usual care in surgical wounds.

**Conclusions:** The authors concluded that treating chronic and large wounds remains a major challenge and that no single therapy is fully effective. ESWT was listed among the innovative strategies for acute and chronic wounds, alongside stem cell and gene therapy, wireless microcurrent stimulation, NPWT, vacuum-assisted closure, cold atmospheric plasma and near-infrared laser treatment. The abstract stated that non-invasive methods such as ESWT and wireless microcurrent stimulation show potential but require confirmation. Remaining obstacles were biological safety, wound-type variability, high costs and limited clinical translation; standardized protocols and larger trials were called for.

**[72]	Guan, X.; Zhu, G.; Jiang, H.; Wang, B.; Cheng, J. Comparative efficacy of advanced therapeutic modalities for diabetic foot ulcers: a systematic review and meta-analysis including NPWT, stem cells, ESWT, and bioengineered treatments. *Front. Bioeng. Biotechnol.* 2026, 14, 1792670. https://doi.org/10.3389/fbioe.2026.1792670.**

**Motivation:** DFUs occur at least once in 15-25% of patients with diabetes and drive hospitalization and treatment costs. The review compared NPWT, therapies based on mesenchymal stem cells, bioengineered skin substitutes, ESWT and PRP.

**Hypothesis:** No formal hypothesis was stated. The review compared these modalities with SWC for complete healing, time to closure, reduction in wound area and adverse events.

**Methods:** PRISMA was followed without protocol registration. PubMed, Embase, the Cochrane Library, Web of Science and Scopus were searched from inception to the end of December 2025. Eligible were RCTs and comparative clinical studies in adults with DFUs; non-English publications, case reports, reviews, data-free studies and gray literature were excluded. Quality, risk of bias and certainty were assessed with the Jadad scale, the Cochrane Risk of Bias 2.0 tool and GRADE.

**Meta-Analysis:** Random-effects inverse-variance pooling gave standardized mean differences (SMDs) with 95% CIs. ESWT formed Group 4 with the other physical modalities. For complete healing, three studies (160 treated, 154 control) gave an SMD of -1.0 (95% CI -1.2 to -0.7) favoring the intervention, without significant heterogeneity; the discussion of the same article reported complete healing from four studies as an SMD of 0.4 (95% CI 0.2-0.6). For time to closure, four studies (210 versus 202) gave an SMD of -1.1 (95% CI -1.3 to -0.8, p < 0.05), likewise homogeneous. The Egger test showed no asymmetry (p = 0.504 and p = 0.472), and GRADE certainty for ESWT was moderate to high.

**Results:** Of 1276 records, 964 were screened after 312 duplicates were removed, 117 full texts assessed and 49 studies included, covering more than 6500 adults with DFUs (sample sizes 30-507, durations 2-24 weeks). Most scored 3 of 5 on the Jadad scale; under GRADE about 35 were high quality and 5 low. The ESWT trials were Snyder et al. (2018; [28]), two multicenter double-blinded phase III RCTs in 336 patients, Omar et al. (2014; [29]) in 60 and Vangaveti et al. (2023; [119]) in 90.

**Conclusions:** The authors concluded that advanced modalities improve DFU outcomes: NPWT produced consistent gains in complete healing, time to healing and wound area reduction, and mesenchymal stem cell-based therapies performed robustly for time to closure. Bioengineered skin substitutes, growth factors, ESWT and PRP also improved healing, although effect magnitude varied. Limitations were clinical diversity in designs and outcome measures, publication bias in some NPWT subgroups, few trials of newer modalities and excluded gray literature.

**[73]	Rodoplu, O. Extracorporeal spark-wave therapy for diabetic wounds and critical limb ischemia: linking angiogenesis, tissue regeneration, and limb salvage outcomes. *Int. J. Drug Deliv.* *Technol.* 2026, 16(66s), 215–226. https://doi.org/10.25258/ijddt.16.66s.28.**

**Motivation:** DFUs and chronic limb-threatening ischemia combine neuropathy, arterial insufficiency and impaired angiogenesis, and about half of those with a foot ulcer also have peripheral arterial disease. The author noted that the diabetic wound, angiogenesis and ischemic limb literatures are reported separately, and that the IWGDF 2023 update did not list ESWT among supported adjuncts.

**Hypothesis:** No single hypothesis was formally tested. The review asked whether ESWT improves angiogenesis-mediated regeneration and limb preservation, addressing wound contraction, time to closure and granulation quality, signals of vascular endothelial growth factor (VEGF), and perfusion outcomes such as walking distance and the ankle-brachial index (ABI).

**Methods:** The review followed PRISMA 2020. PubMed/MEDLINE, Scopus, Web of Science, ScienceDirect, the Cochrane Library and Google Scholar were searched from January 2021 to December 2025. The PICO question covered adults with diabetic or chronic ischemic wounds treated with fESWT, rESWT or low-intensity ESWT against sham, SWC or other adjuncts; peer-reviewed English publications were eligible. Of 1184 records, 872 were screened after duplicate removal, 83 full texts assessed and 25 studies included. Extraction covered ESWT modality, EFD or pressure, impulses per cm^2^ and number of sessions.

**Meta-Analysis:** No meta-analysis was performed.

**Results:** Of the 25 publications, 8 used fESWT (median 6 sessions), 6 rESWT (median 1), 5 low-intensity protocols aimed at the vessels or at claudication (median 6) and 6 mixed or unspecified protocols (median 4). A pilot three-arm double-blind RCT related impulse density to healing time: median healing took 54.0 days at 500 ESWs/cm^2^, 78.5 at 100 EFDs/cm^2^ and 83.0 under sham. In a geriatric chronic-wound study, one rESWT session lowered mean wound area from 9.4 cm^2^ to 6.2 cm^2^ and improved the mean Wound Bed Score by 31.3%. Cited network meta-analyses gave odds ratios of 2.7 (95% CI 1.1-6.7) for healing beyond 12 weeks and 0.4 (95% CI 0.2-0.7) for amputation. All 6 mechanistic papers reported VEGF up-regulation and all 4 that measured it increased capillary density. A claudication meta-analysis of 332 patients gave mean differences of +26.5 (pain-free) and +37.2 (maximum walking distance), with the ABI unchanged. No recurring treatment-related harms were identified.

**Conclusions:** The author concluded that ESWT is a biologically plausible and clinically beneficial adjunct in selected diabetic wounds and ischemic limbs, complementary to revascularization, debridement, infection control and pressure relief, particularly where healing is limited by regenerative failure rather than uncontrolled infection or inflow disease. The evidence was characterized as a consistent pattern rather than a single randomized effect estimate. Limitations were heterogeneity, few direct DFU trials and possible publication bias; multicenter randomized trials with ischemia stratification, standardized energy delivery, sham control and longer follow-up were called for.

**[74]	Su, Y.; Fu, X.; Huang, Y. Consensus statement on the clinical application of extracorporeal shock wave therapy for diabetic foot ulcers (2025 Edition). *Int. J. Surg.* 2026, 112, 71–83. https://doi.org/10.1097/JS9.0000000000003489.**

**Motivation:** DFUs are among the most severe late complications of diabetes, with high disability and mortality. ESWT was approved by the US Food and Drug Administration for DFU wounds in 2017, but Chinese adoption remained limited, protocols being inconsistent and technical standards absent.

**Hypothesis:** No hypothesis was tested. The document assumed ESWT to be a non-invasive and effective adjunct for DFUs whose benefit depends on standardized indications, device selection and parameters derived from the literature and national clinical experience.

**Methods:** This expert consensus of the Wound Repair Committee of the Chinese Medical Doctor Association was pre-registered (PREPARE-2025CN072). Sixty-nine experts formed subgroups for drafting, literature, methodology and appraisal. Five databases were searched from inception to 1 December 2024, studies being appraised by design with the Cochrane risk of bias tool, the Newcastle-Ottawa Scale or the Joanna Briggs Institute criteria. Evidence was graded A for multicenter trials or meta-analyses, B for single-center or large uncontrolled studies and C for expert opinion or small retrospective data. PICO questions were settled in two Delphi rounds and a workshop; recommendation grades ran from I through IIa and IIb to III, with agreement above 90% strong, 70-90% moderate and 50-70% weak.

**Meta-Analysis:** No meta-analysis was performed.

**Results:** Thirty recommendations were organized under nine clinical questions, from indications and contraindications to treatment parameters and post-procedure care. Routine ESWT was recommended for first-onset Wagner grade 1 or 2 DFUs (Level of Evidence A, Recommendation I). Three further indications were graded Level of Evidence C, Recommendation IIb: Wagner grade 3 ulcers after debridement and drainage, ulcers downstaged after amputation for grade 4 or 5 disease, and ulcers with sinus tracts. The contraindications, among them acute thrombosis, malignancy, coagulation disorders and pregnancy, carried the same grade. Radial ESWT was considered particularly suitable, most of its energy reaching superficial tissue (Level of Evidence B, Recommendation IIa), while fESWT was reserved for deep injury such as osteomyelitis (Level of Evidence C, Recommendation IIb). Recommended settings were 1.0-2.5 bar for rESWs and an EFD of 0.03-0.18 mJ/mm^2^ for fESWs, both at 8-13 Hz, higher frequencies for superficial wounds (Level of Evidence A, Recommendation IIa). The treatment field was the wound plus a 2-3 cm margin; each session used 500 EFDs plus 100 per cm^2^ of treated area, at most 100 per point, 2 to 3 times weekly for 4 weeks (Level of Evidence B, Recommendation IIa).

**Conclusions:** The panel concluded that the evidence is too limited to define optimal parameters, treatment duration and the most suitable wound types, and that mechanisms of ESWT are incompletely understood. It called for multicenter, large-scale studies and prioritized standardizing ESWT training and certification. The document was presented as a standardized reference for ESWT in DFUs in China.

**[75]	Zheng, J.; Xie, D.; Wu, M.; Xu, H.; Lin, S.; Dong, L. Effects of non-pharmacological interventions on ulcer healing in patients with diabetic foot: a network meta-analysis of randomized controlled trials. *Front. Endocrinol.* 2026, 17, 1811595. https://doi.org/10.3389/fendo.2026.1811595.**

**Motivation:** DFUs affect 19% to 34% of people with diabetes, and 24% to 33% reportedly heal completely after 12 weeks of SWC. Earlier network meta-analyses mixed pharmacological with non-pharmacological treatments, used no uniform time point and often lacked standardized grading.

**Hypothesis:** No formal hypothesis was stated. Non-pharmacological interventions were assumed to differ in their effect on DFU healing, and to be rankable from RCTs with complete epithelialization at 12 weeks as the endpoint.

**Methods:** The review followed PRISMA and the Cochrane Handbook, was registered in PROSPERO (CRD420251122143) and searched eight Chinese and international databases to August 2025. Eligible were RCTs in DFUs graded Wagner 1-4 or University of Texas 1-3 comparing a non-pharmacological measure with routine care or placebo and reporting the 12-week healing rate or healing time as mean and SD. Screening, extraction and RoB 2 were done in duplicate, certainty with GRADE. Of 4376 records, 24 RCTs with 2352 patients and 15 intervention categories, published 2000-2024, were included; 8 were at low risk of bias, 14 raised some concerns and 2 were at high risk.

**Meta-Analysis:** A frequentist random-effects network meta-analysis used Stata 17.0. The pairwise synthesis gave an odds ratio (OR) of 3.3 (95% CI 2.5-4.3) for the healing rate (I^2^ 40.6%, p = 0.026) and a mean difference (MD) of -17.8 days (95% CI -23.2 to -12.4) for healing time (I^2^ 90.7%, p < 0.001), not reduced below 80% by subgroup analyses. By the surface under the cumulative ranking curve (SUCRA), fESWT plus SWC came twelfth for the healing rate, a position resting on a single RCT and described as dependent on indirect evidence with insufficient precision; it was absent from the healing-time network. The Egger and Begg tests indicated significant publication bias among the 22 healing-rate studies (p < 0.05) but not among the 8 healing-time studies.

**Results:** For the 12-week healing rate, NPWT plus SWC ranked highest (OR 28.3, 95% CI 5.0-159.2; SUCRA 96.1%), then autologous blood-derived products with dressing therapy (86.5%) and exercise therapy (80.4%). Six further interventions followed between 63.3% and 24.7%, then fESWT (17.6%), dressing therapy (6.3%) and SWC alone (5.4%). Gas therapy with dressing therapy plus SWC ranked highest for healing time (MD -64.6 days; SUCRA 100%). GRADE certainty was mostly low to very low.

**Conclusions:** The authors concluded that NPWT plus SWC may be the most beneficial option for the 12-week healing rate and that gas therapy with dressing therapy plus SWC may shorten healing time. They stressed that these results rest on small-sample studies with wide confidence intervals and limited certainty, that the rankings are exploratory and that a lower rank does not make an intervention ineffective. Limitations were the sparse evidence network and the detected publication bias; further large-sample RCTs were called for.
