## Supplementary File 2 for "Extracorporeal Shock Wave Therapy for Wound Management: Clinical Evidence, Energy Delivery Parameters and Mechanistic Insights — A Systematic Review"

**Extracorporeal Shock Wave Therapy in Wound Management: A Comprehensive Systematic Review of Clinical Evidence, Modalities and Mechanisms**

by Brent Musolf, Carmen Nussbaum-Krammer, Michael O’Neal, Nicola Maffulli and Christoph Schmitz

**Supplementary File 2**

**Standardized summaries and 40 key variables (c.f. Table 2 in the main text) from clinical studies on electrohydraulic ESWT for wound management**

**(numbers in brackets refer to the reference numbers in the main text)**

**Note on the standardized summaries:** Each summary condenses one clinical study as it was reported, together with the interpretation that its own authors placed on their findings. The summaries therefore reproduce the position of the respective authors and the state of the field at the time of that publication, and not the assessment of the present systematic review. They follow a uniform structure (Hypothesis, Methods, Results and Conclusions), because the original abstracts differ widely in structure, in length and in the information they report, which makes direct comparison between publications difficult; the standardized form is intended to remove this obstacle.

Abbreviations (in alphabetical order): CABG, coronary artery bypass grafting; DFUs, diabetic foot ulcers; EFD, energy flux density; eNOS, endothelial nitric oxide synthase; ESWT, extracorporeal shock wave therapy; ESWs, extracorporeal shock waves; fESWT, focused ESWT; HBOT, hyperbaric oxygen therapy; LDI, laser Doppler imaging; PCNA; proliferating cell nuclear antigen; PUs, pressure ulcers; RCT, randomized controlled trial; SWC, standard wound care; TcPO₂, transcutaneous partial oxygen pressure; uESWT, unfocused ESWT; V, variable; VEGF, vascular endothelial growth factor; VLUs, venous leg ulcers.

**[76]	Meirer, R.; Kamelger, F.S.; Piza-Katzer, H. Shock wave therapy: An innovative treatment method for partial thickness burns. *Burns* 2005, *31*(7), 921–922. https://doi.org/10.1016/j.burns.2005.02.013.**

**Hypothesis:** This study tested the hypothesis that fESWT accelerates wound healing and improves clinical outcomes in deep partial thickness burns, potentially avoiding the need for surgical interventions such as skin grafting.

**Methods:** This was a single-patient case report evaluating fESWT in a deep partial thickness burn. The subject was a 31-year-old male who sustained a burn on his right forearm caused by hot oil during a domestic accident. He refused the standard treatment of surgical excision and skin grafting for cosmetic reasons and, with informed consent, received fESWT as an alternative. Two fESWT sessions were administered on days 3 and 7 after the burn injury, using an Evotron device (High Medical Technologies (HMT), Lengwil, Switzerland), with an EFD of 0.11 mJ/mm^2^ and 1500 ESWs per treatment session. The frequency of the ESWs (in Hertz) and the number of ESWs per square centimeter were not reported. The primary outcome was the degree of wound re-epithelialization, assessed on day 15 post-injury. Secondary outcomes were cosmetic appearance and adverse events, evaluated six months post-treatment.

**Results:** The patient showed near-complete wound re-epithelialization by day 15 post-injury, without surgical excision or skin grafting. At the six-month follow-up, the wound had fully healed without visible scarring or contractures. fESWT was well-tolerated; no adverse events or complications were reported during or after the procedure.

**Conclusions:** This case report suggested that fESWT may offer a promising non-invasive alternative for promoting wound healing in deep partial thickness burns, with favorable outcomes in healing speed and cosmetic appearance. These findings were encouraging but based on a single patient and require confirmation in well-designed controlled clinical trials to establish the efficacy, safety and optimal treatment parameters of fESWT for burn wound management.

| **V** | **Result** |
| --- | --- |
| V1 | Partial thickness burn |
| V2 | Not provided |
| V3 | Acute; treatment began on day 3 post-injury |
| V4 | Case report |
| V5 | Category 5 |
| V6 | fESWT + SWC |
| V7 | Not applicable |
| V8 | 1 |
| V9 | Not applicable |
| V10 | Evotron (High Medical Technologies (HMT), Lengwil, Switzerland) |
| V11 | Focused, electrohydraulic ESWT |
| V12 | 0.11 mJ/mm^2^ |
| V13 | Not specified |
| V14 | 1500 ESWs per session |
| V15 | Not provided |
| V16 | Not provided |
| V17 | 2 sessions |
| V18 | 4-day interval (Day 3 and Day 7 post-injury) |
| V19 | Degree of wound re-epithelialization |
| V20 | Day 15 post-injury |
| V21 | Cosmetic appearance and presence or absence of adverse events; no biopsies or tissue samples analyzed. |
| V22 | Six months post-treatment |
| V23 | Not applicable |
| V24 | Not applicable |
| V25 | Near-complete reepithelization by Day 15 post-injury; no control group; no statistical comparison |
| V26 | Complete healing at six months post-treatment |
| V27 | Not performed |
| V28 | Not applicable |
| V29 | No treatment-related adverse events observed |
| V30 | Yes |
| V31 | No |
| V32 | No |
| V33 | Not applicable |
| V34 | No |
| V35 | No |
| V36 | No |
| V37 | Yes |
| V38 | Yes |
| V39 | Not applicable |
| V40 | No |

**[77]	Schaden, W.; Thiele, R.; Kölpl, C.; Pusch, M.; Nissan, A.; Attinger, C.E.; Maniscalco-Theberge, M.E.; Peoples, G.E.; Elster, E.A.; Stojadinovic, A. Shock wave therapy for acute and chronic soft tissue wounds: A feasibility study. *J. Surg. Res.* 2007, *143*(1), 1–12. https://doi.org/10.1016/j.jss.2007.01.009.**

**Hypothesis:** This study tested the hypothesis that uESWT is feasible, safe and potentially effective for healing acute and chronic soft tissue wounds of various etiologies, including those unresponsive to standard treatments.

**Methods:** This prospective case series enrolled 208 patients with nonhealing acute or chronic soft tissue wounds: postoperative dehiscence, posttraumatic necrosis, venous and arterial insufficiency ulcers, PUs (decubitus and plaster cast-related) and burns. All received uESWT with SWC; no control group was included. A DermaGold device (TRT, Alpharetta, GA, USA) delivered 100 ESWs/cm^2^ wound surface area at an EFD of 0.1 mJ/mm^2^ and 5 Hz, totaling 100–1000 ESWs per session; the average number of sessions was 2.8, at one- to two-week intervals. The primary outcome was the proportion achieving complete healing (100% epithelialization) over a mean follow-up of 44 days after the first treatment; secondary outcomes were safety and predictors of success.

**Results:** 156 of the 208 patients (75%) achieved complete epithelialization. Smaller wounds (<10 cm^2^), shorter duration (<1 month) and patient age were independent predictors of healing in multivariate logistic regression (age p = 0.01; wound size ≤10 cm^2^ p = 0.01, OR = 0.36, 95% CI 0.16–0.80; duration ≤1 month p < 0.001, OR = 0.25, 95% CI 0.11–0.55). Complete healing was highest in burns, posttraumatic wounds and plaster cast-related ulcers (100%, 86.6% and 85.7%) and by far lowest in venous stasis ulcers (36.0%). No adverse events, infections or wound deteriorations occurred and no anesthesia was required. Average time to healing was 43.5 days.

**Conclusions:** Unfocused ESWT is a feasible, safe and well-tolerated outpatient treatment for varied nonhealing acute and chronic soft tissue wounds. The high proportion of complete healing, particularly in small wounds of short duration, supports its clinical utility. RCTs comparing uESWT with SWC are warranted.

| **V** | **Result** |
| --- | --- |
| V1 | Mixed: post-traumatic, postoperative, venous/arterial ulcers, PUs, burns |
| V2 | Mean wound size: 9.4 cm^2^ |
| V3 | Mixed (acute: 33.2%, chronic: 66.8%) |
| V4 | Case series |
| V5 | Category 5 |
| V6 | uESWT + SWC |
| V7 | Not applicable |
| V8 | 208 |
| V9 | Not applicable |
| V10 | DermaGold (TRT, Alpharetta, GA, USA) |
| V11 | Unfocused, electrohydraulic ESWT |
| V12 | 0.1 mJ/mm^2^ |
| V13 | Not specified |
| V14 | 100–1000 ESWs |
| V15 | 100 ESWs/cm^2^ |
| V16 | 5 Hz |
| V17 | Mean of 2.8 sessions |
| V18 | Weekly to biweekly intervals |
| V19 | Primary endpoints designated feasibility and safety; complete healing (100% epithelialization) the efficacy measure; V19/V21 effectively reversed. |
| V20 | Mean follow-up of 44 days (median 31 days) |
| V21 | Safety (toxicity, infection, deterioration), predictive variables for healing; no biopsies or tissue samples analyzed |
| V22 | Within 6.3 weeks (mean), time frame overlaps with primary |
| V23 | No |
| V24 | Not applicable |
| V25 | 156/208 patients (75%) achieved 100% healing; no control group; no statistical comparison |
| V26 | Predictive factors identified (age, wound size <10 cm^2^, duration <1 month) |
| V27 | No |
| V28 | Not applicable |
| V29 | None reported (no toxicity, infection, or deterioration) |
| V30 | Yes |
| V31 | No |
| V32 | No |
| V33 | Not applicable |
| V34 | No |
| V35 | No |
| V36 | No |
| V37 | No |
| V38 | Yes |
| V39 | Not applicable |
| V40 | Yes |

**[78]	Dumfarth, J.; Zimpfer, D.; Vögele-Kadletz, M.; Holfeld, J.; Sihorsch, F.; Schaden, W.; Czerny, M.; Aharinejad, S.; Wolner, E.; Grimm, M. Prophylactic low-energy shock wave therapy improves wound healing after vein harvesting for coronary artery bypass graft surgery: A prospective, randomized trial. *Ann. Thorac. Surg.* 2008, *86*(6), 1909–1913. https://doi.org/10.1016/j.athoracsur.2008.07.117.**

**Hypothesis:** This study tested the hypothesis that prophylactic uESWT improves postoperative wound healing at the site of saphenous vein harvesting in patients undergoing CABG, compared to SWC without ESWT.

**Methods:** This prospective RCT investigated ESWT for acute surgical wounds after saphenous vein harvesting. 100 consecutive patients undergoing elective CABG were randomized 1:1 to SWC plus a single prophylactic uESWT session (uESWT group) or SWC alone (control group). Unfocused ESWT was applied immediately after wound closure with a DermaGold device (TRT, Alpharetta, GA, USA): 25 ESWs/cm of wound length, an EFD of 0.1 mJ/mm^2^ and a frequency of 5 Hz; only one session was given, with no further sessions or intervals. The primary outcome was wound healing on the ASEPSIS score (a composite of clinical signs of infection and treatment requirements), measured daily from postoperative days 3 through 7; secondary outcomes were need for surgical revision and antibiotics for wound healing disturbances during the in-hospital period.

**Results:** Groups were well matched for demographics, comorbidities and operative variables, including wound length (39 ± 13 cm versus 37 ± 11 cm; p = 0.342). The ASEPSIS score was significantly lower in the ESWT group (4.4 ± 5.3) than in controls (11.6 ± 8.3; p < 0.001), indicating improved healing. Fewer uESWT patients required antibiotics for wound complications (4% versus 22%; p = 0.015), and there was a trend toward fewer surgical revisions (2% versus 10%; p = 0.092). No adverse events related to uESWT were observed.

**Conclusions:** A single prophylactic session of uESWT significantly improved early postoperative wound healing after saphenous vein harvesting for CABG, with better ASEPSIS scores, reduced need for antibiotics and no associated adverse effects. Unfocused ESWT appears a safe, effective and easily applicable strategy to enhance surgical wound healing in cardiac surgery patients.

| **V** | **Result** |
| --- | --- |
| V1 | Surgical wounds from saphenous vein harvesting (post- coronary artery bypass grafting) |
| V2 | ESWT: 39 ± 13 cm wound length; control: 37 ± 11 cm wound length |
| V3 | Acute |
| V4 | RCT |
| V5 | Category 1 |
| V6 | Prophylactic uESWT + SWC |
| V7 | SWC |
| V8 | 50 |
| V9 | 50 |
| V10 | DermaGold (TRT, Alpharetta, GA, USA) |
| V11 | Unfocused, electrohydraulic ESWT |
| V12 | 0.1 mJ/mm^2^ |
| V13 | Not specified |
| V14 | Not provided |
| V15 | 25 ESWs/cm wound length, applied post-wound closure |
| V16 | 5 Hz |
| V17 | 1 |
| V18 | Not applicable |
| V19 | ASEPSIS score (quantitative assessment of wound healing) |
| V20 | Postoperative days 3–7 |
| V21 | Need for surgical revision; need for antibiotics; no biopsies or tissue samples analyzed |
| V22 | During in-hospital stay (not further specified) |
| V23 | Yes |
| V24 | Based on power = 0.8, alpha = 0.05 |
| V25 | ASEPSIS score: ESWT 4.4 ± 5.3 vs control 11.6 ± 8.3, p < 0.001 |
| V26 | Antibiotic use: ESWT 4% vs control 22%, p = 0.015; surgical revision: ESWT 2% vs control 10%, p = 0.092 |
| V27 | No |
| V28 | Not applicable |
| V29 | No SWT-associated adverse events reported |
| V30 | Yes |
| V31 | Yes |
| V32 | No |
| V33 | Yes |
| V34 | No |
| V35 | No |
| V36 | Yes |
| V37 | Yes |
| V38 | Yes |
| V39 | Yes |
| V40 | Yes |

**[79]	Saggini, R.; Figus, A.; Troccola, A.; Cocco, V.; Saggini, A.; Scuderi, N. Extracorporeal shock wave therapy for management of chronic ulcers in the lower extremities. *Ultrasound Med. Biol.* 2008, *34*(8), 1261–1271. https://doi.org/10.1016/j.ultrasmedbio.2008.01.010.**

**Hypothesis:** This study tested the hypothesis that fESWT significantly enhances the healing of chronic lower extremity ulcers compared to SWC.

**Methods:** This prospective controlled cohort study involved 40 patients with chronic lower extremity ulcers unresponsive to SWC for more than three months: posttraumatic ulcers (n=16), VLUs (n=12) and diabetic ulcers (n=4). Thirty patients (32 ulcers) received fESWT + SWC and 10 patients only SWC. Focused ESWT used an Evotron device (High Medical Technologies (HMT), Lengwil, Switzerland), with an EFD of 0.037 mJ/mm^2^ and 100 ESWs/cm^2^ of wound surface per session at 4 Hz (240 ESWs/min), for 4 to 10 sessions at two-week intervals. The primary outcome was healing, defined by complete closure or statistically significant size reduction, assessed 8 to 20 weeks after treatment start, depending on sessions; secondary outcomes were pain (Numeric Box Scale; NBS), exudate, granulation tissue and necrotic tissue, also to 20 weeks post-baseline.

**Results:** Sixteen of 32 ulcers (50%) treated with fESWT healed completely within four to six sessions. Among nonhealed ulcers, wound size was significantly reduced (p < 0.01), by a mean of 37.5% in posttraumatic, 45% in venous and 50% in diabetic ulcers, with significant improvements in granulation tissue percentage and reduction of exudates (p < 0.01). Pain scores (NBS) fell significantly from a mean of 6.7 to lower values post-treatment (p < 0.001). Only one control ulcer healed completely, with no significant improvement in wound size, exudate, granulation tissue or pain. No adverse events were reported in the fESWT group.

**Conclusions:** Focused ESWT appears effective and safe for chronic lower extremity ulcers, particularly posttraumatic cases, with significantly better outcomes than SWC in wound closure, tissue regeneration and pain reduction, suggesting fESWT as a viable noninvasive adjunct. Further RCTs are warranted to optimize the protocol and validate these preliminary results.

| **V** | **Result** |
| --- | --- |
| V1 | Chronic ulcers (venous, posttraumatic, diabetic) |
| V2 | Mean surface area: 5.3 cm^2^ |
| V3 | Chronic; duration >3 months, mean 5.4 months |
| V4 | Cohort study with non-randomized control group |
| V5 | Category 1 |
| V6 | fESWT + SWC |
| V7 | SWC |
| V8 | 30 patients (32 ulcers) |
| V9 | 10 patients |
| V10 | Evotron (High Medical Technologies (HMT), Lengwil, Switzerland) |
| V11 | Focused, electrohydraulic ESWT |
| V12 | 0.037 mJ/mm^2^ |
| V13 | Not specified |
| V14 | Not provided |
| V15 | 100 ESWs/cm^2^ |
| V16 | 4 Hz |
| V17 | 4 to 10 sessions (mean not clearly specified) |
| V18 | Every 2 weeks |
| V19 | Healing of ulcers: complete closure, reduction in surface area, granulation tissue, pain |
| V20 | 8–20 weeks post-baseline (depending on number of sessions) |
| V21 | Pain (NBS score), exudates, granulation tissue, bacterial culture; excision biopsy after fESWT failure: osteomyelitis, *Staphylococcus aureus*; no others |
| V22 | Post each session and at end of treatment (8–20 weeks) |
| V23 | No |
| V24 | Not applicable |
| V25 | fESWT group: 50% healed completely; significant reductions in surface area, pain (p<0.01); control group: minimal improvement |
| V26 | fESWT group: pain reduced (mean NBS from 6.65 to ~5); improved granulation tissue and exudates; control group: no significant changes |
| V27 | No |
| V28 | Not applicable |
| V29 | No adverse events reported |
| V30 | Yes |
| V31 | No |
| V32 | No |
| V33 | No |
| V34 | No |
| V35 | No |
| V36 | No |
| V37 | Yes |
| V38 | Yes |
| V39 | Yes |
| V40 | Yes |

**[23]	Wang, C.J.; Kuo, Y.R.; Wu, R.W.; Liu, R.T.; Hsu, C.S.; Wang, F.S.; Yang, K.D. Extracorporeal shockwave treatment for chronic diabetic foot ulcers. *J. Surg. Res.* 2009, *152*(1), 96–103. https://doi.org/10.1016/j.jss.2008.01.026.**

**Hypothesis:** This study tested the hypothesis that fESWT is more effective than HBOT in healing chronic DFUs by enhancing tissue regeneration, local blood flow and cellular proliferation.

**Methods:** This prospective RCT evaluated fESWT versus HBOT in chronic DFUs: 72 ulcers from 70 patients, non-healing for more than three months, randomly assigned to fESWT or HBOT. fESWT used an Orthowave 180 device (MTS Medical, Konstanz, Germany), delivering 300 ± 100 ESWs/cm^2^ at an EFD of 0.11 mJ/cm^2^, every two weeks for three sessions over six weeks; the frequency in Hertz was not specified. HBOT was given daily (5 sessions per week) for four weeks in a multi-place chamber at 2.5 atmospheres absolute pressure, 90 minutes each. Both groups received SWC. The primary outcome was clinical healing at 6 weeks post-baseline; secondary outcomes were blood flow perfusion, bacterial colony counts and histological and immunohistochemical analyses (eNOS, VEGF, PCNA, TUNEL), at 6 weeks post-treatment and every three months.

**Results:** At six weeks post-treatment, 31% of ulcers in the fESWT group were completely healed, 58% showed more than 50% improvement and 11% were unchanged, versus 22%, 50% and 28% in the HBOT group (p = 0.001). Focused ESWT also significantly improved local blood flow perfusion (p = 0.04); both groups showed a comparable bacteriostatic effect. Histology showed higher cell proliferation, and immunohistochemistry showed significantly increased eNOS, VEGF and PCNA and decreased TUNEL expression, indicating reduced apoptosis, in the fESWT group versus HBOT (all p < 0.05). No treatment-related adverse events were reported in either group.

**Conclusions:** fESWT is a safe and effective adjunctive treatment for chronic DFUs. Compared to HBOT, it results in superior clinical healing, enhanced blood perfusion, increased cellular activity and proliferation, and reduced apoptosis, supporting fESWT as a non-invasive and promising therapeutic option for chronic DFUs.

| **V** | **Result** |
| --- | --- |
| V1 | Chronic DFUs |
| V2 | fESWT: 11.2 ± 20.0 cm^2^; HBOT: 10.5 ± 20.0 cm^2^ |
| V3 | Chronic; mean duration fESWT: 22.7 ± 20.9 months; HBOT: 19.0 ± 19.5 months |
| V4 | RCT |
| V5 | Category 1 |
| V6 | fESWT + SWC |
| V7 | HBOT (20 daily sessions at 2.5 ATA for 90 minutes) + SWC |
| V8 | 34 patients, 36 ulcers |
| V9 | 36 patients, 36 ulcers |
| V10 | Orthowave 180 (MTS Medical, Konstanz, Germany) |
| V11 | Focused, electrohydraulic ESWT |
| V12 | 0.11 mJ/cm^2^ |
| V13 | Not specified |
| V14 | 300 ESWs + 100 ESWs/cm^2^ of ulcer surface per session |
| V15 | Not provided |
| V16 | Not provided |
| V17 | 3 sessions (every 2 weeks over 6 weeks) |
| V18 | 2 weeks |
| V19 | Clinical healing of the ulcer (healed, >50% improved, unchanged) |
| V20 | 6 weeks post-baseline |
| V21 | Bacteriology, histology, immunohistochemistry, blood perfusion; biopsies from the most contaminated ulcer area |
| V22 | 6 weeks post-baseline and every 3 months thereafter |
| V23 | Yes |
| V24 | Sample size of 23 required for α = 0.05, power = 0.8 |
| V25 | fESWT: 31% healed, 58% improved, 11% unchanged; HBOT: 22% healed, 50% improved, 28% unchanged (p = 0.001) |
| V26 | fESWT showed significantly better perfusion, cell concentration, eNOS, VEGF, PCNA; lower TUNEL vs HBOT (p < 0.05) |
| V27 | No |
| V28 | Not applicable |
| V29 | No treatment-related adverse events reported |
| V30 | Yes |
| V31 | No (quasi-random allocation by treatment date) |
| V32 | No |
| V33 | Yes |
| V34 | No |
| V35 | No |
| V36 | Yes |
| V37 | Yes |
| V38 | Yes |
| V39 | Yes |
| V40 | Yes |

**[80]	Arnó, A.; García, O.; Hernán, I.; Sancho, J.; Acosta, A.; Barret, J.P. Extracorporeal shock waves, a new non-surgical method to treat severe burns. *Burns* 2010, *36*(6), 844–849. https://doi.org/10.1016/j.burns.2009.11.012.**

**Hypothesis:** This study tested the hypothesis that uESWT enhances healing in deep partial- and full-thickness burns by increasing tissue perfusion and promoting re-epithelialization, thereby reducing the need for surgical intervention.

**Methods:** This prospective case series investigated uESWT in acute thermal burns in 15 adults (10 men and 5 women) with deep partial- and full-thickness burns involving less than 5% of the total body surface area. All received the same semi-occlusive burn care and two uESWT sessions, on the third and fifth day post-injury, with a dermaPACE device (Sanuwave Health, Eden Prairie, MN, USA): 500 ESWs per session at an EFD of 0.15 mJ/mm^2^, approximately 100 ESWs per cm^2^ of wound area. The frequency in Hertz was not reported; treatment duration per cm^2^ was described qualitatively as 1–2 minutes. The primary outcome was spontaneous re-epithelialization by visual inspection and digital photography up to three weeks post-injury; secondary outcomes were LDI perfusion (days 3 and 5), pain on the visual analogue scale (VAS) and adverse events.

**Results:** Fourteen patients completed the study; one was lost to follow-up. 12 of the 14 (86%) healed completely and spontaneously without surgery, in approximately 15 days on average. Two patients with burns on the dorsum of the hand required surgical debridement and grafting due to insufficient re-epithelialization by day 18. LDI showed significant increases in tissue perfusion after the first uESWT session. Pain was minimal, with all VAS scores below 3. No adverse events such as bleeding, hematoma or infection occurred. One patient developed mild hypertrophic scarring post-grafting.

**Conclusions:** This pilot study suggested that uESWT may be a safe, non-invasive and effective adjunct for deep partial- and full-thickness burns, promoting spontaneous healing and reducing the need for surgery. The absence of a control group limited the conclusions; larger controlled trials are warranted.

| **V** | **Result** |
| --- | --- |
| V1 | Deep partial/full thickness burns |
| V2 | Not provided (only total body surface area <5%) |
| V3 | Acute (treated from day 3 post-injury) |
| V4 | Case series |
| V5 | Category 5 |
| V6 | uESWT + semi-occlusive burn care |
| V7 | Not applicable |
| V8 | 15 (1 lost to follow-up) |
| V9 | Not applicable |
| V10 | dermaPACE (Sanuwave Health, Eden Prairie, MN, USA) |
| V11 | Unfocused, electrohydraulic ESWT |
| V12 | 0.15 mJ/mm^2^ |
| V13 | Not specified |
| V14 | 500 |
| V15 | 100 ESWs/cm^2^ |
| V16 | Not provided |
| V17 | 2 |
| V18 | 2 days (day 3 and day 5 post-injury) |
| V19 | Wound healing (spontaneous re-epithelialization) |
| V20 | Weekly for 1 month, then monthly; spontaneous healing times recorded ranged from 10 to 29 days |
| V21 | Perfusion changes (LDI), pain (VAS), adverse effects; no biopsies or tissue samples analyzed |
| V22 | Day 3 and day 5 post-injury, during and after treatment (VAS), follow-up until healing |
| V23 | No |
| V24 | Not applicable |
| V25 | 12/14 patients (86%) healed without surgery; 2 required grafting |
| V26 | LDI showed improved perfusion post-uESWT; VAS <3; no adverse effects |
| V27 | No |
| V28 | Not applicable |
| V29 | None observed |
| V30 | Yes |
| V31 | No |
| V32 | No |
| V33 | Not applicable |
| V34 | No |
| V35 | No |
| V36 | No |
| V37 | Yes |
| V38 | Yes |
| V39 | Not applicable |
| V40 | No |

**[81]	Larking, A.M.; Duport, S.; Clinton, M.; Hardy, M.; Andrews, K. Randomized control of extracorporeal shock wave therapy versus placebo for chronic decubitus ulceration. *Clin. Rehabil.* 2010, *24*(3), 222–229. https://doi.org/10.1177/0269215509346083.**

**Hypothesis:** This study tested the hypothesis that uESWT accelerates healing in patients with chronic decubitus ulceration compared to placebo treatment.

**Methods:** This randomized, double-blind, placebo-controlled cross-over trial in a long-stay hospital for complex neurological disabilities included chronic decubitus ulcers (PUs) persisting more than three months without healing during a three-week baseline. Ulcers were randomly assigned to uESWT or placebo first, then a two-week washout and crossover. Unfocused ESWT used an Orthowave 180c device (MTS Medical, Konstanz, Germany) with a non-focused applicator: one weekly session for four weeks, 200 ESWs plus 100 ESWs/cm^2^, EFD 0.1 J/mm^2^, 5 Hz. Placebo used an identical device head producing no shock waves but mimicking active therapy. The primary endpoint was change in ulcer area, assessed weekly, particularly healing at 6–8 weeks after the start of uESWT, depending on treatment order; no secondary outcomes were defined.

**Results:** Nine ulcers in eight patients were included, five randomized to placebo first and four to uESWT; all had been static during baseline. Significant improvement in ulcer area occurred 6 weeks after initiation of uESWT in the uESWT-first group and 8 weeks in the placebo-first group (i.e., 8 weeks after crossover, equivalent to 14 weeks after study start). Improvement occurred only after active uESWT began, not during placebo or washout, and mean ulcer area decreased more in the uESWT phases, confirming a time-linked effect. In some ulcers with ischemic borders, a transient size increase preceded faster healing, suggesting debridement-like action. No adverse events were reported.

**Conclusions:** This RCT provided preliminary evidence that uESWT may significantly promote healing of chronic, non-healing decubitus ulcers in severely disabled patients. Improvements were time-linked to active therapy and not placebo, supporting a true therapeutic effect and suggesting uESWT as a valuable non-invasive adjunct for PUs. Larger trials are warranted.

| **V** | **Result** |
| --- | --- |
| V1 | Chronic decubitus ulceration |
| V2 | uESWT-first: 1.23 cm^2^; placebo-first: 1.79 cm^2^ |
| V3 | Chronic; mean duration 54 weeks (range 12–156 weeks) |
| V4 | RCT (cross-over) |
| V5 | Category 1 |
| V6 | uESWT + SWC |
| V7 | Placebo device (inactive head with same appearance/sound) + SWC |
| V8 | 4 ulcers initially; 9 ulcers total after crossover |
| V9 | 5 ulcers initially; 9 ulcers total after crossover |
| V10 | Orthowave 180c (MTS Medical, Konstanz, Germany) |
| V11 | Unfocused, electrohydraulic ESWT |
| V12 | 0.1 J/mm^2^ |
| V13 | Not specified |
| V14 | 200 ESWs + 100 ESWs/cm^2^ |
| V15 | 100 ESWs/cm^2^ |
| V16 | 5 Hz |
| V17 | 4 sessions |
| V18 | 1 week |
| V19 | Change in ulcer area |
| V20 | Weekly; significant change noted at 6–8 weeks post-uESWT start |
| V21 | None explicitly defined or assessed; no biopsies or tissue samples analyzed |
| V22 | Not applicable |
| V23 | No |
| V24 | Not applicable |
| V25 | Significant reduction in ulcer area from baseline in the weeks following uESWT (p < 0.05); no direct between-group uESWT-versus-placebo test was reported |
| V26 | Not applicable |
| V27 | No |
| V28 | Not applicable |
| V29 | None reported |
| V30 | Yes |
| V31 | Yes |
| V32 | Yes |
| V33 | Yes |
| V34 | Yes |
| V35 | No |
| V36 | Yes |
| V37 | Yes |
| V38 | No |
| V39 | No |
| V40 | Yes |

**[82]	Ottomann, C.; Hartmann, B.; Tyler, J.; Maier, H.; Thiele, R.; Schaden, W.; Stojadinovic, A. Prospective randomized trial of accelerated re-epithelization of skin graft donor sites using extracorporeal shock wave therapy. *J. Am. Coll. Surg.* 2010, *211*(3), 361–367. https://doi.org/10.1016/j.jamcollsurg.2010.05.012.**

**Hypothesis:** This study tested the hypothesis that uESWT of split-thickness skin graft (STSG) donor sites significantly accelerates re-epithelialization compared to SWC alone.

**Methods:** This prospective RCT included patients with acute traumatic wounds and burns requiring STSG. 28 patients were enrolled and randomized 1:1. The intervention group (n = 13) received a single intraoperative uESWT session immediately after skin graft harvesting, in addition to SWC; the control group (n = 15) received only SWC, a nonadherent silicone mesh dressing with an antiseptic gel, applied daily. Unfocused ESWT used a DermaGold device (TRT, Alpharetta, GA, USA), with a single application of 100 ESWs/cm^2^ at an EFD of 0.1 mJ/mm^2^. The frequency in Hertz and the total number of ESWs per session in absolute values were not provided; no additional sessions were given and no interval was applicable. The primary outcome was time to complete donor site healing, defined as ≥95% re-epithelialization, assessed daily up to 21 days postoperatively. No secondary outcomes were prespecified or assessed.

**Results:** All patients completed the trial; no adverse events were reported in either group. The mean time to complete donor site epithelialization was significantly shorter in the uESWT group (13.9 ± 2.0 days) than in the control group (16.7 ± 2.0 days) (p < 0.001). This improvement prompted early termination of the study at interim analysis. No infections, allergic reactions or complications associated with uESWT were observed.

**Conclusions:** A single application of uESWT immediately following skin graft harvest significantly accelerates donor site re-epithelialization compared to SWC alone. This noninvasive, well-tolerated therapy holds promise as a clinically relevant adjunct in the management of STSG donor sites. Larger, multicenter trials are warranted to confirm these findings and to assess additional outcomes such as pain, quality of life and cosmetic results.

| **V** | **Result** |
| --- | --- |
| V1 | Acute traumatic wounds and burns requiring skin grafting |
| V2 | Not provided |
| V3 | Acute |
| V4 | RCT |
| V5 | Category 1 |
| V6 | uESWT + SWC (nonadherent silicone mesh + antiseptic gel) |
| V7 | SWC |
| V8 | 13 |
| V9 | 15 |
| V10 | DermaGold (TRT, Alpharetta, GA, USA) |
| V11 | Unfocused, electrohydraulic ESWT |
| V12 | 0.1 mJ/mm^2^ |
| V13 | Not specified |
| V14 | 100 ESWs/cm^2^ |
| V15 | 100 ESWs/cm^2^ |
| V16 | Not provided |
| V17 | 1 |
| V18 | Not applicable (single session) |
| V19 | Time to complete donor site healing (≥95% re-epithelialization) |
| V20 | Up to 21 days (median 15 days), daily assessment |
| V21 | None explicitly defined or assessed; no biopsies or tissue samples analyzed |
| V22 | Not applicable |
| V23 | Yes |
| V24 | 27 subjects per group needed for 80% power to detect 2-day difference with α = 0.01 |
| V25 | ESWT: 13.9 ± 2.0 days; control: 16.7 ± 2.0 days (p < 0.001) |
| V26 | Not applicable |
| V27 | No |
| V28 | Not applicable |
| V29 | No adverse events reported |
| V30 | Yes |
| V31 | Yes |
| V32 | Yes |
| V33 | Yes |
| V34 | Yes |
| V35 | No |
| V36 | Yes |
| V37 | Yes |
| V38 | Yes |
| V39 | Yes |
| V40 | Yes |

**[30]	Wang, C.J.; Wu, R.W.; Yang, Y.J. Treatment of diabetic foot ulcers: A comparative study of extracorporeal shockwave therapy and hyperbaric oxygen therapy. *Diabetes Res. Clin. Pract.* 2011, *92*(2), 187–193. https://doi.org/10.1016/j.diabres.2011.01.019.**

**Hypothesis:** This study tested the hypothesis that fESWT is more effective than HBOT for chronic DFUs, improving healing through enhanced perfusion and cellular regeneration.

**Methods:** This prospective RCT compared fESWT and HBOT for chronic DFUs unhealed for more than three months: 77 patients with 84 DFUs, 39 patients (44 feet) fESWT, 38 patients (40 feet) HBOT. Focused ESWT used a dermaPACE device (Sanuwave Health, Eden Prairie, MN, USA), twice per week for three weeks (six sessions), each of a minimum of 500 ESWs, calculated as 8 times the treatment area in cm^2^ (the ulcer extended by 1.0 cm in all directions); the EFD was 0.23 mJ/mm^2^, the frequency 4 Hz. HBOT comprised 20 daily 90-minute treatments at 2.5 atmospheres absolute pressure; both groups also received SWC. The primary outcome was clinical healing after the first course (3 weeks); secondary outcomes were blood flow perfusion and histopathology (cell proliferation, apoptosis), before and after treatment.

**Results:** After the first course, complete healing occurred in 57% of fESWT feet versus 25% of HBOT feet (p = 0.003); ≥50% improvement in 32% versus 15% (p = 0.071); unchanged ulcers were significantly fewer with fESWT (11% vs. 60%; p < 0.001). A second course healed 50% of remaining fESWT ulcers versus 6% of HBOT ulcers (p = 0.005). Blood perfusion increased significantly after fESWT (p < 0.001) but not after HBOT (p = 0.916). Histology showed increased cell proliferation and decreased apoptosis with fESWT. No treatment-related adverse events occurred with fESWT; four HBOT patients had transient barotrauma-related symptoms.

**Conclusions:** fESWT is significantly more effective than HBOT for chronic DFUs, enhancing local perfusion and cellular activity and producing higher rates of complete healing. It may therefore be a superior adjunctive therapy for chronic diabetic ulcers, offering benefits in both efficacy and safety.

| **V** | **Result** |
| --- | --- |
| V1 | Chronic DFUs |
| V2 | fESWT group: median 4 cm^2^ (range 1.5–9); HBOT group: median 7 cm^2^ (range 2–12) |
| V3 | Chronic; median 6 months (range 3–16 months) |
| V4 | RCT |
| V5 | Category 1 |
| V6 | fESWT + SWC |
| V7 | HBOT + SWC |
| V8 | 39 (44 feet) |
| V9 | 38 (40 feet) |
| V10 | dermaPACE (Sanuwave Health, Eden Prairie, MN, USA) |
| V11 | Focused, electrohydraulic ESWT |
| V12 | 0.23 mJ/mm^2^ |
| V13 | Not specified |
| V14 | Minimum 500 ESWs; actual: treatment area (cm^2^) × 8, treatment area = ulcer perimeter extended 1.0 cm in all directions |
| V15 | 8 ESWs/cm^2^ |
| V16 | 4 Hz |
| V17 | 6 sessions |
| V18 | Twice per week |
| V19 | Ulcer healing status: complete healing, ≥50% improvement, unchanged, worsened |
| V20 | After 6 treatments (~3 weeks); optional second course evaluated 4–6 weeks later |
| V21 | Blood perfusion scan; ulcer-edge biopsies, histopathology: cell proliferation, cell concentration, cell activity, apoptosis |
| V22 | After treatment completion (~3 weeks) |
| V23 | Yes |
| V24 | 40 patients per group needed for α = 0.05 and power = 0.80 |
| V25 | fESWT group: 57% healed; HBOT group: 25% healed (p = 0.003) |
| V26 | fESWT: perfusion ↑ (p < 0.001), cell proliferation ↑, apoptosis ↓; HBOT: perfusion unchanged (p = 0.916) |
| V27 | Not provided |
| V28 | Not provided |
| V29 | HBOT group: 4 patients with barotrauma/sinus pain; fESWT group: no adverse events |
| V30 | Yes |
| V31 | Yes |
| V32 | No |
| V33 | Yes |
| V34 | No |
| V35 | No |
| V36 | No (except pathologists for histology) |
| V37 | Yes |
| V38 | No |
| V39 | Yes |
| V40 | Yes |

**[83]	Wolff, K.S.; Wibmer, A.; Pusch, M.; Prusa, A.M.; Pretterklieber, M.; Teufelsbauer, H.; Schaden, W. The influence of comorbidities and etiologies on the success of extracorporeal shock wave therapy for chronic soft tissue wounds: Midterm results. *Ultrasound Med. Biol.* 2011, *37*(7), 1111–1119. https://doi.org/10.1016/j.ultrasmedbio.2011.04.007.**

**Hypothesis:** This study tested the hypothesis that uESWT is effective for chronic soft tissue wounds, with success not significantly influenced by patient comorbidities or wound etiology.

**Methods:** This one-armed, open, prospective case series enrolled 282 adults with chronic soft tissue wounds persisting ≥30 days, unresponsive to SWC: VLUs, decubitus, arterial, cast pressure, post-surgical, post-traumatic and burn wounds (noncircumferential deep second- or third-degree; circumferential second-degree or higher extremity burns excluded). Unfocused ESWT alone used an Orthowave 180C device (MTS Medical, Konstanz, Germany), delivering unfocused planar ESWs at an EFD of 0.1 mJ/mm^2^ and 5 Hz: median 167 ESWs/cm^2^ wound area (range: 2.7–2400/cm^2^), median two sessions (interquartile range: 1–4, maximum 10), weekly at first, every two weeks after the second session. The primary endpoint was all-cause mortality within 30 days of the final session; secondary outcomes were complete closure, sessions needed and non-healing rate, over a median 31.8-month follow-up.

**Results:** 258 of 282 patients (91.5%) were available for analysis. Median wound surface was 5.0 cm^2^; most wounds (76.4%) had lasted 4 to 12 weeks. Treatment success, defined as complete closure or a final wound bed score (WBS) of ≥14, was achieved in 191 patients (74.1%), with a median of 2 sessions over a median 14 days. Wound duration, surface area and initial WBS were significant predictors in multivariate logistic regression; comorbidities and wound etiology had no statistically significant impact. No treatment-related adverse events occurred and no deaths within 30 days of the final session were due to the therapy. Median WBS improvement was 4 points.

**Conclusions:** uESWT is a safe and effective therapy for chronic soft tissue wounds of varied etiology. Success was not significantly influenced by comorbidities or wound cause but was strongly associated with local characteristics: surface area, duration and wound bed condition.

| **V** | **Result** |
| --- | --- |
| V1 | Chronic soft tissue wounds (postsurgical, post-traumatic, VLUs, decubitus ulcer, arterial ulcer, cast pressure sore, burn) |
| V2 | Median surface = 5.0 cm^2^ (IQR: 2.0–14.0; range: 0.5–300) |
| V3 | Chronic (4–12 weeks: 76.4%; 4–12 months: 12.8%; >1 year: 10.9%) |
| V4 | Case series |
| V5 | Category 5 |
| V6 | uESWT + SWC |
| V7 | Not applicable |
| V8 | 282 (24 lost to follow-up) |
| V9 | Not applicable |
| V10 | Orthowave 180C (MTS Medical, Konstanz, Germany) |
| V11 | Unfocused, electrohydraulic ESWT |
| V12 | 0.1 mJ/mm^2^ |
| V13 | Not specified |
| V14 | Median 167 ESWs/cm^2^ (IQR: 100–300; range: 2.7–2400) |
| V15 | Yes; ESWs/cm^2^ provided |
| V16 | 5 Hz |
| V17 | Median 2 sessions (IQR: 1–4; range: 1–10) |
| V18 | Weekly initially, then biweekly |
| V19 | All-cause mortality within 30 days post-treatment |
| V20 | 30 days after last ESWT session |
| V21 | Complete wound closure, number of treatment sessions, non-healing rate, sessions until dropout; no biopsies or tissue samples analyzed |
| V22 | Up to 31.8 months (median follow-up) |
| V23 | No |
| V24 | Not applicable |
| V25 | 74.1% wound closure (191/258); no control group, no between-group stats |
| V26 | Median WBS improvement = 4 points (initial median = 9; final median = 14) |
| V27 | No |
| V28 | Not applicable |
| V29 | No treatment-related adverse events observed |
| V30 | Yes |
| V31 | No |
| V32 | No |
| V33 | Not applicable |
| V34 | No |
| V35 | No |
| V36 | No |
| V37 | Yes |
| V38 | Yes |
| V39 | No |
| V40 | Yes |

**[84]	Fioramonti, P.; Onesti, M.G.; Fino, P.; Fallico, N.; Scuderi, N. Extracorporeal shock wave therapy for the treatment of VLUs in the lower limbs. *Ann. Ital. Chir.* 2012, *83*(1), 41–44.**

**Hypothesis:** This study tested the hypothesis that fESWT significantly improves the healing of chronic VLUs by enhancing tissue regeneration and promoting revascularization compared to conventional wound care.

**Methods:** This was a case report describing the treatment of a 63-year-old female patient with chronic VLUs on both lower limbs, caused by chronic venous insufficiency. The right leg received fESWT, while the left leg was treated with SWC. Focused ESWT was performed using an Evotron device (High Medical Technologies (HMT), Lengwil, Switzerland) delivering ESWs with an EFD of 0.037 mJ/mm^2^. Each session applied 100 ESWs/cm^2^ at a frequency of 4 Hz. The patient received one session per week over a six-week period, totaling six treatment sessions. The control treatment consisted of weekly disinfection and application of medicated gauze. The primary endpoint was complete wound healing, assessed six weeks after the start of treatment. No secondary outcomes were specified or evaluated.

**Results:** At the conclusion of the six-week treatment period, the ulcers on the right leg treated with fESWT had completely healed, while the ulcer on the left leg treated with conventional dressings showed incomplete healing. No adverse events were reported in this case; bleeding, petechiae, hematoma, seroma formation and pain were mentioned only in the Discussion as complications described in the literature. The findings suggest a notable improvement in wound closure in the limb treated with fESWT compared to the limb treated with SWC.

**Conclusions:** This case report supports the potential effectiveness of fESWT as a non-invasive and well-tolerated therapeutic option for chronic VLUs. Compared to SWC, fESWT resulted in more rapid and complete healing. Further research with larger controlled studies is necessary to confirm these findings and determine the broader clinical applicability of fESWT in chronic wound management.

| **V** | **Result** |
| --- | --- |
| V1 | VLUs |
| V2 | fESWT: 1.5×2 cm and 4×2 cm; control: 4×1.5 cm (no mean provided) |
| V3 | Chronic |
| V4 | Case report |
| V5 | Category 5 |
| V6 | fESWT + SWC |
| V7 | SWC (disinfection + medicated gauze) |
| V8 | 1 |
| V9 | 1 (contralateral leg) |
| V10 | Evotron (High Medical Technologies (HMT), Lengwil, Switzerland) |
| V11 | Focused, electrohydraulic ESWT |
| V12 | 0.037 mJ/mm^2^ |
| V13 | Not specified |
| V14 | Not provided (100 ESWs/cm^2^ mentioned) |
| V15 | 100 ESWs/cm^2^ |
| V16 | 4 Hz |
| V17 | 6 |
| V18 | 1 week |
| V19 | Not provided |
| V20 | After 6 weeks |
| V21 | None explicitly defined or assessed; no biopsies or tissue samples analyzed |
| V22 | Not applicable |
| V23 | Not applicable |
| V24 | Not applicable |
| V25 | fESWT: complete healing; control: incomplete healing (no statistical analysis) |
| V26 | Not applicable |
| V27 | Not applicable |
| V28 | Not applicable |
| V29 | Possible: bleeding, petechiae, hematoma, seroma, pain (not observed in this case) |
| V30 | No |
| V31 | No |
| V32 | No |
| V33 | No |
| V34 | No |
| V35 | No |
| V36 | No |
| V37 | No |
| V38 | No |
| V39 | No |
| V40 | No |

**[85]	Ottomann, C.; Stojadinovic, A.; Lavin, P.T.; Gannon, F.H.; Heggeness, M.H.; Thiele, R.; Schaden, W.; Hartmann, B. Prospective randomized phase II trial of accelerated re-epithelialization of superficial second-degree burn wounds using extracorporeal shock wave therapy. *Ann. Surg.* 2012, *255*(1), 23–29. https://doi.org/10.1097/SLA.0b013e318227b3c0.**

**Hypothesis:** This study tested the hypothesis that uESWT of superficial, second-degree burn wounds significantly accelerates re-epithelialization compared to SWC alone.

**Methods:** This prospective RCT investigated wound healing in acute superficial second-degree burns. Fifty patients were enrolled and randomly assigned to uESWT + SWV or SWC alone. The uESWT group received a single session of uESWT within 24 hours of burn wound debridement using a DermaGold device (TRT, Alpharetta, GA, USA), applying 100 ESWs/cm^2^ with an EFD of 0.1 mJ/mm^2^ over 20 seconds per cm^2^; the frequency in Hertz was not specified and no further sessions were given. The control group received SWC only: debridement and daily topical antiseptic agents and silicone mesh dressings. The primary endpoint was time to complete epithelialization, defined as ≥95% epithelial coverage, assessed daily during hospitalization and at outpatient follow-up. No secondary endpoints were reported.

**Results:** Of the 50 patients enrolled, 44 were evaluable after excluding 6 for incomplete data or loss to follow-up. Mean time to complete epithelialization was significantly shorter in the uESWT group (9.6 ± 1.7 days) than in the control group (12.5 ± 2.2 days) (p < 0.001), and remained significant after adjusting for an age imbalance between groups and under worst-case and best-case imputation in the full cohort. No treatment-related adverse events were reported. Both groups were comparable at baseline except for age, which was higher in the uESWT group.

**Conclusions:** A single application of uESWT significantly accelerates wound healing in superficial second-degree burns. The therapy was safe, noninvasive and well-tolerated, with no observed adverse events, supporting the potential clinical utility of uESWT as an adjunct to standard burn care. Larger trials are needed to confirm the results and explore its mechanism of action and long-term outcomes.

| **V** | **Result** |
| --- | --- |
| V1 | Superficial second-degree burns |
| V2 | Not provided |
| V3 | Acute |
| V4 | RCT |
| V5 | Category 1 |
| V6 | uESWT + SWC (debridement + antiseptic therapy) |
| V7 | SWC alone |
| V8 | 22 |
| V9 | 22 |
| V10 | DermaGold (TRT, Alpharetta, GA, USA) |
| V11 | Unfocused, electrohydraulic ESWT |
| V12 | 0.1 mJ/mm^2^ |
| V13 | Not specified |
| V14 | 100 ESWs/cm^2^ |
| V15 | 100 ESWs/cm^2^ |
| V16 | Not provided (stated as “20 seconds/cm^2^” application time) |
| V17 | 1 |
| V18 | Not applicable (only 1 session) |
| V19 | Time to ≥95% epithelialization |
| V20 | Daily until complete epithelialization; mean time assessed at endpoint |
| V21 | None explicitly defined or assessed; no biopsies or tissue samples analyzed |
| V22 | Not applicable |
| V23 | No (stated sample size target without formal power calculation) |
| V24 | Not applicable |
| V25 | uESWT group: 9.6 ± 1.7 days; control group: 12.5 ± 2.2 days (p < 0.001) |
| V26 | Not applicable |
| V27 | Yes |
| V28 | >80% power for effect size 0.85 (2-day difference in mean time to healing), two-sided test, alpha = 0.05 |
| V29 | No adverse events attributed to ESWT |
| V30 | Yes |
| V31 | Yes |
| V32 | Yes |
| V33 | Yes |
| V34 | Yes |
| V35 | No |
| V36 | Yes |
| V37 | Yes |
| V38 | Yes |
| V39 | Yes |
| V40 | Yes |

**[86]	Saggini, R.; Fioramonti, P.; Bellomo, R.G.; Di Stefano, A.; Scarcello, L.; Di Pancrazio, L.; Iodice, R.; Saggini, A.; Scuderi, N. Chronic ulcers: Treatment with unfocused extracorporeal shock waves. *Eur. J. Inflamm.* 2013, *11*, 499–509. https://doi.org/10.1177/1721727X13011002.**

**Hypothesis:** This study tested the hypothesis that uESWT promotes significant healing of chronic ulcers by reducing wound size and improving pain levels compared to baseline values.

**Methods:** This RCT involved 124 patients aged 28 to 80 years with chronic ulcers of various etiologies (vascular, diabetic, pressure, post-traumatic, iatrogenic) for at least three months. Group A (62 patients) received uESWT with a DermaGold device (TRT, Alpharetta, GA, USA) at an EFD of 0.10 mJ/mm^2^, Group B (62 patients) with an Evotron device (High Medical Technologies (HMT), Lengwil, Switzerland) at an EFD of 0.04 mJ/mm^2^ (considered control treatment). Each session delivered 300 to 600 ESWs at 4 Hz (240 ESWs per minute). The primary outcome was wound area reduction after the seven-week treatment period; secondary outcomes were pain on the Visual Analog Scale (VAS) and perilesional skin trophism, monitored by photo capture.

**Results:** Both groups healed significantly over the seven weeks, Group A with an 80% reduction in wound area versus 67% in Group B; no between-group p-value was reported (the p < 0.001 values were within-group before/after comparisons). Pain decreased by 79% in Group A and 48% in Group B. Group A thus showed a greater area and VAS reduction, but no statistical comparison between the groups was reported. No bleeding or petechiae occurred, but 15 of the 22 group B patients who withdrew did so because of treatment-related pain; no infections were observed.

**Conclusions:** uESWT significantly accelerates wound healing and reduces pain in patients with chronic ulcers, with greater efficacy than the control group. ESWT was found to be safe and effective for chronic ulcers, an alternative to more invasive procedures, and the results support its use as an adjunctive therapy in wound care, particularly where traditional healing methods are insufficient.

| **V** | **Result** |
| --- | --- |
| V1 | Chronic ulcers (vascular, diabetic, pressure, post-traumatic, iatrogenic) |
| V2 | Group A: mean 3.85 cm^2^, Group B: mean 3.4 cm^2^ |
| V3 | Chronic ulcers for at least 3 months, average of 10 months |
| V4 | RCT |
| V5 | Category 2a |
| V6 | uESWT (EFD = 0.1 mJ/mm^2^) |
| V7 | uESWT (EFD = 0.04 mJ/mm^2^) |
| V8 | Group A: 62 patients (final data: 60); |
| V9 | Group B: 62 patients (final data: 40) |
| V10 | Group A: DermaGold (TRT, Alpharetta, GA, USA); Group B: Evotron (HMT, Lengwil, Switzerland) |
| V11 | Unfocused, electrohydraulic ESWT |
| V12 | Group A: 0.10 mJ/mm^2^; Group B: 0.04 mJ/mm^2^ |
| V13 | Not specified |
| V14 | 300 to 600 ESWs per session |
| V15 | Not explicitly provided, but determined by wound area and energy density |
| V16 | 4 Hz (240 ESWs per minute) |
| V17 | 7 sessions, once a week |
| V18 | 7 days |
| V19 | Reduction in wound area (percentage of healing) |
| V20 | After 7 weeks of treatment |
| V21 | Pain reduction (assessed via VAS), improvement in perilesional skin trophism, antibacterial effects; no biopsies or tissue samples analyzed |
| V22 | After 7 weeks of treatment |
| V23 | Not provided |
| V24 | Not applicable |
| V25 | Group A: 80% decrease in wound area; Group B: 67% decrease in wound area |
| V26 | Pain reduction: Group A: 79%, Group B: 48% (VAS scale) |
| V27 | Not provided |
| V28 | Not applicable |
| V29 | No bleeding or petechiae; however, 15 patients in group B withdrew because of pain during treatment |
| V30 | Yes |
| V31 | Yes |
| V32 | No |
| V33 | Yes |
| V34 | No |
| V35 | No |
| V36 | Yes |
| V37 | No |
| V38 | Yes |
| V39 | Yes |
| V40 | Yes |

**[87]	Wang, C.J.; Wu, C.T.; Yang, Y.J.; Liu, R.T.; Kuo, Y.R. Long-term outcomes of extracorporeal shockwave therapy for chronic foot ulcers. *J. Surg. Res.* 2014, *189*(2), 366–372. https://doi.org/10.1016/j.jss.2014.03.002.**

**Hypothesis:** This study tested the hypothesis that fESWT promotes long-term healing of chronic diabetic and nondiabetic foot ulcers, improves local blood flow perfusion and reduces morbidity over five years.

**Methods:** This prospective case series included 67 patients with 72 chronic foot ulcers of at least three months’ duration: 38 diabetic (small-vessel occlusion, peripheral neuropathy) and 29 nondiabetic (venous stasis, peripheral arterial disease). fESWT used the dermaPACE device (Sanuwave Health, Eden Prairie, MN, USA) in six sessions over three weeks (twice weekly), each delivering at least 500 ESWs or 8 ESWs/cm^2^ with an EFD of 0.11 mJ/mm^2^ at 4 Hz. No control group; comparisons used historical institutional controls. Primary endpoint: clinical improvement (complete healing or ≥50% wound reduction) at 1 year; secondary: blood flow perfusion (LDI), quality of life, amputation and mortality at 6 weeks, 1 year and 5 years.

**Results:** At 1 year, 55.6% of ulcers had healed and a further 27.8% showed ≥50% improvement, overall 83.4%, higher in non-diabetic (97%) than diabetic ulcers (73%). At 5 years, 57.4% remained healed and 4.7% maintained ≥50% improvement. Perfusion increased significantly in both groups at 6 weeks and 1 year (p = 0.011 and p = 0.033) but declined from years 1 to 5. The 5-year mortality was significantly higher in diabetic patients (24% vs. 3.5%; p = 0.035); amputation rates were also higher (17% vs. 3.6%), though not significantly. Quality of life favored the nondiabetic group and fESWT over historical HBOT controls (p < 0.001).

**Conclusions:** fESWT is an effective treatment for chronic foot ulcers, with substantial short- and intermediate-term benefits in healing and perfusion. Although effects diminished over five years, particularly in diabetic patients, healing, perfusion and quality of life outcomes were favorable, with few adverse events. Intermittent booster treatments warrant further research.

| **V** | **Result** |
| --- | --- |
| V1 | Chronic foot ulcers (diabetic and non-diabetic) |
| V2 | 9.1 ± 16.2 cm^2^ (range: 0.2–84) |
| V3 | Chronic; mean duration: 17.5 ± 18.6 months (range: 3–72) |
| V4 | Case series |
| V5 | Category 5 |
| V6 | fESWT + SWC |
| V7 | Not applicable |
| V8 | 67 patients (72 ulcers) |
| V9 | Not applicable |
| V10 | dermaPACE (Sanuwave Health, Eden Prairie, MN, USA) |
| V11 | Focused, electrohydraulic ESWT |
| V12 | 0.11 mJ/mm^2^ |
| V13 | Not specified |
| V14 | Treatment area (cm^2^) × 8; minimum 500 ESWs |
| V15 | Not explicitly stated as per cm^2^, but formula provided: ESWs = wound area × 8 |
| V16 | 4 Hz |
| V17 | 6 |
| V18 | Twice per week |
| V19 | Clinical improvement of ulcers (healing and >50% improvement rates) |
| V20 | 3 months, 1 year, 5 years |
| V21 | Local blood flow perfusion, quality of life, morbidity (amputation, mortality); no biopsies or tissue samples analyzed |
| V22 | 6 weeks, 1 year, 5 years |
| V23 | No |
| V24 | Not applicable |
| V25 | At 1 year: healed 55.6%, >50% improved 27.8%; at 5 years: healed 57.4%, >50% improved 4.7% |
| V26 | Perfusion increased at 6 weeks and 1 year (p = 0.011 diabetic, p = 0.033 non-diabetic); QoL favored fESWT over HBOT (p < 0.001) |
| V27 | No |
| V28 | Not applicable |
| V29 | 2 patients had transient burning sensation; no serious adverse events |
| V30 | Yes |
| V31 | No |
| V32 | No |
| V33 | Yes |
| V34 | No |
| V35 | No |
| V36 | No |
| V37 | Yes |
| V38 | No |
| V39 | No |
| V40 | Yes |

**[88]	Aschermann, I.; Noor, S.; Venturelli, S.; Sinnberg, T.; Mnich, C.D.; Busch, C. Extracorporal shock waves activate migration, proliferation and inflammatory pathways in fibroblasts and keratinocytes, and improve wound healing in an open-label, single-arm study in patients with therapy-refractory chronic leg ulcers. *Cell Physiol. Biochem.* 2017, *41*(3), 890–906. https://doi.org/10.1159/000460503.**

**Hypothesis:** This study tested the hypothesis that fESWT promotes healing of chronic leg ulcers by activating cell migration, proliferation and inflammatory pathways in fibroblasts and keratinocytes, improving clinical outcomes in therapy-refractory ulcers.

**Methods:** This prospective case series at a dermatological ulcer clinic included 60 patients with 75 therapy-refractory chronic leg ulcers of 3 months to 50 years’ duration and venous, arterial-venous (mixed) or rare etiologies such as calciphylaxis, PU and autoimmune-related conditions. All received fESWT adjunctive to SWC with a CellSonic device (CellSonic, Las Vegas, NV, USA); each session delivered 100 ESWs/cm^2^ plus 200 additional ESWs over the wound and its edges at an EFD of 0.136 mJ/mm^2^ and 4 Hz, totaling 250 to 2200 ESWs by wound size. Most patients received 4 sessions at 3–4 week intervals (mean 3.3 per ulcer). Primary outcome was the percentage reduction in wound size at the final follow-up visit (no change (<20%), improvement (20–75%), significant improvement (>75%) or complete healing (100%)) with no fixed time point indicated; secondary: tolerance and adverse events.

**Results:** Of the 75 ulcers, 31 (41%) showed complete healing, 12 (16%) significant improvement, 26 (35%) improvement and 6 (8%) no change; overall, 92% improved. The response was independent of ulcer etiology, duration, size or patient age. fESWT was generally well tolerated, although three of the 75 treated patients abandoned treatment because of treatment-induced pain, and was administered in routine outpatient care without complications.

**Conclusions:** fESWT is a safe and potentially effective treatment for chronic, therapy-refractory leg ulcers of various etiologies, with high rates of improvement or complete healing regardless of patient or ulcer characteristics, supporting its routine use as an adjunct to standard wound care. The authors emphasized that RCTs are necessary to validate efficacy and define the role of fESWT in chronic wound management.

| **V** | **Result** |
| --- | --- |
| V1 | Mixed (chronic ulcers and wounds of various etiologies) |
| V2 | Mean 8 cm^2^ (range 1–100 cm^2^) |
| V3 | Chronic, therapy-refractory; ulcer duration 3 months to 50 years (mean 5.9 ± 9.3 years) |
| V4 | Case series (open-label, single-arm study of consecutive routine-care patients; clinical data analyzed retrospectively) |
| V5 | Category 5 |
| V6 | fESWT + SWC |
| V7 | Not applicable |
| V8 | 60 |
| V9 | Not applicable |
| V10 | CellSonic (CellSonic, Las Vegas, NV, USA) |
| V11 | Focused, electrohydraulic ESWT |
| V12 | 0.136 mJ/mm^2^ (energy level 4) |
| V13 | Not specified |
| V14 | 100 impulses/cm^2^ + 200 impulses per session (mean 996 ± 562, range 250–2200; maximum 2200 for ulcers exceeding 20 cm^2^) |
| V15 | 100 impulses per cm^2^ (plus 200 additional impulses per session) |
| V16 | 4 Hz |
| V17 | Usually 4; mean 3.3 ± 1.2 per ulcer (11×1, 7×2, 14×3, 38×4, 4×5, 1×6 treatments) |
| V18 | Once every 3–4 weeks |
| V19 | Not explicitly defined |
| V20 | Not provided |
| V21 | Not explicitly defined; human foreskin from routine circumcision used for in vitro fibroblast and keratinocyte experiments |
| V22 | Not provided |
| V23 | No |
| V24 | Not applicable |
| V25 | Not applicable |
| V26 | Not applicable |
| V27 | No |
| V28 | Not applicable |
| V29 | Generally well tolerated; 3 of 75 patients abandoned treatment because of pain within seconds of starting |
| V30 | Yes |
| V31 | No |
| V32 | No |
| V33 | Not applicable |
| V34 | No |
| V35 | No |
| V36 | No |
| V37 | No |
| V38 | No |
| V39 | Not applicable |
| V40 | No |

**[28]	Snyder, R.; Galiano, R.; Mayer, P.; Rogers, L.C.; Alvarez, O.; Sanuwave Trial Investigators. Diabetic foot ulcer treatment with focused shockwave therapy: Two multicentre, prospective, controlled, double-blinded, randomised phase III clinical trials. *J. Wound Care* 2018, *27*(12), 822–836. https://doi.org/10.12968/jowc.2018.27.12.822.**

**Note: Snyder et al. (2018) analyzed pooled data from two separate RCTs; this summary addresses that pooled analysis.**

**Hypothesis:** This study tested the hypothesis that fESWT used adjunctively with SWC improves healing rates of chronic DFUs compared with SWC plus sham.

**Methods:** Two multicenter, randomized, sham-controlled, double-blinded phase III trials enrolled patients with chronic DFUs (University of Texas grade 1 or 2, stage A) from diabetic peripheral neuropathy, persisting ≥30 days and not reduced in volume by ≥50% during a 2-week SWC run-in. Randomization was to fESWT + SWC or sham + SWC. fESWT used the dermaPACE device (Sanuwave Health, Eden Prairie, MN, USA) at an EFD of 0.23 mJ/mm^2^ with 500 ESWs per session at 4 Hz: four sessions over two weeks (one every 3 ± 1 days) in the first study, up to eight over 12 weeks in the second, the same initial dosing continuing every two weeks. Primary endpoint: complete wound closure by week 12; secondary: time to closure, wound area reduction and adverse events to week 24.

**Results:** Of 336 patients (172 fESWT, 164 control), complete wound closure at 12 weeks did not differ significantly (22.7% vs. 18.3%; p = 0.32) but favored fESWT at 20 weeks (35.5% vs. 24.4%; p = 0.027) and 24 weeks (37.8% vs. 26.2%; p = 0.023). Secondary outcomes also favored fESWT: greater wound area reduction and ≥80% closure earlier. Treatment was well tolerated: no difference in treatment-emergent adverse events, significantly fewer serious adverse events (32.0% vs 43.3%; p = 0.042) and a trend toward fewer target-foot amputations (2.3% vs 6.4%; p = 0.065).

**Conclusions:** Adjunctive fESWT significantly enhanced the healing of hard-to-heal DFUs over time compared with SWC plus sham. It is safe, well-tolerated and clinically meaningful, particularly beyond 12 weeks, supporting use in chronic, non-healing DFUs.

| **V** | **Result** |
| --- | --- |
| V1 | Ulcers (DFUs) |
| V2 | 3.55 ± 3.1 cm^2^ (ESWT); 3.16 ± 2.5 cm^2^ (control) |
| V3 | Chronic (≥30 days); mean ulcer age: 47.2 ± 61.8 weeks (fESWT group), 61.7 ± 91.7 weeks (control group) |
| V4 | RCT |
| V5 | Category 1 |
| V6 | fESWT + SWC |
| V7 | Sham fESWT + SWC |
| V8 | 172 |
| V9 | 164 |
| V10 | dermaPACE (Sanuwave Health, Eden Prairie, MN, USA) |
| V11 | Focused, electrohydraulic ESWT |
| V12 | 0.23 mJ/mm^2^ |
| V13 | Not specified |
| V14 | 500 |
| V15 | Not provided |
| V16 | 4 Hz |
| V17 | 4 sessions over 2 weeks (study 1); up to 8 sessions over 12 weeks (study 2) |
| V18 | Study 1: every 3 ± 1 days; study 2: four every 3 ± 2 days, then 2-weekly |
| V19 | Complete wound closure |
| V20 | 12 weeks |
| V21 | Time to closure, percentage wound closure; no biopsies or tissue samples analyzed |
| V22 | 24 weeks |
| V23 | Yes |
| V24 | Study 1: 200, 75/group, 90% power (α=0.05), 55% vs 29% closure; study 2: 200, Bayesian (study 1 prior) |
| V25 | 35.5% closure (fESWT group) vs 24.4% (control group) at 20 weeks post-baseline (p=0.027) |
| V26 | Greater area reduction weeks 6–24; 80% reduction significant week 14; 24-week closure 37.8% vs 26.2%, not 20% |
| V27 | No |
| V28 | Not applicable |
| V29 | Device-related AEs 9/172 (5.2%) fESWT vs 4/164 (2.4%) control (p = 0.259); serious AEs burning sensation, pain, headache (n=2 each) |
| V30 | Yes |
| V31 | Yes |
| V32 | Yes |
| V33 | Yes |
| V34 | Yes |
| V35 | No |
| V36 | Yes |
| V37 | Yes |
| V38 | Yes |
| V39 | Yes |
| V40 | Yes |

**[28]	Snyder, R.; Galiano, R.; Mayer, P.; Rogers, L.C.; Alvarez, O.; Sanuwave Trial Investigators. Diabetic foot ulcer treatment with focused shockwave therapy: Two multicentre, prospective, controlled, double-blinded, randomised phase III clinical trials. *J. Wound Care* 2018, *27*(12), 822–836. https://doi.org/10.12968/jowc.2018.27.12.822.**

**Note: Snyder et al. (2018) analyzed pooled data from two separate RCTs; this summary addresses the data of the first RCT.**

**Hypothesis:** This study tested the hypothesis that fESWT, as an adjunct to SWC, improves the rate of complete healing in chronic DFUs compared to SWC with sham treatment.

**Methods:** Study 1 was a multicenter, randomized, sham-controlled, double-blinded phase III trial of fESWT in chronic DFUs resulting from peripheral neuropathy. Eligible ulcers were grade 1 or 2, stage A (University of Texas Diabetic Wound Classification), of at least 30 days’ duration (no maximum specified), ≥1.0 cm^2^ and ≤16 cm^2^. Subjects were randomized to fESWT + SWC (n=107) or SWC plus sham (n=99). fESWT used a dermaPACE device (Sanuwave Health, Eden Prairie, MN, USA) at EFD of 0.23 mJ/mm^2^, with 500 ESWs per session at 4 Hz, in four sessions over two weeks, every 3 ± 1 days. Primary outcome was complete wound closure by 12 weeks, confirmed at two consecutive visits; secondary: wound area and volume reduction, time to closure, adverse events and recurrence to 24 weeks post-baseline.

**Results:** At 12 weeks, 20.6% of the fESWT group and 15.2% of the sham group achieved complete wound closure (p = 0.363). At 20 weeks, significantly more fESWT patients achieved complete closure (36.4% vs. 23.2%; p = 0.047); the trend persisted at 24 weeks (39.3% vs. 26.3%; p = 0.054). Secondary outcomes (wound area reduction, amputation rates) were reported only for the pooled dataset, not separately for study 1. No significant differences in adverse event rates were observed; fESWT was well tolerated.

**Conclusions:** Study 1 demonstrated that adjunctive fESWT significantly improved long-term healing outcomes in chronic DFUs compared to sham treatment, with the most pronounced effects after 20 weeks and without increased risk of adverse events.

| **V** | **Result** |
| --- | --- |
| V1 | Ulcer (diabetic foot ulcer) |
| V2 | 3.5 ± 3.2 cm^2^ (fESWT); 2.8 ± 2.4 cm^2^ (sham) |
| V3 | Chronic (≥30 days); mean ulcer age: 48.7 ± 66.6 weeks (ESWT group), 69.5 ± 107.5 weeks (control group) |
| V4 | RCT |
| V5 | Category 1 |
| V6 | fESWT + SWC |
| V7 | Sham fESWT + SWC |
| V8 | 107 |
| V9 | 99 |
| V10 | dermaPACE (Sanuwave Health, Eden Prairie, MN, USA) |
| V11 | Focused, electrohydraulic ESWT |
| V12 | 0.23 mJ/mm^2^ |
| V13 | Not specified |
| V14 | 500 |
| V15 | Not provided |
| V16 | 4 Hz |
| V17 | 4 |
| V18 | Every 3 ± 1 days |
| V19 | Complete wound closure confirmed at two consecutive visits |
| V20 | 12 weeks |
| V21 | Wound area, volume, depth, perimeter, recurrence, amputation, pain (VAS), infection; no biopsies or tissue samples analyzed |
| V22 | Up to 24 weeks |
| V23 | Yes |
| V24 | Target: 200 subjects to detect 26% difference with 90% power |
| V25 | 12 weeks: 20.6% (22/107) vs 15.2% (15/99), p = 0.363; 20 weeks: 36.4% (39/107) vs 23.2% (23/99), p = 0.047 |
| V26 | Area reduction also greater in fESWT group |
| V27 | Not reported for Study 1 separately |
| V28 | Not provided |
| V29 | No significant differences; well tolerated |
| V30 | Yes |
| V31 | Yes |
| V32 | Yes |
| V33 | Yes |
| V34 | Yes |
| V35 | No |
| V36 | Yes |
| V37 | No |
| V38 | Yes |
| V39 | Yes |
| V40 | Yes |

**[28]	Snyder, R.; Galiano, R.; Mayer, P.; Rogers, L.C.; Alvarez, O.; Sanuwave Trial Investigators. Diabetic foot ulcer treatment with focused shockwave therapy: Two multicentre, prospective, controlled, double-blinded, randomised phase III clinical trials. *J. Wound Care* 2018, *27*(12), 822–836. https://doi.org/10.12968/jowc.2018.27.12.822.**

**Note: Snyder et al. (2018) analyzed pooled data from two separate RCTs; this summary addresses the data of the second RCT.**

**Hypothesis:** This study tested the hypothesis that increasing the number of fESWT sessions enhances healing of chronic DFUs compared to SWC with sham treatment.

**Methods:** Study 2 was a multicenter, randomized, sham-controlled, double-blinded phase III trial of adjunctive fESWT in chronic DFUs from diabetic peripheral neuropathy. Eligible ulcers were grade 1 or 2, stage A (University of Texas classification), ≥30 days’ duration, 1.0–16.0 cm^2^. Of 130 patients, 65 received ESWT and 65 sham alongside SWC. fESWT used a dermaPACE device (Sanuwave Health, Eden Prairie, MN, USA) at EFD of 0.23 mJ/mm^2^, with 500 ESWs per session at 4 Hz. Up to 8 sessions over 12 weeks: first four every 3 ± 2 days, remaining four every 2 weeks. Primary endpoint was complete wound closure by week 12 (blinded assessors); secondary: wound area, depth and volume reduction, amputations and adverse events to week 24.

**Results:** 26.2% of the ESWT group and 23.1% of the sham group achieved complete closure by week 12 (p = 0.684). By week 24, closure rates were 35.4% for ESWT and 26.2% for sham (p = 0.254), not statistically significant, though trends consistently favored fESWT. The fESWT group showed greater mean reductions in wound area and volume. No significant differences in adverse events were observed.

**Conclusions:** Study 2 missed significance in its primary endpoint; increasing applications from four to eight did not improve outcomes (closure at 24 weeks was in fact lower than in study 1 (35.4% vs 39.3%) with identical sham results (26.2% vs 26.3%)) and the authors attributed the absent dose effect to the dose not being scaled to wound size. Treatment was nonetheless well tolerated.

| **V** | **Result** |
| --- | --- |
| V1 | DFUs |
| V2 | 3.7 ± 2.8 cm^2^ (fESWT); 3.7 ± 2.8 cm^2^ (sham) |
| V3 | Chronic (≥30 days); mean ulcer age: 44.6 ± 53.4 weeks (fESWT group), 49.7 ± 59.2 weeks (control group) |
| V4 | RCT |
| V5 | Category 2b |
| V6 | fESWT + SWC |
| V7 | Sham fESWT + SWC |
| V8 | 65 |
| V9 | 65 |
| V10 | dermaPACE (Sanuwave Health, Eden Prairie, MN, USA) |
| V11 | Focused, electrohydraulic ESWT |
| V12 | 0.23 mJ/mm^2^ |
| V13 | Not specified |
| V14 | 500 |
| V15 | Not provided |
| V16 | 4 Hz |
| V17 | Up to 8 |
| V18 | First 4 every 3 ± 2 days, then every 2 weeks |
| V19 | Complete wound closure confirmed by visual and photographic evaluation |
| V20 | 12 weeks |
| V21 | Wound size metrics, recurrence, amputation, infection, adverse events; no biopsies or tissue samples analyzed |
| V22 | Up to 24 weeks |
| V23 | Yes |
| V24 | Powered to detect HR > 1; Bayesian design with informative prior from Study 1 |
| V25 | fESWT: 26.2% (17/65); sham: 23.1% (15/65); p = 0.684 |
| V26 | Non-significant trends favoring fESWT in wound size reduction and amputation rates |
| V27 | Not reported for Study 2 separately |
| V28 | Not provided |
| V29 | No significant differences; well tolerated |
| V30 | Yes |
| V31 | Yes |
| V32 | Yes |
| V33 | Yes |
| V34 | Yes |
| V35 | No |
| V36 | Yes |
| V37 | Yes |
| V38 | Yes |
| V39 | Yes |
| V40 | Yes |

**[89]	Chou, W.Y.; Wang, C.J.; Cheng, J.H.; Chen, J.H.; Chen, C.C.; Kuo, Y.R. Extended extracorporeal shockwave therapy for chronic diabetic foot ulcers: A case series. *Wounds* 2019, *31*(5), 132–136.**

**Hypothesis:** This study tested the hypothesis that extended and repeated application of fESWT helps maintaining the therapeutic effects of fESWT and improves long-term outcomes in patients with chronic DFUs.

**Methods:** This case series included four patients with chronic DFUs (non-healing ulcers of more than three months’ duration in diabetes), all Wagner grade II or less, without a control group. All received fESWT adjunctive to SWC (dressings, glucose control, offloading) with a dermaPACE device (Sanuwave Health, Eden Prairie, MN, USA) at an EFD of 0.11 mJ/mm^2^ and 4 Hz. Each session involved at least 500 ESWs, calculated as 8 ESWs/cm^2^ of wound area; three patients received 6 sessions and one 12, twice weekly. Primary outcome: clinical healing; secondary: blood flow perfusion, immunohistochemical markers of angiogenesis, inflammation and tissue regeneration, and the Diabetic Foot Ulcer Scale-Short Form, all assessed at 48 weeks post-baseline.

**Results:** At 48 weeks post-baseline, two patients exhibited complete healing and two showed partial improvement (86% and 40% reduction in ulcer size, respectively). One patient showed improved blood flow perfusion, the others no significant change. Immunohistochemistry showed elevated angiogenic and tissue repair biomarkers in three patients, decreased in the fourth. The Diabetic Foot Ulcer Scale Short-Form score decreased in one patient and was unchanged in three at 48 weeks; from immediately post-fESWT to 48 weeks, patient 3 showed a worse score. No serious adverse events occurred; mild local redness or swelling resolved with conservative measures.

**Conclusions:** fESWT may promote healing in chronic DFUs, and extended, intermittent treatment may help sustain the benefits. Although limited by the small sample and lack of a control group, the series indicated a possible contribution to long-term wound healing and tissue viability in chronic DFUs. Further RCTs are warranted to validate these results and optimize treatment protocols.

| **V** | **Result** |
| --- | --- |
| V1 | Chronic DFUs |
| V2 | Not provided |
| V3 | Chronic, >3 months |
| V4 | Case series |
| V5 | Category 5 |
| V6 | fESWT + SWC |
| V7 | Not applicable |
| V8 | 4 |
| V9 | Not applicable |
| V10 | dermaPACE (Sanuwave Health, Eden Prairie, MN, USA) |
| V11 | Focused, electrohydraulic ESWT |
| V12 | 0.11 mJ/mm^2^ |
| V13 | Not specified |
| V14 | ≥500 |
| V15 | 8/cm^2^ |
| V16 | 4 Hz |
| V17 | 6 (3 patients), 12 (1 patient) |
| V18 | Twice weekly |
| V19 | Ulcer healing, perfusion, histological markers |
| V20 | 48 weeks post-baseline |
| V21 | DFU scale, perfusion; biopsies taken from the ulcers were analyzed using histology and immunohistochemistry |
| V22 | 48 weeks |
| V23 | No |
| V24 | Not applicable |
| V25 | 2 healed, 2 improved |
| V26 | Mixed results |
| V27 | No |
| V28 | Not applicable |
| V29 | Mild redness/swelling; no systemic issues |
| V30 | Yes |
| V31 | No |
| V32 | No |
| V33 | Not applicable |
| V34 | No |
| V35 | No |
| V36 | No |
| V37 | Yes |
| V38 | Yes |
| V39 | Not applicable |
| V40 | No |

**[90]	Galiano, R.; Snyder, R.; Mayer, P.; Rogers, L.C.; Alvarez, O.; Sanuwave Trial Investigators. Focused shockwave therapy in diabetic foot ulcers: Secondary endpoints of two multicentre randomised controlled trials. *J. Wound Care* 2019, *28*(6), 383–395. https://doi.org/10.12968/jowc.2019.28.6.383.**

**Hypothesis:** This study tested the hypothesis that adjunctive fESWT leads to superior healing outcomes compared to sham in patients with chronic neuropathic DFUs unresponsive to SWC.

**Methods:** Pooled analysis of two multicenter, randomized, double-blind, sham-controlled trials (Snyder et al., 2018) in chronic (≥30 days), non-ischemic, neuropathic DFUs (grade 1A or 2A, University of Texas classification). After a 2-week SWC run-in, subjects whose wounds had not reduced by ≥50% were randomized to fESWT or sham with continued SWC. fESWT used a dermaPACE device (Sanuwave Health, Eden Prairie, MN, USA) at EFD of 0.23 mJ/mm^2^, with 500 ESWs per session at 4 Hz; study 1 gave four sessions over two weeks, study 2 up to eight over 12 weeks. Primary outcome: complete wound closure (100% epithelialization, no drainage) at 12 weeks; secondary: wound area change, time to closure, infection, recurrence and adverse events to 24 weeks.

**Results:** Of 336 patients (172 fESWT, 164 control), complete closure at 12 weeks did not differ significantly in the pooled set (22.7% vs 18.3%), reaching significance at 20 weeks (35.5% vs 24.4%, p = 0.027) and 24 weeks (37.8% vs 26.2%, p = 0.023). (These primary-endpoint data came from Snyder et al., 2018; the present paper reported secondary endpoints only.) The secondary-endpoint benefit applied to study 1 only; in study 2 no wound-closure-related secondary endpoint differed significantly, and these were not analyzed in the pooled set. Time to 25% closure was significantly shorter with fESWT (84 days vs. 112 days, p = 0.035). Adverse events, including infections and procedural pain, were comparable; no serious treatment-related events.

**Conclusions:** Adjunctive fESWT significantly improves healing in chronic DFUs unresponsive to SWC alone, with higher closure rates and faster healing and no increase in adverse events, supporting it as an advanced therapy for recalcitrant DFUs.

| **V** | **Result** |
| --- | --- |
| V1 | Ulcers (DFUs) |
| V2 | Study 1 baseline: area 3.5/2.8 cm^2^, perimeter 6.9/6.4 cm (fESWT/sham); inclusion 1-16 cm^2^; study 2/pooled unreported |
| V3 | Chronic (minimum 30 days duration) |
| V4 | RCT |
| V5 | Category 1 |
| V6 | fESWT + SWC |
| V7 | sham fESWT + SWC |
| V8 | 172 |
| V9 | 164 |
| V10 | dermaPACE (Sanuwave Health, Eden Prairie, MN, USA) |
| V11 | Focused, electrohydraulic ESWT |
| V12 | 0.23 mJ/mm^2^ |
| V13 | Not specified |
| V14 | 500 |
| V15 | Not provided |
| V16 | 4 Hz |
| V17 | Study 1: 4 sessions over the 2-week treatment phase; study 2: up to 8 sessions over the 12-week treatment phase |
| V18 | Study 1: 4 applications every 3 +/- 1 days over 2 weeks; study 2: 4 every 3 +/- 2 days, then 2-weekly |
| V19 | Complete wound closure (100% epithelialization with no drainage) |
| V20 | 12 weeks after first treatment |
| V21 | Percent change in wound area; time to closure; no biopsies or tissue samples analyzed |
| V22 | Efficacy-related secondary endpoints were measured at 12, 20 and 24 weeks (study visits every 14 +/- 2 days) |
| V23 | Yes |
| V24 | Target 200 randomized 1:1; assumed 55% vs 29% closure; 75 per group for 90% power (alpha 0.05) |
| V25 | Not the topic of this study |
| V26 | Study 1: area reduction 48.6% vs 10.7% (p = 0.015), perimeter 46.4% vs 25.0% (p = 0.022); study 2/pooled: none significant |
| V27 | No |
| V28 | Not applicable |
| V29 | Mild, transient pain; no serious treatment-related adverse events |
| V30 | Yes |
| V31 | Yes |
| V32 | Yes |
| V33 | Yes |
| V34 | Yes |
| V35 | No |
| V36 | Yes |
| V37 | No |
| V38 | Yes |
| V39 | Yes |
| V40 | Yes |

**[91]	Holsapple, J.S.; Cooper, B.; Berry, S.H.; Staniszewska, A.; Dickson, B.M.; Taylor, J.A.; Bachoo, P.; Wilson, H.M. Low intensity shockwave treatment modulates macrophage functions beneficial to healing chronic wounds. *Int. J. Mol. Sci.* 2021, *22*(15), 7844. https://doi.org/10.3390/ijms22157844.**

**Hypothesis:** This study tested the hypothesis that fESWT improves healing outcomes in chronic VLUs by modulating macrophage function and promoting tissue regeneration.

**Methods:** This case series investigated fESWT in chronic VLUs in ten patients, nine of whom completed the study. The treatment-resistant ulcers had shown no healing for at least eight weeks before intervention; there was no control group. fESWT used a DermaGold device (TRT, Alpharetta, GA, USA) at an EFD of 0.11 mJ/mm^2^ and 4 Hz. Each patient received at least 500 ESWs per session, tailored to wound area at 8 ESWs/cm^2^, in up to 6 sessions (initially 3, plus 3 more if not fully healed), biweekly. Primary outcome was the change in wound area; secondary: angiogenesis (CD31), smooth muscle cell (SMC) actin, cell proliferation (Ki67), macrophage abundance and activation – all assessed 2 weeks after the first fESWT session.

**Results:** fESWT improved healing in 7 of the 9 patients who completed the study, with mean wound area reduced from 93.5 ± 38.6 cm^2^ to 88.2 ± 37.4 cm^2^ (p = 0.051) and a median wound size reduction of 11% (interquartile range 1.5–22.5%). Angiogenesis increased significantly in most patients (elevated CD31 staining, p = 0.0391). Macrophage counts per wound biopsy area significantly decreased (p = 0.048), suggesting resolution of chronic inflammation. Ki67 and SMC actin increased in some patients but did not reach statistical significance. Macrophage activation varied, with an overall trend toward increased activation, not exclusively toward M1 or M2 phenotypes.

**Conclusions:** fESWT may support healing in chronic VLUs by reducing macrophage infiltration and promoting angiogenesis, suggesting modulation of immune and regenerative processes in wounds that have failed SWC. Although the absence of a control group limited conclusions, the observed biological effects and clinical improvements warrant further controlled trials.

| **V** | **Result** |
| --- | --- |
| V1 | Chronic VLUs |
| V2 | 93.5 cm^2^ pre-treatment |
| V3 | Chronic; non-healing for ≥8 weeks |
| V4 | Case series |
| V5 | Category 5 |
| V6 | fESWT + SWC |
| V7 | Not applicable |
| V8 | 9 (after exclusion of one) |
| V9 | Not applicable |
| V10 | DermaGold (TRT, Alpharetta, GA, USA) |
| V11 | Focused, electrohydraulic ESWT |
| V12 | 0.11 mJ/mm^2^ |
| V13 | Not specified |
| V14 | Minimum 500 ESWs |
| V15 | 8 ESWs/cm^2^ |
| V16 | 4 Hz |
| V17 | Up to 6 sessions (3 initially, 3 more if not healed) |
| V18 | Every 2 weeks |
| V19 | Change in wound area size |
| V20 | 2 weeks post-treatment |
| V21 | Biopsies of ulcers collected both immediately before and two weeks post-treatment with fESWT, analyzed using histology and immunohistochemistry |
| V22 | 2 weeks post-treatment |
| V23 | No |
| V24 | Not applicable |
| V25 | Wound area reduced in 7/9 patients; mean reduction from 93.5 to 88.2 cm^2^ (p = 0.051) |
| V26 | CD31 increased (p = 0.039); macrophage counts decreased (p = 0.048); others not statistically significant |
| V27 | No |
| V28 | Not applicable |
| V29 | Not reported |
| V30 | Yes |
| V31 | No |
| V32 | No |
| V33 | Not applicable |
| V34 | No |
| V35 | No |
| V36 | No |
| V37 | Yes |
| V38 | Yes |
| V39 | Not applicable |
| V40 | Yes |

**[92]	Jeong, D.; Lee, J.H.; Lee, G.B.; Shin, K.H.; Hwang, J.; Jang, S.Y.; Yoo, J.; Jang, W.Y. Application of extracorporeal shockwave therapy to improve microcirculation in diabetic foot ulcers: A prospective study. *Medicine* 2023, *102*(11), e33310. https://doi.org/10.1097/MD.0000000000033310.**

**Hypothesis:** This study tested the hypothesis that fESWT significantly improves microcirculation, as measured by TcPO₂, in patients with DFUs after a three-week treatment period.

**Methods:** This prospective cohort study included 25 patients with type 2 diabetes mellitus and Wagner grade I or II DFUs from peripheral arterial disease and neuropathy. The ulcerated feet (n = 32) formed the ESWT group, the unaffected contralateral feet (n = 18) the controls. fESWT used an orthoPACE device (Sanuwave Health, Eden Prairie, MN, USA) at an EFD of 0.2 mJ/mm2 and 4 Hz. Each session consisted of 1500 ESWs, given three times per week for three weeks (9 sessions); the interval between sessions is not stated. Primary outcome: TcPO₂, measured weekly, with recovery to ≥43 mm Hg as endpoint, assessed at baseline and weeks 1, 2 and 3 post-baseline. Secondary: subgroup analysis of Wagner grade I versus grade II ulcers.

**Results:** TcPO₂ improved significantly in the ESWT group from the second week (baseline 34.3 ± 3.9 mm Hg vs week 2 post-baseline 43.5 ± 3.5 mm Hg, p = 0.003; week 3 post-baseline 45.3 ± 3.6 mm Hg, p = 0.001), whereas controls showed no significant change. Between-group differences were significant at all points: baseline (p = 0.003), week 1 (p = 0.015), week 2 (p = 0.014), week 3 post-baseline (p = 0.032). Both Wagner grades responded favorably, TcPO₂ surpassing the 43 mm Hg threshold by the second week post-baseline. No treatment-related adverse events were reported.

**Conclusions:** fESWT significantly improves microcirculation in chronic DFUs within two weeks, raising TcPO₂ above the prognostic threshold of 43 mm Hg. It was well tolerated and effective in mild and moderate DFUs regardless of Wagner grade, supporting fESWT as a promising adjunctive therapy for vascularization in DFUs.

| **V** | **Result** |
| --- | --- |
| V1 | DFUs |
| V2 | Mean size not provided; inclusion criteria: ulcer < 2.5 cm in diameter |
| V3 | Chronic; DFU duration criteria: ulcer present ≥30 days before enrollment |
| V4 | Prospective cohort study (non-randomized, with intra-patient control) |
| V5 | Category 1 |
| V6 | fESWT + SWC |
| V7 | No treatment (contralateral unaffected foot used as control) |
| V8 | 32 feet |
| V9 | 18 feet |
| V10 | orthoPACE (Sanuwave Health, Eden Prairie, MN, USA) |
| V11 | Focused, electrohydraulic ESWT |
| V12 | 0.2 mJ/mm^2^ |
| V13 | Not specified |
| V14 | 1500 ESWs |
| V15 | Not provided |
| V16 | 4 Hz |
| V17 | 9 sessions |
| V18 | 3 sessions per week |
| V19 | Increase in TcPO₂ to ≥43 mm Hg |
| V20 | Weekly assessments; significance observed at week 2 post-baseline |
| V21 | Subgroup analysis by Wagner grade (I vs II); no biopsies or tissue samples analyzed |
| V22 | Weekly for 3 weeks |
| V23 | Yes |
| V24 | Based on α = 0.05, power = 0.8, minimum 14 feet in ESWT group required |
| V25 | ESWT: TcPO₂ 34.3 ± 3.9 → 45.3 ± 3.6 mm Hg at week 3 (p = 0.001); control: no significant change |
| V26 | Wagner I: TcPO₂ 34.0 ± 5.9 → 45.5 ± 5.4 mm Hg (p = 0.029); Wagner II: 36.9 ± 5.4 → 44.2 ± 5.2 (p = 0.009) |
| V27 | No |
| V28 | Not applicable |
| V29 | No treatment-related adverse events observed |
| V30 | Yes |
| V31 | No |
| V32 | No |
| V33 | No |
| V34 | No |
| V35 | No |
| V36 | No |
| V37 | Yes |
| V38 | Yes |
| V39 | Yes |
| V40 | Yes |

**[93]	Nemeth, D.; Shah, J. Extracorporeal shockwave therapy (ESWT) in an outpatient wound care clinic: Case series analysis of a non-invasive technology in the management of chronic wounds for wound bed preparation. *Wound Manag. Prev.* 2024, *70*(2). https://doi.org/10.25270/wmp.22090.**

**Hypothesis:** This study tested the hypothesis that fESWT improves healing outcomes in patients with complex chronic wounds that were refractory to SWC.

**Methods:** This retrospective case series in a medically underserved outpatient wound care clinic covered complex chronic wounds (DFUs, surgical dehiscence, flap necrosis) unhealed after more than 30 days of SWC. Thirteen patients with 18 wounds were enrolled; after exclusions, 10 patients with 13 wounds were analyzed. All received fESWT adjunctive to SWC with a dermaPACE device (Sanuwave Health, Eden Prairie, MN, USA); EFD and frequency in Hertz were not reported. fESWT was applied weekly, ESWs per session tailored to wound size per manufacturer guidelines, for a maximum recorded 12 weeks; the one wound that did not close received 17 treatments. Primary outcome was complete wound closure to 12 weeks post-baseline; secondary: wound dimension reduction and correlations of healing time with wound volume or tendon/bone involvement.

**Results:** Of the 13 wounds, 12 achieved complete closure within 12 weeks, with a mean of 6.8 treatment sessions (range 3–11). One wound did not fully close but shrank substantially and healed shortly after a biological graft, suggesting effective wound bed preparation. A priori power analysis required a sample size of 67; the actual 13 wounds were therefore underpowered. No significant correlation was found between initial wound volume or tendon/bone involvement and weeks to closure. No adverse events were reported.

**Conclusions:** This case series suggests that fESWT may be a safe, non-invasive and potentially effective adjunct for chronic wounds in outpatient settings, particularly in patients with multiple comorbidities and limited healthcare access. Despite the small sample and lack of statistical significance, the high healing rate supports the utility of fESWT in wound bed preparation and chronic wound management; larger controlled studies are warranted.

| **V** | **Result** |
| --- | --- |
| V1 | Chronic lower extremity wounds (including DFUs, flap necrosis and surgical dehiscence) |
| V2 | Mean initial wound volume 1.7 cm^3^ (SD 3.5, range 0.02-13); diabetic subcohort 0.7 cm^3^ (SD 0.9, range 0.02-2.2) |
| V3 | Chronic (>30 days refractory to SWC) |
| V4 | Case series |
| V5 | Category 5 |
| V6 | fESWT + SWC |
| V7 | not applicable |
| V8 | 10 patients (13 wounds) |
| V9 | not applicable |
| V10 | dermaPACE (Sanuwave Health, Eden Prairie, MN, USA) |
| V11 | Focused, electrohydraulic ESWT |
| V12 | Not provided |
| V13 | Not specified |
| V14 | Variable, based on wound size; specific values not provided |
| V15 | Not provided |
| V16 | Not provided |
| V17 | Mean 6.8 weeks to closure (SD 2.8; range 3-11); diabetic cohort 7 (SD 2.8); non-healing wound 17 treatments |
| V18 | Weekly intervals |
| V19 | Complete wound healing (wound closure) |
| V20 | Up to 12 weeks |
| V21 | Wound dimension reduction, correlation of healing time with wound volume, tendon/bone involvement; no biopsies or tissue samples analyzed |
| V22 | Within 12 weeks |
| V23 | Yes |
| V24 | Sample size needed = 67; study sample = 13 wounds |
| V25 | 12 of 13 wounds healed completely by week 12; 1 wound improved but not healed, no statistical significance |
| V26 | No significant correlation between wound volume and healing duration; trend toward faster healing with tendon/bone involvement (non-significant) |
| V27 | No |
| V28 | not applicable |
| V29 | No adverse events reported |
| V30 | Yes |
| V31 | No |
| V32 | No |
| V33 | Not applicable |
| V34 | No |
| V35 | No |
| V36 | No |
| V37 | No |
| V38 | Yes |
| V39 | Not applicable |
| V40 | Yes |

**[94]	Marzella, L.; Riccio, M.; D’Agostino, M.C.; Lazzerini, A.; De Francesco, F. Shock wave-induced regeneration in soft tissue reconstruction: Clinical application in hand surgery. Surgeries 2026, 7(1), 4. https://doi.org/10.3390/surgeries7010004.**

**Hypothesis:** This study tested the hypothesis that fESWT added to SWC acts as a regenerative rather than symptomatic treatment, improving healing, pain, inflammation, microvascular remodeling, sensory recovery and mobility in hand soft tissue defects, with nailfold videocapillaroscopy tracking functional recovery.

**Methods:** This prospective single-arm case series included 64 patients with hand soft tissue defects; mean time since injury or surgery 6.2 ± 3.4 weeks, so defects were predominantly subacute, although the abstract and conclusions called them chronic ulcers. All received fESWT added to SWC, without a control group, using an Orthogold 100 device (MTS Medical, Konstanz, Germany) at an EFD of 0.10 mJ/mm^2^ with 350–1000 impulses per cm^2^ per session; frequency and absolute ESW number were not reported. Sessions were weekly, three to six (mean 4.3 ± 1.2); no single primary endpoint was designated.

**Results:** All 64 patients completed treatment and follow-up. From baseline to end of treatment BWAT fell from 45.1 ± 3.9 to 17.2 ± 3.0 (p < 0.01), capillaroscopy rose 3.6 ± 0.8 to 7.7 ± 0.6 (p < 0.05) and ROM recovery from 40.2 ± 9.7% to 94.7 ± 5.3% (p < 0.001); VAS, CEA, IGA-I, SWMT and 2PD improved likewise. Complete closure took 4.5 ± 1.1 weeks; diabetics and smokers healed more slowly (5.8 ± 1.3 vs. 4.2 ± 0.9 weeks, p = 0.03). Capillaroscopy correlated with BWAT (ρ = −0.64) and ROM (ρ = 0.58, p < 0.01). No adverse events were recorded.

**Conclusions:** fESWT added to routine wound care was associated with significant improvements in wound status, pain, inflammation, microvascular architecture, sensory function and mobility. The authors emphasized that the study was not designed to show superiority over conventional techniques; without a control group and with short follow-up, benefits cannot be attributed to fESWT alone, and RCTs are required.

| **V** | **Result** |
| --- | --- |
| V1 | Hand defects: fingertip amputation/skin loss 46 (72%), flap necrosis 8 (13%), postoperative 5 (8%), chronic ulcer/scar 5 (8%) |
| V2 | Mean wound area at baseline 2.4 ± 1.1 cm^2^ (range 0.8–5.0) |
| V3 | Mixed; 6.2 ± 3.4 weeks since injury (range 3–16) |
| V4 | Case series |
| V5 | Category 5 |
| V6 | fESWT + SWC |
| V7 | Not applicable |
| V8 | 64 |
| V9 | Not applicable |
| V10 | Orthogold 100 (MTS Medical, Konstanz, Germany) |
| V11 | Focused, electrohydraulic ESWT |
| V12 | 0.10 mJ/mm^2^ |
| V13 | Not specified |
| V14 | 350–1000 ESWs/cm^2^ per session |
| V15 | 350–1000 ESWs/cm^2^ per session |
| V16 | Not provided |
| V17 | 3–6 sessions (mean 4.3 ± 1.2) |
| V18 | One session per week |
| V19 | No single primary endpoint; VAS, CEA, IGA-I, BWAT, capillaroscopy score, SWMT, 2PD and ROM recovery all primary |
| V20 | Baseline, after 3 sessions, end of treatment, 8 weeks after treatment (also 1, 2 and 4 weeks) |
| V21 | Not designated; time to closure, smokers/diabetics vs. non-smokers/non-diabetics subgroups, capillaroscopy correlations with BWAT, SWMT, 2PD, ROM; no biopsies |
| V22 | Same as primary: baseline, after 3 sessions, end of treatment, 8-week follow-up; capillaroscopy only to end of treatment |
| V23 | No |
| V24 | Not applicable |
| V25 | VAS 5.9 ± 0.7→0.8 ± 0.5 (p < 0.001); BWAT 45.1 ± 3.9→17.2 ± 3.0; CEA 2.6 ± 0.5→0.6 ± 0.5; IGA-I 3.4 ± 0.6→0.8 ± 0.4 (p < 0.01); capillaroscopy 3.6 ± 0.8→7.7 ± 0.6; SWMT 5.12 ± 0.7→3.61 ± 0.6; 2PD 12.8 ± 2.3→6.4 ± 1.8 mm (p < 0.05) |
| V26 | Diabetics/smokers vs others: healing 5.8 ± 1.3/4.2 ± 0.9 weeks (p = 0.03); VAS 1.3 ± 0.6/0.8 ± 0.4 (p = 0.02); BWAT 20.2 ± 3.1/17.0 ± 2.8 (p = 0.03); values converged by week 8 |
| V27 | No |
| V28 | Not applicable |
| V29 | No adverse events reported; no skin breakdown, hematoma or infection and no treatment discontinuation |
| V30 | Yes |
| V31 | No |
| V32 | No |
| V33 | Not applicable |
| V34 | No |
| V35 | No |
| V36 | No |
| V37 | Yes |
| V38 | Yes |
| V39 | Not applicable |
| V40 | Yes |
