## Supplementary File 3 for "Extracorporeal Shock Wave Therapy for Wound Management: Clinical Evidence, Energy Delivery Parameters and Mechanistic Insights — A Systematic Review"

**Extracorporeal Shock Wave Therapy in Wound Management: A Comprehensive Systematic Review of Clinical Evidence, Modalities and Mechanisms**

by Brent Musolf, Carmen Nussbaum-Krammer, Michael O’Neal, Nicola Maffulli and Christoph Schmitz

**Supplementary File 3**

**Standardized summaries and 40 key variables (c.f. Table 2 in the main text) from clinical studies on electromagnetic ESWT for wound management**

**(numbers in brackets refer to the reference numbers in the main text)**

**Note on the standardized summaries:** Each summary condenses one clinical study as it was reported, together with the interpretation that its own authors placed on their findings. The summaries therefore reproduce the position of the respective authors and the state of the field at the time of that publication, and not the assessment of the present systematic review. They follow a uniform structure (Hypothesis, Methods, Results and Conclusions), because the original abstracts differ widely in structure, in length and in the information they report, which makes direct comparison between publications difficult; the standardized form is intended to remove this obstacle.

Abbreviations (in alphabetical order): CABG, coronary artery bypass grafting; DFUs, diabetic foot ulcers; EFD, energy flux density; eNOS, endothelial nitric oxide synthase; ESWT, extracorporeal shock wave therapy; ESWs, extracorporeal shock waves; fESWT, focused ESWT; HBOT, hyperbaric oxygen therapy; LDI, laser Doppler imaging; PCNA; proliferating cell nuclear antigen; RCT, randomized controlled trial; SWC, standard wound care; TcPO₂, transcutaneous partial oxygen pressure; V, variable; VEGF, vascular endothelial growth factor; VLUs, venous leg ulcers.

**[95]	Moretti, B.; Notarnicola, A.; Maggio, G.; Moretti, L.; Pascone, M.; Tafuri, S.; Patella, V. The management of neuropathic ulcers of the foot in diabetes by shock wave therapy. *BMC Musculoskelet. Disord.* 2009, *10*, 54. https://doi.org/10.1186/1471-2474-10-54.**

**Hypothesis:** This study tested the hypothesis that fESWT, in addition to SWC, is more effective in promoting healing of chronic neuropathic DFUs compared to SWC alone.

**Methods:** A total of 30 patients with plantar DFUs of at least six months’ duration and a minimum area of 1 cm^2^ were randomly assigned to two groups. The intervention group (n = 15) received fESWT + SWC (therapeutic footwear, debridement, silvercell dressing); the control group (n = 15) received SWC alone. fESWT was performed using a Minilith SL1 device (Storz Medical, Tägerwilen, Switzerland) at EDF of 0.03 mJ/mm^2^, delivering 100 ESWs/cm^2^ per treatment at undisclosed frequency. Three sessions were administered at 72-hour intervals. The primary endpoint was complete wound healing at 20 weeks post-baseline; secondary endpoints were re-epithelization rate (mm^2^/day) and healing time (days), also evaluated over the 20-week study period.

**Results:** At 20 weeks post-baseline, complete wound closure was achieved in 53.3% of the fESWT group compared to 33.3% of the control group. Mean healing time among those achieving complete closure was significantly shorter in the fESWT group (60.8 ± 4.7 days) than in the control group (82.2 ± 4.7 days) (p < 0.001). The re-epithelization index was also significantly higher in the fESWT group (3.0 ± 0.3 mm^2^/day) than in the control group (1.3 ± 0.3 mm^2^/day) (p < 0.001). No major adverse events were reported. One case of local infection occurred in each group; both resolved with oral antibiotics.

**Conclusions:** These results indicate that fESWT, as an adjunct to SWC, significantly improves the healing rate, accelerates wound closure and enhances re-epithelization in chronic neuropathic DFUs. The treatment was well tolerated, with no significant adverse events. These findings support the potential role of fESWT in managing chronic DFUs that are otherwise difficult to heal.

| **V** | **Result** |
| --- | --- |
| V1 | Neuropathic DFUs |
| V2 | ESWT group: 297.8 ± 129.4 mm^2^; control group: 245 ± 100.9 mm^2^ |
| V3 | Chronic; ulcers present for ≥6 months |
| V4 | RCT |
| V5 | Category 1 |
| V6 | fESWT + SWC (therapeutic footwear, debridement, silvercell dressing) |
| V7 | SWC |
| V8 | 15 |
| V9 | 15 |
| V10 | Minilith SL1 (Storz Medical, Tägerwilen, Switzerland) |
| V11 | Focused, electromagnetic ESWT |
| V12 | 0.03 mJ/mm^2^ |
| V13 | Not specified |
| V14 | 100 ESWs/cm^2^ of wound |
| V15 | 100 ESWs/cm^2^ |
| V16 | Not provided |
| V17 | 3 |
| V18 | Every 72 hours |
| V19 | Complete wound healing |
| V20 | 20 weeks post-baseline |
| V21 | Re-epithelization index; healing time; no biopsies or tissue samples analyzed |
| V22 | 20 weeks post-baseline |
| V23 | Not provided |
| V24 | Not applicable |
| V25 | Complete healing: 53.3% vs. 33.3%; healing time: 60.8 ± 4.7 vs. 82.2 ± 4.7 days; p < 0.001 |
| V26 | Re-epithelization index: 3.0 ± 0.3 mm^2^/day (fESWT group) vs. 1.3 ± 0.3 mm^2^/day (control group); p < 0.001 |
| V27 | Not provided |
| V28 | Not applicable |
| V29 | One patient in each group had local infection; resolved with oral antibiotics |
| V30 | Yes |
| V31 | Yes |
| V32 | No |
| V33 | Yes |
| V34 | No |
| V35 | No |
| V36 | No |
| V37 | Yes |
| V38 | Yes |
| V39 | Yes |
| V40 | Yes |

**[96]	Jankovic, D. Case study: Shock waves treatment of diabetic gangrene. *Int. Wound J.* 2011, *8*(2), 206–209. https://doi.org/10.1111/j.1742-481X.2011.00779.x.**

**Hypothesis:** This study tested the hypothesis that fESWT promotes wound healing and reduces pain in chronic, poorly healing diabetic gangrene, potentially avoiding limb amputation.

**Methods:** This case report describes a 75-year-old man with diabetic gangrene of both feet due to peripheral arterial occlusive disease (PAOD) and type II diabetes mellitus. Conventional therapies including antibiotics, surgical debridement and peripheral perfusion enhancement had failed. The intervention was fESWT, initially combined with rESWT and later fESWT alone, using a Duolith SD1 device (Storz Medical, Tägerwilen, Switzerland). fESWT was applied at an EFD of 0.03–0.10 mJ/mm^2^, 4 Hz and 1000–1500 ESWs per foot. In the second session, 1000 focused ESWs at 0.07 mJ/mm^2^ and 1000 radial ESWs at 2.6 bar were applied. Treatment comprised 11 sessions over approximately 11 months, at 2–4-week intervals after an initial 4-day gap between the first two. The primary outcome was resolution of necrotic tissue (wound healing), the secondary pain reduction, with long-term follow-up.

**Results:** The gangrenous necrosis resolved completely through autolytic healing, without surgical intervention. Pain, initially rated between 7 and 9 on a visual analogue scale, decreased to 2, at which point all pain medications were discontinued. Quality of life improved significantly, with reduced need for wound dressings. A transient inflammatory response after the second treatment led to a temporary pause in therapy and prophylactic antibiotic use; no adverse events occurred after resumption. Follow-up confirmed sustained wound healing and absence of pain up to two years after the last session.

**Conclusions:** This case supports fESWT as a non-invasive, well-tolerated and effective option for chronic, non-healing diabetic wounds, particularly where standard treatments have failed. The outcomes suggest that fESWT can stimulate tissue regeneration and alleviate pain, avoiding more invasive measures such as limb amputation. Further controlled studies are needed to validate these findings.

| **V** | **Result** |
| --- | --- |
| V1 | Diabetic gangrene |
| V2 | Not provided |
| V3 | Chronic |
| V4 | Case report |
| V5 | Category 5 |
| V6 | Focused and radial ESWT |
| V7 | Not applicable |
| V8 | 1 |
| V9 | Not applicable |
| V10 | Duolith SD1 (Storz Medical, Tägerwilen, Switzerland) |
| V11 | Focused, electromagnetic ESWT and radial, ballistic ESWT |
| V12 | 0.03–0.10 mJ/mm^2^ (focused); 2.6 bar (radial) |
| V13 | Not specified |
| V14 | 1000–1500 ESWs per foot (focused); 1000 ESWs (radial in initial session) |
| V15 | Not provided |
| V16 | 4 Hz |
| V17 | 11 |
| V18 | Every 2–4 weeks (after first 2 sessions) |
| V19 | Wound healing (necrosis resolution) and pain reduction |
| V20 | >2 years post-baseline |
| V21 | Pain reduction; no biopsies or tissue samples analyzed |
| V22 | >2 years post-baseline |
| V23 | No |
| V24 | Not applicable |
| V25 | Complete resolution of necrosis; pain reduction from 7–9 to 2 |
| V26 | Significant reduction in pain and wound care needs |
| V27 | No |
| V28 | Not applicable |
| V29 | Temporary inflammatory reaction after second session; resolved with antibiotics |
| V30 | Yes |
| V31 | No |
| V32 | No |
| V33 | Not applicable |
| V34 | No |
| V35 | No |
| V36 | No |
| V37 | Yes |
| V38 | Yes |
| V39 | Not applicable |
| V40 | No |

**[97]	Stieger, M.; Schmid, J.P.; Bajrami, S.; Hunziker, T. Extrakorporale Stoßwellentherapie eines komplizierten chronischen Ulcus cruris venosum. *Hautarzt* 2013, *64*(6), 443–446. https://doi.org/10.1007/s00105-012-2527-4.**

**Hypothesis:** This study tested the hypothesis that fESWT promotes wound healing in a chronic, therapy-resistant VLU by enhancing granulation tissue formation and vascularization.

**Methods:** This case report involved a 56-year-old woman with a chronic VLU of six years’ duration on the lower leg, associated with chronic venous insufficiency, lipedema with secondary lymphedema and morbid obesity. Previous conventional and advanced wound care had failed. The intervention consisted of adjuvant fESWT combined with continued compression therapy using zinc paste bandages. Focused ESWT was performed using a Duolith SD1 device (Storz Medical, Tägerwilen, Switzerland) at EFD of 0.25 mJ/mm^2^, delivering 2000 ESWs per session at a frequency of 4 Hz. Sessions were conducted once per week, totaling 30 sessions. The primary outcome was complete re-epithelialization, assessed at the end of the 30-session treatment period. The secondary outcome was improvement in lymphatic drainage, particularly lymphatic stasis and fistula formation, assessed during follow-up after initial ulcer closure.

**Results:** Following five weekly fESWT sessions, progressive granulation and re-epithelialization were observed. After completing 30 treatment sessions, the chronic ulcer showed complete closure. A recurrence occurred several weeks after therapy cessation, accompanied by increased lymphatic stasis. A second fESWT cycle, including the proximal lower leg, resulted in reduced lymphedema and partial ulcer resolution. The residual 3×3 cm ulcer was successfully closed with split-thickness skin grafting. No treatment-related adverse events were reported throughout the course of therapy.

**Conclusions:** Focused ESWT demonstrated the potential to stimulate wound healing in a long-standing, non-healing venous ulcer by promoting granulation tissue development and improving vascularization. Additionally, the therapy appeared to facilitate lymphatic drainage, potentially contributing to long-term wound management. This case highlights the utility of fESWT as a non-invasive adjunctive treatment option in complex chronic wound cases refractory to SWC.

| **V** | **Result** |
| --- | --- |
| V1 | Chronic VLU |
| V2 | 15×10 cm (approx. 150 cm^2^) |
| V3 | Chronic, present for at least 6 years |
| V4 | Case report |
| V5 | Category 5 |
| V6 | fESWT + conventional wound therapy with repeated surgical debridement and continued compression therapy with zinc paste bandages |
| V7 | Not applicable |
| V8 | 1 |
| V9 | Not applicable |
| V10 | Duolith SD1 (Storz Medical, Tägerwilen, Switzerland) |
| V11 | Focused, electromagnetic ESWT |
| V12 | 0.25 mJ/mm^2^ |
| V13 | Not specified |
| V14 | 2000 ESWs per session |
| V15 | Not stated; 2000 ESWs over approx. 200 cm^2^ (wound approx. 150 cm^2^ plus margin), i.e. approx. 10 ESWs/cm^2^ |
| V16 | 4 Hz |
| V17 | 30 sessions in the first cycle; a second ESWT cycle of unspecified length followed after the recurrence |
| V18 | Weekly intervals |
| V19 | Complete re-epithelialization |
| V20 | After 30 sessions (approx. 30 weeks) |
| V21 | Improvement in lymphatic drainage; no biopsies or other tissue samples collected for analysis related to fESWT |
| V22 | After additional sessions (not precisely defined) |
| V23 | No |
| V24 | Not applicable |
| V25 | Complete re-epithelialization observed after 30 sessions |
| V26 | Reduction of lymphatic stasis; one small ulcer remained, later closed surgically |
| V27 | No |
| V28 | Not applicable |
| V29 | No treatment-related adverse events reported |
| V30 | No |
| V31 | No |
| V32 | No |
| V33 | Not applicable |
| V34 | No |
| V35 | No |
| V36 | No |
| V37 | No |
| V38 | No |
| V39 | Not applicable |
| V40 | No |

**[98]	Variji, Z.; Aghazadeh, N.; Hasanzadeh, H.; Firooz, A. Extracorporeal shock wave therapy in the treatment of non-healing diabetic ulcer: A pilot study. *J. Clin. Exp. Dermatol. Res.* 2015, *6*, 289. https://doi.org/10.4172/2155-9554.10000289.**

**Hypothesis:** This study tested the hypothesis that fESWT is a safe and effective adjunctive treatment for non-healing DFUs, improving wound healing and local circulation.

**Methods:** This prospective case series evaluated fESWT in five patients (four male, one female; mean age 58.2 ± 19.6 years) with chronic DFUs of at least six months' duration. All had type 2 diabetes with peripheral arterial disease and neuropathy. fESWT was performed using a Duolith SD1 device with the C-ACTOR handpiece (Storz Medical, Tägerwilen, Switzerland) at EFD of 0.25 mJ/mm^2^, delivering 500 ESWs to the wound margin and 1000 ESWs to the distal limb per session (frequency not specified). Each patient received 6 to 8 weekly sessions; no control group was used. The primary outcome was reduction in ulcer surface area, measured by digital photography two weeks after the final session. Secondary outcomes, assessed concurrently, were Ankle Brachial Index (ABI) and monofilament test scores.

**Results:** Mean ulcer duration was 1.6 ± 1.0 years and mean baseline ulcer size 7.5 ± 5.1 cm^2^. Following fESWT, ulcer surface area was significantly reduced in four of the five patients, with a mean reduction of 1.2 ± 0.8 cm^2^ (p = 0.03). The fifth patient, who did not respond, had the lowest baseline ABI. ABI improved significantly in all patients, from a mean of 0.6 ± 0.1 to 0.9 ± 0.1 (p < 0.001). Monofilament test scores also improved, reflecting reduced neuropathy. No adverse events were reported.

**Conclusions:** Focused ESWT appears to be a safe and potentially effective adjunctive treatment for chronic DFUs, particularly in patients with underlying vascular and neuropathic complications. Complete re-epithelialization was not achieved in this small cohort, but improvements in ulcer size, circulation and neuropathy were observed. Further RCTs are needed to confirm efficacy and determine optimal treatment parameters.

| **V** | **Result** |
| --- | --- |
| V1 | Diabetic ulcer |
| V2 | Mean: 7.5 ± 5.1 cm^2^ |
| V3 | Chronic; mean duration: 1.6 ± 1.0 years |
| V4 | Case series |
| V5 | Category 5 |
| V6 | fESWT + SWC |
| V7 | Not applicable |
| V8 | 5 |
| V9 | Not applicable |
| V10 | Duolith SD1 with C-ACTOR handpiece (Storz Medical, Tägerwilen, Switzerland) |
| V11 | Focused, electromagnetic ESWT |
| V12 | 0.25 mJ/mm^2^ |
| V13 | Not specified |
| V14 | 500 (wound margin) + 1000 (distal limb) = 1500 per session |
| V15 | Not provided |
| V16 | Not provided |
| V17 | 6–8 |
| V18 | Weekly |
| V19 | Reduction in ulcer surface area |
| V20 | 2 weeks after final treatment session |
| V21 | ABI, monofilament test; no biopsies or tissue samples analyzed |
| V22 | 2 weeks after final treatment session |
| V23 | No |
| V24 | Not applicable |
| V25 | 4 of 5 patients had mean ulcer size reduction of 1.2 ± 0.8 cm^2^ (p = 0.03) |
| V26 | ABI improved from 0.6 ± 0.1 to 0.9 ± 0.1 (p < 0.001); monofilament test improved from 6/10 to 4/10 |
| V27 | No |
| V28 | Not applicable |
| V29 | No adverse events reported |
| V30 | Yes |
| V31 | No |
| V32 | No |
| V33 | Not applicable |
| V34 | No |
| V35 | No |
| V36 | No |
| V37 | Yes |
| V38 | No |
| V39 | Not applicable |
| V40 | Yes |

**[99]	Jeppesen, S.M.; Yderstraede, K.B.; Rasmussen, B.S.; Hanna, M.; Lund, L. Extracorporeal shockwave therapy in the treatment of chronic diabetic foot ulcers: A prospective randomised trial. *J. Wound Care* 2016, *25*(11), 641–649. https://doi.org/10.12968/jowc.2016.25.11.641.**

**Hypothesis:** This study tested the hypothesis that fESWT improves healing outcomes in patients with chronic DFUs compared with SWC alone.

**Methods:** In this prospective, open-label RCT, 23 patients with chronic DFUs were randomized 1:1 to ESWT + SWC (n=11) or SWC alone (n=12). The fESWT group received six sessions over three weeks in addition to SWC. Focused ESWT was performed using a Duolith SD1 device (Storz Medical, Tägerwilen, Switzerland) at EFD of 0.2 mJ/mm^2^ and a frequency of 5 Hz. Each session delivered 250 ESWs/cm^2^ to the ulcer surface plus a 1 cm perimeter (focal area 0–30 mm) and 500 deep ESWs (focal area 15–45 mm) to the arteries supplying the ulcer. The primary outcome was percentage reduction in ulcer area at 7 weeks post-baseline; secondary outcomes were TcPO_2_ and ulcer-related pain at 3, 5 and 7 weeks.

**Results:** At 7 weeks post-baseline, mean ulcer area reduction was 34.5% in the fESWT group and 5.6% in controls; the within-group improvement with fESWT was statistically significant (p < 0.01), although the between-group difference was not (p = 0.387). TcPO_2_ increased significantly in the fESWT group compared with controls at 3 weeks (p = 0.044), but not later. Pain scores did not differ significantly between groups at any time, and no fESWT-related adverse events were reported. A retrospective power analysis indicated that 76 patients would be required to detect a significant difference in the primary outcome (80% power, 5% significance level).

**Conclusions:** fESWT may contribute to improved local tissue oxygenation and within-group reductions in ulcer size in chronic DFUs. However, given the lack of statistically significant between-group differences and the small sample size, the authors concluded that larger, blinded RCTs are needed to confirm these findings and define the role of fESWT in diabetic wound care.

| **V** | **Result** |
| --- | --- |
| V1 | Diabetic foot ulcer |
| V2 | ESWT group: 2.3 cm^2^; control group: 2.4 cm^2^ |
| V3 | Chronic; ESWT group: 22.6 months; control group: 15.2 months |
| V4 | RCT |
| V5 | Category 2b |
| V6 | fESWT + SWC |
| V7 | SWC |
| V8 | 11 (10 analyzed) |
| V9 | 12 (11 analyzed) |
| V10 | Duolith SD1 (Storz Medical, Tägerwilen, Switzerland) |
| V11 | Focused, electromagnetic ESWT |
| V12 | 0.2 mJ/mm^2^ |
| V13 | Not specified |
| V14 | 250 ESWs/cm^2^ on ulcer + 500 ESWs on supplying arteries |
| V15 | 250 ESWs/cm^2^ |
| V16 | 5 Hz |
| V17 | 6 |
| V18 | Twice weekly over 3 weeks |
| V19 | Ulcer area reduction |
| V20 | 7 weeks |
| V21 | TcPO_2_, pain score; no biopsies or tissue samples analyzed |
| V22 | 3 weeks, 5 weeks, 7 weeks |
| V23 | No |
| V24 | Not applicable |
| V25 | fESWT group: 34.5% reduction (p < 0.01); control group: 5.6% (not significant); between-group: p = 0.387 |
| V26 | TcPO_2_ increase at 3 weeks: fESWT group +12.3%, control group –5.3%, p = 0.044; pain: not significant |
| V27 | Yes |
| V28 | 76 patients needed for 80% power |
| V29 | None reported |
| V30 | Yes |
| V31 | Yes |
| V32 | Yes |
| V33 | No |
| V34 | No |
| V35 | No |
| V36 | No |
| V37 | Yes |
| V38 | Yes |
| V39 | Yes |
| V40 | Yes |

**[100]	Saito, S.; Ishii, T.; Kamogawa, Y.; Watanabe, R.; Shirai, T.; Fujita, Y.; Shirota, Y.; Fujii, H.; Ito, K.; Shimokawa, H.; Yamaguchi, T.; Kawaguchi, Y.; Harigae, H. Extracorporeal shock wave therapy for digital ulcers of systemic sclerosis: A phase 2 pilot study. *Tohoku J. Exp. Med.* 2016, *238*(1), 39–47. https://doi.org/10.1620/tjem.238.39.**

**Hypothesis:** This study tested the hypothesis that fESWT accelerates healing and improves clinical outcomes in patients with refractory digital ulcers associated with systemic sclerosis (SSc), beyond the effects of conventional pharmacological treatments.

**Methods:** This case series included nine patients with systemic sclerosis with at least one newly developed digital ulcer of ischemic origin due to SSc-related vascular impairment. There was no control group; patients continued standard vasodilator and immunosuppressive therapy. fESWT was performed using a Duolith SD1 device (Storz Medical, Tägerwilen, Switzerland) at EFD ranging from 0.08 to 0.25 mJ/mm^2^ and a frequency of 4 Hz, with 7,000 ESWs per session, once weekly for nine weeks. The primary endpoint was ulcer number and diameter at 20 weeks after the first ESWT session. Secondary endpoints were pain (VAS and PainVision), skin thickness (Rodnan score), disability (HAQ), health-related quality of life (EQ-5D) and fingertip temperature, evaluated at baseline, 9, 15 and 20 weeks.

**Results:** The number of ulcers decreased significantly from a mean of 5.4 at baseline to 2.2 at 20 weeks (p < 0.05). Among 18 ulcers larger than 5 mm, 10 healed completely and their mean diameter decreased from 10.9 mm to 2.5 mm. VAS Pain scores improved from a mean of 43.3 to 30.2 and Rodnan skin scores improved significantly. HAQ and EQ-5D improved during the treatment period but were not statistically significant at 20 weeks. Finger temperature changes were variable and not significant. No adverse events were reported and treatment was well tolerated.

**Conclusions:** This pilot study suggests that fESWT may offer a beneficial, safe and noninvasive treatment option for improving ulcer healing and clinical symptoms in systemic sclerosis-related digital ulcers. Further RCTs with larger populations are necessary to confirm these findings and determine the long-term efficacy and optimal regimen of fESWT.

| **V** | **Result** |
| --- | --- |
| V1 | Digital ulcers associated with systemic sclerosis |
| V2 | Mean diameter of large ulcers: 10.9 mm (18 ulcers >5 mm) |
| V3 | Chronic; disease duration 5–32 years (mean 12.6 years) |
| V4 | Case series |
| V5 | Category 5 |
| V6 | fESWT + SWC |
| V7 | Not applicable |
| V8 | 9 |
| V9 | Not applicable |
| V10 | Duolith SD1 (Storz Medical, Tägerwilen, Switzerland) |
| V11 | Electromagnetic ESWT |
| V12 | 0.08–0.25 mJ/mm^2^ |
| V13 | Not specified |
| V14 | 7,000 ESWs per session |
| V15 | Not provided |
| V16 | 4 Hz |
| V17 | 9 sessions |
| V18 | 1 session per week |
| V19 | Number and diameter of ulcers at 20 weeks after the start of treatment |
| V20 | 20 weeks after the start of treatment |
| V21 | Pain VAS, PainVision score, HAQ, EQ-5D, Rodnan skin score and fingertip temperature; no biopsies or tissue samples analyzed |
| V22 | 9, 15 and 20 weeks after the start of treatment |
| V23 | No |
| V24 | Not applicable |
| V25 | Mean number of ulcers decreased from 5.4 to 2.2 at 20 weeks after the start of treatment (p < 0.05) |
| V26 | VAS 43.3 → 30.2; PainVision improved; Rodnan improved, p < 0.05; HAQ/EQ-5D improved at 9, not 20 weeks |
| V27 | No |
| V28 | Not applicable |
| V29 | No adverse events reported |
| V30 | Yes |
| V31 | No |
| V32 | No |
| V33 | Not applicable |
| V34 | No |
| V35 | No |
| V36 | No |
| V37 | Yes |
| V38 | Yes |
| V39 | Not applicable |
| V40 | Yes |

**[101]	Porso, M.; Loreti, S.; Nusca, S.M.; Luziatelli, S.; Caccia, D.; Taborri, G.; Trischitta, D.; Taurino, M.; Padua, L.; Saraceni, V.M.; Vulpiani, M.C.; Vetrano, M. Defocused shock wave therapy for chronic soft tissue wounds in the lower limbs: A pilot study. *Ultrasound Med. Biol.* 2017, *43*(1), 362–369. https://doi.org/10.1016/j.ultrasmedbio.2016.08.038.**

**Hypothesis:** This study tested the hypothesis that defocused ESWT leads to significant wound healing and pain reduction in patients unresponsive to advanced dressing treatments.

**Methods:** This pilot cohort study involved ten patients with chronic soft tissue ulcers of varying etiology (venous, diabetic, post-traumatic, mixed), unresponsive to standard treatments for more than three months. Defocused ESWT was performed using a Duolith SD1 device (Storz Medical, Tägerwilen, Switzerland) at EFD of 0.15 mJ/mm^2^ and a frequency of 4 Hz. Each patient received 3 sessions at 72-hour intervals, each with 300 ESWs to the wound edge plus 100 ESWs/cm^2^ wound area. The primary outcome was wound size reduction, assessed by computerized digital photo documentation (Woundsoft software). Secondary outcomes were wound healing (Bates-Jensen Wound Assessment Tool, BJWAT) and pain (Visual Analogue Scale, VAS), assessed at baseline, 15, 30 and 90 days post-treatment.

**Results:** At the 90-day follow-up, seven of ten ulcers (70%) achieved complete closure with 100% epithelialization and one showed partial healing (33% epithelialization). Two ulcers remained unchanged, though these patients experienced significant pain relief. Median wound size at 90 days was 0 cm^2^ and the BJWAT score improved significantly. Median VAS score decreased to 0, with complete pain relief in nine of ten patients. BJWAT reductions were significant at all time points (15, 30 and 90 days) and wound size reductions at 30 and 90 days (p = 0.005 each); the reduction at 15 days was not significant (p = 0.059).

**Conclusions:** This pilot study suggests that defocused ESWT is a non-invasive, effective therapy for chronic soft tissue ulcers, promoting significant wound healing and pain reduction in patients unresponsive to SWC. It may serve as an alternative or adjunct for difficult-to-heal ulcers, although larger controlled studies are necessary to confirm these results and establish optimal protocols.

| **V** | **Result** |
| --- | --- |
| V1 | Chronic soft tissue wounds (arterial, diabetic, mixed, venous, post-traumatic) |
| V2 | Median baseline wound size ranged from 0.5 to 8.8 cm^2^ |
| V3 | Chronic (>3 months); median duration: 10 months (range 3–52 months) |
| V4 | Case series |
| V5 | Category 5 |
| V6 | Defocused ESWT + SWC |
| V7 | Not applicable |
| V8 | 10 |
| V9 | Not applicable |
| V10 | Duolith SD1 (Storz Medical, Tägerwilen, Switzerland) |
| V11 | Defocused, electromagnetic ESWT |
| V12 | 0.15 mJ/mm^2^ |
| V13 | Not specified |
| V14 | 300 ESWs + 100 ESWs/cm^2^ |
| V15 | 100 ESWs/cm^2^ |
| V16 | 4 Hz |
| V17 | 3 sessions |
| V18 | 72 hours |
| V19 | Wound size, BJWAT, VAS score |
| V20 | 15, 30 and 90 days post-treatment |
| V21 | VAS pain score assessed alongside healing; no biopsies or tissue samples analyzed |
| V22 | 15, 30 and 90 days post-treatment |
| V23 | No |
| V24 | Not applicable |
| V25 | 7/10 complete healing, 1/10 improved, 2/10 unchanged; significant size, BJWAT and VAS improvements (p < 0.05) |
| V26 | Not applicable |
| V27 | No |
| V28 | Not applicable |
| V29 | None reported (no adverse events) |
| V30 | Yes |
| V31 | No |
| V32 | No |
| V33 | Not applicable |
| V34 | No |
| V35 | No |
| V36 | No |
| V37 | Yes |
| V38 | Yes |
| V39 | Not applicable |
| V40 | Yes |

**[102]	Taheri, P.; Shahbandari, M.; Parvaresh, M.; Vahdatpour, B. Extracorporeal shockwave therapy for chronic venous ulcers: A randomized controlled trial. *Galen Med. J.* 2021, *10*, e1931. https://doi.org/10.31661/gmj.v10i0.1931.**

**Hypothesis:** This study tested the hypothesis that fESWT, as an adjunct to SWC, significantly improves wound healing, reduces pain and enhances quality of life in chronic VLUs compared to SWC alone.

**Methods:** In this RCT, 50 patients with chronic VLUs due to chronic venous insufficiency were randomized to ESWT (n=25) or sham ESWT (n=25), both with standard compression bandaging; after 3 dropouts per group, 22 per group (44 total) were analyzed. Focused ESWT was performed using a Duolith SD1 device (Storz Medical, Tägerwilen, Switzerland) at undisclosed EFD and 5 Hz, with 100 ESWs/cm^2^ of wound area per session, weekly for four weeks. The primary outcome was wound size reduction; secondary outcomes were pain intensity (Visual Analogue Scale), patient satisfaction and quality of life (Charing Cross Venous Ulcer Questionnaire, CCVUQ), at baseline and weeks 4 and 8.

**Results:** Wound size decreased in both groups over 8 weeks, but the between-group difference was not significant (p = 0.281). Between-group differences significantly favored fESWT for patient satisfaction (p < 0.001) and for the aesthetics and emotional-state subscales and total CCVUQ score (p < 0.001 to p = 0.005); on the CCVUQ a lower score indicates better quality of life, yet the total score rose in both groups (ESWT 55.1 ± 6.5 to 55.9 ± 6.6; control 59.0 ± 5.3 to 61.8 ± 5.5), i.e. it deteriorated in both, significantly less with ESWT. Pain reduction was greater with fESWT, though not statistically significant. No serious adverse events occurred, although some patients initially experienced discomfort during fESWT.

**Conclusions:** fESWT was found to be a safe and effective adjunct in chronic VLUs, contributing to significant improvements in patient satisfaction and quality of life. While wound size differences were not statistically significant, the therapy showed beneficial clinical trends and merits further investigation.

| **V** | **Result** |
| --- | --- |
| V1 | Chronic VLUs |
| V2 | ESWT group: 567.8 ± 644.2 mm^2^; control group: 715.8 ± 769.7 mm^2^ |
| V3 | Chronic; mean duration ~62 weeks (fESWT group) and ~57 weeks (control group) |
| V4 | RCT |
| V5 | Category 2b |
| V6 | ESWT + SWC |
| V7 | Sham ESWT (device off) + SWC |
| V8 | 22 patients |
| V9 | 22 patients |
| V10 | Duolith SD1 (Storz Medical, Tägerwilen, Switzerland) |
| V11 | Focused, electromagnetic ESWT |
| V12 | Not specified |
| V13 | Not specified |
| V14 | 100 ESWs/cm^2^ |
| V15 | 100 ESWs/cm^2^ |
| V16 | 5 Hz |
| V17 | 4 sessions |
| V18 | 1 week |
| V19 | Changes in wound size, pain intensity, patient satisfaction and quality of life (CCVUQ) |
| V20 | Weeks 4 and 8 post-baseline |
| V21 | Quality of life (CCVUQ subscales), satisfaction score; no biopsies or tissue samples analyzed |
| V22 | Weeks 4 and 8 |
| V23 | Not provided |
| V24 | Not provided |
| V25 | Wound size 94.8 ± 176.2 (fESWT) vs. 307.6 ± 433.6 (control), p = 0.281; satisfaction p < 0.001; VAS p = 0.860 |
| V26 | CCVUQ significantly better in the fESWT group for aesthetics and emotional state (p < 0.001); total score p = 0.005 |
| V27 | Not provided |
| V28 | Not provided |
| V29 | ESWT-induced pain initially, no serious adverse events |
| V30 | Yes |
| V31 | Yes |
| V32 | No |
| V33 | Yes |
| V34 | Yes |
| V35 | No |
| V36 | Yes |
| V37 | Yes |
| V38 | Yes |
| V39 | Yes |
| V40 | Yes |

**[103]	Rassweiler, J.J.; Scheitlin, W.; Goezen, A.S.; Rassweiler-Seyfried, M.C. Low-energy shockwave therapy in the management of wound healing following Fournier’s gangrene. *Eur. Urol. Open Sci.* 2022, *45*, 8–11. https://doi.org/10.1016/j.euros.2022.08.019.**

**Hypothesis:** This study tested the hypothesis that fESWT effectively promotes wound healing in patients with secondary wound complications following surgical treatment of Fournier’s gangrene.

**Methods:** This case series without a control group describes fESWT in four male patients (ages 27, 57, 61 and 68 years) with acute necrotizing fasciitis (Fournier’s gangrene) requiring extensive surgical debridement of scrotal and perineal skin. Three patients underwent reconstructive surgery with skin flaps, which subsequently developed wound dehiscence. The fourth had complete necrosis of the scrotal and penile skin with insufficient tissue for plastic reconstruction and thus received fESWT as the sole wound management strategy. Focused ESWT was performed using a Duolith SD1 device (Storz Medical, Tägerwilen, Switzerland) with an EFD of 0.25 mJ/mm^2^ and a frequency of 3 Hz, with 2000 ESWs per session, three times weekly for six weeks. The primary outcome was complete wound closure at 12 weeks post-baseline; no secondary outcomes were defined.

**Results:** All four patients demonstrated significant wound healing. The three patients with wound dehiscence following skin flap procedures experienced near-complete closure without further surgery. The fourth patient, managed exclusively with fESWT due to the lack of reconstructive options, showed complete restoration of scrotal and penile skin within 12 weeks. No adverse events were reported and no further surgical intervention was necessary. The clinical outcomes were considered dramatic and highly positive.

**Conclusions:** This case series provides the first report of successful fESWT in patients with Fournier’s gangrene-related wounds. The observed restoration of local tissue, rather than fibrotic closure, may suggest regenerative effects potentially involving stem cell recruitment. Given the favorable outcomes in all four cases, fESWT appears a promising adjunct or alternative to surgical management in selected patients. However, controlled trials are needed to confirm efficacy and elucidate the underlying mechanisms.

| **V** | **Result** |
| --- | --- |
| V1 | Fournier’s gangrene |
| V2 | Not provided |
| V3 | Acute; dehiscence after initial healing |
| V4 | Case series |
| V5 | Category 5 |
| V6 | fESWT |
| V7 | Not applicable |
| V8 | 4 |
| V9 | Not applicable |
| V10 | Duolith SD1 (Storz Medical, Tägerwilen, Switzerland) |
| V11 | Focused, electromagnetic ESWT |
| V12 | 0.25 mJ/mm^2^ |
| V13 | Not specified |
| V14 | 2000 |
| V15 | Not provided |
| V16 | 3 Hz |
| V17 | Approximately 18 sessions (3 per week for 6 weeks) |
| V18 | Three treatment sessions per week |
| V19 | Not provided |
| V20 | 12 weeks |
| V21 | None explicitly defined or assessed; no biopsies or tissue samples analyzed |
| V22 | Not provided |
| V23 | No |
| V24 | Not applicable |
| V25 | Almost complete healing of three flap-dehiscence wounds at 12 wk; fourth patient: full closure, scrotal/penile skin restored |
| V26 | Not provided |
| V27 | No |
| V28 | Not applicable |
| V29 | None reported |
| V30 | No |
| V31 | No |
| V32 | No |
| V33 | Not applicable |
| V34 | No |
| V35 | No |
| V36 | No |
| V37 | Yes |
| V38 | No |
| V39 | Not applicable |
| V40 | No |

**[104]	Ishii, T.; Kawaguchi, Y.; Ishikawa, O.; Takemori, H.; Takasawa, N.; Kobayashi, H.; Takahashi, Y.; Yasuoka, H.; Kodera, T.; Takai, O.; Nakaya, I.; Sato, Y.; Izumiyama, T.; Fujii, H.; Kamogawa, Y.; Shirota, Y.; Shirai, T.; Fujita, Y.; Saito, S.; Chiu, S.W.; Yamaguchi, T.; Shimokawa, H.; Harigae, H. Effectiveness and safety of low-energy shock wave therapy for digital ulcers associated with systemic sclerosis: A phase 3 pivotal clinical trial. *Mod. Rheumatol.* 2025, *35*(3), 484–495. https://doi.org/10.1093/mr/roae104.**

**Hypothesis:** This study tested the hypothesis that fESWT is effective and safe in reducing the number of digital ulcers in systemic sclerosis (SSC), compared to conventional treatment.

**Methods:** This non-randomized controlled cohort study included 60 patients with SSc-associated refractory digital ulcers persisting despite over 4 weeks of conventional treatment; 30 received fESWT plus conventional treatment (vasodilators, prostaglandins etc.), 30 conventional treatment alone. Focused ESWT was performed using a Duolith SD1 device (Storz Medical, Tägerwilen, Switzerland) at an EFD of 0.08 to 0.25 mJ/mm^2^ and 4 Hz, 2000 ESWs per session, once weekly for 8 consecutive weeks. The primary outcome was the mean reduction in digital ulcer number at 8 weeks post-baseline; secondary outcomes at 4, 8 and 12 weeks.

**Results:** At 8 weeks, mean ulcer count reduction was significantly greater with fESWT than with conventional treatment (4.5, SD 2.6 versus 0.8, SD 2.8; p < 0.001). The ulcer reduction rate favored fESWT at 4 and 8 weeks (p < 0.001) but not at 12 weeks (p = 0.178); more fESWT patients reached ≥20%, ≥50% and ≥70% reduction and fewer new ulcers appeared (p = 0.013). No significant between-group differences were found for VAS pain improvement rate, HAQ, EQ-5D or modified Rodnan TSS (p = 0.082, 0.084; 0.257, 0.896; 0.507, 0.430; 0.463, respectively). Adverse events affected 18/30 (60.0%) of fESWT and 16/30 (53.3%) of control patients (no significant difference); six were possibly fESWT-related, all mild and resolving. No deaths occurred and all patients completed the protocol.

**Conclusions:** The findings suggest that fESWT is a safe and effective adjunctive treatment for digital ulcers in SSc, significantly reducing ulcer number and new ulcer incidence compared to conventional treatment alone, whereas no significant between-group differences were found for pain, disability, quality of life or skin sclerosis.

| **V** | **Result** |
| --- | --- |
| V1 | Digital ulcers (systemic sclerosis-associated) |
| V2 | Not provided |
| V3 | Chronic; >4 weeks duration prior to study entry |
| V4 | Cohort study with non-randomized control group |
| V5 | Category 1 |
| V6 | fESWT + conventional treatment (vasodilators, prostaglandins etc.) |
| V7 | Conventional treatment |
| V8 | 30 |
| V9 | 30 |
| V10 | Duolith SD1 (Storz Medical, Tägerwilen, Switzerland) |
| V11 | Electromagnetic focused ESWT |
| V12 | 0.08–0.25 mJ/mm^2^ |
| V13 | Not specified |
| V14 | 100 ESWs per area (10 hand, five forearm); stated as 2000 pulses per upper extremity, inconsistent with areas |
| V15 | Not provided |
| V16 | 4 Hz |
| V17 | 8 sessions |
| V18 | Weekly |
| V19 | Mean reduction in number of ulcers at 8 weeks post-baseline |
| V20 | 8 weeks post-baseline |
| V21 | Ulcer count reduction at 4 and 12 weeks, pain (VAS), m-Rodnan, HAQ, EQ-5D; no biopsies or tissue samples |
| V22 | 4, 8 and 12 weeks post-baseline |
| V23 | Yes |
| V24 | 25 patients per group for 80% power (SD=5, α=0.05), final n=30 per group for safety |
| V25 | Mean ulcer reduction at 8 weeks: ESWT group = 4.5 (SD 2.6); control group = 0.8 (SD 2.8); p<0.001 |
| V26 | 4 wk 3.33 ± 2.35 vs 0.30 ± 1.91, p < 0.001; 12 wk 3.93 ± 2.32 vs 1.60 ± 2.49, p < 0.001; new ulcers 0.23 ± 0.43 vs 1.57 ± 2.81, p = 0.013 |
| V27 | No |
| V28 | Not applicable |
| V29 | Six mild fESWT-related AEs (ulcer infection, 2x hand swelling, periungitis, Wolff-Parkinson-White, pretibial edema), resolved; four severe, none device-related |
| V30 | Yes |
| V31 | No |
| V32 | No |
| V33 | No |
| V34 | No |
| V35 | No |
| V36 | No |
| V37 | Yes |
| V38 | Yes |
| V39 | Yes |
| V40 | Yes |

**[105]	Landscheidt, K.; Alabdulmohsen, A.; Hübscher, M.; Geber, B.; Hernekamp, J.F.; Goertz, O. Extracorporeal shock waves therapy for the treatment of acute and chronic wounds—A prospective, monocentric clinical trial to examine the effect of shock waves on wound healing. *Health Sci. Rep.* 2025, *8*(1), e70311. https://doi.org/10.1002/hsr2.70311.**

**Hypothesis:** This study tested the hypothesis that fESWT accelerates and improves epithelialization at skin graft donor and recipient sites versus placebo or sham.

**Methods:** This prospective RCT enrolled 35 patients (mean age 68.6 years) with chronic wounds requiring split-thickness skin grafts: DFUs and VLUs (25.7% each), soft tissue defects after necrotizing fasciitis (8.6%), decollement injuries (5.7%), healing disorders after osteosynthesis or unknown cause (34.3%). Group 1 (n = 25) received intraindividual fESWT (area A) versus placebo (area B); Group 2 (n = 10) received sham treatment with a shockwave-absorbing transducer (both areas). Focused ESWT was delivered by a Duolith SD1 device (Storz Medical, Tägerwilen, Switzerland), EDF 0.25 mJ/mm^2^, 100 ESWs/cm^2^ at 4 Hz, in three sessions on postoperative days 5, 7 and 9. The primary endpoint was re-epithelialization of donor and recipient areas by planimetric image analysis on days 7, 9, 12 and 90. Secondary endpoints were wound infection rate, moisture, pain (VAS) and microbial colonization.

**Results:** In Group 1, fESWT-treated areas re-epithelialized significantly faster than placebo: day 7 (donor site 0.7 vs. 0.6, p < 0.01), day 9 (0.8 vs. 0.7, p < 0.01) and day 12 (0.9 vs. 0.8, p < 0.05). Recipient sites improved similarly (days 7, 9 and 12; p < 0.001 to p = 0.008). No significant difference was found versus the sham group, likely due to Group 2's low sample size. Wound infection was significantly lower in fESWT-treated than placebo areas on days 7, 9 and 12. Moisture and pain did not differ significantly; no adverse events occurred.

**Conclusions:** Repeated fESWT significantly accelerated healing and reduced infection rates at donor and recipient sites compared to placebo in chronic wounds treated by skin grafts. It was well tolerated and supported as a noninvasive adjunct to conventional wound care.

| **V** | **Result** |
| --- | --- |
| V1 | Mixed |
| V2 | >50 cm^2^ |
| V3 | Acute (donor), chronic (recipient) |
| V4 | RCT |
| V5 | Category 1 |
| V6 | fESWT |
| V7 | Placebo/sham treatment (device off or with shockwave-absorbing transducer) |
| V8 | 25 |
| V9 | 10 |
| V10 | Duolith SD1 (Storz Medical, Tägerwilen, Switzerland) |
| V11 | Focused, electromagnetic ESWT |
| V12 | 0.25 mJ/mm^2^ |
| V13 | Not specified |
| V14 | ≥5000 |
| V15 | 100 ESWs/cm^2^ |
| V16 | 4 Hz |
| V17 | 3 |
| V18 | Days 5, 7 and 9 post-baseline |
| V19 | Intraindividual difference in epithelialized area (fESWT vs. placebo/sham) |
| V20 | Days 7, 9, 12 and 90 post-baseline |
| V21 | Wound infection, moisture, pain, microbial colonization, dropout; no biopsies or tissue samples analyzed |
| V22 | Days 0, 5, 7, 9, 12 and 90 post-baseline |
| V23 | No |
| V24 | Not applicable |
| V25 | Day 12: donor 0.92 (fESWT) vs. 0.82 (placebo), p < 0.05; recipient smaller residual area, p = 0.008 |
| V26 | Lower infection rate in fESWT group (p < 0.05); other differences not significant |
| V27 | No |
| V28 | Not applicable |
| V29 | No treatment-related adverse events |
| V30 | Yes |
| V31 | Yes |
| V32 | No |
| V33 | Yes |
| V34 | Yes |
| V35 | No |
| V36 | Yes |
| V37 | Yes |
| V38 | Yes |
| V39 | Yes |
| V40 | Yes |
