## Supplementary File 4 for "Extracorporeal Shock Wave Therapy for Wound Management: Clinical Evidence, Energy Delivery Parameters and Mechanistic Insights — A Systematic Review"

**Extracorporeal Shock Wave Therapy in Wound Management: A Comprehensive Systematic Review of Clinical Evidence, Modalities and Mechanisms**

by Brent Musolf, Carmen Nussbaum-Krammer, Michael O’Neal, Nicola Maffulli and Christoph Schmitz

**Supplementary File 4**

**Standardized summaries and 40 key variables (c.f. Table 2 in the main text) from clinical studies on piezoelectric ESWT for wound management**

**(numbers in brackets refer to the reference numbers in the main text)**

**Note on the standardized summaries:** Each summary condenses one clinical study as it was reported, together with the interpretation that its own authors placed on their findings. The summaries therefore reproduce the position of the respective authors and the state of the field at the time of that publication, and not the assessment of the present systematic review. They follow a uniform structure (Hypothesis, Methods, Results and Conclusions), because the original abstracts differ widely in structure, in length and in the information they report, which makes direct comparison between publications difficult; the standardized form is intended to remove this obstacle.

Abbreviations (in alphabetical order): CABG, coronary artery bypass grafting; DFUs, diabetic foot ulcers; EFD, energy flux density; eNOS, endothelial nitric oxide synthase; ESWT, extracorporeal shock wave therapy; ESWs, extracorporeal shock waves; fESWT, focused ESWT; HBOT, hyperbaric oxygen therapy; LDI, laser Doppler imaging; PCNA; proliferating cell nuclear antigen; RCT, randomized controlled trial; SWC, standard wound care; TcPO₂, transcutaneous partial oxygen pressure; V, variable; VEGF, vascular endothelial growth factor; VLUs, venous leg ulcers.

**[106]	Dolibog, P.; Dolibog, P.; Franek, A.; Brzezińska-Wcisło, L.; Arasiewicz, H.; Wróbel, B.; Chmielewska, D.; Ziaja, J.; Błaszczak, E. Randomized, controlled clinical pilot study of venous leg ulcers treated with using two types of shockwave therapy. *Int. J. Med. Sci.* 2018, *15*(12), 1275–1285. https://doi.org/10.7150/ijms.26614.**

**Note: Dolibog et al. (2018) investigated both piezoelectric fESWT and rESWT; only the fESWT results are presented here.**

**Hypothesis:** fESWT was hypothesized to heal VLUs more effectively than SWC, with a faster reduction in ulcer size and a higher percentage of complete healing.

**Methods:** Three-arm RCT: 65 patients assessed, 57 randomized, 50 completed (17 rESWT, 15 fESWT, 18 SWC); the 33 patients referred to here are only the fESWT arm (n=15) and the SWC arm (n=18), receiving SWC (saline dressings, compression bandages). The fESWT group received 6 sessions with a Piezowave device (Richard Wolf, Knittlingen, Germany) at 5-day intervals, 100 ESWs/cm^2^, EFD 0.173 mJ/mm^2^ and 5 Hz. The primary endpoint was change in total ulcer surface area 4 weeks post-treatment; secondary outcomes were percentage change in ulcer dimensions, the Gilman index and completely healed ulcers, assessed 4 weeks after the last therapy session.

**Results:** The fESWT group showed a statistically significant reduction in ulcer size, an average decrease of 63.5% versus 38.9% in controls (p = 0.084, not statistically significant). Completely healed ulcers were more frequent with fESWT (4/15 = 26% vs 0/18); no statistical test was reported for healing. Maximum ulcer length and width improved more with fESWT, whereas the Gilman index was essentially the same (0.3 ± 0.1 cm vs 0.3 ± 0.2 cm; p(B C) = 1); none of these fESWT vs SWC comparisons was significant.

**Conclusions:** fESWT was concluded to be more effective than SWC; rESWT and fESWT not differing significantly; for the fESWT arm, none of the between-group comparisons with standard care in Table 3 reached statistical significance, although the Abstract and the nonlinear T1/2 analysis (Table 4) claimed a significant difference versus standard care. Further, larger studies with longer follow-up are needed.

| **V** | **Result** |
| --- | --- |
| V1 | VLUs |
| V2 | 8.1 ± 12.4 cm^2^ (fESWT group); 8.3 ± 4.6 cm^2^ (SWC group) |
| V3 | Chronic; duration 3–24 months (fESWT group), 1–48 months (SWC group) |
| V4 | RCT |
| V5 | Category 2b |
| V6 | fESWT |
| V7 | SWC (saline dressings and compression bandages) |
| V8 | 15 |
| V9 | 18 |
| V10 | Piezowave (Richard Wolf, Knittlingen, Germany) |
| V11 | Focused, piezoelectric ESWT |
| V12 | 0.173 mJ/mm^2^ |
| V13 | Not specified |
| V14 | 100 ESWs/cm^2^ |
| V15 | 100 ESWs/cm^2^ |
| V16 | 5 Hz |
| V17 | 6 sessions |
| V18 | Every 5 days |
| V19 | Change in total ulcer surface area and linear dimensions inside groups |
| V20 | 4 weeks after last treatment (i.e., ~8 weeks total) |
| V21 | Completely healed wounds; Gilman index; ulcer surface area change; nonlinear time-to-halving; no biopsies |
| V22 | 4 weeks after last treatment (same as primary endpoint) |
| V23 | Not provided |
| V24 | Not provided |
| V25 | Ulcer area 63.5% (fESWT) vs 38.9% (control), not significant: p(B C) = 0.084 (8 weeks), 0.159 (4 weeks) |
| V26 | 26% complete healing (fESWT), none in control; Gilman index 0.3 ± 0.1 vs 0.3 ± 0.2 cm; significance not always specified |
| V27 | Not provided |
| V28 | Not provided |
| V29 | Not provided (the paper did not report on adverse events or tolerability) |
| V30 | Yes |
| V31 | Yes |
| V32 | No |
| V33 | Yes |
| V34 | No |
| V35 | No |
| V36 | No |
| V37 | Yes |
| V38 | Yes |
| V39 | Yes |
| V40 | Yes |

**[107]	Hitchman, L.H.; Totty, J.P.; Cai, P.; Smith, G.E.; Carradice, D.; Chetter, I.C. Extracorporeal shockwave therapy for diabetic foot ulcers: A feasibility study. *J. Wound Care* 2023, *32*(3), 182–192. https://doi.org/10.12968/jowc.2023.32.3.182.**

**Hypothesis:** The primary aim was feasibility of delivering fESWT to DFU patients and of a definitive trial; clinical effect on wound healing and patient-reported benefit were secondary. Failure of conventional treatment was not an eligibility criterion.

**Methods:** Prospective, single-center, mixed-methods feasibility study; 106 patients were screened, 51 (48.1%) eligible and 24 recruited (22.6% of screened, 47.1% of eligible). The neuropathic/ischemic breakdown was not reported. All patients received fESWT with a PiezoWave2 device (Richard Wolf, Knittlingen, Germany): 3 sessions over 7 day, EFD 0.1 mJ/mm^2^ and 120 ESWs /cm^2^ per session; the frequency was not stated. The primary outcome was feasibility, assessed by improvements in wound size, healing rate and quality of life (QoL); secondary outcomes were wound size reduction, ulcer healing at 12 weeks post-baseline and patient satisfaction, measured at baseline and 4, 8 and 12 weeks.

**Results:** Of 24 recruited patients, two withdrew before treatment; 22 underwent the intervention and were analyzed; 18 (81.8%) attended all three sessions and 10 (45.5%) the 12-week follow-up. At 12 weeks, nine ulcers (40.9%) had healed (the Abstract gives 45.5%; the Results section nine of 22, i.e. 40.9%), with mean ulcer surface area reduced by 83.3% and mean volume by 91.6% (no statistical test; the 95% confidence intervals for the 12-week values include zero). No average or median time to healing was reported; QoL improved. Mild adverse effects (pain, burning, itching) did not stop treatment; no major adverse events occurred; 90.9% attended at least two of the three sessions.

**Conclusions:** fESWT for DFUs was concluded to be feasible, well-tolerated and promising for healing and QoL. Despite the absence of a control group, fESWT could be a viable therapeutic option for DFU management, warranting a larger RCT to confirm efficacy and optimal parameters.

| **V** | **Result** |
| --- | --- |
| V1 | DFUs |
| V2 | Surface area: 4.4 ± 10.2 cm^2^, depth: 0.4 ± 0.5 cm |
| V3 | Ulcer age: 36.5 ± 312.9 days (0–1095 days) |
| V4 | Prospective single-arm feasibility (cohort) study |
| V5 | Category 5 |
| V6 | fESWT |
| V7 | Not applicable |
| V8 | 22 |
| V9 | Not applicable |
| V10 | PiezoWave2 (Richard Wolf, Knittlingen, Germany) |
| V11 | Focused, piezoelectric ESWT |
| V12 | 0.1 mJ/mm^2^ |
| V13 | Not specified |
| V14 | 120 ESWs/cm^2^ |
| V15 | 120 ESWs/cm^2^ |
| V16 | Not specified |
| V17 | 3 sessions over 7 days |
| V18 | Not specified (the paper stated only that three sessions were given over a seven-day period) |
| V19 | Feasibility of delivering fESWT; secondary outcomes include wound size reduction, DFU healing at 12 weeks post-baseline and QoL improvement |
| V20 | 12 weeks post-baseline |
| V21 | Change in ulcer size, ulcers healed at 12 weeks, HRQoL (EQ-5D-3L, SF-12); no biopsies |
| V22 | 1, 4, 8 and 12 weeks post-baseline |
| V23 | No |
| V24 | Not applicable |
| V25 | nine DFUs (40.9%) healed at 12 weeks (Abstract 45.5%; Results 40.9%); ulcer surface area reduced 83.3%, volume 91.6% |
| V26 | HRQoL improved; pain decreased, but completion rates for questionnaires dropped over time |
| V27 | No |
| V28 | Not applicable |
| V29 | Mild pain, burning, or itching during treatment, resolved after treatment |
| V30 | Yes |
| V31 | No |
| V32 | No |
| V33 | Not applicable |
| V34 | No |
| V35 | No |
| V36 | No |
| V37 | Yes |
| V38 | Yes |
| V39 | Not applicable |
| V40 | Yes |

**[24]	Dolibog, P.T.; Dolibog, P.; Bergler-Czop, B.; Grzegorczyn, S.; Chmielewska, D. The efficacy of extracorporeal shockwave therapy compared with compression therapy in healing venous leg ulcers. *J. Clin. Med.* 2024, *13*(7), 2117. https://doi.org/10.3390/jcm13072117.**

**Note: Dolibog et al. (2024) investigated both piezoelectric fESWT and rESWT; only the fESWT results are presented here.**

**Hypothesis:** fESWT was hypothesized to improve the healing of VLUs significantly compared to SWC and to produce a clinically meaningful difference in wound healing rates and wound size reduction.

**Methods:** RCT in chronic VLUs; 69 patients were allocated to four groups (IPC n=20, rESWT n=16, fESWT n=15, SWC n=18); the 33 patients referred to here are only the fESWT arm (n=15) and the SWC arm (n=18). fESWT used a Piezowave device with an F10G4 applicator (Richard Wolf, Knittlingen, Germany), 6 sessions spaced 5 days apart over 4 weeks, each delivering 100 ESWs/cm^2^ at 5 Hz and an EFD of 0.173 mJ/mm^2^. Controls received SWC (daily gauze dressings, elastic bandages). The primary outcome was the relative change in ulcer surface area after 4 weeks, assessed at baseline and the 4-week follow-up; the secondary outcome was the weekly wound healing rate (WHR).

**Results:** After 4 weeks, the fESWT group showed a median relative reduction of 18% in wound area, the control group only 16%. Both groups had statistically significant reductions from baseline, but no significant difference between them. The mean weekly healing rate was higher with fESWT (4.3 vs 3.1 mm/week), but the paper explicitly stated that there were no differences in WHR among the rESWT, fESWT and SWC groups, and the weekly medians in Table 3 favored SWC at weeks 2 and 4.

**Conclusions:** fESWT had a positive effect on the healing of VLUs, but its efficacy was similar to that of SWC. Both led to significant wound healing, though the difference was not statistically significant. fESWT may offer an additional option for VLU wound management, but further studies are needed to establish clinical superiority over traditional therapies.

| **V** | **Result** |
| --- | --- |
| V1 | VLUs |
| V2 | Median initial wound surface area 7.4 cm^2^ (25Q-75Q 4.6-16.2) fESWT; 10.5 cm^2^ (25Q-75Q 5.8-12.0) SWC |
| V3 | Chronic (duration of VLU: fESWT group 12 months, control group 18.5 months) |
| V4 | RCT |
| V5 | Category 2b |
| V6 | fESWT |
| V7 | Standard care (gauze dressing saturated in 0.9% sodium chloride and elastic bandages) |
| V8 | 15 |
| V9 | 18 |
| V10 | Piezowave with F10G4 applicator (Richard Wolf, Knittlingen, Germany) |
| V11 | Focused, piezoelectric ESWT |
| V12 | 0.173 mJ/mm^2^ |
| V13 | Not specified |
| V14 | 100 ESWs/cm^2^ |
| V15 | 100 ESWs/cm^2^ |
| V16 | 5 Hz |
| V17 | 6 sessions |
| V18 | 5 days |
| V19 | Wound relative change in the ulcer surface area and perimeter after 4 weeks of treatment |
| V20 | 4 weeks |
| V21 | WHR; no biopsies or tissue samples analyzed |
| V22 | Weekly (W1, W2, W3, W4) |
| V23 | Yes |
| V24 | Estimated sample size 60 patients (p = 0.05, power 0.8), related to 4 different groups |
| V25 | fESWT group: 18% reduction in wound area (median relative percentage change); control group: 16% reduction |
| V26 | WHR 4.3 (4.1 elsewhere) fESWT vs 3.1 SWC mm/week; Table 3 medians favored SC W2/W4; no group differences |
| V27 | Not provided |
| V28 | Not provided |
| V29 | No adverse events were described, but the paper stated that ESWT was generally well tolerated |
| V30 | Yes |
| V31 | No - patients allocated sequentially by order of presentation and recruitment phase, not randomly |
| V32 | No |
| V33 | Yes |
| V34 | No |
| V35 | No |
| V36 | No |
| V37 | Yes |
| V38 | Yes |
| V39 | Yes |
| V40 | Yes |

**[108]	Martínez Herraiz, A.; Sánchez Ibáñez, I.; Cuenca González, C.; Garvín Ocampos, L. Terapia de ondas focales de choque para úlcera plantar. *Angiología* 2024, 76(5), 339–342. https://doi.org/10.20960/angiologia.00649.**

**Hypothesis:** This case report tested whether piezoelectric fESWT, added to standard wound care, promotes re-epithelialization and closure of a diabetic plantar (malperforans) ulcer complicated by osteomyelitis and improves ulcer-related quality of life and pain.

**Methods:** Case report of a 70-year-old male with insulin-dependent type II diabetes mellitus of nine years' duration, peripheral vascular disease and previous right forefoot amputations, referred for a plantar ulcer of several weeks' duration; magnetic resonance imaging (MRI) showed osteomyelitis and a fistula. The affected area was 1 cm^2^. Six sessions of focused ESWs were given with a PiezoWave F10G4 device (Richard Wolf, Knittlingen, Germany) at 15-day intervals, giving 370, 390, 360, 360, 350 and 350 impulses (area in cm^2^ × 10 plus 350), i.e. approximately 10 ESWs/cm^2^ plus 350. EFD was not reported; Table I gives only "intensity" 2 and frequency 4, without units.

**Results:** Ulcer area in Table I was 1.5, 2, 1, 1, 0 and 0 cm^2^ at sessions one to six, i.e. complete closure. At four weeks the Charing Cross Venous Ulcer Questionnaire score fell from 90/100 to 10/100, the visual analogue scale pain score from 7/10 to 1/10 and DN4 from 7/10 to 1/10. Control MRI at two months showed disappearance of the fistula and resolution of the osteomyelitis of the proximal first and second metatarsals, with a persisting minimal cuneiform focus. No adverse events were reported and no statistical analysis was performed.

**Conclusions:** The report supported piezoelectric fESWT as a well-tolerated adjunct in the multidisciplinary management of diabetic plantar ulcers. Because a single patient was treated concurrently with SWC, offloading orthoses and previous revascularization, without a control group, defined primary endpoint or statistical analysis, no causal attribution was possible; controlled studies are required.

| **V** | **Result** |
| --- | --- |
| V1 | Diabetic plantar (malperforans) ulcer of the right foot, with underlying osteomyelitis and fistula |
| V2 | 1 cm^2^ initially; Table I: 1.5 cm^2^ first session, 2 cm^2^ second session |
| V3 | Several weeks ("de semanas de evolución"); recurrent right forefoot ulceration since 2014 |
| V4 | Case report |
| V5 | Category 5 |
| V6 | Focused, piezoelectric ESWT in addition to SWC (nurse-led dressings) and offloading plantar orthoses |
| V7 | Not applicable |
| V8 | 1 |
| V9 | Not applicable |
| V10 | PiezoWave F10G4 (Richard Wolf, Knittlingen, Germany) |
| V11 | Focused, piezoelectric ESWT |
| V12 | Not provided; Table I reported only an "intensity" of 2 per session, without units |
| V13 | Not specified |
| V14 | 370, 390, 360, 360, 350, 350 ESWs (sessions 1–6; area cm^2^ × 10 plus 350) |
| V15 | Approximately 10 ESWs/cm^2^ plus a fixed 350 ESWs per session; dose per cm^2^ alone not provided |
| V16 | Not provided; Table I lists a frequency of 4 per session, without units |
| V17 | 6 |
| V18 | 15 days |
| V19 | Not provided (no formally defined primary endpoint; clinical resolution of the ulcer, with the ulcer area recorded at each session) |
| V20 | At each of the 6 sessions; ulcer area 0 cm^2^ from the fifth session (approximately 2 months) |
| V21 | Quality of life (Charing Cross Venous Ulcer Questionnaire), pain (visual analogue scale), DN4, control MRI; no biopsies |
| V22 | 4 weeks post-baseline (questionnaires and pain scales); 2 months post-baseline (MRI) |
| V23 | Not applicable |
| V24 | Not applicable |
| V25 | Complete closure; ulcer area 1.5, 2, 1, 1, 0, 0 cm^2^ at sessions 1–6 (no statistical analysis) |
| V26 | CCVUQ 90/100→10/100, VAS 7/10→1/10, DN4 7/10→1/10 (4 weeks); MRI 2 months: fistula resolved, minimal cuneiform focus persisting |
| V27 | Not applicable |
| V28 | Not applicable |
| V29 | Not reported (no treatment-related adverse events were described) |
| V30 | Yes |
| V31 | No |
| V32 | No |
| V33 | Not applicable |
| V34 | No |
| V35 | No |
| V36 | No |
| V37 | Yes |
| V38 | Yes |
| V39 | Not applicable |
| V40 | Yes |

**[27]	Hitchman, L.; Lathan, R.; Ravindhran, B.; Sidapra, M.; Long, J.; Cowling, A.; Keding, A.; Watson, J.; Iglesias, C.; Smith, G.; Twiddy, M.; Russell, D.; Chetter, I.C. Extracorporeal shockwave therapy for diabetes related foot ulcers: A pilot three-arm double-blinded randomised controlled trial. *Int. Wound J.* 2025, 22, e70740. https://doi.org/10.1111/iwj.70740.**

**Note: This was a pilot RCT following the feasibility case series of Hitchman et al. (2023) [107]; no between-group statistical testing was performed.**

**Hypothesis:** This study tested whether a multi-center RCT of fESWT for DFUs is deliverable, and clinically that fESWT accelerates healing versus sham dose-dependently.

**Methods:** Single-center, pilot, three-arm, double-blinded RCT, follow-up 24 weeks. 74 patients with a DFU present ≥ 4 weeks received standard DFU care plus high-dose fESWT (500 ESWs/cm^2^, n=25), low-dose fESWT (100 ESWs/cm^2^, n=23) or sham fESWT (0 EFDs/cm^2^, n=26). fESWT was delivered with a PiezoWave2 device (Richard Wolf, Knittlingen, Germany) at 0.1 mJ/mm^2^ and 5 Hz in three sessions over 7 +/- 2 days; the primary endpoint was trial deliverability.

**Results:** Of 904 screened patients, 141 (15.6%) were eligible, 74 (52.5%) recruited; treatment adherence 94.6% (70/74); follow-up attendance 97.3%, 93.2%, 87.8% at 6, 12, 24 weeks (65/74 abstract, 64/74 results text); 6-month attrition 12.2%. DFUs healed by 24 weeks in 64.0% (16/25) high-dose, 56.5% (13/23) low-dose and 42.3% (11/26) sham; median time to healing 54.0, 78.5 and 83.0 days; no p-values were calculated. Side effects were mild, mostly tingling, more frequent in the sham arm; no serious adverse event was ESWT-related; major amputation in 2 (8.0%), 1 (4.3%) and 1 (3.8%) participants; no deaths.

**Conclusions:** The definitive trial was concluded deliverable, adherence high and attrition well below the anticipated 30%; eligibility, follow-up attendance and questionnaire completeness require modification. The clinical ordering favored fESWT but was exploratory: the trial was not powered for efficacy, no between-group comparisons were performed, and the arms differed at baseline in age, ulcer duration, size and etiology. The trial supported a fully powered multi-center RCT but did not establish that fESWT accelerates healing.

| **V** | **Result** |
| --- | --- |
| V1 | DFUs (surgical secondary-intention wounds: 5 high dose, 3 low dose, 1 sham) |
| V2 | Mean DFU area 9.5 ± 14.8, 6.6 ± 12.1, 4.6 ± 5.5 cm^2^ (high/low/sham); overall 6.9 ± 11.5 cm^2^; mean depth 0.4 ± 0.6, 0.2 ± 0.2, 0.3 ± 0.2 cm |
| V3 | Chronic; DFU ≥ 4 weeks required. Mean DFU age 133.5 ± 167.5, 239.7 ± 411.6, 180.6 ± 184.2 days (high/low/sham) |
| V4 | RCT (single-center, pilot, three-arm, double-blinded) |
| V5 | Category 2b |
| V6 | fESWT plus standard DFU care, in two dose arms: high dose 500 ESWs/cm^2^ and low dose 100 ESWs/cm^2^ |
| V7 | Sham ESWT plus standard DFU care: 0 ESWs/cm^2^ |
| V8 | 48 (25 high dose, 23 low dose) |
| V9 | 26 |
| V10 | PiezoWave2 (Richard Wolf, Knittlingen, Germany) |
| V11 | Focused, piezoelectric ESWT |
| V12 | 0.1 mJ/mm^2^ (high dose and low dose arms). |
| V13 | Not specified |
| V14 | Not reported per session; prescribed per wound area, minimum 500 ESWs (high dose), 100 (low dose), 0 (sham) |
| V15 | 500 ESWs/cm^2^ (high dose); 100 ESWs/cm^²^ (low dose) |
| V16 | 5 Hz (both active arms) |
| V17 | 3 sessions (all three arms) |
| V18 | Three sessions over a 7 ± 2 day period; the interval between individual sessions was not reported |
| V19 | Trial deliverability: eligibility, recruitment, treatment and follow-up adherence, missing data; green/amber/red criteria (> 60%, 30–60%, < 30%) |
| V20 | Assessed at each follow-up point and overall, i.e., across the 18-month recruitment period and the 24-week follow-up |
| V21 | Healing time, proportion healed, DFU size, QoL (EQ-5D-5L, DFS-SF, Wound-QoL-14), adverse events, resource use; no biopsies |
| V22 | After the third ESWT session and at 6, 12 and 24 weeks post-randomization; healing recorded whenever it occurred |
| V23 | No power analysis; target size set to estimate standard deviation for a definitive trial, allowing 30% attrition |
| V24 | Target 90 participants; 74 (82.2% of target) were actually recruited |
| V25 | 904 screened, 141 (15.6%) eligible, 74/141 (52.5%) recruited; adherence 94.6% (70/74); follow-up 97.3%/93.2%/87.8% at 6/12/24 weeks; attrition 12.2% |
| V26 | Healing at 24 weeks 64.0% (16/25), 56.5% (13/23), 42.3% (11/26) high/low/sham; median time 54.0/78.5/83.0 days; no p-values |
| V27 | Not provided |
| V28 | Not provided |
| V29 | Mild: tingling, pain, discomfort; more in sham; no ESWT-related serious AE; major amputation 2/1/1 (8.0%/4.3%/3.8%); no deaths |
| V30 | Yes |
| V31 | Yes |
| V32 | Yes |
| V33 | No |
| V34 | Yes |
| V35 | No |
| V36 | Yes |
| V37 | Yes |
| V38 | Yes |
| V39 | No |
| V40 | Yes |

**[109]	Crevenna, R.; Vass, Z.; Mickel, M.; Leutner, M.; Li, S.; Keilani, M. Extracorporeal shockwave therapy as a key component of interdisciplinary wound management *in diabetic foot syndrome: A case report. Wien. Med. Wochenschr.* 2026, ePub ahead of print. https://doi.org/10.1007/s10354-026-01184-1.**

**Hypothesis:** This case report tested whether fESWT, an adjunctive regenerative therapy within a multimodal, interdisciplinary conservative concept, can act as a "therapeutic enabler" in diabetic foot syndrome complicated by osteomyelitis and restart healing in a deep, stagnant wound.

**Methods:** Case report of a 57-year-old male with a first manifestation of severe type II diabetes mellitus and diabetic foot syndrome, with a wound of the right first toe (digitus I pedis), osteomyelitis and bony sequestrum. Concurrent care was multimodal (wound care, antibiotics, metabolic stabilization, diabetes management, offloading, infection control). Five fESWT sessions, usually weekly, were applied peri-ulcerously without local anesthesia using a PiezoWave2 device (Richard Wolf, Knittlingen, Germany), at approximately 0.08–0.25 mJ/mm^2^, 1000–2000 ESWs per session and 3–5 Hz following ISMST-oriented protocols; these are protocol ranges, not the settings used.

**Results:** After five fESWT sessions, local wound management extracted a relevant bony sequestrum, leaving a cavity approximately the size of a phalanx. Within only four days, rapid and substantial granulation of the deep cavity was observed, after approximately five weeks of largely stagnant wound healing. Hemoglobin A1c improved from pathological to normal under concurrent diabetes management. No wound measurements, healing times, pain or quality-of-life data, adverse events or statistical analysis were reported.

**Conclusions:** Focused ESWT may be a clinically relevant adjunctive option in diabetic foot syndrome with complicated wound healing, with a striking temporal association between fESWT and pronounced granulation. The authors were explicit that causal conclusions cannot be derived from a single case report and that fESWT is additive, not a replacement for established wound care. Because of simultaneous antibiotic therapy, sequestrectomy and metabolic correction, the independent contribution of fESWT cannot be determined; prospective randomized controlled trials are called for.

| **V** | **Result** |
| --- | --- |
| V1 | Diabetic foot syndrome; wound of the right first toe (digitus I pedis) with osteomyelitis and bony sequestrum |
| V2 | Not provided (residual cavity after sequestrum removal approximately the size of an entire phalanx) |
| V3 | Not explicitly provided; approximately 5 weeks of largely stagnant wound healing preceded the observed response |
| V4 | Case report |
| V5 | Category 5 |
| V6 | Focused piezoelectric ESWT plus multimodal conservative care (wound care, antibiotics, metabolic stabilization, diabetes management, offloading, infection control) |
| V7 | Not applicable |
| V8 | 1 |
| V9 | Not applicable |
| V10 | PiezoWave2 (Richard Wolf, Knittlingen, Germany) |
| V11 | Focused, piezoelectric ESWT |
| V12 | Approximately 0.08–0.25 mJ/mm^2^ |
| V13 | Not specified |
| V14 | 1000–2000 ESWs per session (protocol range; the number actually applied was not reported) |
| V15 | Not provided |
| V16 | 3–5 Hz (protocol range) |
| V17 | 5 |
| V18 | Usually 1 week |
| V19 | Not provided (no formally defined primary endpoint; the outcomes described were sequestrum extraction and granulation of the residual wound cavity) |
| V20 | After 5 fESWT sessions (sequestrum extraction) and 4 days after sequestrum removal (granulation) |
| V21 | None formally defined; the hemoglobin A1c level was reported qualitatively; no biopsies or tissue samples analyzed |
| V22 | Not provided |
| V23 | Not applicable |
| V24 | Not applicable |
| V25 | After 5 fESWT sessions: sequestrum extracted; rapid, substantial granulation of cavity within 4 days after 5 weeks' stagnation |
| V26 | Hemoglobin A1c improved from pathological to normal values (no numerical values, no statistical analysis) |
| V27 | Not applicable |
| V28 | Not applicable |
| V29 | Not reported (no treatment-related adverse events were described) |
| V30 | Yes |
| V31 | No |
| V32 | No |
| V33 | Not applicable |
| V34 | No |
| V35 | No |
| V36 | No |
| V37 | Yes |
| V38 | Yes |
| V39 | Not applicable |
| V40 | No |
