## Supplementary File 5 for "Extracorporeal Shock Wave Therapy for Wound Management: Clinical Evidence, Energy Delivery Parameters and Mechanistic Insights — A Systematic Review"

**Extracorporeal Shock Wave Therapy in Wound Management: A Comprehensive Systematic Review of Clinical Evidence, Modalities and Mechanisms**

by Brent Musolf, Carmen Nussbaum-Krammer, Michael O’Neal, Nicola Maffulli and Christoph Schmitz

**Supplementary File 5**

**Standardized summaries and 40 key variables (c.f. Table 2 in the main text) from clinical studies on radial ESWT for wound management**

**(numbers in brackets refer to the reference numbers in the main text)**

**Note on the standardized summaries:** Each summary condenses one clinical study as it was reported, together with the interpretation that its own authors placed on their findings. The summaries therefore reproduce the position of the respective authors and the state of the field at the time of that publication, and not the assessment of the present systematic review. They follow a uniform structure (Hypothesis, Methods, Results and Conclusions), because the original abstracts differ widely in structure, in length and in the information they report, which makes direct comparison between publications difficult; the standardized form is intended to remove this obstacle.

Abbreviations (in alphabetical order): CABG, coronary artery bypass grafting; DFUs, diabetic foot ulcers; EFD, energy flux density; eNOS, endothelial nitric oxide synthase; ESWT, extracorporeal shock wave therapy; ESWs, extracorporeal shock waves; fESWT, focused ESWT; HBOT, hyperbaric oxygen therapy; LDI, laser Doppler imaging; PCNA; proliferating cell nuclear antigen; RCT, randomized controlled trial; SWC, standard wound care; TcPO₂, transcutaneous partial oxygen pressure; V, variable; VEGF, vascular endothelial growth factor; VLUs, venous leg ulcers.

**[110]	Nossair, A.A.; Eid, M.M.; Salama, A.B. Advanced protocol of shock wave therapy for diabetic foot ulcer. *J. Am. Sci.* 2013, *9*, 633–638. https://doi.org/10.7537/marsjas090413.70.**

**Hypothesis:** This study tested the hypothesis that rESWT enhances wound healing in DFUs by improving epithelialization and reducing wound surface area compared to SWC.

**Methods:** A RCT was conducted with 40 diabetic patients who had stage II or III lower limb DFUs. The patients were randomly divided into two groups (ESWT and control). The ESWT group received three treatment sessions (one treatment session per week) of rESWT using a BTL-5000 SWT device (BTL, Prague, Czech Republic), with 500 ESWs/cm^2^ of wound area delivered at EFD of 0.1 mJ/mm^2^, in addition to medications and the same standard wound care that the control group received. The control group received SWC, including debridement and infection management. The primary outcome was the reduction in wound surface area (WSA); the secondary outcome was the rate of epithelialization. Both outcomes were assessed at baseline and 12 weeks after the start of treatment.

**Results:** The ESWT group showed a significant reduction in wound surface area, from 8.9 ± 3.4 cm^2^ at baseline to 1.9 ± 3.3 cm^2^ after 12 weeks, compared to the control group's reduction from 8.3 ± 3.9 cm^2^ to 4.7 ± 3.4 cm^2^ (p < 0.01). Additionally, the epithelialization rate in the ESWT group increased significantly to 83.3 ± 27.4%, while the control group showed a much lower epithelialization rate of 48.7 ± 31.7% (p < 0.001). These results indicate that rESWT significantly accelerated wound healing compared to SWC.

**Conclusions:** The results of this study suggest that rESWT is an effective and safe modality for accelerating the healing of DFUs. It significantly reduced wound surface area and increased the epithelialization rate compared to SWC. Shock wave therapy could provide a valuable adjunct treatment in managing DFUs and improving patient outcomes.

| **V** | **Result** |
| --- | --- |
| V1 | DFUs |
| V2 | ESWT group: 8.9±3.4 cm^2^, control group: 8.3±3.9 cm^2^ |
| V3 | Chronic (DFUs present for at least 3 months) |
| V4 | RCT |
| V5 | Category 1 |
| V6 | rESWT |
| V7 | SWC (debridement, infection treatment) |
| V8 | 20 |
| V9 | 20 |
| V10 | BTL-5000 SWT (BTL, Prague, Czech Republic) |
| V11 | rESWT |
| V12 | 0.1 mJ/mm^2^ |
| V13 | Not measured; for the radial BTL-5000 SWT the quoted 0.1 mJ/mm^2^ "flux density" was taken from an earlier publication rather than determined for this device; neither an air pressure nor a measurement basis was reported |
| V14 | 500 ESWs/cm^2^ of wound area |
| V15 | 500 ESWs/cm^2^ |
| V16 | Not provided |
| V17 | 3 sessions |
| V18 | 1 week between each session |
| V19 | WSA and epithelialization rate |
| V20 | 12 weeks post-treatment |
| V21 | Epithelialization rate; no biopsies or tissue samples analyzed |
| V22 | 12 weeks post-treatment |
| V23 | Not provided |
| V24 | Not applicable |
| V25 | ESWT group: 1.9 ± 3.3 cm^2^ (WSA), control group: 4.7 ± 3.4 cm^2^ (WSA), p < 0.01 |
| V26 | ESWT group: 83.3 ± 27.4% epithelialization rate, control group: 48.7 ± 31.7%, p < 0.001 |
| V27 | Not provided |
| V28 | Not applicable |
| V29 | Not provided |
| V30 | Yes |
| V31 | Yes |
| V32 | Not provided |
| V33 | Yes |
| V34 | No |
| V35 | No |
| V36 | No |
| V37 | Yes |
| V38 | Yes |
| V39 | Yes |
| V40 | Yes |

**[111]	Fekete, L.; Nagy, G.Á.; Diamant, P.K.; Halmy, C.; Zentai, A. A radiális lökéshullám-kezelés szerepe a nagyméretű ulcus cruris gyógyításában. *Orv. Hetil.* 2014, *155*(45), 1794–1799. https://doi.org/10.1556/OH.2014.30018.**

**Hypothesis:** This study tested the hypothesis that rESWT is effective in chronic VLUs, assessing its impact on wound size reduction and readiness for surgical intervention.

**Methods:** This case report involved a single patient with a chronic VLU caused by venous circulatory failure for five years. The ulcer, measuring 25×18 cm^2^, was treated with rESWT twice weekly for 10 weeks, using a Swiss DolorClast device (Electro Medical Systems, Nyon, Switzerland), with an EFD of 0.08 mJ/mm^2^ and 1000 ESWs/cm^2^ of wound area, in addition to wound cleansing, necrectomy/keratotomy as required and dressings adapted to the actual phase of wound healing. No control group was utilized. The primary outcome was the reduction in ulcer size; the secondary outcome, though not specifically defined, included clinical signs of healing such as granulation tissue formation and readiness for surgical closure. The primary endpoint was assessed at 10 weeks post-baseline, just before surgery for skin grafting.

**Results:** At 10 weeks post-baseline, the ulcer had substantially decreased from 25×18 cm^2^ to a size suitable for surgical intervention. The wound showed substantial granulation tissue formation; inflammatory signs such as edema and erythema were notably reduced, and the patient was deemed ready for skin grafting. The paper did not report adverse events or tolerability; at the time of publication the patient was asymptomatic and had healed well post-surgery.

**Conclusions:** rESWT was effective in reducing the size of a chronic VLU and preparing the wound for surgical closure, with a favorable impact on the clinical progression of wound healing, although adverse events and tolerability were not reported. This case supports the potential use of rESWT as a supplementary treatment for chronic VLUs, especially when conservative treatment alone fails to achieve adequate healing.

| **V** | **Result** |
| --- | --- |
| V1 | VLU |
| V2 | 25×18 cm^2^ |
| V3 | Chronic (5 years) |
| V4 | Case report |
| V5 | Category 5 |
| V6 | Radial ESWT |
| V7 | Not applicable |
| V8 | 1 |
| V9 | Not applicable |
| V10 | Swiss DolorClast (Electro Medical Systems, Nyon, Switzerland) |
| V11 | Radial ESWT |
| V12 | 0.08 mJ/mm^2^ |
| V13 | Not specified; radial Swiss DolorClast, 0.08 mJ/mm^2^ quoted as "energy density", no air pressure or measurement basis |
| V14 | 1000 ESWs/cm^2^ of wound area |
| V15 | 1000 ESWs/cm^2^ |
| V16 | Not provided |
| V17 | 20 sessions (2 times per week for 10 weeks) |
| V18 | Twice per week |
| V19 | Not clearly defined, but wound size reduction and healing readiness for surgery were primary measures |
| V20 | 10 weeks post-baseline (final assessment before surgery) |
| V21 | None explicitly defined or assessed; no biopsies or tissue samples analyzed |
| V22 | Not applicable |
| V23 | No |
| V24 | Not applicable |
| V25 | The VLU significantly decreased in size in the rESWT-treated patient |
| V26 | Not applicable |
| V27 | No |
| V28 | Not applicable |
| V29 | Not provided |
| V30 | Yes |
| V31 | No |
| V32 | No |
| V33 | Not applicable |
| V34 | No |
| V35 | No |
| V36 | No |
| V37 | Yes |
| V38 | Yes |
| V39 | No |
| V40 | No |

**[106]	Dolibog, P.; Dolibog, P.; Franek, A.; Brzezińska-Wcisło, L.; Arasiewicz, H.; Wróbel, B.; Chmielewska, D.; Ziaja, J.; Błaszczak, E. Randomized, controlled clinical pilot study of venous leg ulcers treated with using two types of shockwave therapy. *Int. J. Med. Sci.* 2018, *15*(12), 1275–1285. https://doi.org/10.7150/ijms.26614.**

**Note: Dolibog et al. (2018) investigated both piezoelectric fESWT and rESWT; only the rESWT results are presented here.**

**Hypothesis:** This study tested the hypothesis that rESWT heals VLUs more effectively than SWC, with faster reduction in ulcer size and a higher percentage of complete healing.

**Methods:** This RCT comparison involved 35 patients: 17 in the rESWT group and 18 in the SWC control group (the whole trial analyzed 50 patients in three groups; 57 were randomized and 7 did not complete treatment). The rESWT group received 6 sessions at 5-day intervals with a ShockMaster 500 and classic 15 mm applicator (Gymna Uniphy, Bilzen-Hoeselt, Belgium), 100 ESWs/cm^2^ at 5 Hz and 0.2 MPa air pressure (manufacturer-specified nominal EFD 0.17 mJ/mm^2^), plus the same standard care as controls (wet saline gauze dressings, gently compressing elastic bandages). The primary endpoint was change in total ulcer surface area; secondary outcomes were percentage change in ulcer dimensions (length, width, circumference), the Gilman index and the number of completely healed ulcers, measured before treatment and 4 weeks after the last session.

**Results:** The rESWT group showed a statistically significant reduction in ulcer size, averaging 67.7% versus 38.9% in controls (p < 0.05). A complete cure was achieved in 35% of the rESWT patients; the paper reported the corresponding control rate only graphically (Figure 3) and gave no statistical test for this comparison. The rESWT group also showed greater improvements in maximum ulcer length and width and in the Gilman index, although not all reached statistical significance.

**Conclusions:** rESWT significantly improved VLU healing compared to SWC, with a more pronounced reduction in ulcer surface area and a higher percentage of complete healing. Further studies with larger samples and longer follow-up are needed to confirm these findings and the long-term benefits of rESWT.

| **V** | **Result** |
| --- | --- |
| V1 | VLUs |
| V2 | 5.8 ± 7.9 cm^2^ (rESWT group); 8.3 ± 4.6 cm^2^ (control group) |
| V3 | Chronic; duration 2–24 months (rESWT), 1–48 months (control) |
| V4 | RCT |
| V5 | Category 1 |
| V6 | rESWT |
| V7 | SWC (saline dressings and compression bandages) |
| V8 | 17 |
| V9 | 18 |
| V10 | ShockMaster 500 (Gymna Uniphy, Bilzen-Hoeselt, Belgium) |
| V11 | rESWT |
| V12 | 0.17 mJ/mm^2^ |
| V13 | Not measured; manufacturer's nominal 0.17 mJ/mm^2^ for 0.2 MPa (2 bar), 15 mm applicator, radial ShockMaster 500 |
| V14 | 100 ESWs/cm^2^ |
| V15 | 100 ESWs/cm^2^ |
| V16 | 5 Hz |
| V17 | 6 sessions |
| V18 | Every 5 days |
| V19 | Change in total ulcer surface area and linear dimensions inside groups |
| V20 | 4 weeks after last treatment (i.e., ~8 weeks total) |
| V21 | Number of completely healed wounds, Gilman index, percentage change of ulcer area; no biopsies or tissue samples analyzed |
| V22 | 4 weeks after last treatment (same as primary endpoint) |
| V23 | Not provided |
| V24 | Not provided |
| V25 | Ulcer area reduced by 67.7% (rESWT) vs 38.9% (control); p < 0.05 |
| V26 | Healed: 35% (rESWT) versus a control rate that the paper reports only graphically (Figure 3); Gilman 8wk 0.75 ± 1.07/0.31 ± 0.22 cm p = 0.447; area 67.7 ± 30.5/38.9 ± 27.1% p = 0.023; length 58.2 ± 35.9/24.6 ± 21.5% p = 0.029; width 59.6 ± 34.5/30.6 ± 18.6% p = 0.041 |
| V27 | Not provided |
| V28 | Not provided |
| V29 | Not provided |
| V30 | Yes |
| V31 | Yes |
| V32 | No |
| V33 | Yes |
| V34 | No |
| V35 | No |
| V36 | No |
| V37 | Yes |
| V38 | Yes |
| V39 | Yes |
| V40 | Yes |

**[112]	Dolibog, P.; Dolibog, P.T.; Franek, A.; Brzezińska-Wcisło, L.; Wróbel, B.; Arasiewicz, H.; Chmielewska, D.; Ziaja, J.; Błaszczak, E. Comparison of ultrasound therapy and rESWT in the treatment of venous leg ulcers—Clinical, pilot study. *Postepy Dermatol. Alergol.* 2018, *35*(5), 454–461. https://doi.org/10.5114/ada.2018.79191.**

**Hypothesis:** This study tested the hypothesis that rESWT promotes VLU healing more effectively than SWC and is comparably effective to ultrasound therapy.

**Methods:** This RCT involved 51 patients (17 in each group) with VLUs of varying durations (1–31 months), assigned to rESWT, ultrasound therapy or a control group receiving SWC. The rESWT group received 6 sessions at 5-day intervals over 4 weeks, each delivering 100 ESWs/cm^2^ at a frequency of 5 Hz and an air pressure of 0.2 MPa, corresponding to a manufacturer-specified nominal EFD of 0.17 mJ/mm^2^ (classic 15 mm applicator), using a ShockMaster 500 device (Gymna Uniphy, Bilzen-Hoeselt, Belgium); these patients additionally received the same standard care as controls (gauze dressing saturated in 0.9% sodium chloride and elastic bandages changed daily). The primary outcome was the reduction in ulcer area and the secondary outcome the Gilman index, a linear parameter of wound healing, both measured at baseline and 4 weeks post-baseline.

**Results:** After 4 weeks, ultrasound therapy achieved the greatest reduction in ulcer area (mean 67.6%), followed by rESWT (38.2%) and the control group (15.8%). The Gilman index also favored ultrasound therapy (0.49 cm), followed by rESWT (0.24 cm) and the control group (0.13 cm). Both ultrasound and rESWT were more effective than SWC in healing VLUs, with ultrasound therapy the most effective.

**Conclusions:** rESWT was effective in reducing the area of VLUs, though less effective than ultrasound therapy. Both treatments were significantly more effective than SWC, which showed minimal improvement. The study supports rESWT as a potential treatment for VLUs, though further research is needed to confirm its long-term effectiveness and compare it with other therapies.

| **V** | **Result** |
| --- | --- |
| V1 | VLUs |
| V2 | Ultrasound therapy: 9.8 cm^2^, radial ESWT: 9.2 cm^2^, control: 11.8 cm^2^ |
| V3 | Ultrasound therapy: 1-24 months, radial ESWT: 3-24 months, control: 1-31 months |
| V4 | RCT |
| V5 | Category 4 |
| V6 | Radial ESWT |
| V7 | SWC (Gauze dressing with 0.9% sodium chloride and elastic bandages changed daily) |
| V8 | 17 |
| V9 | 17 |
| V10 | ShockMaster 500 (Gymna Uniphy, Bilzen-Hoeselt, Belgium) |
| V11 | Radial ESWT |
| V12 | 0.17 mJ/mm^2^ |
| V13 | Not measured; manufacturer's nominal 0.17 mJ/mm^2^ for 0.2 MPa (2 bar), 15 mm applicator, radial ShockMaster 500 |
| V14 | 100 ESWs/cm^2^ |
| V15 | 100 ESWs/cm^2^ |
| V16 | 5 Hz |
| V17 | 6 sessions (over 4 weeks) |
| V18 | 5 days |
| V19 | Reduction in ulcer area (cm^2^) |
| V20 | 4 weeks |
| V21 | Gilman index (wound healing rate); no biopsies or tissue samples analyzed |
| V22 | 4 weeks |
| V23 | Not provided |
| V24 | Not applicable |
| V25 | Area reduction: ultrasound 67.6 ± 29.6%, rESWT 38.2 ± 28.3%, control 15.8 ± 12.4%; p(A-B-C)<0.001, p(A-B)<0.05, p(A-C)<0.001, p(B-C)<0.05 |
| V26 | Gilman index: ultrasound 0.49 ± 0.20, rESWT 0.24 ± 0.17, control 0.13 ± 0.01 cm; p(A-B-C)<0.001, p(A-B)<0.05, p(A-C)<0.001, p(B-C)>0.05 (rESWT vs control NOT significant) |
| V27 | Not provided |
| V28 | Not applicable |
| V29 | Not provided |
| V30 | Yes |
| V31 | Yes |
| V32 | No |
| V33 | Yes |
| V34 | No |
| V35 | No |
| V36 | No |
| V37 | Yes |
| V38 | Yes |
| V39 | Yes |
| V40 | Yes |

**[113]	Duan, H.; Li, H.; Liu, H.; Zhang, H.; Liu, N.; Dong, Q.; Li, Z. Extracorporeal shockwave therapy combined with alginate dressing for treatment of sacroiliac decubital necrosis in older adults: A case report. *Medicine* 2020, *99*(19), e19849. https://doi.org/10.1097/MD.0000000000019849.**

**Hypothesis:** This study hypothesized that rESWT combined with alginate dressing effectively promotes the healing of sacroiliac decubital necrosis (PUs) in older adults.

**Methods:** This case report focused on the treatment of a 62-year-old male patient diagnosed with stage IV sacroiliac decubital necrosis, caused by long-term pressure and exacerbated by cognitive impairment following carbon monoxide poisoning. The patient received a combined treatment of rESWT using a Swiss DolorClast device (Electro Medical Systems, Nyon, Switzerland) and alginate dressing over 12 weeks. rESWT was administered once a week, with the patient receiving 12 sessions in total. The therapy parameters included a frequency of 4-5 Hz, air pressure of 2-3 bar and a total of 200-300+100 ESWs/cm^2^ per treatment session. The primary outcome measure was the healing of the pressure ulcer, assessed using the Pressure Ulcer Scale for Healing (PUSH) score at baseline, 4, 8 and 12 weeks as well as at 2 weeks post-discharge.

**Results:** At baseline, the patient’s PUSH score was 17, which indicated severe ulceration. After 12 weeks of therapy, the PUSH score decreased to 5, and the ulcer was considered healed (PUSH score = 0) 2 weeks post-discharge. No adverse reactions or side effects were reported during the treatment period.

**Conclusions:** rESWT combined with alginate dressing proved to be an effective treatment for sacroiliac decubital necrosis in this older adult patient, significantly promoting wound healing. The findings suggest that this combined therapy may offer a valuable solution for similar cases of PUs, particularly in elderly patients. However, further studies are necessary to confirm these results in larger populations and with RCTs.

| **V** | **Result** |
| --- | --- |
| V1 | PU (sacroiliac decubital necrosis) |
| V2 | Approximately 7.5 x 5.5 cm (stage IV sacrococcygeal pressure sore) |
| V3 | Approximately 2 weeks at admission; first noticed by family 2 weeks before hospitalization, not effectively treated |
| V4 | Case report |
| V5 | Category 5 |
| V6 | rESWT + alginate dressing; debridement every 2 days; neurorehabilitation 1.5h twice daily 6d/wk 12wk; cognitive therapy 6×/wk 12wk |
| V7 | Not applicable (no control group) |
| V8 | 1 |
| V9 | Not applicable (no control group) |
| V10 | Swiss DolorClast (Electro Medical Systems, Nyon, Switzerland) |
| V11 | rESWT |
| V12 | Not provided (only air pressure of 2-3 bar reported; no energy flux density given) |
| V13 | Not specified |
| V14 | 200/300 + 100 ESWs per cm^2^ |
| V15 | 200/300 + 100 ESWs per cm^2^ |
| V16 | 4-5 Hz |
| V17 | 12 sessions (once a week for 12 weeks) |
| V18 | Once a week |
| V19 | Wound healing as measured by the PUSH score |
| V20 | 4, 8 and 12 weeks, plus 2 weeks post-discharge |
| V21 | Rancho Levels of Cognitive Functioning before treatment and at 4, 8, 12 weeks; no biopsies or tissue samples |
| V22 | Not applicable |
| V23 | No |
| V24 | Not applicable |
| V25 | PUSH score: 17 baseline, 13 (4 weeks), 9 (8 weeks), 5 (12 weeks), 0/healed 2 weeks post-discharge |
| V26 | Not applicable |
| V27 | No |
| V28 | Not applicable |
| V29 | No adverse events or side effects were reported |
| V30 | Yes |
| V31 | No |
| V32 | No |
| V33 | Not applicable |
| V34 | No |
| V35 | No |
| V36 | No |
| V37 | Yes |
| V38 | Yes |
| V39 | Not applicable |
| V40 | No |

**[114]	Kang, N.; Yu, X.; Ma, Y. Radial extracorporeal shock wave therapy in a patient with decubitus ulcer after spinal cord injury: A case report. *Am. J. Transl. Res.* 2020, *12*(5), 2093–2098.**

**Hypothesis:** This study tested the hypothesis that rESWT promotes the healing of decubitus ulcers in a patient with spinal cord injury (SCI), as an alternative to invasive procedures such as skin flap transplantation.

**Methods:** This case report describes a 51-year-old male with a decubitus ulcer on the right heel following a spinal cord injury caused by a lumbar vertebral fracture. The ulcer had a surface area of 4.8 × 4.5 cm^2^ and a depth of 2 cm. rESWT was delivered with a radial ESWT device from Storz Medical (Tägerwilen, Switzerland); model not named, an R15 transmitter (15 mm) was used first and a DEEP transmitter (15 mm) from week 3. No control group. Treatment was given once or twice a week for three months (a single session in the first week, twice weekly thereafter) at 2.0 to 3.5 bar (no energy flux density reported), with 3000 to 6000 ESWs per session at 10 Hz. The primary outcome was complete healing of the ulcer, assessed three months after baseline; secondary outcomes were not specified.

**Results:** The decubitus ulcer exhibited substantial healing. After three months of treatment, it was completely healed with no infection or complications, with a reduction in wound size and improvement in the surrounding tissue. The paper made no statement about tolerability or adverse events; it reported only that the wound did not become infected

**Conclusions:** This case report suggests rESWT can be an effective and non-invasive treatment for promoting the healing of decubitus ulcers in patients with spinal cord injury, and may provide a viable alternative to invasive surgical procedures like skin flap transplantation, especially in patients with poor circulation and nutritional status. Further research with larger sample sizes and control groups is needed to confirm the generalizability of these findings.

| **V** | **Result** |
| --- | --- |
| V1 | Decubitus ulcer |
| V2 | 4.8 × 4.5 cm^2^ |
| V3 | 21 days |
| V4 | Case report |
| V5 | Category 5 |
| V6 | rESWT after surgical debridement, combined with intermittent ultra-short-wave therapy |
| V7 | Not applicable |
| V8 | 1 |
| V9 | Not applicable |
| V10 | rESWT, Storz Medical (Tägerwilen, Switzerland); model unnamed; R15 then DEEP transmitter (15 mm) from week 3 |
| V11 | Radial ESWT |
| V12 | Not provided |
| V13 | Not specified |
| V14 | 3000–6000 ESWs per session |
| V15 | Not provided |
| V16 | 10 Hz |
| V17 | Once or twice per week for 3 months (1 session in week 1, then 2 sessions per week) |
| V18 | Once or twice per week |
| V19 | Complete healing of the decubitus ulcer |
| V20 | 3 months |
| V21 | None explicitly defined or assessed; no biopsies or tissue samples analyzed |
| V22 | Not applicable |
| V23 | No |
| V24 | Not applicable |
| V25 | Complete wound healing after 3 months of treatment |
| V26 | Not applicable |
| V27 | No |
| V28 | Not applicable |
| V29 | Not provided |
| V30 | Yes |
| V31 | No |
| V32 | No |
| V33 | Not applicable |
| V34 | No |
| V35 | No |
| V36 | No |
| V37 | Yes |
| V38 | Yes |
| V39 | Not applicable |
| V40 | No |

**[25]	Fan, Y.; Shen, W.; Chai, C.; Hua, F. Application value of extracorporeal shock wave therapy of patients with recurrent diabetic foot ulcer infection. *Acta Med. Mediterr.* 2022, *38*, 2769–2775.**

**Hypothesis:** This study hypothesized that rESWT improves wound healing and clinical outcomes in patients with recurrent DFUs compared to SWC.

**Methods:** This RCT involved 87 patients with chronic DFUs: 46 received rESWT combined with SWC and 41 received SWC only. rESWT was performed using a Swiss DolorClast device (Electro Medical Systems, Nyon, Switzerland) at an air pressure of 2-4 bar with a quoted energy density of 0.25 mJ/mm^2^, 4000-6000 ESWs per session, a frequency of 9-18 Hz and 4-6 sessions, one per week. The primary outcome was the reduction in wound size and healing time, evaluated at the end of treatment (4-6 weeks). Secondary outcomes included wound exudation, histology, bacterial clearance rate, blood glucose, blood pressure, inflammatory markers and life quality scores, assessed before and after treatment, except life quality scores, assessed at the 6-month follow-up.

**Results:** The rESWT group showed significantly better clinical outcomes than the control group, with a higher wound reduction rate and faster healing time (p < 0.05). Secondary outcomes, including bacterial clearance, wound exudation, histology scores and clinical efficacy, were also significantly improved. The incidence of adverse reactions (bleeding, pain, reinfection) was lower in the rESWT group (8.7%) than in the control group (24.4%) (p < 0.05). Blood glucose, blood pressure and inflammatory factors also improved more in the rESWT group (p < 0.05), and life quality scores at the 6-month follow-up were significantly higher.

**Conclusions:** rESWT significantly improved the healing of recurrent DFUs, enhanced clinical efficacy and reduced adverse reactions compared to SWC, with a positive impact on both primary and secondary outcomes, including wound healing, bacterial clearance and overall patient well-being. rESWT is a valuable therapeutic option for managing DFUs, with potential to improve patient outcomes and quality of life.

| **V** | **Result** |
| --- | --- |
| V1 | DFUs |
| V2 | rESWT group: 28.3 +/- 3.5 cm^2^; control group: 27.7 +/- 3.6 cm^2^ |
| V3 | Chronic (recurrent DFUs) |
| V4 | Non-randomized controlled (cohort) study |
| V5 | Category 1 |
| V6 | rESWT |
| V7 | SWC |
| V8 | 46 |
| V9 | 41 |
| V10 | Swiss DolorClast Smart (Electro Medical Systems, Nyon, Switzerland) |
| V11 | rESWT |
| V12 | 0.25 mJ/mm^2^ |
| V13 | Not specified |
| V14 | 4000-6000 ESWs |
| V15 | Not provided |
| V16 | 9-18 Hz |
| V17 | 4-6 |
| V18 | 1 session per week |
| V19 | Wound healing time, wound reduction rate, exudation, histology, bacterial clearance rate and clinical efficacy |
| V20 | After 4-6 weeks of treatment |
| V21 | Blood glucose, blood pressure, hs-CRP, WBC, transcutaneous oxygen partial pressure, life quality scores; no biopsies or histological analysis |
| V22 | After treatment for blood glucose, blood pressure and inflammatory markers; 6-month follow-up for the life quality scores |
| V23 | Not provided |
| V24 | Not provided |
| V25 | rESWT superior to control in wound reduction rate, healing time, bacterial clearance and other clinical parameters (p < 0.05) |
| V26 | rESWT group better than control for blood glucose, blood pressure, inflammation and quality of life (p < 0.05) |
| V27 | Not provided |
| V28 | Not provided |
| V29 | Mild bleeding, pain, redness, reinfection; lower incidence in rESWT (8.7%) than control (24.4%) |
| V30 | Yes |
| V31 | No |
| V32 | No |
| V33 | Yes |
| V34 | No |
| V35 | No |
| V36 | No |
| V37 | Yes |
| V38 | Yes |
| V39 | Yes |
| V40 | Yes |

**[115]	Dymarek, R.; Kuberka, I.; Rosińczuk, J.; Walewicz, K.; Taradaj, J.; Sopel, M. The immediate clinical effects following a single rESWT in pressure ulcers: A preliminary randomized controlled trial of the SHOWN project. *Adv. Wound Care* 2023, *12*(8), 440–452. https://doi.org/10.1089/wound.2021.0015.**

**Hypothesis:** This study tested whether a single session of rESWT promotes immediate clinical improvements in PU healing compared to placebo.

**Methods:** This RCT involved 40 patients aged 61 to 92 years with PUs of chronic etiology, treated in long-term inpatient care and outpatient wound care settings. An experimental group received active rESWT and a control group placebo ESWT. rESWT used a Cellactor device (Storz Medical, Tägerwilen, Switzerland) with 300 + 100 ESWs/cm^2^ in a single session, an air pressure of 2.5 bar with a quoted EFD of 0.15 mJ/mm^2^ and a frequency of 5 Hz. The primary outcome was wound healing assessed by the Wound Bed Score (WBS) and Bates–Jensen Wound Assessment Tool (BWAT); secondary outcomes were not provided. Per the Methods, the final measurement (M1) was taken 1 week after the session, although the title and abstract describe the effects as "immediate"

**Results:** In the rESWT group wound area decreased from 11.5 cm^2^ to 8.1 cm^2^, length from 5.0 cm to 4.4 cm and width from 3.2 cm to 2.5 cm (all p < 0.001). WBS increased from 3.9 to 9.7 points and BWAT improved from 45.5 to 30.7 points (both p < 0.001). The placebo group deteriorated, with increases in wound area (p = 0.002), length (p = 0.16) and width (p = 0.005), no change in WBS and a small, non-significant worsening in BWAT (from 34.1 to 35.4 points, i.e. +1.3 points; p = 0.28).

**Conclusions:** Even a single session of rESWT can produce significant clinical improvements in patients with PUs, with reduced wound size and improved clinical scores. rESWT may be an effective adjunctive therapy for chronic PUs, but further studies with larger sample sizes and longer follow-up are needed to confirm its clinical utility and long-term efficacy.

| **V** | **Result** |
| --- | --- |
| V1 | PUs |
| V2 | rESWT group: 11.5 +/- 9.6 cm^2^; placebo group: 12.3 +/- 12.5 cm^2^ |
| V3 | Chronic, with a mean duration of 5.8 ± 3.3 months for the rESWT group |
| V4 | RCT |
| V5 | Category 1 |
| V6 | rESWT |
| V7 | Placebo rESWT (same device, but with a covering to absorb energy, no therapeutic effect) |
| V8 | 20 |
| V9 | 20 |
| V10 | Cellactor (Storz Medical, Tägerwilen, Switzerland) |
| V11 | rESWT |
| V12 | 0.15 mJ/mm^2^ |
| V13 | Not specified |
| V14 | 300 + 100 ESWs/cm^2^ (300 impulses at baseline plus 100 impulses per cm^2^ of wound area) |
| V15 | 100 ESWs/cm^2^ (plus a fixed 300 baseline impulses per wound) |
| V16 | 5 Hz |
| V17 | 1 session |
| V18 | Not applicable (only 1 session) |
| V19 | Planimetric wound area, length, width; WBS; BWAT |
| V20 | 1 week after the single treatment session (M1) per the Methods; the title and abstract call the effects "immediate" |
| V21 | None explicitly defined or assessed; no biopsies or tissue samples analyzed |
| V22 | Not applicable |
| V23 | No |
| V24 | Not applicable |
| V25 | rESWT: WBS +5.8, BWAT -14.8 points (p < 0.05); placebo: WBS 0.0, BWAT +1.4 |
| V26 | Not provided |
| V27 | No |
| V28 | Not applicable |
| V29 | Not provided |
| V30 | Yes |
| V31 | Yes |
| V32 | No (allocation concealment is not described) |
| V33 | Yes |
| V34 | Yes |
| V35 | No |
| V36 | Yes |
| V37 | Yes |
| V38 | Yes |
| V39 | Yes |
| V40 | Yes |

**[24]	Dolibog, P.T.; Dolibog, P.; Bergler-Czop, B.; Grzegorczyn, S.; Chmielewska, D. The efficacy of extracorporeal shockwave therapy compared with compression therapy in healing venous leg ulcers. *J. Clin. Med.* 2024, *13*(7), 2117. https://doi.org/10.3390/jcm13072117.**

**Note: Dolibog et al. (2024) investigated both piezoelectric fESWT and rESWT; only the rESWT results are presented here.**

**Hypothesis:** This study tested the hypothesis that rESWT significantly improves the healing of VLUs compared to SWC, routinely used for chronic wounds.

**Methods:** This RCT enrolled 69 patients with chronic VLUs, primarily caused by chronic venous insufficiency, allocated to four arms (IPC n = 20, rESWT n = 16, fESWT n = 15, SWC n = 18); the present entry covers the rESWT arm (n = 16) and the SWC control arm (n = 18), i.e. 34 of the 69 patients. rESWT was performed using a ShockMaster 500 device (Gymna Uniphy, Bilzen, Belgium) in 6 sessions every 5 days for 4 weeks, with 100 ESWs/cm^2^ of wound area, a frequency of 5 Hz and EFD of 0.17 mJ/mm^2^. The control group received SWC with daily gauze dressings and elastic bandages. The primary outcome was the relative change in wound surface area after 4 weeks; the secondary outcome was the weekly wound healing rate (WHR), measured each week during treatment.

**Results:** After 4 weeks, the rESWT group showed a median relative wound area reduction of 31.6% versus 16% in the control group. Both groups showed statistically significant reductions from baseline, but the median reduction was notably larger with rESWT. The weekly healing rate was higher in the rESWT group, but the between-group difference was not statistically significant.

**Conclusions:** rESWT improved wound healing in VLUs more effectively than SWC. Although both modalities led to significant improvements, the rESWT group exhibited a larger reduction in wound area. These findings support rESWT as a viable treatment option for chronic VLUs, although additional studies are needed to validate its clinical benefits over standard therapies.

| **V** | **Result** |
| --- | --- |
| V1 | VLUs |
| V2 | Median initial wound area: rESWT group 6.1 cm^2^ (25Q-75Q 2.5-12.3); control (SC) group 10.5 cm^2^ (25Q-75Q 5.8-12.0) |
| V3 | Chronic (Duration of VLUs: rESWT group 10 months, control group 18.5 months) |
| V4 | RCT |
| V5 | Category 2b |
| V6 | rESWT |
| V7 | SWC (gauze dressing saturated in 0.9% sodium chloride and elastic bandages) |
| V8 | 16 |
| V9 | 18 |
| V10 | ShockMaster 500 (Gymna Uniphy, Bilzen, Belgium) |
| V11 | rESWT |
| V12 | 0.17 mJ/mm^2^ |
| V13 | Not specified |
| V14 | 100 ESWs/cm^2^ |
| V15 | 100 ESWs/cm^2^ |
| V16 | 5 Hz |
| V17 | 6 sessions |
| V18 | 5 days |
| V19 | Wound relative change in the ulcer surface area and perimeter after 4 weeks of treatment |
| V20 | 4 weeks |
| V21 | Weekly WHR; red blood cell deformability (elongation index, EI) before and after treatment; no biopsies |
| V22 | Weekly (W1, W2, W3, W4) |
| V23 | Yes |
| V24 | Estimated sample size 60 patients across 4 groups (p = 0.05, power 0.8) |
| V25 | rESWT group: 31.6% reduction in wound area (median relative percentage change); control group: 16% reduction |
| V26 | Weekly healing rate: rESWT > SWC |
| V27 | Not provided |
| V28 | Not provided |
| V29 | No adverse events reported; the paper stated only that "treatment with rESWT was generally well tolerated" |
| V30 | Yes |
| V31 | No |
| V32 | No |
| V33 | Yes |
| V34 | No |
| V35 | No |
| V36 | No |
| V37 | Yes |
| V38 | Yes |
| V39 | Yes |
| V40 | Yes |

**[116]	Dymarek, R.; Kuberka, I.; Walewicz, K.; Taradaj, J.; Rosińczuk, J.; Sopel, M. Is shock wave application effective on various chronic wounds in the geriatric population? Preliminary clinical study. *Clin. Interv. Aging* 2024, *19*, 665–679. https://doi.org/10.2147/CIA.S448298.**

**Hypothesis:** This study tested whether a single session of rESWT significantly improves the healing of chronic wounds in geriatric patients, particularly DFUs, VLUs and PUs.

**Methods:** A prospective uncontrolled interventional study included 19 patients (31 wounds) with chronic wounds: 2 DFUs, 7 VLUs and 22 PUs. rESWT was performed using a Cellactor device (Storz Medical, Tägerwilen, Switzerland) in a single session with 300 baseline pulses + 100 pulses per cm^2^ of wound area, EFD of 0.15 mJ/mm^2^, pressure of 2.5 bar and a frequency of 5 Hz. The primary outcome was improvement in wound size by planimetric assessment; secondary outcomes were clinical improvements assessed with the Wound Bed Score (WBS) and Bates-Jansen Wound Assessment Tool (BWAT), before treatment (M0) and one week post-treatment (M1).

**Results:** After the single session, wound area was significantly reduced from 9.4 ± 9.1 cm^2^ to 6.2 ± 7.1 cm^2^ (p < 0.001), with reductions in wound length and width. WBS increased significantly from 4.6 ± 2.8 to 10.4 ± 3.6 points (a 31.3% improvement) and BWAT decreased from 43.8 ± 7.9 to 30.7 ± 7.5 points (a 20.0% improvement) (p < 0.001). A significant negative correlation was found between wound area and WBS (r = -0.446, p = 0.012) and a positive correlation with BWAT (r = 0.327, p = 0.073). No adverse events were reported; the treatment was well-tolerated.

**Conclusions:** A single session of rESWT can lead to significant short-term improvements in the healing of chronic wounds in older adults, highlighting its potential as an adjunctive therapy, with notable improvements in both wound size and clinical assessment scores. Future studies should include larger samples, a control group and a longer follow-up.

| **V** | **Result** |
| --- | --- |
| V1 | DFUs, VLUs and PUs |
| V2 | Mean area: 9.4 ± 9.1 cm^2^ |
| V3 | Mean duration: 9.1 ± 9.2 months |
| V4 | CS |
| V5 | Category 5 |
| V6 | rESWT |
| V7 | Not applicable |
| V8 | 19 patients with 31 wounds (2 DFUs, 7 VLUs and 22 PUs) |
| V9 | Not applicable |
| V10 | Cellactor (Storz Medical, Tägerwilen, Switzerland) |
| V11 | rESWT |
| V12 | 0.15 mJ/mm^2^ |
| V13 | Not specified |
| V14 | 300 baseline pulses + 100 pulses per cm^2^ of wound area |
| V15 | 100 ESWs/cm^2^ (plus a fixed 300 baseline pulses per wound) |
| V16 | 5 Hz |
| V17 | 1 session |
| V18 | Not applicable |
| V19 | Objective improvement in wound parameters (area, length, width) by digital planimetry |
| V20 | 1 week post-treatment (M1) |
| V21 | Secondary: WBS, BWAT; tertiary: planimetric–clinical correlation, etiology/location subgroups; no biopsies |
| V22 | M0 (immediately before rESWT) and M1 (one week after rESWT) |
| V23 | No |
| V24 | Not applicable |
| V25 | Significant wound decrease and clinical improvement in WBS and BWAT (p < 0.001) |
| V26 | One week post-session: WBS 4.6 ± 2.8 to 10.4 ± 3.6 (+31.3%), BWAT 43.8 ± 7.9 to 30.7 ± 7.5 points (20.0%); both p < 0.001 |
| V27 | No |
| V28 | Not applicable |
| V29 | No treatment-related adverse events reported; the procedure was performed without local anesthesia and without causing pain |
| V30 | Yes |
| V31 | No |
| V32 | No |
| V33 | Not applicable |
| V34 | No |
| V35 | No |
| V36 | No |
| V37 | Yes |
| V38 | Yes |
| V39 | Not applicable |
| V40 | Yes |

**[117]	Sopel, M.; Kuberka, I.; Szczuka, I.; Taradaj, J.; Rosińczuk, J.; Dymarek, R. Can shockwave treatment elicit a molecular response to enhance clinical outcomes in pressure ulcers? The SHOck Waves in wouNds Project. *Biomedicines* 2024, *12*(2), 359. https://doi.org/10.3390/biomedicines12020359.**

**Hypothesis:** This study tested the hypothesis that rESWT enhances the healing of PUs by promoting cellular proliferation, improving vascularization and increasing myofibroblast formation.

**Methods:** This case series involved 10 patients (mean age 85.8 years) with EPUAP grade II–III PUs, treated with two sessions of rESWT using a Cellactor device (Storz Medical, Tägerwilen, Switzerland). The protocol included 300 initial ESWs followed by 100 ESWs/cm^2^ of wound surface area (WSA), a pressure of 2.5 bar, an EFD of 0.15 mJ/mm^2^ and a frequency of 5 Hz, performed twice a week with a 3-day interval. The primary outcome was wound healing, assessed by WSA and wound bed score (WBS) at baseline (M0) and after the first (M1) and second (M2) treatments. Secondary outcomes included microvascular density (CD31), proliferation index (Ki-67) and myofibroblast presence (α-SMA), measured at the same time points.

**Results:** Mean WSA decreased from 13.5 cm^2^ at baseline to 9.9 cm^2^ at M1 and 7.2 cm^2^ at M2, all comparisons statistically significant (M0 vs M1 p = 0.005, M1 vs M2 p = 0.005, M0 vs M2 p = 0.005; overall Friedman p < 0.001). WBS increased from a mean of 2.7 points at baseline to 7.3 points at M1 and 11.3 points at M2 (p < 0.001). The proliferation index (Ki-67) and microvascular density (CD31) both increased significantly post-treatment, indicating enhanced cellular activity and angiogenesis, and α-SMA expression also increased significantly, suggesting improved extracellular matrix remodeling.

**Conclusions:** rESWT is a promising adjunct therapy for PUs, significantly improving wound area, wound bed score, cellular proliferation and microvascular density. The increased myofibroblast formation and angiogenesis further suggest that ESWT promotes a more favorable environment for wound healing. Future studies with larger sample sizes and control groups are necessary to confirm these findings and establish optimal treatment protocols.

| **V** | **Result** |
| --- | --- |
| V1 | PUs, classified between grades II and III according to EPUAP |
| V2 | Mean WSA: 13.5 ± 13.6 cm^2^ |
| V3 | Chronic wounds, mean duration of 7.5 months |
| V4 | Case series |
| V5 | Category 5 |
| V6 | rESWT |
| V7 | Not applicable |
| V8 | 10 |
| V9 | Not applicable |
| V10 | Cellactor (Storz Medical, Tägerwilen, Switzerland) |
| V11 | rESWT |
| V12 | 0.15 mJ/mm^2^ |
| V13 | Not specified |
| V14 | 300 ESWs initial + 100 ESWs/cm^2^ of wound surface area |
| V15 | 100 ESWs/cm^2^ |
| V16 | 5 Hz |
| V17 | 2 sessions |
| V18 | 3 days |
| V19 | Wound healing as assessed by WSA and WBS |
| V20 | M0 (baseline), M1 (24 hours after first session), M2 (24 hours after second session) |
| V21 | Microvascular density, proliferation index, α-SMA expression; collected tissue specimens underwent histological and immunohistochemical assessments |
| V22 | M0, M1, M2 |
| V23 | Not provided |
| V24 | Not provided |
| V25 | Statistically significant improvement in WSA and WBS after rESWT |
| V26 | Significant increase in proliferation index (Ki-67), microvascular density (CD31) and myofibroblasts (α-SMA) |
| V27 | Not provided |
| V28 | Not provided |
| V29 | Not provided |
| V30 | Yes |
| V31 | No |
| V32 | No |
| V33 | Not applicable (no control group) |
| V34 | No |
| V35 | No |
| V36 | No |
| V37 | Yes |
| V38 | Yes |
| V39 | Not applicable (no control group) |
| V40 | Yes |

**[118]	Yang, R.; Ren, L.; Wang, H.; Guo, L.; Liu, L.; Chen, M.; Tian, X. Extracorporeal shockwave therapy for the treatment of deep dermal burns of the hand: A preliminary study. *J. Plast. Reconstr. Aesthet. Surg.* 2025, *102*, 185–194. https://doi.org/10.1016/j.bjps.2025.01.050.**

**Hypothesis:** This study tested the hypothesis that rESWT enhances wound healing in deep dermal burns of the hand, potentially through improved blood perfusion and reduced inflammation.

**Methods:** This double-blind RCT involved 40 patients with deep dermal burns of the hand caused by flame, hydrothermal, chemical or flash burns, randomized to rESWT (n=20) or sham rESWT plus routine dressing treatment (control, n=20). rESWT used a Swiss DolorClast device (Electro Medical Systems, Nyon, Switzerland), 15 mm applicator, 1.0 bar, quoted energy level 0.01 mJ/mm^2^, 300 baseline impulses + 100 impulses/cm^2^ of wound surface, 8 Hz, in 3 sessions on days 3, 5 and 7 post-injury. The control (RT) group received identical routine dressing treatment plus a sham procedure with the device switched off; patients wore opaque masks. The primary endpoints were the percentage reduction in wound area between day 1 and day 14 (termed "percentage of wound healing" by the authors) and wound blood perfusion. Secondary outcomes were infection scores (days 3, 5, 7, 14), Visual Analog Scale (VAS) pain score on days 3, 5 and 7 and Vancouver Scar Scale (VSS) at 6 months after discharge.

**Results:** At day 14, the median percentage reduction in wound area was significantly higher with rESWT (78%; IQR 67-85%) than in the control group (33%; IQR 26-45%) (p < 0.001); complete wound closure was not reported. Blood perfusion increased significantly versus baseline on days 3, 5 and 7. Infection scores were significantly lower in the rESWT group on day 7 (p < 0.05) and VAS pain scores were lower on days 3, 5 and 7 (p < 0.05). At 6 months post-discharge, VSS scores were significantly lower in the ESWT group (p < 0.05), suggesting better scar quality.

**Conclusions:** rESWT significantly improved the healing of deep dermal burns of the hand compared with sham treatment plus routine dressings, reducing infection, alleviating pain and producing better scar outcomes. These findings support rESWT as an effective adjunctive treatment, offering a non-invasive alternative to surgery, especially for patients unable to undergo surgical intervention.

| **V** | **Result** |
| --- | --- |
| V1 | Deep dermal burns of the hand |
| V2 | %TBSA only, no cm^2^: range 0.5-3%; median (IQR) rESWT 2.00 (1.25-2.50)%, sham control 1.75 (1.00-2.50)%, p = 0.383 |
| V3 | Acute (since 3 days post-injury) |
| V4 | RCT |
| V5 | Category 1 |
| V6 | rESWT |
| V7 | Sham rESWT: identical routine dressing plus same procedure with device switched off; patients wore opaque masks |
| V8 | 20 |
| V9 | 20 |
| V10 | Swiss DolorClast (Electro Medical Systems, Nyon, Switzerland) |
| V11 | rESWT |
| V12 | 0.01 mJ/mm^2^ |
| V13 | Not specified |
| V14 | 300 baseline impulses + 100 impulses per cm^2^ of wound surface |
| V15 | 100 ESWs/cm2 (plus a fixed 300 baseline impulses per wound) |
| V16 | 8 Hz |
| V17 | 3 sessions (days 3, 5 and 7 post-injury) |
| V18 | 2 days |
| V19 | Wound healing percentage (days 1 and 14) and blood perfusion by laser speckle contrast imaging; both primary |
| V20 | 14 days post-injury |
| V21 | Wound infection, VAS pain, VSS; no biopsies or tissue samples analyzed |
| V22 | Infection: days 3, 5, 7, 14 post-injury; VAS pain: days 3, 5, 7; VSS: 6 months post-discharge |
| V23 | No |
| V24 | Not applicable |
| V25 | rESWT group: median 78% (IQR 67-85%) healing at 14 days; control group: median 33% (IQR 26-45%) (p < 0.001) |
| V26 | rESWT: lower infection (day 7), lower VAS pain (days 3, 5, 7), better VSS (6 months post-dsscharge); p < 0.05 |
| V27 | No |
| V28 | Not applicable |
| V29 | No adverse events in the rESWT group (e.g., blisters, infection) |
| V30 | Yes |
| V31 | Yes |
| V32 | No |
| V33 | Yes |
| V34 | Yes |
| V35 | No |
| V36 | Yes |
| V37 | Yes |
| V38 | Yes |
| V39 | Yes |
| V40 | Yes |
