## Supplementary File 6 for "Extracorporeal Shock Wave Therapy for Wound Management: Clinical Evidence, Energy Delivery Parameters and Mechanistic Insights — A Systematic Review"

**Extracorporeal Shock Wave Therapy in Wound Management: A Comprehensive Systematic Review of Clinical Evidence, Modalities and Mechanisms**

by Brent Musolf, Carmen Nussbaum-Krammer, Michael O’Neal, Nicola Maffulli and Christoph Schmitz

**Supplementary File 6**

**Standardized summaries and 40 key variables (c.f. Table 2 in the main text) from clinical studies on ESWT with undisclosed ESWT modality in wound management**

**(numbers in brackets refer to the reference numbers in the main text)**

**Note on the standardized summaries:** Each summary condenses one clinical study as it was reported, together with the interpretation that its own authors placed on their findings. The summaries therefore reproduce the position of the respective authors and the state of the field at the time of that publication, and not the assessment of the present systematic review. They follow a uniform structure (Hypothesis, Methods, Results and Conclusions), because the original abstracts differ widely in structure, in length and in the information they report, which makes direct comparison between publications difficult; the standardized form is intended to remove this obstacle.

Abbreviations (in alphabetical order): CABG, coronary artery bypass grafting; DFUs, diabetic foot ulcers; EFD, energy flux density; eNOS, endothelial nitric oxide synthase; ESWT, extracorporeal shock wave therapy; ESWs, extracorporeal shock waves; fESWT, focused ESWT; HBOT, hyperbaric oxygen therapy; LDI, laser Doppler imaging; PCNA; proliferating cell nuclear antigen; RCT, randomized controlled trial; SWC, standard wound care; TcPO₂, transcutaneous partial oxygen pressure; V, variable; VEGF, vascular endothelial growth factor; VLUs, venous leg ulcers.

**[29]	Omar, M.T.; Alghadir, A.; Al-Wahhabi, K.K.; Al-Askar, A.B. Efficacy of shock wave therapy on chronic diabetic foot ulcer: A single-blinded randomized controlled clinical trial. *Diabetes Res. Clin. Pract.* 2014, *106*(3), 548–554. https://doi.org/10.1016/j.diabres.2014.09.024.**

**Hypothesis:** This study tested the hypothesis that ESWT improves the healing rate, wound surface area (WSA) reduction and wound bed preparation in chronic DFUs compared to a control group receiving SWC.

**Methods:** This RCT involved 38 patients with 45 chronic DFUs, of which 24 ulcers (19 patients) were treated with ESWT and 21 ulcers (19 patients) with SWC. The ESWT group received ESWT twice a week for a total of 8 sessions. The ESWs were applied at a frequency of 100 ESWs/cm^2^ and an EFD of 0.11 mJ/cm^2^. The primary outcome measure was the percentage of wound surface area reduction, while secondary outcomes included the percentage of granulation tissue, the amount of exudates and the wound bed preparation scores. Outcomes were assessed at baseline, after 8 weeks of treatment and at a 20-week follow-up.

**Results:** The ESWT group showed significantly greater reductions in wound size than the control group. After 8 weeks of treatment, the mean percentage reduction in WSA was 60.1% in the ESWT group and 36.2% in the control group (p < 0.05); after 20 weeks it was 83.3% versus 63.3% (p < 0.05). The healing time in the ESWT group was also significantly shorter, averaging 64.5 days compared to 81.2 days in the control group (p < 0.05). Furthermore, 54% of DFUs in the ESWT group were completely healed by the 20-week follow-up, compared to only 28.5% in the control group.

**Conclusions:** ESWT significantly improved the healing rate and reduced the healing time in chronic DFUs compared to standardized wound care. It also produced a significant reduction in wound size and improved wound bed preparation without any adverse reactions. These results suggest that ESWT is a beneficial adjunctive therapy for chronic DFUs.

| **V** | **Result** |
| --- | --- |
| V1 | Chronic DFUs |
| V2 | ESWT group: 7.9 ± 3.0 cm^2^; Control group: 8.6 ± 3.5 cm^2^ |
| V3 | Chronic wounds (resisted to conservative treatment > 3 months) |
| V4 | RCT |
| V5 | Category 1 |
| V6 | ESWT |
| V7 | Standardized wound care (debridement, blood-glucose control agents, footwear modification) |
| V8 | 19 patients (24 ulcers) |
| V9 | 19 patients (21 ulcers) |
| V10 | Not provided |
| V11 | Not specified |
| V12 | 0.11 mJ/cm^2^ as printed in the paper; almost certainly a typographical error for 0.11 mJ/mm^2^ |
| V13 | Not specified |
| V14 | 100 ESWs/cm^2^ |
| V15 | 100 ESWs/cm^2^ |
| V16 | Not provided |
| V17 | 8 sessions (twice per week) per the Methods; the Discussion states that between six and eight sessions were actually given |
| V18 | Ambiguous: twice weekly, one-week interval, eight sessions (Section 2.5, p. 550); inter-session interval not stated |
| V19 | Percentage of WSA reduction, wound healing rate |
| V20 | After 8 weeks (W8), follow-up at 20 weeks (W20) |
| V21 | Wound bed preparation, percentage of granulation tissue and exudates presence; no biopsies or tissue samples analyzed |
| V22 | After 8 weeks (W8), follow-up at 20 weeks (W20) |
| V23 | Yes |
| V24 | 38 total (19 per group), 44 for dropout; detect 20% WSA difference, alpha = 0.05, 85% power |
| V25 | ESWT: 83.3% ± 20.7 reduction in WSA at W20; Control: 63.3% ± 24.9 reduction in WSA at W20 |
| V26 | ESWT group: 54% healed at W20; control group: 28.5% healed at W20 |
| V27 | Not provided |
| V28 | Not applicable |
| V29 | No adverse reactions were observed |
| V30 | Yes |
| V31 | Yes |
| V32 | No |
| V33 | Yes |
| V34 | No |
| V35 | No |
| V36 | Yes |
| V37 | Yes |
| V38 | Yes |
| V39 | Yes |
| V40 | Yes |

**[119]	Vangaveti, V.N.; Jhamb, S.; Goodall, J.; Bulbrook, J.; Biros, E.; Malabu, U.H. Extracorporeal shockwave therapy (ESWT) in the management of diabetic foot ulcer: A prospective randomized clinical trial. *J. Foot Ankle Surg.* 2023, *62*(5), 845–849. https://doi.org/10.1053/j.jfas.2023.04.013.**

**Hypothesis:** This study tested whether adding ESWT to SWC results in significantly better wound healing outcomes in patients with DFUs than SWC alone.

**Methods:** This prospective, single-center RCT randomly allocated 48 patients with DFUs to ESWT plus SWC (25 patients) or SWC only (23 patients). The index ulcer was the largest full-thickness ulcer present. ESWT was applied at 0.11 mJ/mm^2^ EFD, 310 ESWs/cm^2^ and treatment session, and 5 Hz frequency, every 2 weeks over the 6-week treatment period; the total number of sessions was not stated. The primary outcome was wound size change at 6 weeks; secondary outcomes were inflammatory markers (e.g., IL-6, TNF-α, CRP) at baseline and 6 weeks post-baseline. The ESWT device was not disclosed.

**Results:** After 6 weeks there was no statistically significant difference in wound healing between the groups. Ulcer size was reduced in 18 ESWT + SWC patients (86%) versus 14 (70%) with SWC; these denominators are patients with 6-week data (21 and 20, Table 4), not the 25 and 23 randomized. This difference was not statistically significant (unadjusted OR 2.5, 95% CI 0.54-12.1, p = 0.23; adjusted OR 2.9, 95% CI 0.44-19.0, p = 0.26). p = 0.33 refers to mean absolute reduction in wound surface area. Inflammatory marker levels (IL-6, TNF-α, CRP) also did not differ significantly. Complete healing at 6 weeks was slightly more frequent with SWC only (6/23, 26.1%) than with ESWT + SWC (5/25, 20.0%), a non-significant difference (p = 0.73).

**Conclusions:** In this RCT, ESWT did not provide a statistically significant benefit in improving wound healing compared to SWC in patients with DFUs. While ESWT may hold potential in enhancing diabetic wound healing, larger and longer studies are required to confirm its effectiveness.

| **V** | **Result** |
| --- | --- |
| V1 | DFUs |
| V2 | ESWT + SWC: 70 mm^2^ (25-226 mm^2^); SWC: 48 mm^2^ (20-408 mm^2^) |
| V3 | ESWT + SWC: 8.5 weeks (3.7-55.2 weeks); SWC: 7.5 weeks (1.7-47.5 weeks) |
| V4 | RCT |
| V5 | Category 2b |
| V6 | ESWT + SWC |
| V7 | SWC |
| V8 | 25 |
| V9 | 23 |
| V10 | Not provided |
| V11 | Not specified |
| V12 | 0.11 mJ/mm^2^ |
| V13 | Not specified |
| V14 | Minimum 310 ESWs/cm^2^ (adjusted based on wound size) |
| V15 | Minimum 310 ESWs/cm^2^ |
| V16 | 5 Hz |
| V17 | Not stated. Patients were seen and treated every 2 weeks over a 6-week treatment period, implying about 3 sessions. |
| V18 | 2 weeks |
| V19 | Wound healing, measured by ulcer size reduction |
| V20 | 6 weeks post-baseline |
| V21 | Changes in inflammatory markers (IL-6, TNF-α, CRP, etc.); no biopsies or tissue samples analyzed |
| V22 | 6 weeks post-baseline |
| V23 | Yes |
| V24 | Sample size of 25 patients per arm, total 50 (80% power, 12% difference in healing) |
| V25 | No significant difference in wound healing between groups at 6 weeks post-baseline |
| V26 | No significant changes in inflammatory markers between groups at 6 weeks post-baseline |
| V27 | Not provided |
| V28 | Not applicable |
| V29 | Not provided |
| V30 | Yes |
| V31 | Yes |
| V32 | Yes |
| V33 | Yes |
| V34 | No |
| V35 | No |
| V36 | No |
| V37 | Yes |
| V38 | Yes |
| V39 | Yes |
| V40 | Yes |
